# Discovery of a novel antibiotic, Transitmycin, from *Streptomyces* sp unveils highly efficient activities against tuberculosis and human immunodeficiency virus

**DOI:** 10.1101/2024.04.03.24305291

**Authors:** Vanaja Kumar, Balagurunathan Ramasamy, Mukesh Doble, Radhakrishnan Manikkam, Luke Elizabeth Hanna, Gandarvakottai Senthilkumar Arumugam, Kannan Damodharan, Suresh Ganesan, Azger Dusthakeer, Precilla Lucia, Shainaba A Saadhali, Shanthi John, Poongothai Eswaran, Selvakumar Nagamiah, Jaleel UCA, Rakhila M, Ayisha Safeeda, Sathish S

## Abstract

HIV is identified as a factor that aggravates tuberculosis disease pathogenesis and its progression to latent TB. While, TB is declared as one of the major causes for AIDS-associated mortality. So there is a dire need for new drugs to combat such ailments that have a synergistic interaction.This has led us to study a novel antibiotic purified from a marine Streptomyces sp isolated from the coral reef ecosystem of South Indian coast. Streptomyces sp. R2 (MTCC 5597; DSM 26035)., isolated from the marine water was grown on agar plates and the crude yellowish orange pigment secreted was extracted using various solvents. The antibiotic, named as Transitmycin, was purified and tested against M. tuberculosis, drug resistant strains, and M. tuberculosis biofilm. The compound was also tested against HIV-1 viruses belonging to six subtypes. Several characterisation tools were used to elucidate the structure of this novel antibiotic. Transitmycin was derivitaised to elucidate the absolute configurations of the amino acids present in it. Tr, unlike actinomycin D, has L-valine in both the rings instead of D-valine (found in the latter). Also, one of the proline in Tr is in D–configuration while it is in L configuration in actinomycin D suggesting that ours is a novel compound and is not reported so far. It exhibits dual activities against the standard H37Rv, 49 drug sensitive clinical isolates, and MtB biofilm as well as standard and 20 clinical isolates of HIV. This is the first paper that reports the isolation of a new antibiotic from marine actinobacteria exhibiting unusual anti-TB and HIV activities which could be exploited further as a lead molecule in the quest for the design of drug with dual activities.

**Highlights:**

- A novel antibiotic was purified from a marine Streptomyces sp isolated from the coral reef of S. India
- Presence of L-valine, not observed in actinomycin D, and one of the proline in D configuration suggest that it is a novel structure not reported before
- It exhibits activity against standard MtB strain as well as clinical isolates and drug resistance ones
- It exhibits anti-HIV activity against several clinical isolates

## Introduction

Despite, the history of infectious diseases is time immemorial, an urgent need for protection against infectious diseases has one of the major health concerns. Antibiotics have revolutionized the diagnostic factor in various aspects and the drug discovery is considered as a hallmark in human history. However, the concomitant development of resistance against diseases is the most serious consequence [1]. In 2020, an overall rate of 1.5 million people pretentious from TB. In fact, TB is one of the leading sources of death, wherein occupied the 13^th^ position and moreover the second foremost infective destroyer after COVID-19 (above HIV/AIDS). Since it is an airborne infectious disease caused by *M tuberculosis*, continues to be a major cause of mortality and morbidity worldwide [2].

An upsurge of multi drug resistance (MDR) and extensively drug resistance (XDR) is an additional perturbing factor in tuberculosis chemotherapy. Comprehensively, the existing treatment for TB based on half yearly quadruple therapy containing rifampicin, isoniazid, ethambutol and pyrazinamide results the cure rate of 90-95% [3] for drug-sensitive TB at global level. Nevertheless, this regimen is inadequate to treat MDR and XDR-TB infections, sometimes the second line antibiotics are employed to treat MDR-TB infections for prolonged time whereas XDR infections are almost untreatable. Different TB drugs with unique mode of action are desperately necessary to fight against drug resistant tuberculosis. For example, bedaquiline and delamanid are approved for drug resistant TB, however due to the reported toxicity issues, they may employ only on last resort [4]. In addition, the second line drugs used to treat MDR-TB with several disadvantages such as less efficacious, more toxic, and more expensive than the first-line drugs [5]. At presently progressing anti TB drugs should partake essential properties like succeed a short therapy period (sterilizing effect), a simpler regimen, capable of act on MDR/XDR-TB, concurrent treatment of TB/HIV and advanced safety level than the existing drugs [6]. Alike the human immunodeficiency virus (HIV) leads to destruction of the immune system resulting a condition termed as Acquired Immuno Deficiency Syndrome (AIDS). After its finding AIDS becomes a great risk to humans, reflected to be pandemic and it is the greatest public health crisis in globally. As of 2021, millions of people living with HIV were rescuing antiretroviral therapy [7]. Several researches have been developed the drugs against HIV disease, such as the growth of many antiretroviral drugs (ARVs), including the atazanavir, ibalizumab, darunavir, abacavir, zidovudine, lamivudine, efavirenz, tenofovir disoproxil fumarate, tenofovir alafenamide, emtricitabine, bictegravir, cabotegravir, rilpivirine etc. nevertheless, the competency of HIV intensive to mutate rapidly leads to the upsurge of drug resistance to existing anti-retrovirals [8]. Since the finding of penicillin, many of the microbial natural products tends to the new paradigm for novel drug discovery development [9,10].

Over the past 60 years, [11] the microbial bioprospecting has gained a remarkable quantity of medicinal components, including actinobacteria, the major bacterial phylum group of Gram-positive bacteria with guanine and cytosine (G+C) rich content in their genomic sequence, which are the important prokaryotes in economic and ecological manner. Significantly the existence of actinobacterial multiplicity in several rare ecosystems liable for prolific producers of many biologically active natural compounds with different potential activities [12–15]. Some of the pharmaceutical companies have dramatically declined in the result of novel wide range antibiotics in the past few decades [16]. Obviously, this critical state has caused in the surplus of such antibiotics with distinct mode of activity targeting the arcade, wherein this precluded the assessment of usual terrestrial sources in particular for actinobacteria and led scientists to pursuit unique habitats on the marine family for novel bioactive molecules [12,17,18]. Indeed, actinobacteria are ubiquitous in the marine environment, which are occupying a substantial ecological characteristic in the recycling and production of novel natural products with huge pharmaceutical applications [19].

Generally, the 16S rRNA gene sequencing has concomitantly employed to identify, classify and quantitation of microbes in multifaceted biological molecules. It is obvious from the 16S rRNA sequencing that marine microbial species as bacteria and archae have an extreme different taxonomy. Even the 16S rRNA sequences of more than ten thousand actinomycetes had been isolated hitherto from marine outsources. The discovery of novel secondary metabolites from marine actinobacteria has just surpassed that of their terrestrial counterparts [15]. According to the review of Blunt et al., [20] shown that there are 179 new natural products isolated during 2016, in which the actinobacterial genus *Streptomyces* remains to be the dominant source. An extensive study on typical marine actinobacterial genera like *Salinispora* and *Verrucosispora* produces salinosporamide and abyssomycin respectively, which suggest that actinobacteria improves significant feature towards marine drug discovery research [21]. Further, marine based antibiotics are more ingenious in combat infections due to the terrestrial bacteria do not have chance to develop resistance against them [22]. During the microbial bioprospecting process, the actinobacterial extracts isolated from various less-explored ecosystems were screened for anti-TB and anti-HIV activity. The isolation, characterization and potential bioactivities against TB and HIV of an antibiotic obtained from a marine *Streptomyces* sp. R2 is being reported herewith.

## Results and Discussion

### Description of the actinobacterial strain

*Transitmycin* is a depsipeptide (bicyclic) molecule produced by actinobacterial strain R2, which was isolated from the sediment samples from Rameswaram coral reef ecosystem (Lat. 9.2876° N; Long. 79.3129° E), Tamil Nadu, South India using Starch Casein Agar (SCA) supplemented with nalidixic acid (20µg/ml) and nystatin (100µg/ml). Viability of strain was maintained in International Streptomyces Project-2 (ISP2) agar slants, 30% glycerol broth followed by lyophilisation. It was also deposited in Microbial Type Culture Collection (MTCC), India and in DSMZ – German Collection of Microorganisms and Cell Cultures, Germany. (R2 = MTCC5597; DSM26035).

### Characterization and taxonomy

The characterization of a strain is the key element in classification of prokaryotes including actinobacteria [23]. Actinobacterial classification was initially based mainly on morphological and physiological characteristics [24–25]. The onset of chemotaxonomic standards has provided reproducible and reliable data to identify the genera at genus level [26]. This microbial strain R2 under microscopic observation showed the presence of dense aerial and substrate mycelia with long, un-fragmented spore chains with hairy structures (Supplementary Fig. 1a-b). The physiological and biochemical characteristics of the strain R2 are given in Supplementary S-Table 1. Strain R2 showed good growth in various ISP media and utilized a wide range of carbon and nitrogen sources. In addition, good growth was also observed at different physiological conditions. Notably, the extracellular yellow pigment production was greatly influenced by nitrogen substrates, pH, temperature and NaCl. Strain R2 produced lipase and amylase. High sensitivity was observed for most of the antibiotics tested (Supplementary S-Table 1). The cell wall analysis revealed that the strain R2 is rich in LL-2,6-Diaminopimelic acid (L-DAP) and glycine. No diagnostic sugars were found in the cell wall constituents. In our present study, the results of phenotypic characterization and cell wall analysis indicated that the actinobacterial strain R2 belongs to the genus *Streptomyces*, however it is not adequate for differentiation at species level.

Furthermore, the PCR (Polymerase Chain Reaction) amplification of 16S rRNA gene of strain R2 formed around 1400 base pair sequence and the BLAST (Basic Local Alignment Search Tool) analysis have shown 99% similarity to the 16S rRNA gene sequence of *Streptomyces variabilis* (EU570414) and other closely related species submitted in GenBank. Phylogenetic relationship of the strain R2 and related taxa are given in Supplementary S-Fig. 1c.

The 16S rRNA gene sequence of strain R2 has the accession number HQ012501at GenBank, but this gene provides limited resolution for species level identification, since it discloses more extensive genotypic differences. At this stage, the average nucleotide identity (ANI) [27] of all preserved genes between any two genomes shows likely to reform taxonomy, because it also correlates with the best DNA: DNA Hybridization (DDH) values. The most often used standards for species delineation *i.e*. the 70% DDH [28] which is closely equivalent to 95% ANI (Average Nucleotide Identity) values. Moreover, no organisms have been defined hitherto has shown <98.7% identity in their 16s rRNA gene and shown <95% ANI or 70% DDH. These results supported the substitution of cumbersome DDH and related procedures with simple sequence-based standards. Significantly the metabolic property of transitmycin production had not been reported before from any other meticulously associated *Streptomyces* species. Based on the aforesaid facts, the actinobacterial strain R2 is identified to be a novel strain of *Streptomyces variabilis*, but still it showed 99% similarity with its associated proximity neighbors. It is very important to mention here that there is no literature evidence on any commercially available antibiotic, in particular anti-TB and/or anti-HIV compounds from *Streptomyces variabilis*.

### Transitmycin production

The growth of *Streptomyces* sp. R2 is simultaneously observed with grey colour aerial mycelia and soluble yellow orange pigment in good yield. The crude compound is extracted well in several solvents namely, methanol, chloroform and dichloromethane, followed by diethyl ether and ethyl acetate (Supplementary Table 2). The extracts in the former solvents were intensely coloured when compared to the extracts from the latter solvents. Salts and debris were present in the extracts while using former solvents, so ultimately ethyl acetate was chosen for extracting the bioactive pigment.

The activity of the crude extract was also tested on different strains of *M. <u>tuberculosis</u>.* More than 95% reduction in Relative Light Units (RLU) was observed through Luciferase Reporter Phage Assay against all three *M. <u>tuberculosis</u>* strains. When 1.0 L of YEME agar was used for production it yielded 800 mg crude antibiotic extract in ethyl acetate. Major antibiotics reported from actinobacteria are extracellular in nature [29–30]. Industrial production of many antibiotics from *Streptomyces* is achieved through submerged fermentation process [31]. In contrast, some actinobacterial strains are found to produce antibiotics only on solid media and very little reason is reporeted as to why activity is restricted to solid culture and not observed in submerged cultures. Shomura *et al reported* [32] that about 1300 out of 6500 actinomycetes showed antimicrobial activity against one or more of the test organisms when tested by agar plug method. In the secondary screening, about 25 (1.9%) of the 1300 strains were found to be non-producers in submerged cultures. So, we conlcude that the reports of antibiotics isolated from only agar cultures of actinomycetes are very rare.

According to Mayurama *et al*. [33] fumaridamycin was detected with much difficulty in submerged cultures, because the mycelium of the producing strain inactivate the antibiotic more readily in liquid than in agar culture. Similarly, Shomura*et al* [32] demonstrated that the antibiotic produced by *Streptomyces halstedii* has shown activity against Gram negative bacteria only in agar dishes, which correlates well with its mycelial morphology. The aerial mycelium was filamentous during antibiotic production in solid cultures, but fragmented in non-producing liquid cultures. Similar to report from Shomura *et al*. [32] the bioactive pigment production by the *Streptomyces* strain R2 was observed only in agar culture, where the vegetative mycelium was filamentous. While decreasing the concentration of YEME broth from 2X to 1/10X the mycelia filamentation was found to increase. However, none of the five concentrations of YEME broth produced the pigment (unpublished data). This outcome evidenced that the filamentous mycelial structure does not influence pigment production by the strain R2 in broth while the bioactive pigment production was observed in all the concentrations of YEME agar when filamentous mycelia was formed. In accordance to Ohnishi *et al*. [34] who reported that 2-aminophenoxazin-3-one containing grixazone A and B, afford yellow pigments during phosphate depletion by *Streptomyces griseus*. By adopting these strategies, it was concluded that optimizing the medium components may trigger the pigmented bioactive compound production by our isolated *Streptomyces* sp. R2 in liquid cultures.

### Transitmycin purification, characterization and structure elucidation

The *Streptomyces* sp. R2 was found to produce 800 mg of crude pigment per litre of yeast extract malt extract medium in agar surface fermentation. The yellow-orange pigment was separated from the crude ethyl acetate extract by TLC and column chromatography and its purity was confirmed by HPLC analysis (Supplementary Fig S13-S17).

Primarily the three well separated spots *viz.* R1, R2 and R3 with R_f_ values of 0.8, 0.6, 0.3, respectively were observed on analytical TLC using EtOAc:MeOH (9.5:0.5) solvent system. In bioassay directed isolation, fraction R1 (named as Transitmycin) showed more than 95% inhibition against *M. tuberculosis* strain H37Rv in LRP assay. It was isolated as an orange colour amorphous powder with [α]_D_^25^: -106° (*c* = 0.2, MeOH). The R1, R2, R3 in crude extract and as purified compounds R1, R2, R3 showed similar retention time in RP-HPLC chromatograph (Fig, 1a-d). A single peak of transitmycin at a RT of 5.8 minutes confirmed its purity (Fig. 1c).

**Fig. 1.**
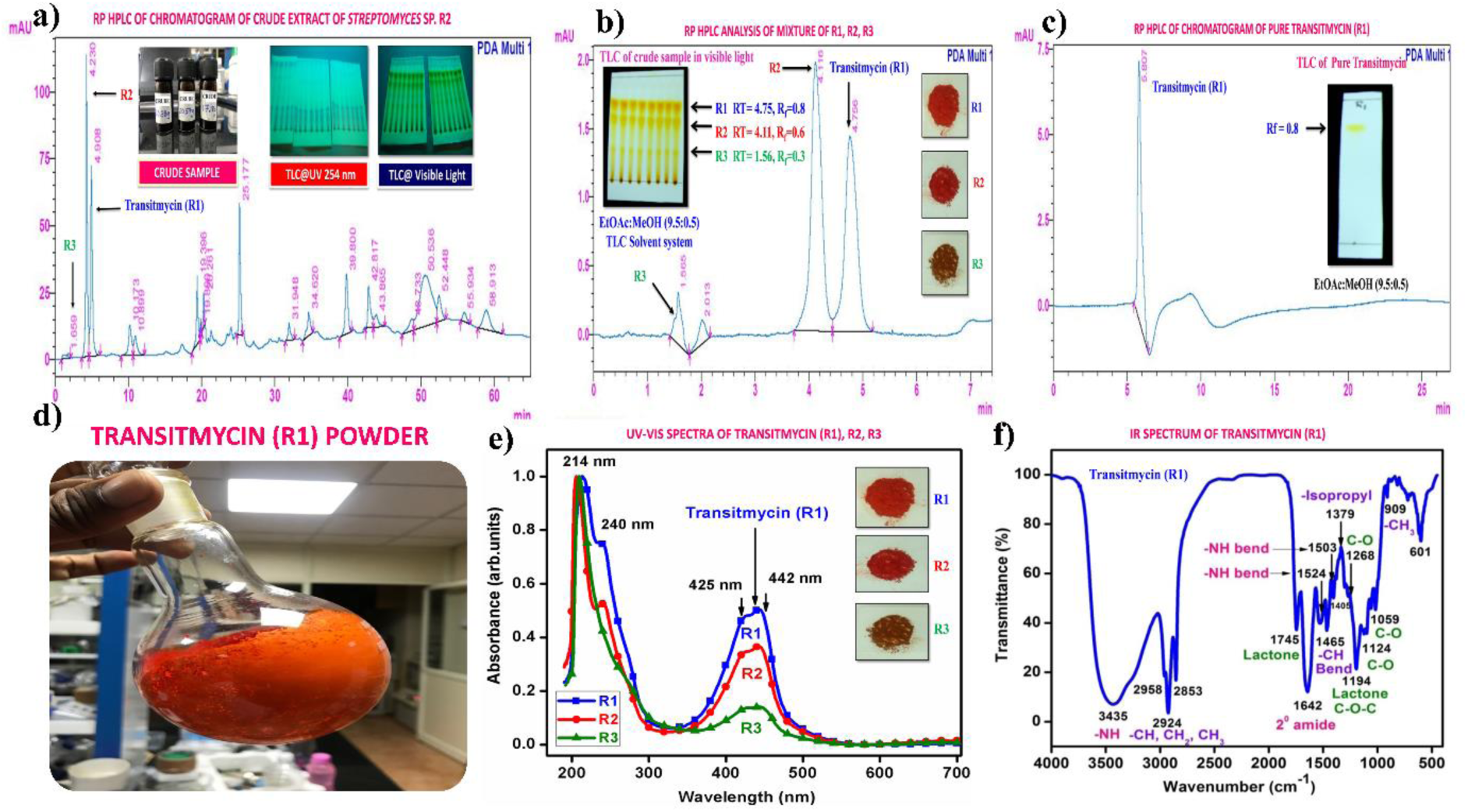
Representative HPLC chromatogram of crude and isolated samples R1, R2 and R3. **a** RP HPLC of Chromatogram of crude extract of *Streptomyces* sp. R2. **b** RP HPLC analysis of mixture of R1, R2, R3. **c** RP HPLC of Chromatogram of pure Transitmycin R1. **d** Transitmycin R1 obtained after column chromatographic purification. **e** UV-Vis spectra of Transitmycin R1, R2 and R3. **f** IR Spectrum of Transitmycin R1.

The chemical structures of these molecules (R1, R2 and R3) were elucidated by UV-Visible, IR, CD, ^1^H, ^13^C, DEPT 135 NMR, 2D NMR (^1^H-^1^H COSY, ^1^H-^1^H DQF-COSY, ^1^H-^1^H TOCSY, ^1^H-^13^C HSQC, ^1^H-^13^C HMBC, NOSEY, ROESY) and MALDI-TOF-MS, HR-ESIMS, HR-EIMS, HR-LCMS, and 3200 QTRAP-LC/MS/MS analyses and also compared with the previously reported NMR and Mass data of actinomycins (Supplementary Information) [35–57]. Advances in spectroscopic techniques have mainly utilized for compound identification and immensely accelerated the unambiguous representation of compound characterization and structural elucidations. The UV-Vis spectrum of Transitmycin showed a strong absorption band around (λ_max_ 214, 240, 425, 442 nm (in MeOH) (Fig. 1e). The colour and absorption peak in UV-Vis analysis revealed the presence of phenoxazinone chromophore. Singh *et al*. [58] and Maskey *et al*. [59] detected the presence of phenoxazinone chromophore in bioactive metabolites from *Streptomyces* sp. and *Actinomadura* sp. The absolute configuration of the amino acids were supposed to be identical to that of actinomycin D, as indicated by the negative optical rotation values and the strong cotton effect at about 210 nm in the CD spectra. The CD values of Transitmycin (R1) are: [MeOH, [nm], (mdeg)] λ_max_(Δε) 195 (+24.0), 210 (-21.5), 241 (+1.7).

In the IR spectrum, a strong absorption broad band appears at around 3435 cm^-1^, an intense strong peak at 1746 cm^-1^ and a band around 1099 cm^-1^ are assignable to be amino (or hydroxyl), lactone ring and alicyclic 6-membered ether type (C-O) groups, respectively. IR data of transitmycin (R1) (KBr cm^-1^), 3435 cm^-1^ for NH, 2958, 2924 cm^-1^ (m, -CH str, asym, CH_3_ and CH_2_), 2872 cm^-1^, 2853 cm^-1^ (m, -CH str, sym, CH_3_ and CH_2_),1746 cm^-1^ (s, C=O str, lactone ring), 1642 cm^-1^(s, -C=O str, 2° amide), 1524, 1503 (m, -NH bend, 2° amide), 1466 (m, CH bend (scissoring), CH_2_), 1379 cm^-1^ (s, -CH bend, isopropyl group), 1268 (s, C-O str, ester), 1194 (C-O-C of lactone) 1099, 1059, 1017 (s, C-O or C-N), 720, 712, 694, 689 (s, - CH bend, oop, aromatic ring), 909 (w, CH_3_ rocking) [48] (Fig. 1f).

The ^1^H and ^13^C NMR spectra exhibited the typical features of two (alpha and beta ring) pentapeptido lactone ring attached with phenoxazinone chromophore, i.e., each ring contained four amide carbonyl resonances and one ester carbonyl in one ring (δ_C_ 168.8, 173.4, 173.1, 166.2, 167.4 (α-ring) and 168.9, 173.9, 172.7, 166.4, 167.4 (β-ring), together with phenoxazinone chromophore (101.8 (C-1), 147.3 (C-2), 179.0 (C-3), 113.4 (C-4), 144.9 (C-4a), 140.3 (C-5a), 127.6 (C-6), 130.2 (C-7), 126.0 (C-8), 132.1(C-9), 128.4 (C-9a), 14.9 (C-11), 7.6 (C-12), 166.0 (C-13), 165.8 C-14). One of the amino acid proline in the β-ring contained a keto group (208.0 in the ^13^C NMR). From ^1^H NMR spectrum, NH of amino acids (δ_H_ 8.23, β-L-Valine), (δ_H_ 7.74, α-L-Valine), (δ_H_ 7.69, β-L-Threonine, (δ_H_ 7.2, α-L-Threonine), (δ_H_ 6.55, 2H of β-proline) (δ_H_ 5.93 2H of α-proline) and four N-methyl groups (δ_H_ 2.91, 2.88, 2.89, 2.87) (Fig. 2 a-d). In addition, the ^1^H NMR spectra of Transitmycin indicated the presence of eight methyl groups arising from four isopropyls.

**Fig. 2.**
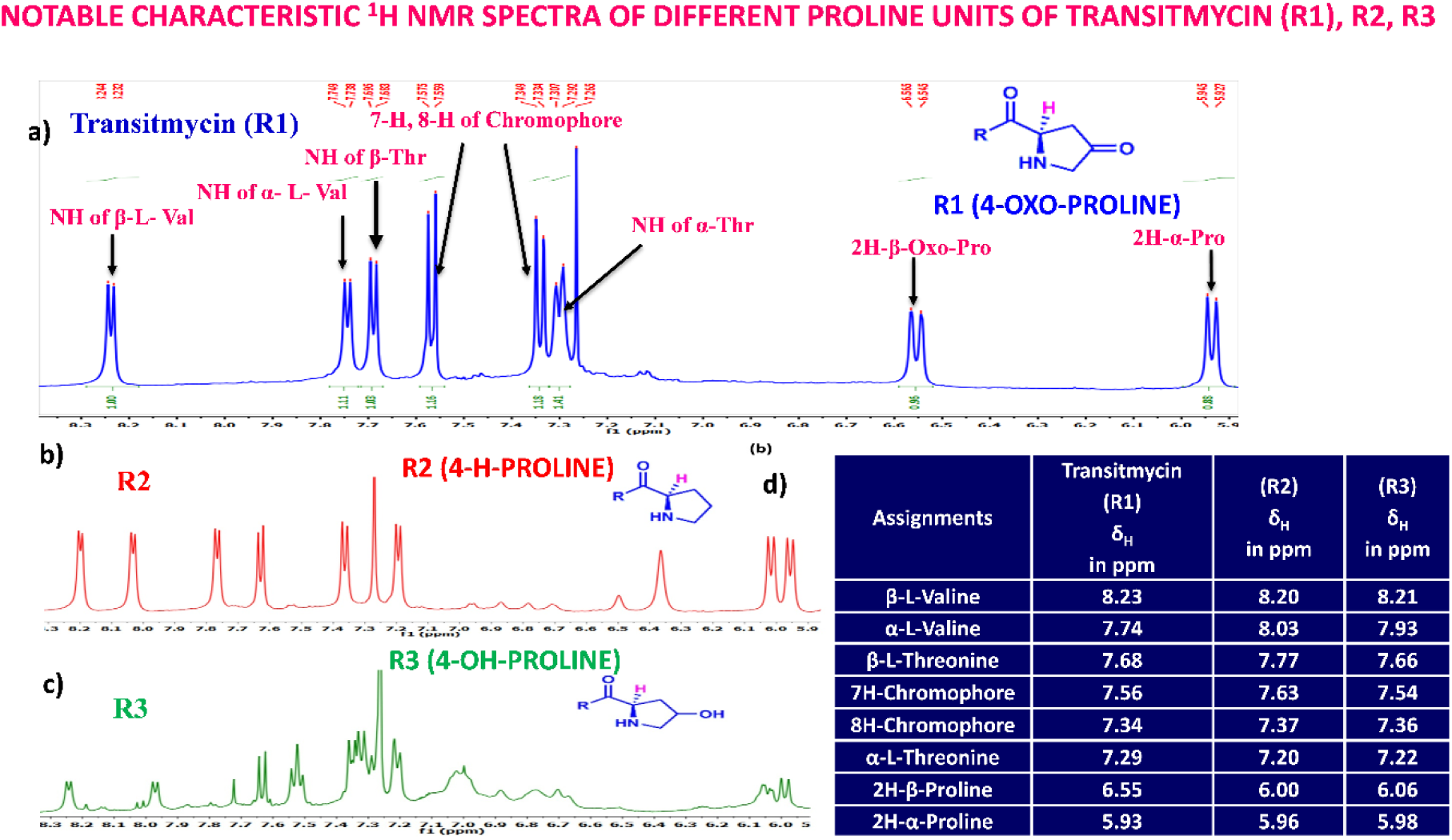
Notable characteristic ^1^H NMR spectra of different proline units of Transitmycin as **a** R1 (4-oxoproline). **b** R2 (proline) and **c** R3 (4-OH proline). **d** Comparative studies for ^1^H NMR (500 MHz, CDCl_3_) chemical shift value of NH containing amino acid residue and chromophore motifs of Transitmycin (R1), R2 and R3.

The UV/Vis absorption spectra with maximal absorbance at 240 nm and 442 nm support the presence of an amino phenoxazinone chromophore in its structure. From ^1^H - ^1^H COSY and TOCSY experiments, five amino acid systems namely Pro, Thr, Val, N-Methyl Val, and Sar were identified. The assignments of the protonated carbons were obtained from the HSQC spectrum, in combination with inspection of the HMBC spectrum. By comparison of the UV spectrum *(λ_max_* 442 nm, in MeOH) of Transitmycin with that of actinomycin series *(λ_max_* 440 nm, in MeOH) (Fig. 1e) it was concluded that the former contained an aminophenoxazinone chromophore residue. In ^1^H NMR, two *ortho* coupled protons at 7-H, 8-H; δ_H_ 7.56 and 7.34 corresponding to 1,2,3,4-tetrasubstituted aromatic ring, and two 3H singlet at δ_H_ 2.52 (11-CH_3_) and 1.95 (12-CH_3_) of methyl groups in peri-position of the aromatic system were identified. This is the characteristics of phenoxazinone chromophore found in various actinomycins.

The result was further confirmed by HMBC correlations between the 7-H (δ_H_ 7.59) and 8-H (δ_H_ 7.34) of the tetra substituted double bond and the carbonyl resonances at δ_C_ 166.0. The carbonyl carbons of Pro, Thr, Sar, Val, and N-methyl Val, were clearly assigned to (δ_C_ 179.0, 174.0, 173.1, 169.02, 198.8, 167.5, 166.5, 166.56, 166.3, 166.1 and 165.9) on the basis of the observed correlations between carbonyl groups protons of the same amino acid residue in the HMBC spectrum. All the residues were connected on the basis of DQF-COSY, TOCSY, HMBC, ROESY and NOESY correlations (Fig. 3a-c), thus establishing the amino acid sequences and overall constitution. (Supplementary Fig. S14-65, S-Table 5). The detailed analysis of ^1^H-^1^H COSY, ^1^H−^1^H DQF-COSY, ^1^H−^13^C HMBC, and ^1^H−^13^C HSQC, ^1^H−^1^H TOCSY, ^1^H−^1^H NOESY, ^1^H−^1^H ROESY NMR spectra [35–51] (Supplementary Information Fig. S14-65, S-Table S5) and MALDI-TOF-MS (Fig. 4a-m and Fig. 5) and HR-ESIMS, HR-LCMS, ESI-MS, QTRAP LC-MS/MS [52–57] (Fig. 5, S-Table S6), (Supplementary Information Fig. S14-115, S-Table S7a-b) spectral fragmentation pattern revealed ten amino acids in Transitmycin (Fig. 3d), which is identical to those present in actinomycin X2 [35–57] (2 X MeVal, 2 X Thr, 2 X Sar, 2 X Val, proline and ketoproline). OPro was identified by the ketone moiety (δ_C_ 208.6 ppm) and the altered chemical shifts and coupling patterns of the neighbouring methylene groups (Table 1).

**Fig. 3.**
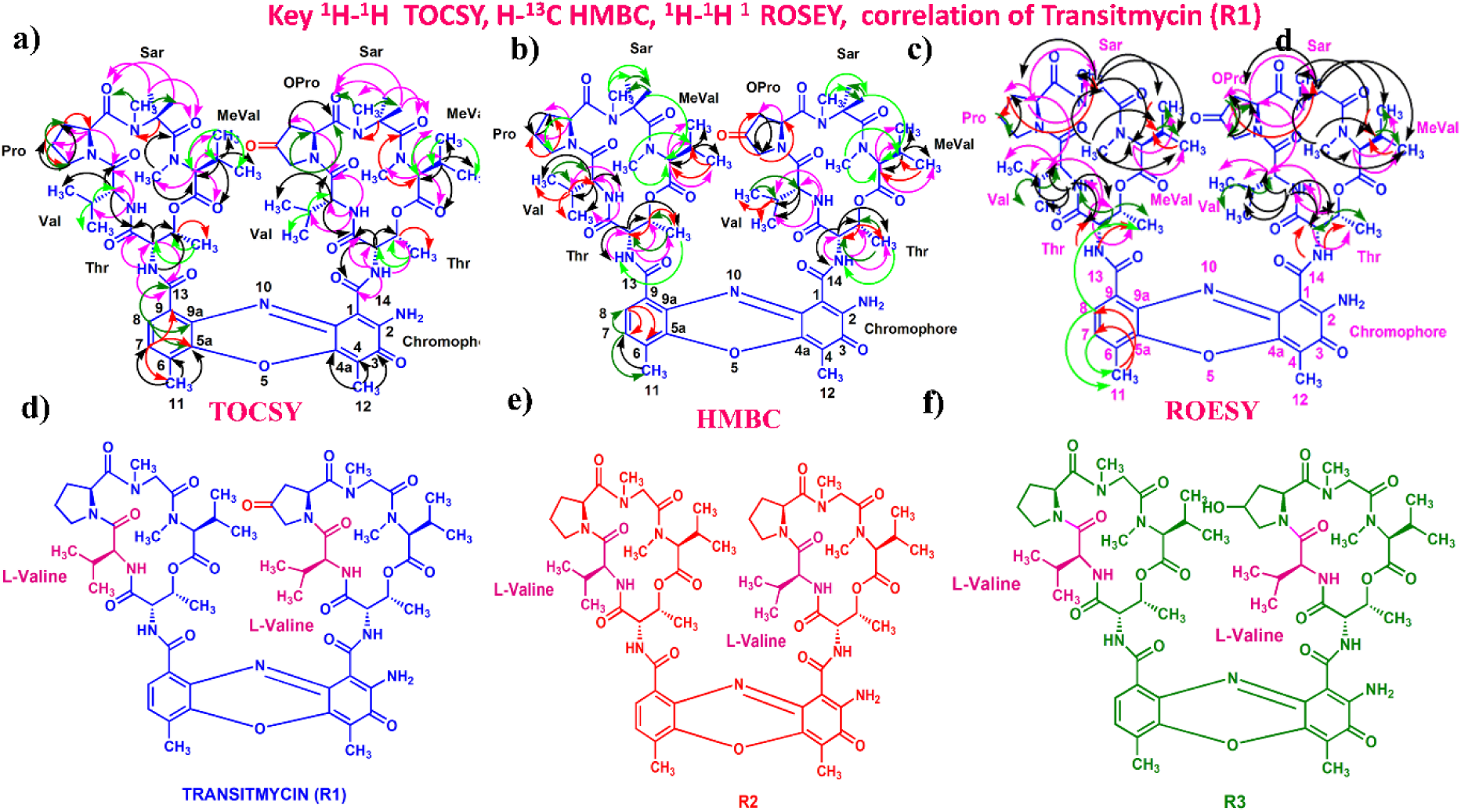
**a** Key ^1^H-^1^H COSY correlation of Transitmycin (R1). **b** Key ^1^H-^1^H TOCSY correlation of Transitmycin (R1). **c** Key ^1^H-^13^C HMBC connectivity for Transitmycin (R1). **c** ^1^H-^1^H ROESY correlation of Transitmycin (R1). **d** Chemical structure of isolated compounds Transitmycin (R1). **e** R2. **f** R3.

**Fig. 4.**
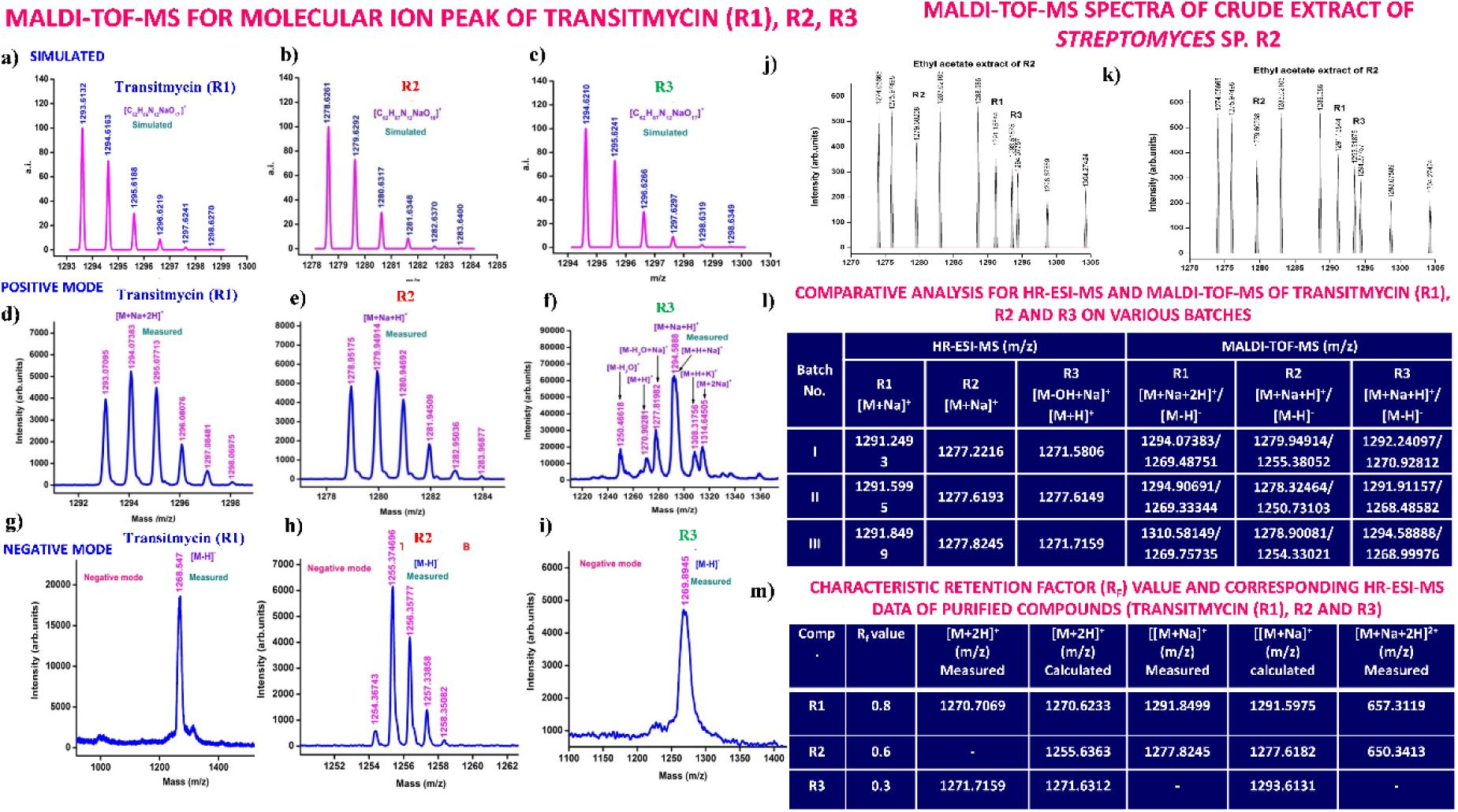
**a-m**. Comparative analysis of MALDI-TOF-MS for molecular ion peak of Transitmycin (R1), R2, R3. **a** R1 (Simulated). **b** R2 (Simulated). **c** R3 (Simulated). **d** R1 (Measured, positive mode. **e** R2 (Measured, Positive mode). **f R**3 (Measured, Positive mode). **g** R1 (Measured, Negative mode). **h** R2 (Measured, Negative mode). **i** R3 (Measured, Negative mode), MALDI-TOF-MS spectra of crude extract of *Streptomyces* sp. R2. **j** positive mode. **k** negative mode **l** Comparative analysis for HR-ESI-MS and MALDI-TOF-MS of Transitmycin (R1), R2 and R3 on various batches. **m** Characteristic retention factor (R_f_) value and corresponding HR-ESI-MS data of purified compounds (Transitmycin (R1), R2 and R3).

**Fig. 5.**
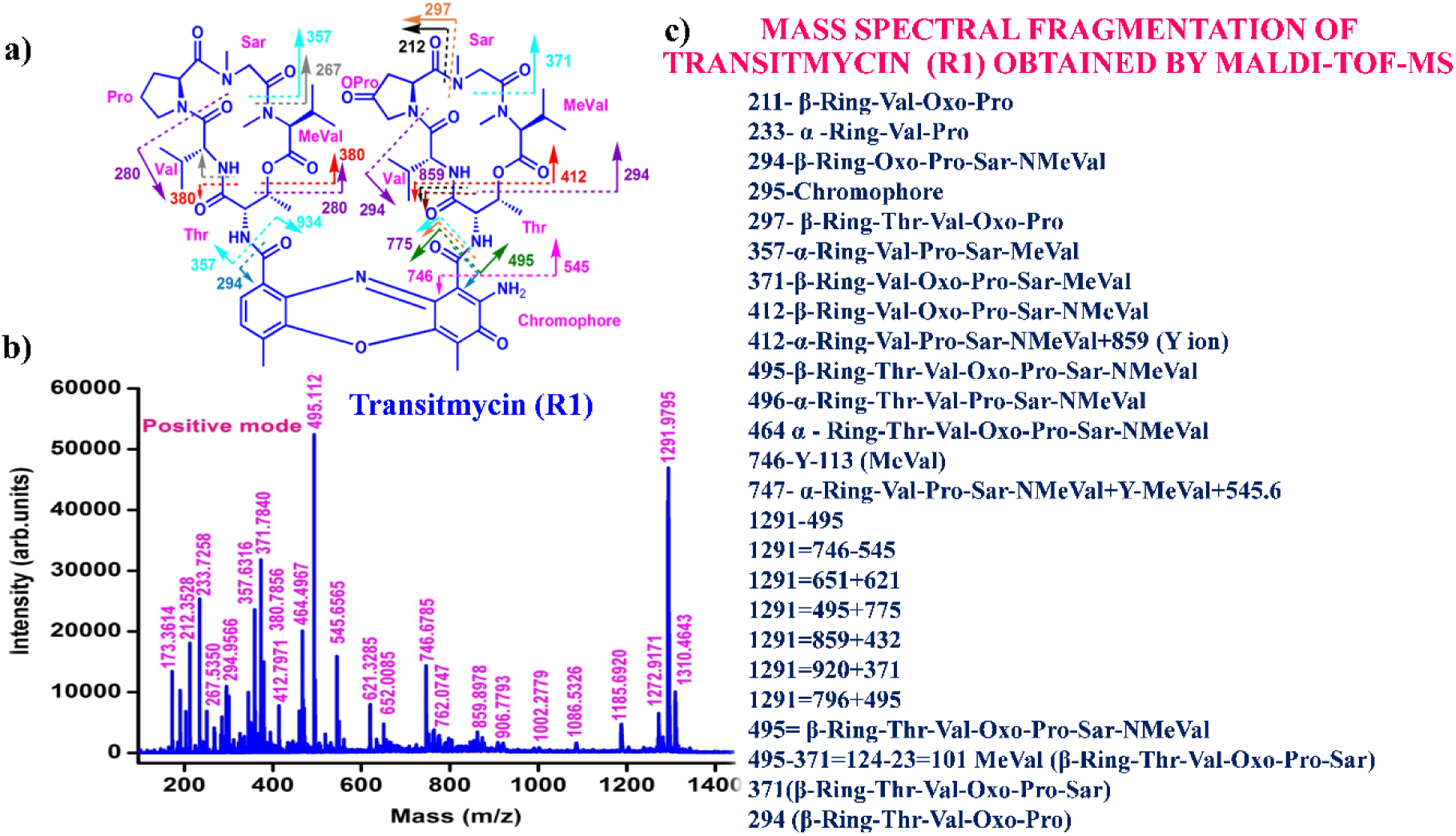
**a** Mass spectral fragmentation of Transitmycin (R1) obtained by MALDI-TOF-MS, **b** MALDI-TOF-MS spectrum of Transitmycin (R1). **c** Mass spectral fragmentation of Transitmycin (R1) obtained by MALDI-TOF-MS, 211-β-Ring-Val-Oxo-Pro, 233-α -Ring- Val-Pro, 294-β-Ring-Oxo-Pro-Sar-NMeVal, 295-Chromophore, 297-β-Ring-Thr-Val-Oxo-Pro, 357-α-Ring-Val-Pro-Sar-MeVal, 371-β-Ring-Val-Oxo-Pro-Sar-MeVal, 412-β-Ring-Val-Oxo-Pro-Sar-NMeVal, 412-α-Ring-Val-Pro-Sar-NMeVal+859 (Y ion), 495-β-Ring-Thr-Val-Oxo-Pro-Sar-NMeVal, 496-α-Ring-Thr-Val-Pro-Sar-NMeVal, 464 α - Ring-Thr- Val-Oxo-Pro-Sar-NMeVal, 746-Y-113 (MeVal), 747- α-Ring-Val-Pro-Sar-NMeVal+Y- MeVal+545.6, 495= β-Ring-Thr-Val-Oxo-Pro-Sar-NMeVal, 495-371=124-23=101 MeVal (β-Ring-Thr-Val-Oxo-Pro-Sar), 371(β-Ring-Thr-Val-Oxo-Pro-Sar), 294 (β-Ring-Thr-Val- Oxo-Pro).

**Table 1.** ^1^H NMR (500 MHz, CDCl_3_)/^13^C NMR (125 MHz, CDCl_3_) and 2D NMR (500 MHz, CDCl_3_) correlation spectral data of Transitmycin (R1) ^1^ H−^1^ H COSY, ^1^H−^1^H TOCSY ^1^H−^13^C HMBC, ^1^H−^1^H ROESY.

| α- ring | Position | $\delta_C$ | $\delta_H$ | $J$ (Hz) | COSY | TOCSY | HMBC | ROSEY |
| --- | --- | --- | --- | --- | --- | --- | --- | --- |
| **Thr** | 1 | 168.8 | - | - | - | - | - | - |
|  | 2 | 54.6 | 4.60 | d, 10.0 | 3, 4, -NH | 3, 4 | 1, 3, 13 | 3, 4, 2-Val, Val-NH |
|  | 3 | 74.7 | 5.13 | m | 2, 4, -NH | 2, 4, | 4 | 2, 4, Val-NH |
|  | 4 | 17.0 | 1.10 | d, 6.5 | 2, 3 | 2, 3, -NH | 2, 3 | 2, 3, Val-NH |
|  | NH | - | 7.29 | d, 7.0 | 2, 3, 4 | 2, 3, 4 | 1, 3, 13 | 2, 4 |
| **L-Val** | 1 | 173.4 | - | - | - | - | - | - |
|  | 2 | 58.7 | 3.58 | dd, 10.5, 6.5 | 3, 4, 5, -NH | 3, 4, 5, -NH | 1, 5, 1-Thr | 3, 4, 5, 2-Pro |
|  | 3 | 31.7 | 2.08 | m | 2, 4, 5 | 2, 4, 5 | 2, 5 | 4, 5, 4-Thr |
|  | 4 | 19.0 | 0.89 | d, 7.0 | 2, 3 | 2, 3, -NH | 2, 3, 5 | 2, 3, 5 |
|  | 5 | 18.9 | 1.13 | d, 6.5 | 2, 3 | 2, 3, -NH | 2, 3, 4 | 2, 3, 2-Val, Val-NH |
|  | NH | - | 7.74 | d, 5.5 | 2, 3, 4, 5 | 2, 3, 4, 5 | 2, 3, 1-Thr | 2, 3, 4, 5, 2-Thr, 3-Thr |
| **Pro** | 1 | 173.1 | - | - | - | - | - | - |
|  | 2 | 56.4 | 5.93 | d, 9.5 | 3a, 3b, 4a, 4b | 3a, 3b 4a, 4b | 3, 4, 5, 1-Val | 3a, 3b, 2a-Sar, 2b-Sar, Sar-NMe |
|  | 3 | 31.6 | 2.73<br>1.84 | dd, 17.5, 7.0<br>dd, 19.5, 6.5 | 3b, 4a, 4b<br>3a, 4a, | 3b, 4a, 4b<br>3a, 4a, | 1, 2, 4<br>1, 4, 5 | 3b, 2a-Sar, 2b-Sar<br>3a, 4a, 4b |
|  | 4 | 23.3 | 2.26<br>2.01 | d, 16.5<br>d, 8.0 | 3a, 3b, 4b<br>3b, 4a | 3a, 3b, 4b<br>3b, 4a | 3, 5<br>3, 5 | 4b, 5a, 5b<br>4a, 5a |
|  | 5a<br>5b | 47.8 | 3.86<br>3.67 | d, 18.5<br>d, 18.5 | 3a, 4a, 4b<br>3a, 4a, 4b | 3a, 4a, 4b<br>3a, 4a, 4b | 4, 1-Val | 5b |
| **Sar** | 1 | 166.2 | - | - | - | - | - | - |
|  | 2 | 51.2 | 2a, 4.70<br>2b, 3.64 | d, 17.5<br>d, 17.5 | 2b<br>2a | 2b, NMe<br>2a, NMe | 1, NMe, 1-pro<br>1, NMe, 1-Pro | 2b, NMe, 2-Pro<br>4-MeVal, 5-MeVal, NMe |
|  | N-Me | 34.9 | 2.87 | s |  | 2a, 2b | 2, 1-Pro | 2b, 3-MeVal |
| **Me-Val** | 1 | 167.4 | - | - | - | - | - | - |
|  | 2 | 71.3 | 2.70 | d, 9.0 | 3, 4, 5 | 3, 4, 5 | 1, 3, 4, NMe | 4, 5 |
|  | 3 | 26.9 | 2.62 | d, 7.0 | 2, 4, 5 | 2, 4, 5 | 1, 3, 4, 5, NMe | 4, 5 |
|  | 4 | 21.6 | 0.96 | d, 6.0 | 2, 3 | 2, 3, 5 | 2, 3, 5 | 3 |
|  | 5 | 19.2 | 0.72 | d, 6.5 | 2, 3 | 2, 3, 4 | 2, 3, 4 | 3 |
|  | N-Me | 39.2 | 2.89 | s | - | 2, 3 | 2, 1-Sar | 2, 3, 5, 2a-Sar, 2b-Sar |

| β-ring | position | $\delta_C$ | $\delta_H$ | $J$ (Hz) | COSY | TOCSY | HMBC | NOESY |
| --- | --- | --- | --- | --- | --- | --- | --- | --- |
| **Thr** | 1 | 168.9 | - | - | - | - | - | - |
|  | 2 | 54.9 | 4.50 | d, 9.0 | 3,4, -NH | 3, 4 | 1, 3, 14 | 3, 4, 2-Val, Val-NH |
|  | 3 | 74.6 | 5.23 | m | 2,4,- NH | 2, 4 | 4 | 2,4, Val-NH |
|  | 4 | 17.6 | 1.20 | d, 6.0 | 2, 3 | 2, 3, - NH | 2, 3 | 2,3, Val-NH |
|  | NH | - | 7.68 | d, 6.0 | 2, 3, 4 | 2, 3, 4 | 1, 3, 14 | 2, 4 |
| **L-Val** | 1 | 173.9 | - | - | - | - | - | - |
|  | 2 | 57.1 | 3.71 | dd,<br>10.5,<br>6.5 | 3, 4, 5, -<br>NH | 3, 4, 5, -NH | 1, 3, 5, 1-Thr, | 3, 4, 5, 2-Oxo pro |
|  | 3 | 31.8 | 2.20 | m | 2, 4, 5 | 2, 4, 5 | 2, 5 | 4, 5, 4-Thr |
|  | 4 | 19.0 | 0.86 | d, 7.0 | 2, 3 | 2, 3, -NH | 2, 3, 5 | 2, 3, 5 |
|  | 5 | 18.8 | 1.14 | d, 7.0 | 2, 3 | 2, 3, -NH | 2, 3, 4 | 2, 3, 2-Val, Val-NH |
|  | NH |  | 8.23 | d, 6.0 | 2, 3, 4, 5 | 2, 3, 4, 5 | 2, 3, 1-Thr | 2, 3, 4, 5, 2-Thr, 3-Thr |
| **Oxo-Pro** | 1 | 172.7 | - |  |  |  |  |  |
|  | 2 | 54.2 | 6.55 | dd,<br>10.0,<br>2.0 | 3, 5 | 3, 5 | 3, 1-Val | 3, 5, 2a-Sar, 2b-sar-NMe |
|  | 3 | 41.8 | 2.32<br>2.13 | d, 18.5<br>d, 13.0 | 2, 5<br>2, 5 | 2, 5<br>2, 5 | 1<br>1 | 2, 5<br>2, 5 |
|  | 4 | 208.0 | - | - | - | - | - | - |
|  | 5 | 53.7 | 3.83<br>3.90 | d, 18.5<br>dd,<br>18.5,<br>12.0 | 2, 3<br>2, 3 | 2, 3<br>2, 3 | 1, 2<br>1, 2 | 2, 3<br>2, 3 |
| **Sar** | 1 | 166.4 | - | - | - | - | - | - |
|  | 2 | 51.28 | 2a,<br>4.54<br>2b,<br>3.95 | d, 19.5<br>d, 19.0 | 2b<br>2a | 2b, NMe<br>2a, NMe | 1, NMe, 1-Pro<br>1, NMe, 1-Pro | 2b, NMe, 2-oxo-pro<br>4-MeVal, 5-MeVal,<br>NMe |
|  | N-Me | 34.7 | 2.88 | s | - | 2a, 2b | 2, 1-Pro | 2b, 2MeVal |
| **Me-Val** | 1 | 167.4 | - | - | - | - | - | - |
|  | 2 | 71.1 | 2.67 | d, 8.5 | 3, 4, 5 | 3, 4, 5 | 1, 3, 4, 5,<br>NMe | 4, 5 |
|  | 3 | 26.8 | 2.60 | d, 8.0 | 2, 4, 5 | 2, 4, 5 | 1, 3, 4, 5,<br>NMe | 4, 5 |
|  | 4 | 21.5 | 0.93 | d, 6.0 | 2, 3 | 2, 3, 5 | 2, 3, 5 | 3 |
|  | 5 | 19.1 | 0.73 | d, 6.0 | 2, 3 | 2, 3, 4 | 2, 3, 4 | 3 |
|  | N-Me | 39.0 | 2.91 | s | - | 2, 3 | 2, 1- Sar | 2, 5, 2a-Sar, 2b-Sar |

**Chromophore**
| Position | $\delta_C$ | $\delta_H$ | $J$ (Hz) | COSY | TOCSY | HMBC | NOESY |
| --- | --- | --- | --- | --- | --- | --- | --- |
| 1 | 101.8 | - | - | - | - | - | - |
| 2 | 147.3 | - | - | - | - | - | - |
| 3 | 179.0 | - | - | - | - | - | - |
| 4 | 113.4 | - | - | - | - | - | - |
| 4a | 144.9 | - | - | - | - | - | - |
| 5 | - | - | - | - | - | - | - |
| 5a | 140.3 | - | - | - | - | - | - |
| 6 | 127.6 | - | - | - | - | - | - |
| 7 | 130.2 | 7.34 | d, 8.0 | 8, 11 | 8, 11 | 5a, 9, 11-Me | 11 |
| 8 | 126.0 | 7.56 | d, 7.5 | 7, 11 | 7, 11 | 9a, 5a, 13 | 11 |
| 9 | 132.1 | - | - | - | - | - | - |
| 9a | 128.4 | - | - | - | - | - | - |
| 10 | - | - | - | - | - | - | - |
| 11 | 14.9 | 2.52 | s | 7, 8 | 7, 8 | 5a, 6, 7 | 7, 8 |
| 12 | 7.6 | 2.18 | s | - | - | 3, 4, 4a | - |
| 13 | 166.0 | - | - | - | - | - | - |
| 14 | 165.8 | - | - | - | - | - | - |

The MALDI-TOF-MS spectrum of crude extract of Streptomyces sp. R2 (Fig. 4j-k) and measured, calculated mass of purified compounds Transitmycin (R1), R2, R3 were given in (Fig. 4a-i, Supplementary Figure S70-115). The molecular formula was established as C_62_H_84_N_12_NaO_17_ based on Positive HRESI-MS, which showed protonated pseudo molecular ion peak [M+H] ^+^ at m/z 1270.7069 (Fig. 4a-m). It also showed intense peaks, due to Na and K adducts, at m/z 1291.8307 [M+Na]^+^ and 1307.8124. [M+K]^+^ respectively (Calcd. for C_62_H_84_N_12_NaO_17_: 1291.5975: Found: 1291.8307). Fig. 4a-i Similarly, the MALDI TOF MS spectrum of transitmycin showed intense peak in positive mode at *m/z* 1293.61316 [M+Na+2H] ^+3^ and at m/z 1309.93062 [M+K]^+^ and in negative mode at m/z 1269.33344 [M-H]- (Fig. 4a-m, Fig. 4-5, Supplementary Information Fig. S115a-b). The compound R2 was isolated as orange red powder and its molecular formula was established as C_62_H_86_N_12_O_16_ [M+Na]^+^ by positive HRESIMS. The MALDI TOF molecular ion of R2 showed peak at *m/z* 1278.95175 [M+Na+H]^+^ and negative ion mode at *m/z* 1255.38052 [M-H]^-^ For a molecular formula of C_62_H_87_N_12_O_16_Na its molecular weight is calculated as 1278.6261 [M+Na+H]^+^ which matches with 1278.95175, that is similar to that of actinomycin D (Fig. 3e). The compound R3 was obtained as an oranges red powder and the molecular formula of R3 was determined to be C_62_H_87_N_12_O_17_ from HR-ESIMS peak at *m/z* 1271.7159z [M+H]^+^ and when calculated for the molecular formula, C_62_H_87_N_12_O_17_ 1271.6312, found to be 1271.7159 and 1277.6149 [M-OH-Na]^+^. The molecular formula of R3 was established as C_62_H_87_N_12_O_17_ by positive MALDI-TOF 1294.5888 [M+H+Na]^+^ and negative mode 1268.4852 [M-H]- (Fig. 4f) which is identical to that of actinomycin 0β (Supplementary Information Fig. S70-115, S-Table S7a-b) [52–57].

The differences between compounds R1, R2, R3 were becaue of the variation in proline at 4^th^ position. Transitmycin (R1) has keto group in the 4^th^ position, R2 does not have keto group and compound R3 has hydroxyl group in the 4^th^ position (Fig. 3a-c).

### HPLC Analysis of L-FDAA Derivatives of Transitmycin (R1)

HPLC experiment was used to determine the absolute configuration of isolated compounds. The absolute configurations of the amino acids were assumed to be identical to that of actinomycin X_2_, as indicated by the negative optical rotation values and the strong cotton effect at about 210 nm in the CD spectra. The assignment of the amino acids was carried out primarily by the analysis of the ^1^H-^13^C HSQC and ^1^H-^1^H-COSYcorrelations, MALDI-TOF-MS (Fig. 4 a-m, S-Table S6), QTRAP LC-MS/MS (Supplementary Information Fig. S14-114, S115a-b, S-Table S7a-b), and completed with the help of HMBC spectrum (Fig. 3a-c). Additionally, a small amount of compound Transitmycin and R3 were hydrolyzed and the free amino acid were analyzed by HPLC, HR-LCMS, HR-ESIMS after chiral derivatization with Marfey’s reagent. Altered proline and sarcocine were not available as reference. Although neither the altered proline and threonine moieties nor N-methylated alanine were available as references, they can be assumed to possess L-configurations due to the fact that the exchange of a single amino acid with its enantiomer leads to significant conformational changes of the respective peptidolactone ring, resulting in reduced biological activities as well as chemical shift deviations that have not been observed. The absolute configuration of the threonine, valine, methyl valine, proline in Transitmycin (R1) was clarified by Marfey’s method applied for the acid hydrolysate of Transitmycin (R1) in comparison with standard amino acid analysis [60–64]. Retention times of the standard N^α^-(5-fluro-2,4-di-nitrophenyl)- L-alanamide (FDAA) derivatives were as follows: D-threonine, 15.682 min; L-threonine, 13.987 min; D-proline, 17.035 min; L-proline, 16.519 min; D-valine, 21.244 min; L-valine, 19.248 min; D-N-methyl valine, 22.089 min; L-Methyl valine 20.814 min. The chromatogram of the FDAA derivatives of acid hydrolysate Transitmycin (R1) showed peaks corresponding to L-Threonine (14.021 min), L-proline (16.520 min), L-valine (19.252), L-N-methyl valine (21.263 min) were obtained in the hydrolysate. The above same mentioned procedure for compound R3 was also done and similar results was obtained. (Supplementary Information Fig. S116-125, Table S-8a-b) [60–64]. Comparison, with authentic standards, revealed the presence of L-MeVal, L-Thr and D-Val as expected, however the D-Valine is in L-configuration as well as one of the proline is in D-proline instead of L-configuration. (Fig. 6a-c).

**Fig. 6.**
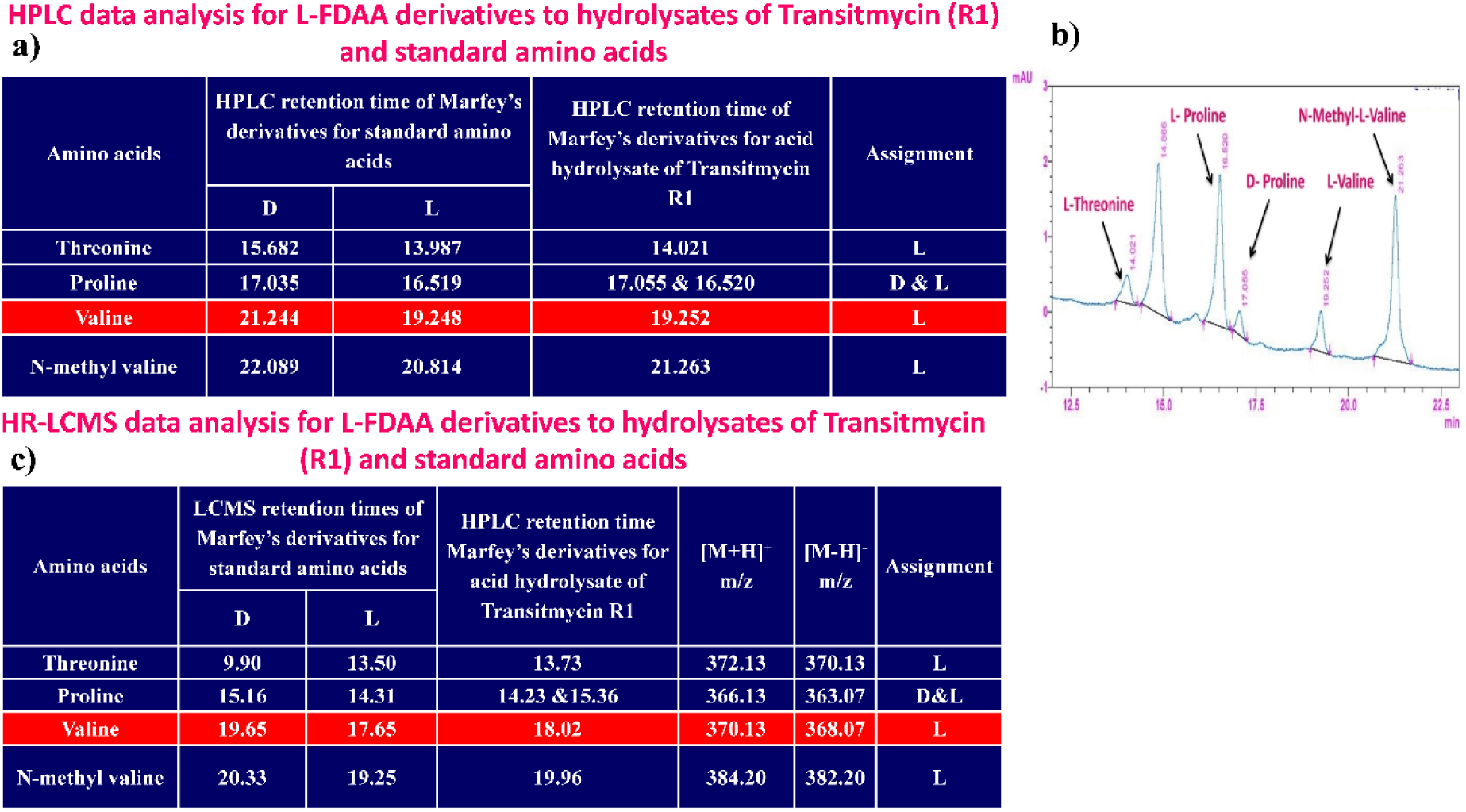
**a** HPLC data analysis for L-FDAA derivatives to hydrolysates of Transitmycin R1 and standard amino acids. **b** Graphical representation for RP-HPLC chromatogram of L-FDAA (Marfey’s) derivatives of hydrosylated Transitmycin (R1). **c** HR-LCMS data analysis for L-FDAA derivatives to hydrolysates of Transitmycin R1 and standard amino acids.

### LC-MS Analysis of L-FDAA Derivatives of Transitmycin (R1)

The retention times of the D- and L-FDAA derivatives of standard amino acid, respectively, were as follows: Pro: 14.35, 14.97, min, *m/z* 366.13 [M +H]^+^, *m/z* 363.07 [M-H]^+^;Val:19.63,17.52 min, *m/z* 370.13 [M+H]^+^, 368.13 [M-H]^-^; Thr: 9.90, 13.50 min, *m/z*372.07 [M+H]^+^, 370.13 [M − H]^-^; NMeVal: 20.33, 19.23 min, *m/z* 384.20[M+H]^+^382.20 [M –H]^-^. The L-FDAA was used to derivatize the acid hydrolysates of Transitmycin (R1), R3 and eight standard amino acids (D-Val, L-Val, D-Thr, L-Thr, D-N-MeVal, L-N-MeVal, D-Pro and L-Pro). The reaction with L-FDAA was performed with the same procedure as above. [63–64].The retention times of the L-FDAA derivatives were as follows: D-Pro:15.16 min, L-Pro:14.31 min, *m/z* 366.13 [M+H]^+^ 363.07 [M−H]^-^;D-Val: 19.65 min, L-Val 17.65 min *m/z* 370.13 [M+H]^+^368.07[M−H]^−^; D-Thr: 9.90 min, L-Thr, 13.50 min, *m/z* 372.13[M+H]^+^370.13 [M−H]^-^; D-N-MeVal 20.33 min, L-N-MeVal 19.25 min, *m/z* 384.20[M+H]^+^,382.20 [M+H]^-^. The retention time of L-FLDA derivatives of acid hydrolysates Transitmycin (R1) were as follows: L-threonine (13.73 min), L-Proline (14.23 min), D-Proline or keto-proline (15.36 min), L-valine (18.02 min), N-Methyl valine (19.96 min) as illustrated in Fig. 6 a-b. The retention time of L-FLDA derivatives of acid hydrolysates (R3) (Supplementary Information Fig. S-128-161, S-Table S9 a and b) were as follows: to L-threonine (12.45 min) or D-threonine (9.36 min), L-Proline (14.25 min), D-Proline or keto-proline (15.21 min), L-valine (17.49 min), N-Methyl valine (19.93). Comparison with authentic standards revealed the presence of L-MeVal, L-Thr, L-Proline, L-Valine and one of the Proline as in D-configuration (Fig. 6c). Hence, we named the unusual newly found compound as Transitmycin (Fig. 3d, Table 1& 2), a member of the X-type [35–64].

### Determination of anti TB and anti-HIV activity

#### Activity against planktonic cultures of M. tuberculosis

Despite the introduction of new anti TB drugs, emergence of antibiotic resistance among *M. tuberculosis* strains remains a major challenge in tuberculosis therapy. According to WHO Global Tuberculosis report - 2022, there is a 3.1% increase in incidence of multi drug resistant and rifampicin resistant TB cases from 2020 to 2021[65]. Thus, there is a burning need for more new antitubercular drugs to tackle drug resistant tuberculosis [66]. Since 1940s, secondary metabolites and their associated derivatives have played a key role in anti-TB drugs development. This is best exemplified by an extremely active aminoglycoside, namely streptomycin, the first clinical drug that was made available against TB [67]. In a previous study, Streptocytosines A, Bamitecin and Amitecin isolated from sea water *Streptomyces* in Japan showed activity against *M. smegmatis* at 32, 16 and 8 µg/ml concentrations, respectively [68]. In another study, actinomycin X2 and actinomycin D isolated from marine *Streptomyces* sp. MS449 in China showed activity against *M. tuberculosis* H37Rv at 1.92mg/ml and 1.77 mg/ml, respectively [69]. In the present study Transitmycin, a novel molecule isolated from a marine *Streptomyces* sp. R2 picked up from coral reef ecosystem showed activity notably against drug sensitive, multi drug resistant (MDR) and mono resistant strains of *M. tuberculosis* at concentrations of 5 and 10 µg/ml (Fig. 7). Based on the preliminary experiment on laboratory strain, *M. tuberculosis* H37Rv, minimum inhibitory concentration for transitmycin was determined on clinical isolates which included 49 drug sensitive strains and 48 drug resistant strains. The drug resistant profile of the clinical isolates are given in (supplementary S-table 12). Out of 97 clinical isolates, MIC at 5 µg/ml was seen in 89 isolates and MIC at 10 µg/ml was seen in 8 isolates for transitmycin based on LRP assay.

**Fig. 7.**
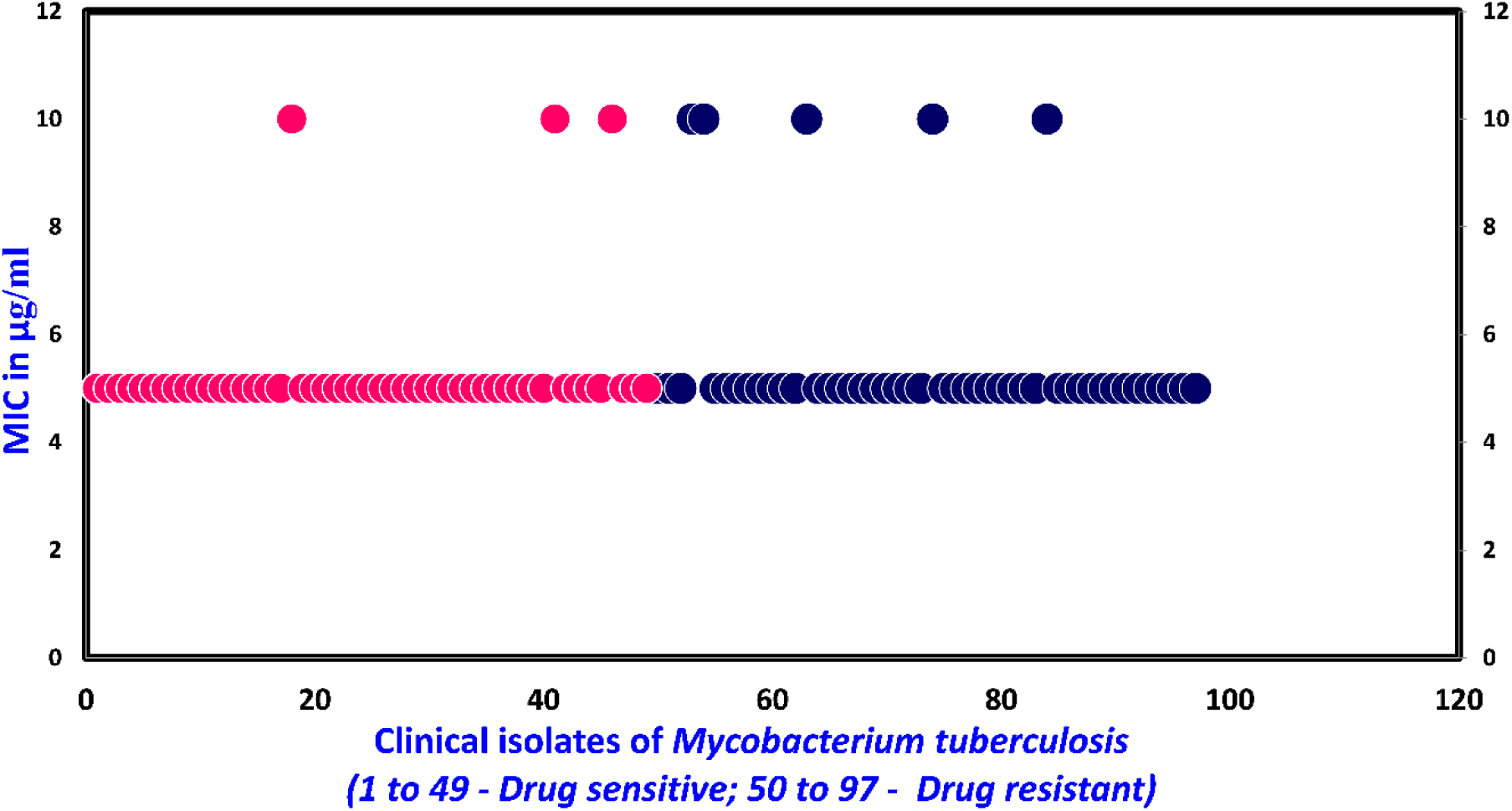
MIC of Transitmycin in micrograms per millilitre for *Mycobacterium tuberculosis* isolates susceptible to anti-TB drugs (n=49) and drug resistant to one or more anti-TB drugs (n=48) as determined by LRP assay. Sensitive strains are represented in blue colour and resistant strains are represented in brown colour.

### Activity against M. tuberculosis biofilm

Bacteria can persist for extended periods of time inside the biofilm due to their ability to resist the immune system, display increased virulence and become phenotypically more resistant to antibiotics. Moreover, antibiotic concentrations required to control bacteria within a biofilm are estimated to be 100–1,000 fold greater than that is needed to treat planktonic forms. As planktonically-grown *M. tuberculosis* are unlikely to be entirely representing the bacterial load during human infection, we set out to determine how effective transitmycin can be against *M. tuberculosis* growing as a biofilm, a bacterial phenotype known to be more resistant to antibiotic treatment [70]. In the present study, *M. tuberculosis* culture showed biofilm formation in the control wells alone which could be observed by the naked eye. The wells with cells and transitmycin failed to produce biofilm. CFU determined prior to the addition of transitmycin was 1.9 x 10^6^ to 2.3 x 10^6^/ml. After 4 days of exposure with the compound, the CFU dropped to 9 x 10^4^ to 10 x 10^4^ ml. Addition of transitmycin completely killed all the cells at the end of 5 weeks (Table 3).

**Table 2.** Physico-chemical properties of Transitmycin.

| Transitmycin (R1) | Properties |
| --- | --- |
| **TLC** | Single yellow spot with R<sub>f</sub> value 0.8 (EtOAc:MeOH (9.5:0.5)) |
| **Color and consistency** | Orange color amorphous powder |
| **Yield** | 200 mg, 20% |
| **Melting point (mp)** | 240-242°C |
| **[α]<sub>D</sub><sup>25</sup>** | -106 ° (c = 0.2, MeOH) |
| **Solubility** | Soluble in chloroform, dichloromethane, ethyl acetate, methanol, ethanol, acetonitrile, DMSO and water; Insoluble in Hexane |
| **UV** | (MeOH) λ<sub>max</sub> (log ε) 214 (3.07), 240 (2.30), 425 (1.44), 442 (1.51) nm |
| **CD** | [MeOH, [nm], (mdeg)]: λ<sub>max</sub>(Δε) 195 (+11.1), 210 (-21.0), 242 (+4.7) |
| **IR (KBr), ν<sub>max</sub>** | 3435, 2958, 2924, 2853, 1745, 1642, 1524, 1465, 1379, 1194, 1099, 1059 cm<sup>-1</sup> |
| **<sup>1</sup>H NMR (500MHz)** | Table 2 |
| **<sup>13</sup>C NMR (125MHz, CDCl<sub>3</sub>):** | 179.0, 174.0, 173.5, 173.17, 169.0, 168.8, 167.5, 167.5, 166.5, 166.3, 166.1, 165.9, 144.34, 145.93, 145.04, 140.5, 132.19, 130.3, 129.2, 127.8, 126.1, 113.6, 101.8, 76.7, 74.76, 74.67, 71.4, 71.2, 58.5, 57.2, 56.4, 54.9, 54.7, 54.3, 51.3, 29.6, 29.6, 29.3, 22.6, 21.7, 21.6, 19.2, 19.2, 19.09, 19.06, 18.8, 17.14, 14.11, 7.77. |
| **HRESI-MS** | *m/z* (pos.ions) 656.9243 [M+2H]<sup>2+</sup>, 1270.7069 [M+ H]<sup>+</sup>, 1291.8449 [M + Na]<sup>+</sup> 1307.9286 [M + K]<sup>+</sup><br>C<sub>62</sub>H<sub>84</sub>N<sub>12</sub>O<sub>17</sub>Na [M + Na]<sup>+</sup> calc. 1291.5975, found. 1291.8449 |
| **MALDI-TOF-MS** | *m/z* (pos.ions) 1293.61316 [M + Na+2H]<sup>+</sup>, 1309.93062 [M+K]<sup>+</sup><br>*m/z* (neg.ions) 1269.33344 [M-H]<sup>-</sup><br>C<sub>62</sub>H<sub>84</sub>N<sub>12</sub>O<sub>17</sub>Na [M+Na+2H]<sup>+</sup> calc. 1293.61950, found. 1291.61316 |
| **EI-MS: (70 ev)** | *m/z*(pos.ions) 1348.1437, 1291.4173 [M+Na]<sup>+</sup>, 1224.7363, 1191.8994, 1023.6241, 886.0243, 743.2058, 614.8185, 347.6111, 202.5464, 138.5079 |
| **LCESI-MS** | *m/z* (pos.ions) 1291.5995 [M + Na]<sup>+</sup> C<sub>62</sub>H<sub>84</sub>N<sub>12</sub>O<sub>17</sub>Na [M+Na]<sup>+</sup> calc. 1291.5975, found. 1291.5995 |
| **CHN** | Anal. calcd for C<sub>62</sub>H<sub>84</sub>N<sub>12</sub>O<sub>17</sub>: C, 58.66; H, 6.67; N, 13.24.<br>Found: C, 59.71; H, 7.28; N, 10.19. |

**Table 3.** Activity of transitmycin on biofilm formation.

| Culture | Zero days<br>10<sup>5</sup> x CFU/ml |  | After 4 days<br>10<sup>5</sup> x CFU/ml |  | Biofilm formation<br>after 5 weeks |
| --- | --- | --- | --- | --- | --- |
| *M. tuberculosis*<br>alone | 1.1 | 1.8 | 6.6 | 3.6 | Present |
| *M. tuberculosis*+<br>Transitmycin<br>(10 µg/ml) | 19.0 | 23.0 | 0.9 | 1.0 | 0.0 |

#### Anti-HIV activity

The symbiotic association of TB and HIV poses a challenge to human survival in which HIV complicates the treatment and diagnosis of TB. Besides, HIV–TB patients encounter other unique problems such as cumulative toxicity, immune reconstitution inflammatory syndrome (IRIS), drug-drug interactions, lower plasma drug levels, and the emergence of drug resistance during treatment [71]. The currently available therapy for treating patients co-infected with HIV and TB requires a very high pill load. Therefore, a class of drugs that can be used to treat TB and HIV would be a real breakthrough in TB and AIDS Research. Hence, the present study also evaluated the anti-HIV activity of transitmycin. A dose-dependent reduction was observed in HIV-1 p24 levels in viral culture indicating that transitmycin possessed significant anti-HIV activity. Transitmycin demonstrated good anti-viral activity against the different subtypes of HIV-1 as well as clinical isolates obtained directly from HIV-infected persons. In addition, transitmycin was also active against HIV-1 viruses resistant to nevirapine and AZT (Fig. 8a). The estimated IC_50_ value ranged between 0.19 and 0.65µg/ml for the viruses tested. There was a >50% inhibition at a concentration of 0.1µg/ml and 80-95% inhibition at a concentration of1µg/ml in the clinical isolates (Fig. 10b). The compound demonstrated a remarkable inhibitory effect on primary isolates belonging to various subtypes as well as to clinical isolates obtained from HIV-infected individuals. IC_50_ values calculated for the various strains tested indicate that transitmycin is a potent inhibitor of HIV-1 under *in vitro* experimental conditions. Importantly, transitmycin also inhibited drug resistant forms of the virus, in a dose-dependent manner. These findings suggest that transitmycin holds promise as the first potent compound that can be used to treat TB and HIV infections when they occur singly, as well as in combination as HIV/TB co-infection. (Supplementary S-table 13).

**Fig. 8.**
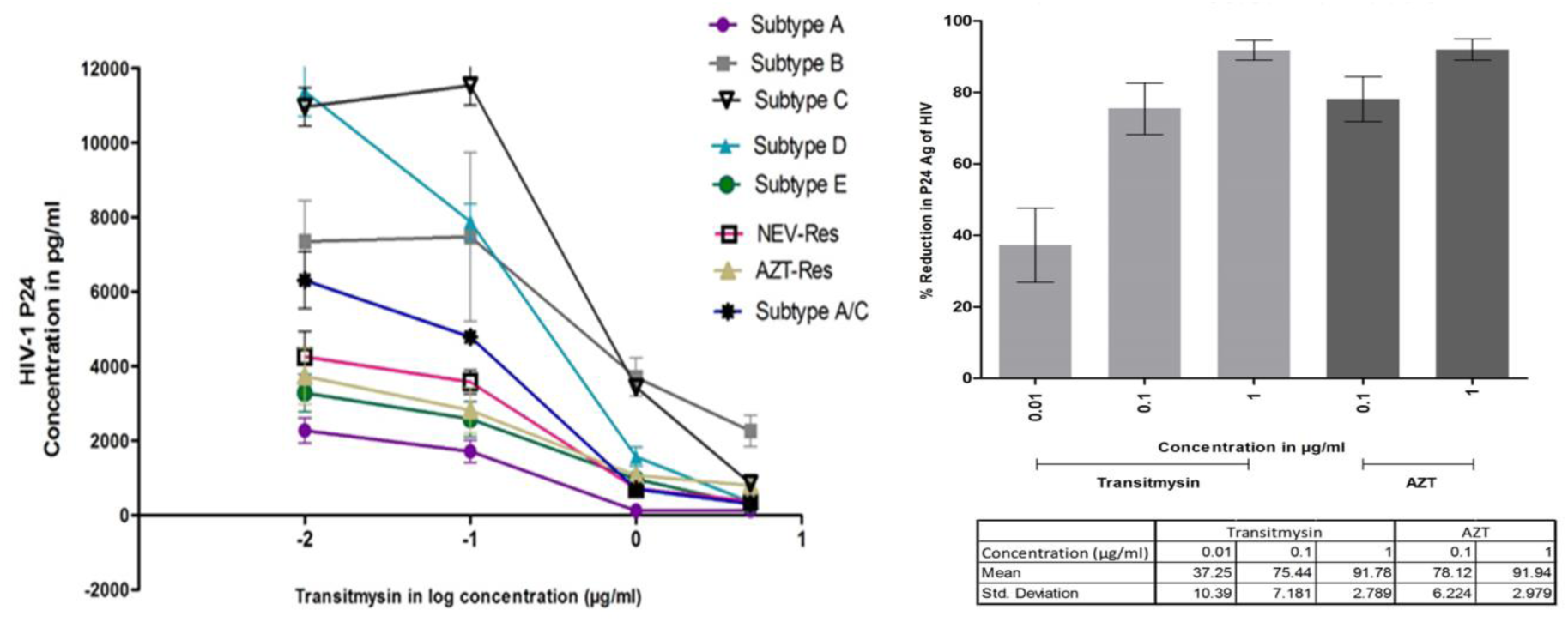
**a** Activity of Transitmycin against different HIV-1 clades. Various concentrations of Transitmycin were tested using 100 TCID_50_ of virus belonging to 6 different HIV-1 clades as well as two drug-resistant strains. Each concentration was tested in triplicate on all the indicated viruses and mean ± SEM values are shown in the graph. Log_10_ -2 in the figure equals 0.01 µg/ml, Log_10_ -1 equals 0.1 µg/ml, Log_10_ 0 equals 1 µg/ml and Log_10_ 1 equals 10 µg/ml. **b** Activity of Transitmycin on clinical isolates obtained from HIV-1 infected individuals. Three concentrations of Transitmycin and two concentrations of AZT were tested against 20 clinical isolates and mean ± SD values are shown on the graph. This experiment was performed on two different occasions.

### R1, R2 and R3 intercalates with the genomic DNA of Mycobacterium tuberculosis

Actinomycin D is the structurally similar compound for R1, R2 and R3. Actinomycin D and its derivatives are reported to intercalate with the DNA and exhibits fluorescence. So, we tested R1, R2 and R3 for its properties of DNA intercalation and fluorescence. Ethidium bromide 0.5 µg/ml is used as a positive control. The relative fluorescence unit (RFU) of R1, R2 and R3 with the DNA is compared with the RFU of R1, R2 and R3 without DNA. The average RFU of Ethidium bromide was 8561429. From the experiment, it was observed that all the three compounds, R1, R2 and R3 have DNA intercalating property and exhibits fluorescence (Fig. 9).

**Fig. 9.**
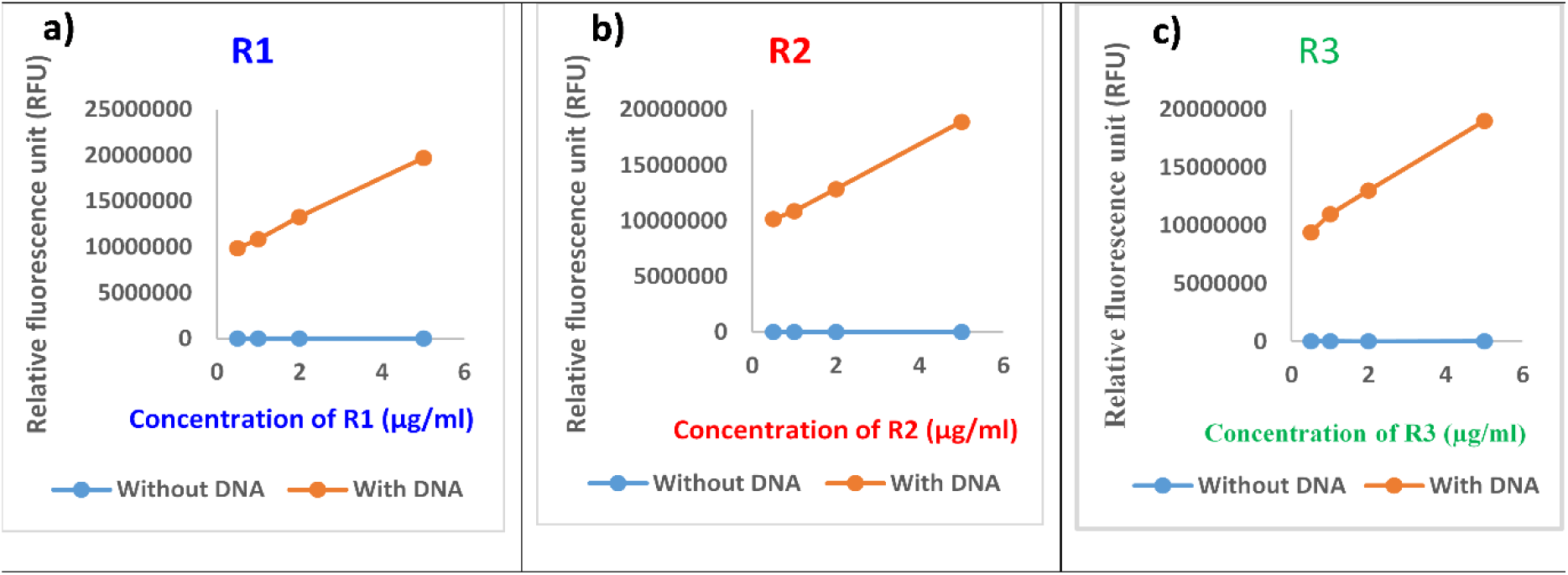
a) R1, b) R2 and c) R3 binds with the DNA and emit fluorescence in a dose dependent manner. The concentrations of R1, R2 and R3 tested were 0.5 µg/ml, 1 µg/ml, 2 µg/ml and 5 µg/ml. As the concentration increases, the Relative fluorescence unit (RFU) also increases.

### Docking and molecular dynamics studies

The concept behind docking was to assess the potential interaction between the transitmycin Fig. 10a or its derivative molecules and various DNA/Protein targets of interest. Through docking studies, we aimed to validate the anticipated interactions, even though comprehensive molecular dynamics studies were restricted, particularly for the entire transitmycin or its derivatives. Nevertheless, the chromophore part Fig. 10b molecular dynamics dynamics displayed enhanced stability, indicating promising outcomes in our research

**Fig 10.**
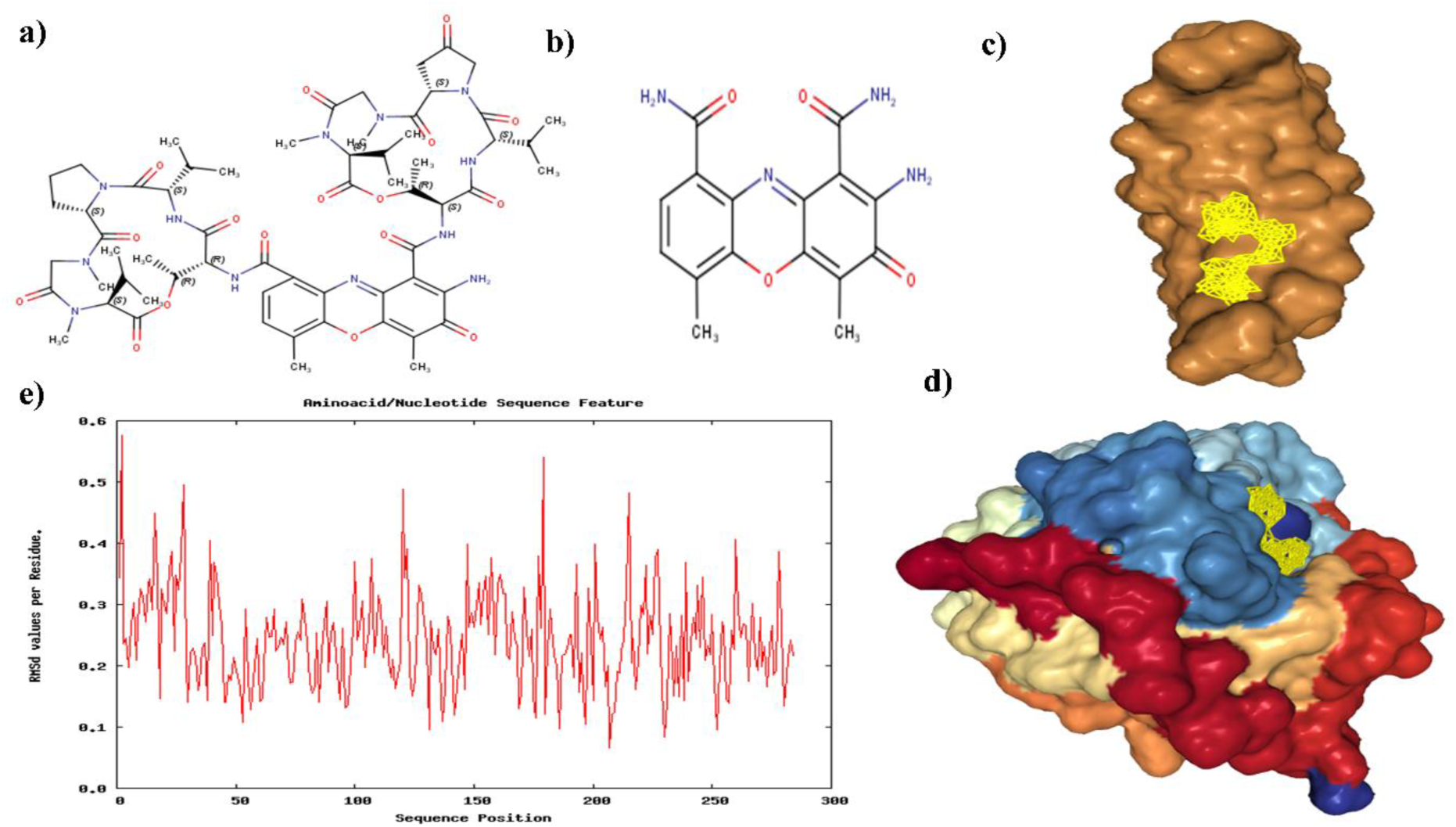
a) Transitmycin structure b) Chromophore part of transitmycin or its derivatives, c) 1MNV R2 Docking Pose d) 3PKE with R3 Docking pose e) RMSD values of Residues during the simulation Plot (3PKE with R3).

We performed docking and molecular docking experiments with a specific target and three ligands: R1, R2, and R3. Our analysis showed that 1MNV displayed favorable interactions with R2, whereas 3PKE exhibited superior binding with R3 Fig. 10c. The docking score data is presented in Table 4.

**Table 4.** The docking score data of specific target (1MNV, 3PKE, 1MO3, 3IU8) and three ligands: R1, R2, and R3.

| Name of the Molecule | Docking Score | Confidence Score | Ligand RMSD (A) |
| --- | --- | --- | --- |
| 1MNV with R1 | -489.71 | 0.9989 | 22.52 |
| 1MNV with R2 | -518.80 | 0.9994 | 17.61 |
| 1MNV with R3 | -532.55 | 0.9995 | 25.15 |
| 1MO3 with R1 | -397.33 | 0.9929 | 83.36 |
| 1MO3 with R2 | -424.84 | 0.9959 | 83.95 |
| 1MO3 with R3 | -420.99 | 0.9956 | 83.36 |
| 3PKE with R1 | -391.53 | 0.9921 | 52.95 |
| 3PKE with R2 | -493.89 | 0.9990 | 50.96 |
| 3PKE with R3 | -503.74 | 0.9992 | 57.70 |
| 3IU8 with R1 | -337.77 | 0.9771 | 37.59 |
| 3IU8 with R2 | -418.79 | 0.9954 | 47.58 |
| 3IU8 with R3 | -437.87 | 0.9969 | 50.75 |

Docking Score: docking scores were determined using the knowledge-based iterative scoring functions ITScorePP or ITScorePR. A more negative docking score implies a higher likelihood of a binding model. However, it’s important to note that the score doesn’t represent the true binding affinity, as it hasn’t been calibrated against experimental data.

Confidence Score: To gauge the likelihood of binding between two molecules, we introduced a confidence score based on docking scores. The formula for the confidence score is:

Confidence_score=1.01.0+e0.02×(Docking_Score+150)Confidence_score=1.0+e0.02×(Doc king_Score+150)1.0

In this context, when the confidence score surpasses 0.7, it suggests a high probability of binding. Scores between 0.5 and 0.7 indicate a possible binding, whereas scores below 0.5 imply an unlikely binding.

Ligand RMSD: Ligand RMSD values were computed by comparing ligands in the docking models with the input or modelled structures.

We conducted Molecular Dynamics (MD) simulations for the complex 1MNV with R2 and R3. Unfortunately, MD simulations for R2 and R3 couldn’t be performed due to the large size of their peptide side chains, preventing accommodation into the minor groove of DNA (1MNV). However, we successfully carried out simulations with the chromophores of R2 and R3. The RMSD of the residues remained within acceptable limits during these simulations.

Additionally, MD simulations were conducted for the complex 3PKE with R3. Similar challenges were encountered with the peptide side chains, leading to simulations being performed with the chromophore, as illustrated in Figure 10b. The RMSD values plotted against residues are presented in Figure 10e.

The results of the MD simulations indicate the stability of the protein backbone throughout the simulation period. However, it is noteworthy that the simulations faced computational challenges, primarily due to the size of the peptide side chains. Despite these difficulties, the obtained data provides valuable insights into the behavior of the complexes during the simulations.

## Methods

### Characterization and taxonomy of *Streptomyces* sp. R2

Micromorphology of potential *Streptomyces* sp. R2 was studied by adopting Transplantation Embedding Technique [72]. Briefly, a rectangular trough was dug out of an ISP2 agar plate using sterile knife. Then the spores of *Streptomyces* strain were inoculated on the edges of the trough under aseptic condition. A sterile cover-slip was placed over the agar, touching the inoculated area on the ISP2 agar plate. The plate was incubated at 28°C for 7-14 days. The cover-slip was aseptically removed using sterile forceps and placed over clean microscopic slide fixing the same using cellophane tape. Micromorphology of *Streptomyces* sp. R2 was observed under bright field microscope (Olympus) under 10x and 40x magnifications. Spore structure and spore surface morphology were recorded using a scanning electron microscope (JEOL model JSM5600LV). Media and procedures used for determination of cultural characteristics and carbon and nitrogen source utilization were those described originally by Shirling and Gottileb [24]. Effect of pH, temperature, NaCl concentration and anaerobic condition were studied using modified ISP2 medium. Antibiotic susceptibility pattern was determined by disc diffusion method using standard antibiotic discs (Hi media) following the standard protocol [73]. Biomass for cell wall analysis was prepared by growing *Streptomyces* sp. R2 in shake flasks (120 rpm) containing ISP2 broth at 28 ^0^C for 5 days. Amino acid and sugar content analyses of whole cell hydrolysates were performed according to the original procedure described by Staneck and Roberts [74].

### Molecular characterization and phylogenetic analysis

*Streptomyces* sp. R2 was grown in 50 ml of ISP2 broth at 28^°^C for 48 h. The genomic DNA was extracted using Chromous genomic DNA isolation kit. Polymerase chain reaction (PCR) was performed for the amplification of 16S rRNA gene using the primers 5’ – AGAGTRTGATCMTYGCTWAC – 3’ and 5’ – CGYTAMCTTWTTACGRCT – 3’ on a ABI12720 thermal cycler (Applied Biosystems). The conditions used for thermal cycling were as follows: initial denaturation at 94^0^C for 4 min, followed by 35 cycles consisting of denaturation at 94^0^C for 30 sec, primer annealing at 55^0^C for 30 sec and primer extension at 72^0^C for 2 min, followed by a final extension at 72^0^C for 5 min. The amplified 16s rRNA gene fragment (∼ 1.4 kb) was separated by agarose gel electrophoresis and the purified fragment was used for sequencing in an ABI3130 genetic analyser. The nearly complete 16s rRNA gene sequence of strain R2 (1400 nt) was subjected to BLAST comparison against the 16s rRNA sequences given in GenBank/DDBJ/EMBL databases.

Phylogenetic analysis was performed using CLUSTAL-W and MEGA version 3.1. Evolutionary distances (Kimura’s two parameter model) [75] and clustering were calculated employing the neighbour-joining method. The topology of phylogenetic tree was evaluated by the bootstrap re-sampling method with 1000 replicates.

### Production of Transitmycin

Hundred microliters of *Streptomyces* sp. R2 spore suspension was transferred into 10 YEME agar plates and spread using sterile L-rods. The plates were incubated at 28^°^C for 10 days. After every 24 hours of fermentation, the mycelial growth was scrapped out and the crude pigment secreted into the agar medium was extracted using equal volume (1:1) of different organic solvents such as n-hexane, dichloromethane, chloroform, ethyl acetate and methanol for 24 hours. The solvent portion was collected and dried at 40^°^C using Concentrator plus (Eppendorf) [76]. Anti TB activity of crude extracts were tested against *M. tuberculosis* H37Rv at 100µg concentration adopting LRP assay [77].

*Streptomyces* sp. R2 was cultured for 10 days at 28^0^C on YEME agar plates (2000 ml of medium in 100 petriplates of 90 mm diameter) to produce the culture extract in bulk. After the incubation period, the cell material was aseptically removed and discarded after autoclaving. The yellow pigmented antibiotic containing agar medium was cut in to pieces and extracted twice with equal volume (1:1 ratio) of ethyl acetate for 24 hours.

### Purification of transitmycin

Transitmycin was purified by preparative thin layer chromatography (TLC) using Merck silica gel 60 (GF254) pre coated aluminium (6x8 cm size) plates. The extract was separated using different solvents in different proportions. After running, the 200 sheets were kept at room temperature for complete drying of the plate. Spots on TLC were detected through naked eye as well as under UV light (254 and 365 nm). After drying, three major yellow colour spots (R1, R2 and R3) were scrapped, mixed with ethyl acetate and filtered using a funnel fitted with Whatman filter paper. Ethyl acetate was evaporated to dryness under vacuum to obtain the compounds as dry amorphous powder. (Supplementary Fig. S2).

All three compounds were tested against *M. tuberculosis* H37Rv at 100 µg concentration by LRP assay. All three compounds from the ethyl acetate extract was purified using column chromatography packed with neutral alumina using a gradient of 1% methanol/chloroform mixture (CH_3_OH/ CHCl_3_) as the eluent. Fractions were collected and concentrated under vacuum to obtain pure transitmycin. The product was visualized in a silica gel coated TLC sheet (Supplementary Fig. S2 and S3).

The compound R1 (named as transitmycin) that showed maximum activity was taken for characterization and other studies testing its biological activity. Purity of transitmycin (Fig. 2c). was analysed by HPLC using Shimadzu (Japan) RID-10A gradient high-performance liquid chromatographic instrument, equipped with two LC-20AD pumps controlled by a CBM-10 inter-face module. Refractive index Detector RID 10A (Shimadzu) was used for the peak. Analysis was performed on a Luna 5u C_18_ (2) reversed-phase column, 100 (150X4.6mm). The analytical parameters were selected after screening a number of solvent systems and gradient profiles. Separation was achieved using a two-pump gradient program for pump A (0.1% Acetic acid in CH_3_CN) and pump B (0.1% Acetic acid in H_2_O) in a linear gradient of acetonitrile and water from 0:100 to 65:35 in 65 minutes at a flow rate of 2 ml/min. Detection was done at 254 nm, the absorption maxima close to that of majority of the compounds. Injection size for sample was 20 µl. Column temperature was 30°C. (Supplementary Fig. S4 and S5).

### Characterization and structure elucidation

Colour and consistency of the purified antibiotic was visually observed. Solubility was tested in water, methanol, acetone, ethyl acetate, diethyl ether, dichloromethane, chloroform, and n-hexane by dissolving 1 mg of purified antibiotic. Optical rotations were measured with a Autopol IV Automatic polarimeter, and the [α]_D_ values are given in deg cm^2^ g^-1^. Melting point was analysed using Mettler Toledo Model FP62 [78]. Ultraviolet (UV) spectrum was determined using Shimadzu UV-1700 series. One milligram of sample was dissolved in 10ml of methanol and the spectra were recorded at wavelength between 190 – 900 nm. The Infrared (IR) spectrum of the purified antibiotic was determined on Perklin Elmer Spectrum One FT-IR. The spectrum was obtained using potassium bromide (KBr) pellet technique in the range of 450 to 4000 cm^-1^ at a resolution of 1.0 cm^-1^. Potassium bromide (AR grade) was dried under vacuum at 100°C and 100 mg of KBr with 1mg of purified antibiotic was used to prepare KBr pellet. The spectrum was plotted as intensity versus wave number [79]. ^1^H and ^13^C NMR spectra were recorded on a Bruker Advance 500 NMR spectrometer in CDCl_3_ with TMS as internal Standard and with chemical shifts (*δ*) reported in ppm. Two-dimensional ^1^H–^1^H COSY, DQF-COSY, NOESY, ROESY, ^1^H–^13^C HSQC, HMBC, and spectra were recorded on a Bruker Advance 500 NMR spectrometer. MALDI-TOF MS analyses were performed using an Applied Biosystems ABI4700 TOF mass spectrometer in reflector mode with an accelerating voltage of 20 kV. HRESIMS were measured on a Q-TOF micro mass spectrometer (Waters USA) in positive ion mode with methanol as solvent. QTOF-MS was recorded on an Agilent 6520-QTOF LCMS having an ESI source in positive mode.

### HPLC Analysis of L-FDAA Derivatives of Transitmycin

Transitmycin (3.0 mg) was dissolved in 1 ml of 6NHCl and heated in a sealed glass tube at 110^°^C for 24 h. After removing the solvents, the hydrolysate mixture (3 mg) and the amino acid standards (0.5 mg) were separately dissolved in 0.1 mL of water and treated with 0.2 mL of 1% 1-fluoro-2,4-dinitrophenyl-5-L-alaninamide (FDAA) (Marfey’s reagent) in acetone (10 mg/mL in acetone) and 0.04 mL of 1.0 M sodium bicarbonate. The vials were heated at 50^°^C for 90 min, and the contents after cooling at room temperature were neutralized with 1N HCl. After degassing, an aliquot of the FDAA derivative was diluted in CH_3_CN, Water (1:1) and analysed by reversed phase HPLC column Luna 5u C_18_ (2) 100 (150X4.6mm) and a linear gradient of acetonitrile and water containing 0.05% trifluoroacetic acid from 10:90 to 50:50 in 20 min and then isocratic. The flow rate was adjusted to 1 mL/min and the absorbance detection was at 340 nm. The chromatogram was compared with those of amino acid standards treated in the same conditions [60–61].

### LC-MS Analysis of L-FDAA Derivatives of Transitmycin

The analysis of the L- and D-FDLA derivatives of Transitmycin was performed on a Waters Acquity UPLC coupled with a Thermo LCQ Deca XP MAX. QTOF-MS was recorded on an Agilent 6520-QTOF LCMS having a ESI source in Positive mode and employing a linear gradient of from 25% to 70% CH_3_CN in 0.01 M formic acid at 0.5 mL/min over 60 min [62–64].

#### Transitmycin (R1)

R1 was obtained as an orange red solid; [α]_D_^25^ -106 ° (c 0.2, MeOH); UV (MeOH) λ _max_ (log ε) 214 (3.07), 240 (2.30), 425 (1.44), 442 (1.51) nm; CD [(MeOH),(mdeg)] λ_max_(Δε) 195 (+11.1), 210 (-21.0), 242 (+4.7) nm; IR (KBr), *ν*_max_ 3435, 2958, 2924, 2853, 1745, 1642, 1524, 1465, 1379, 1194, 1099, 1059 cm^-1^; HRESI-MS (pos.ions): *m/z* 1270.7069 [M+2H]^+^,1291.8449 [M+Na]^+^, 1307.9286 [M+K]^+^657.3119 [M+2H]^+2^; MALDI-TOF-MS (pos.ions):*m/z* 1293.07095 [M+Na+2H]^+^, 1309.93062 [M+K]^+^ MALDI-TOF-MS (neg.ions) 1269.33344 [M-H]^-1^HNMR, ^13^NMR and 2DNMR details (Supplementary Fig. S14-52, S-Table 5).

#### Compound (R2)

R2 was obtained as a red solid; [α] ^25^: -24° (c 0.2, MeOH); UV(MeOH) λ_max,_ (log ε) 205(1.25),240(0.63),425 (0.39), 442 (0.42) nm; CD [MeOH, (mdeg)] λ_max,_ (Δε) 195 (+8.8), 210 (-22.0), 240 (+4.3) nm;IR (KBr), *ν*_max_3436, 2961, 2924, 2853, 1744, 1650, 1565, 1415, 1204, 1140, 1045, 1019 cm^-1^; HRESI-MS (pos. ions) :1277.8245 [M + Na]^+^1293.8735 [M + K]^+^*m/z* 650.3413 [M+2H]^+2^; MALDI-TOF-MS *m/z* (pos.ions) 1278.95175 [M+Na+H]^+^; MALDI-TOF-MS (neg.ions) *m/z* 1255.38052 [M-H]^-^, ^1^HNMR, ^13^NMR and 2D NMR details (Supplementary Fig. S53-63).

#### Compound (R3)

R3 was obtained as orange solid; [α] ^25^:-27 ° (c 0.2, MeOH); UV (MeOH) λ _max_ (log ε) 206 (1.90), 240 (0.69) 424 (0.191), 442.2 (0.19) nm; CD [MeOH, (mdeg)] λ_max_ (Δε) 195 (+24.0), 210 (-21.5), 241 (+1.7) nm; IR (KBr), *ν*_max_ 3415, 2957, 2924, 2853, 1745, 1642, 1583, 1464, 1384, 1193, 1093, 1078 cm^-1^; HRESI-MS (pos.ions):1271.7159 [M +H]^+^ *m/z*1277.6149 [M-OH+Na]^+^; MALDI-TOF-MS (pos.ions) *m/z* 1294.5888 [M + Na]^+^ MALDI-TOF-MS (neg. ions) *m/z* 1268.48582 [M-H]^-^, ^1^HNMR,^13C^NMR and 2D NMR details, (Supplementary Fig. S64-69).

### Determination of anti TB and anti-HIV activity

Anti-TB activity was determined by adopting Luciferase Reporter Phage (LRP) assay against the standard laboratory strain, *Mycobacterium tuberculosis* H_37_Rv, and 97 clinical *M. tuberculosis* isolates including drug sensitive and cultures exhibiting different drug resistant patterns. Different concentrations (5 – 50 µg/ml) of purified antibiotic – transitmycin were prepared using 10% dimethyl sulfoxide (DMSO). About 50µl of antibiotic solution was added to 350 µl of glycerol 7H9 broth in cryo vials. Effect of DMSO was also tested by adding 50 µl of 10% DMSO instead of the antibiotic. Mycobacterial cell suspension equivalent of 2 McFarland units was prepared from log phase culture and 100 µl of the same was added to all the vials before incubating at 37°C for 72 hours. After incubation, 50 µl of high titre luciferase reporter phage phAE129 and 40 µl of 0.1M CaCl2 were added to test and control vials. All the vials were incubated at 37°C for 4 hours. After incubation 100 µl of suspension from each vial was transferred to a luminometer cuvette. 100 µL of D-luciferin was added and relative light unit (RLU) was measured in a luminometer [77]. Percentage of inhibition was calculated using the RLU of control and test and MIC was determined. [81,82]

### Activity against *M. tuberculosis* biofilm

Cell suspensions of *M. <u>tuberculosis</u>* H37Rv were prepared using 7H9 broth. Biofilms of *M. tuberculosis* were developed on 6 well tissue culture plates by adding 2 ml of Sautons medium (without Tween 80) and inoculating 20 μL of saturated planktonic culture of *M. tuberculosis* H_37_Rv. The plate was wrapped with parafilm and incubated without shaking at 37°C in humidified conditions for 7 to 14 days. The plate was observed regularly after 7 days for biofilm formation by *M. <u>tuberculosis</u>* which can be visibly seen. When the biofilm was formed, 10 μg/ml of transitmycin was added to the transitmycin test wells leaving the controls and considered as zero day. The viable counts of tubercle bacilli were determined from the wells on zero day, 4^th^ day and after 5 weeks and expressed in cfu/ml [80].

### Anti-HIV activity

**Viruses:** HIV-1 viruses belonging to subtypes A, B, C, D, E and A/C (Subtype A: 92RW020, Subtype B: JR-FL, Subtype C: 92BR025, Subtype D: 92UG001, Subtype E: 92TH021 and Subtype A/C: 92RW009), were obtained from the NIH AIDS Repository (Germantown, MD, USA). Clinical isolates of HIV-1 were produced in our laboratory by co-culture of HIV-infected peripheral blood mononuclear cells (PBMC) with activated donor PBMC.

### Testing for anti-HIV activity

Anti-HIV activity of Transitmycin was determined using the HIV-1 gag p24 inhibition assay. Initially, testing was performed on a lab-adapted HIV-1 subtype B isolate, HIV-1 IIIB. Donor PBMC were obtained from healthy volunteers after obtaining the approval of the Institutional Ethics Committee of the National Institute for Research in Tuberculosis as well as the informed consent of the participants. Donor PBMC were stimulated with PHA (Phytohemeagglutinin) for 72 hours and incubated with 100 TCID_50_ of the virus per 1 x 10^6^ cells for 4 h at 37°C. The cells were washed to remove the un-adsorbed virus and plated at a concentration of 10,000 cells/well in a 96-well tissue culture plate. Varying concentrations of Transitmycin (0.01µg/ml, 0.1µg/ml, 1.0µg/ml and 5.0µg/ml) were added to triplicate wells. Control cultures were set up without the addition of the compound. AZT was used as the reference compound. Cultures were maintained for 7 days at 37°C in a CO_2_ incubator. On day 7, HIV-1 gag p24 antigen production was determined as an indirect measure of viral replication in the culture supernatants using the Alliance HIV-1 p24 ELISA kit (Perkin Elmer, USA).

### Testing of Transitmycin against different HIV-1 subtypes

Anti-viral activity of Transitmycin was also tested on primary HIV-1 isolates belonging to different subtypes - Subtype A: 92RW020, Subtype B: JR-FL, Subtype C: 92BR025, Subtype D: 92UG001, Subtype E: 92TH021 and Subtype A/C: 92RW009, as well as a Nevirapine resistant and AZT resistant strain, using the method described above. The activity of Transitmycin on different viruses was determined by measuring HIV-1 p24 antigen in 7-day culture supernatants. The IC_50_value (concentration of compound required to inhibit 50% of virus replication) of Transitmysin for the different HIV-1 subtypes was calculated by fitting a dose response curve using a non-linear regression analysis to generate a sigmoidal three parameter dose response curve (GraphPad Prism, version 6).

### Activity against clinical strains of HIV

Anti-viral activity of Transitmycin was further evaluated on 20 clinical isolates of HIV-1 obtained by co-culture of patient PBMC with PHA-stimulated donor PBMC in the laboratory. For this analysis, only three concentrations (0.01 µg/ml, 0.1 µg/ml and 1µg/ml) of transitmycin was used based on the results of the above experiment.

### DNA binding and Fluorescence assay

0.5 µg/ml, 1 µg/ml, 2 µg/ml and 5 µg/ml of R1, R2 and R3 were separately added to 100 µl of PBS in triplicates in 96 well plate. *Mycobacterium tuberculosis* genomic DNA was extracted and 200 ng of DNA is added to each test wells. DNA was not added to the control wells of R1, R2 and R3. For the positive control, 0.5 µg/ml of Ethidium bromide is taken in PBS and 200 ng of DNA is added to it. After 15 mins, the plate is read in Multimode - plate reader, Spinco Biotech, with excitation 546 nm and emission 595 nm. The relative fluorescence unit is recorded. Average of RFU for each concentration of R1, R2 and R3 was taken and plotted as graph.

## Conclusion

Globally, the general public has latent or active tuberculosis, and this is expected to increase in the future, causing the medical system a major challenge. The need of the hour is to identify novel antibiotics to support the current regimens with reduced side effects, duration and cost of treatment. Transitmycin is a novel antibiotic isolated from a novel *Streptomyces* sp. MTCC from the coral reef soil from Rameshwaram waters in India. It has the unique property of killing latent and active forms of TB bacilli irrespective of the resistance they have towards major antiTB drugs and also sterilising the different resistant and recombinant clades of standard and clinical HIV virus. When brought into the market for human use, transitmycin can strategize treating TB and HIV simultaneously, which could be a major breakthrough, none the less. Of course, preclinical trials and toxicity studies need to be performed before it can be tested on human volunteers which will require sufficient resources. Countries which are burdened with this dual ailment may have to take considerable interest in this direction. Structural medications to the parent compound can further improve its activity.

## Data Availability

SUPPORTINg information

## Acknowledgement

Professor Balasubramanian Kalpattu Kuppusami, INSA Senior Scientist, Department of Chemistry, IIT Madras Chennai 600 036, India and Professor Krishna Kumari Gadepalli Narasi, Department of Medicinal Chemistry, Sri Ramachandra University, Chennai 600 116, India, acknowledged for discussion of structure elucidation of Transitmycin. Technical assistance of Dr. S. Balaji and Dr. A. S. Shainaba of National Institute for Research in Tuberculosis is acknowledged in fine tuning and uploading the manuscript.

## Supplementary materials

The comprehensive characterization of the anticipated products have been subjected through UV/Vis, IR, CD, CHNS analyses, whilst, followed by ^1^H-NMR ^13^C-NMR, ^1^H-^1^H COSY, ^1^H−^1^H DQF-COSY, ^1^H−^13^C HMBC, and ^1^H−^13^C HSQC, ^1^H−^1^H TOCSY, ^1^H−^1^H NOESY, ^1^H−^1^H NOESY 2D NMR spectra and MALDI-TOF-MS, HR-ESIMS, HR-LCMS, ESI-MS, QTRAP LC-MS/MS, RP-HPLC and LCMS analyses respectively. The following L-FDAA derivatives of R1, R2, R3 spectral information on all compounds of this article can be found in the online version.

## Funding Received for this study

1. Potential Tuberculosis Drugs from Marine Actinomycetes, Funded by Department of Science and Technology, New Delhi (November, 2007 to November, 2009).
2. Study to evaluate the baseline anti TB and anti-HIV properties of novel antibiotic transitmycin (Tr) isolated from novel *Streptomyces* Sp. R2 funded by Indian Council of medical Research (2009-2012).
3. Purity and in vitro efficacy studies on transitmycin funded by Indian Council of Medical Research (2015-2016).

## Authors’ individual contributions

Vanaja Kumar – Principal Investigator, planning, supervision, coordination and data compilation, manuscript writing and editing.

Balagurunthan Ramasamy–Facilitating Growing of producer strain, coordination in crude extract preparation as lab head.

Mukesh Doble – Purification of compound from crude extract, structure analysis and chemical characterisation and report preparation as lab head.

Gandarvakottai Senthilkumar Arumugam – Isolation, purification, characterization, complete structural elucidation and a key role in Innovation Of Transitmycin drug discovery development and manuscript writing and editing.

Kannan Damodharan - Isolation, purification, characterization, structural elucidation of Transitmycin compounds and final manuscript drafting.

Radhakrishnan Manikkam-Isolation and characterisation of producer strain, biological and biochemical characterisation, testing biological activities, data compilation, manuscript writing.

Hanna Luke Elizabeth - Testing crude extract and purified compound on HIV standard and clinical clades. Preparation of write up.

Suresh Ganesan – Isolation, purification, characterization, structural elucidation of Transitmycin compounds and final manuscript drafting

Azger Dusthackeer- Testing biological activities against latent bacilli and biofilms, preparing the write up.

Precilla Lucia – Laboratory work for testing the compound on HIV clades.

Shainaba A Saadhali- Testing biological activities against latent bacilli and biofilms, preparing the write up.

Shanthi John – laboratory work with respect to growing the producer strain and preparation of crude extract.

Poongothai Eswaran - laboratory work with respect to growing the producer strain and preparation of crude extract.

Selvakumar Nagamiah– supervision of laboratory work on testing biological activities, editing manuscript.

Soumya Swaminathan – Facilitating coordinated activity for fund release, conduct of experiments against HIV clades as Head of the laboratory and the Institute.

Jaleel UCA-Provided the overall conceptualization of theoretical study, and offered guidance throughout the research process and reviewed and edited the manuscript to ensure clarity, consistency, and scientific accuracy.

Rakhila M-Conducted the data analysis, molecular docking simulations, and statistical interpretation of the results and edited the manuscript to ensure clarity, consistency, and scientific accuracy.

Ayisha Safeeda-Conducted the data analysis, molecular docking simulations, and statistical interpretation of the results and edited the manuscript to ensure clarity, consistency, and scientific accuracy.

Satheesh S -was responsible for creating and editing the figures and tables that visually represented the key findings.

## Additional Information

### Competing Interests

The authors declare no competing interests.

## Supplementary Information

**S-Table 1.**
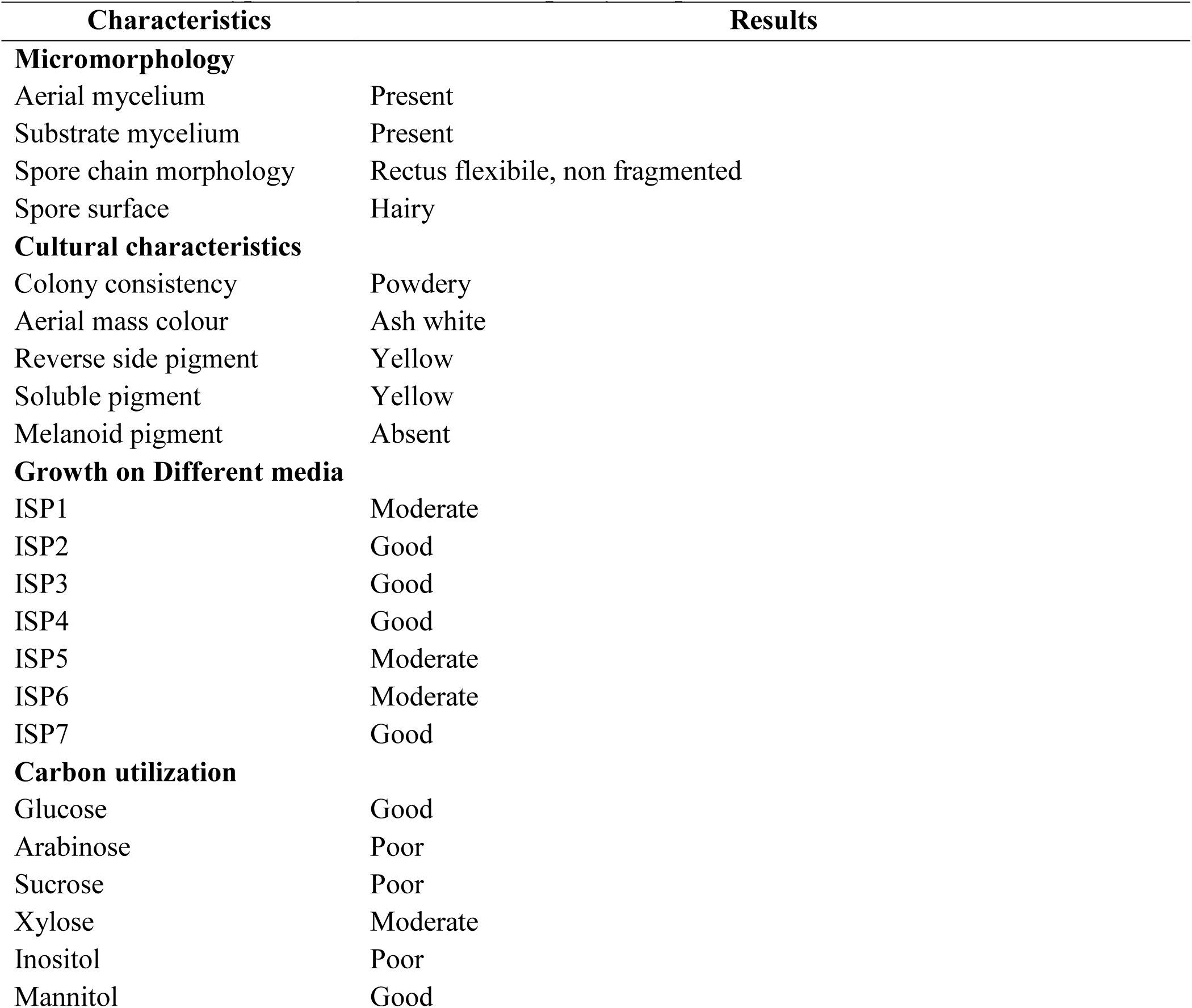
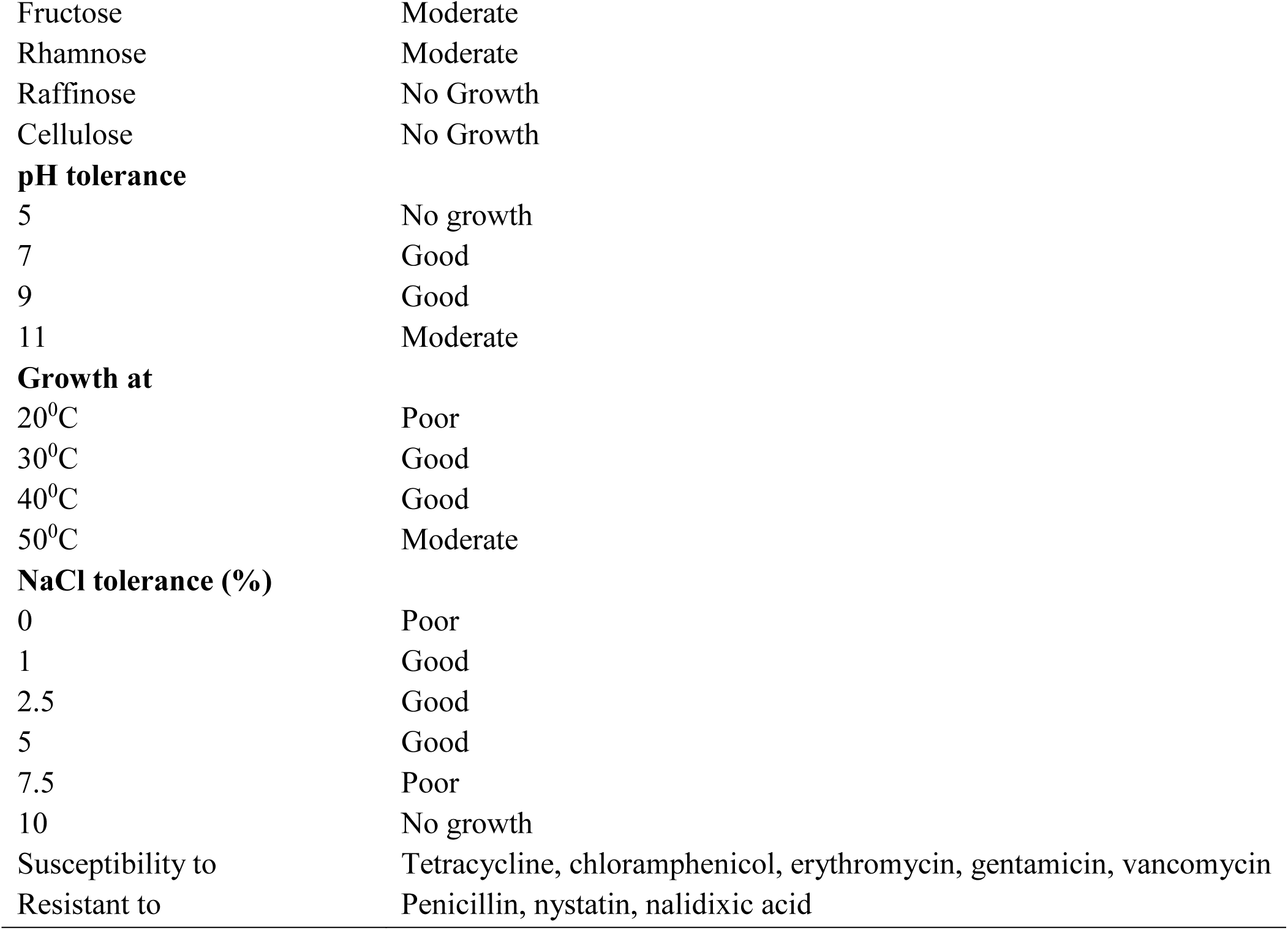
Phenotypic characteristics of *Streptomyces* sp. R2.

| Characteristics |  | Results |
| --- | --- | --- |
| **Micromorphology** |  |  |
| Aerial mycelium |  | Present |
| Substrate mycelium |  | Present |
| Spore chain morphology |  | Rectus flexibile, non fragmented |
| Spore surface |  | Hairy |
| **Cultural characteristics** |  |  |
| Colony consistency |  | Powdery |
| Aerial mass colour |  | Ash white |
| Reverse side pigment |  | Yellow |
| Soluble pigment |  | Yellow |
| Melanoid pigment |  | Absent |
| **Growth on Different media** |  |  |
| ISP1 |  | Moderate |
| ISP2 |  | Good |
| ISP3 |  | Good |
| ISP4 |  | Good |
| ISP5 |  | Moderate |
| ISP6 |  | Moderate |
| ISP7 |  | Good |
| **Carbon utilization** |  |  |
| Glucose |  | Good |
| Arabinose |  | Poor |
| Sucrose |  | Poor |
| Xylose |  | Moderate |
| Inositol |  | Poor |
| Mannitol |  | Good |

|  |  |
| --- | --- |
| Fructose | Moderate |
| Rhamnose | Moderate |
| Raffinose | No Growth |
| Cellulose | No Growth |
| **pH tolerance** |  |
| 5 | No growth |
| 7 | Good |
| 9 | Good |
| 11 | Moderate |
| **Growth at** |  |
| 20<sup>0</sup>C | Poor |
| 30<sup>0</sup>C | Good |
| 40<sup>0</sup>C | Good |
| 50<sup>0</sup>C | Moderate |
| **NaCl tolerance (%)** |  |
| 0 | Poor |
| 1 | Good |
| 2.5 | Good |
| 5 | Good |
| 7.5 | Poor |
| 10 | No growth |
| Susceptibility to | Tetracycline, chloramphenicol, erythromycin, gentamicin, vancomycin |
| Resistant to | Penicillin, nystatin, nalidixic acid |

**S-Table 2.** Effect of solvents on the extraction of Transitmycin.

| **Solvents** | **Quantity of extracts** | **Antibacterial activity against *S. aureus* MTCC 96** | **Anti TB activity (% reduction in RLU)** |
| --- | --- | --- | --- |
| **Methanol** | **40** | **14** | **58.31** |
| **Chloroform** | **41** | **9** | **18.07** |
| **Dichloromethane** | **40** | **13** | **22.71** |
| **Diethyl ether** | **10** | **14** | **74.23** |
| **Ethyl acetate** | **9** | **19** | **83.40** |
| **n-hexane** | **-** | **-** | **-** |

**Fig. S1.**
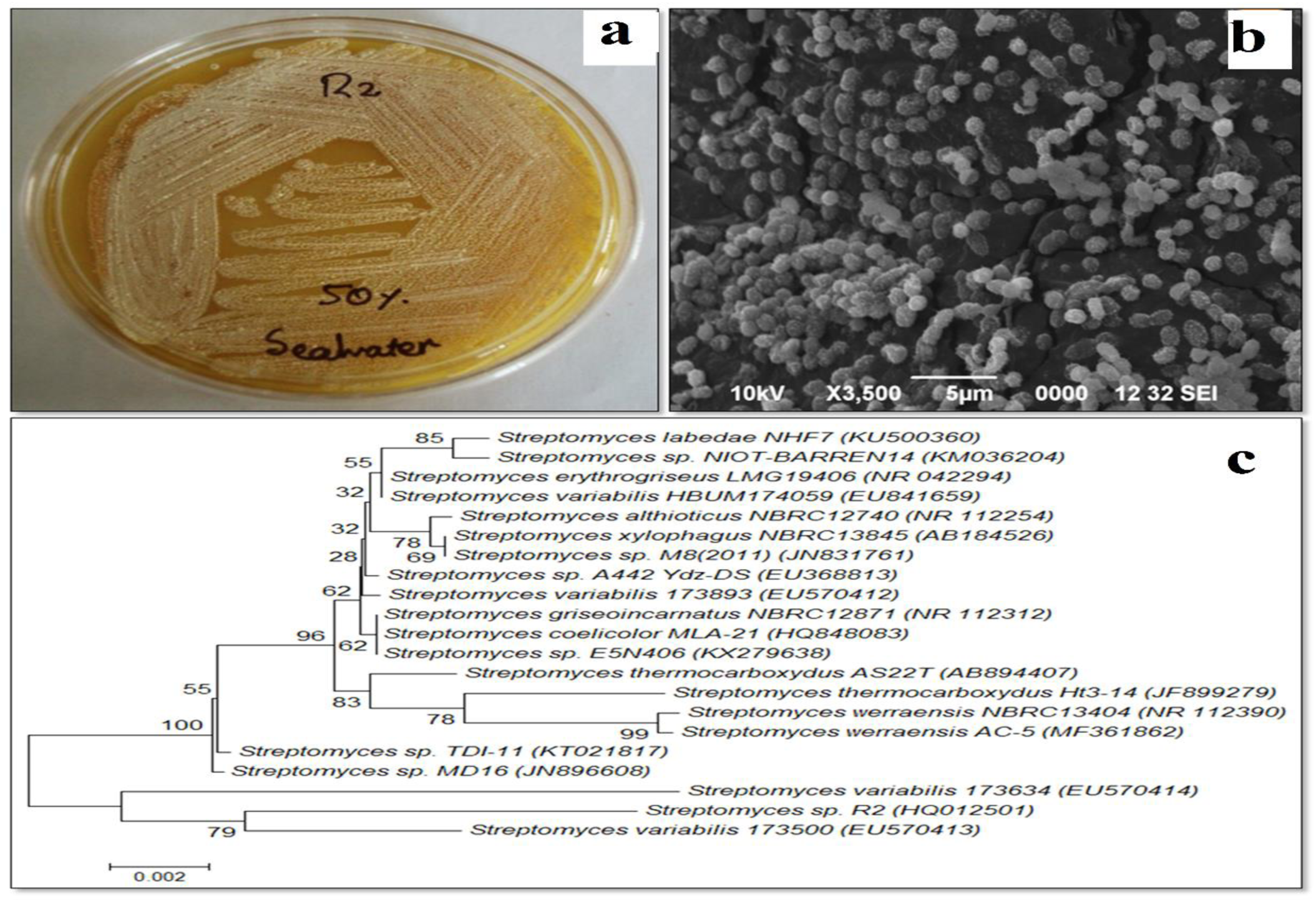
Characteristic microbial strain and its morphological arrangement. **a** Cultural morphology and microscopic visualization. **b** SEM image (3,500X) of Streptomyces variabilis R2 on ISP2 agar medium. **c** Phylogenetic dendrogram obtained by distance matrix analysis of 16s rRNA gene sequences, showing the position of strain R2 among its phylogenetic neighbours.

**Figure S2.**
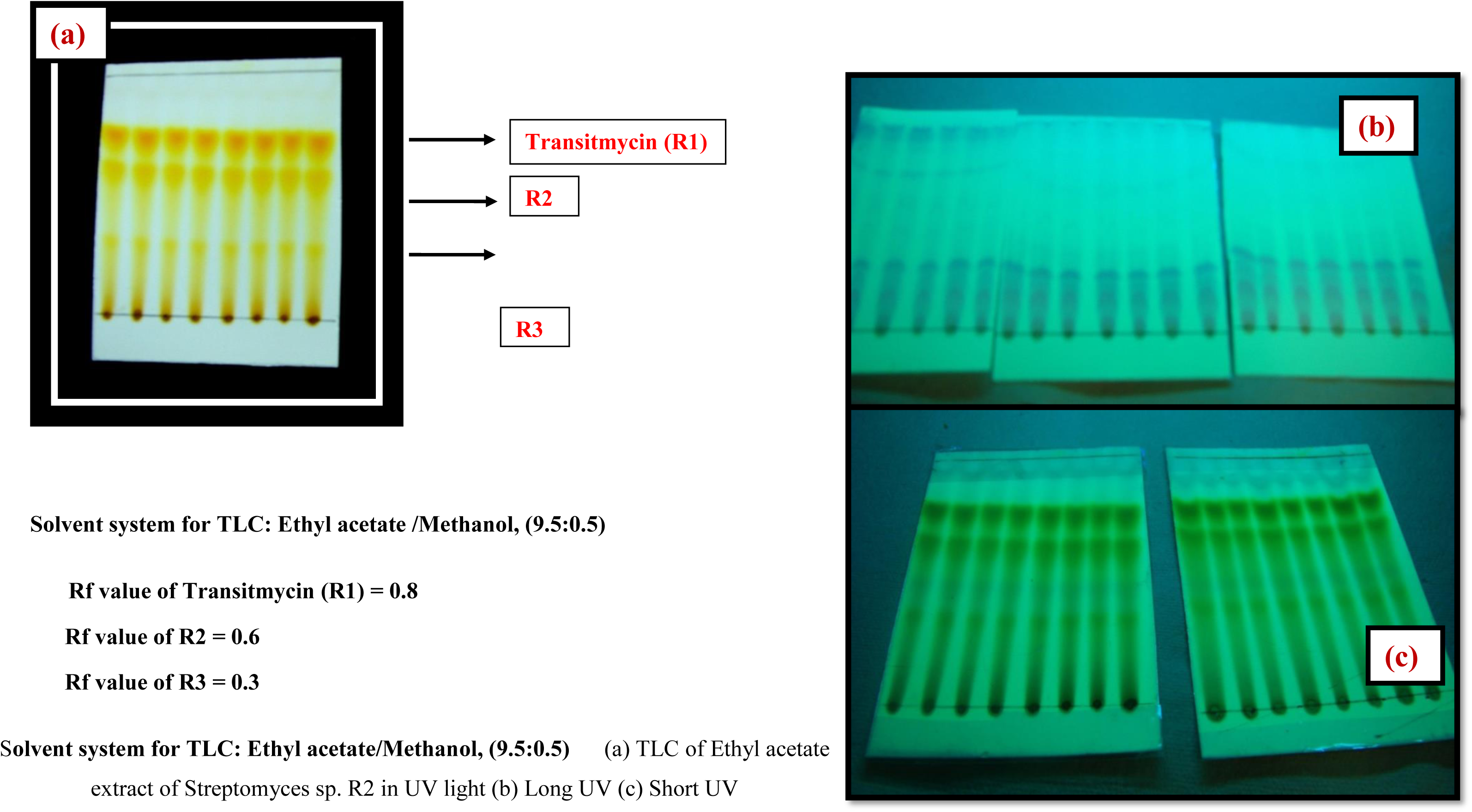
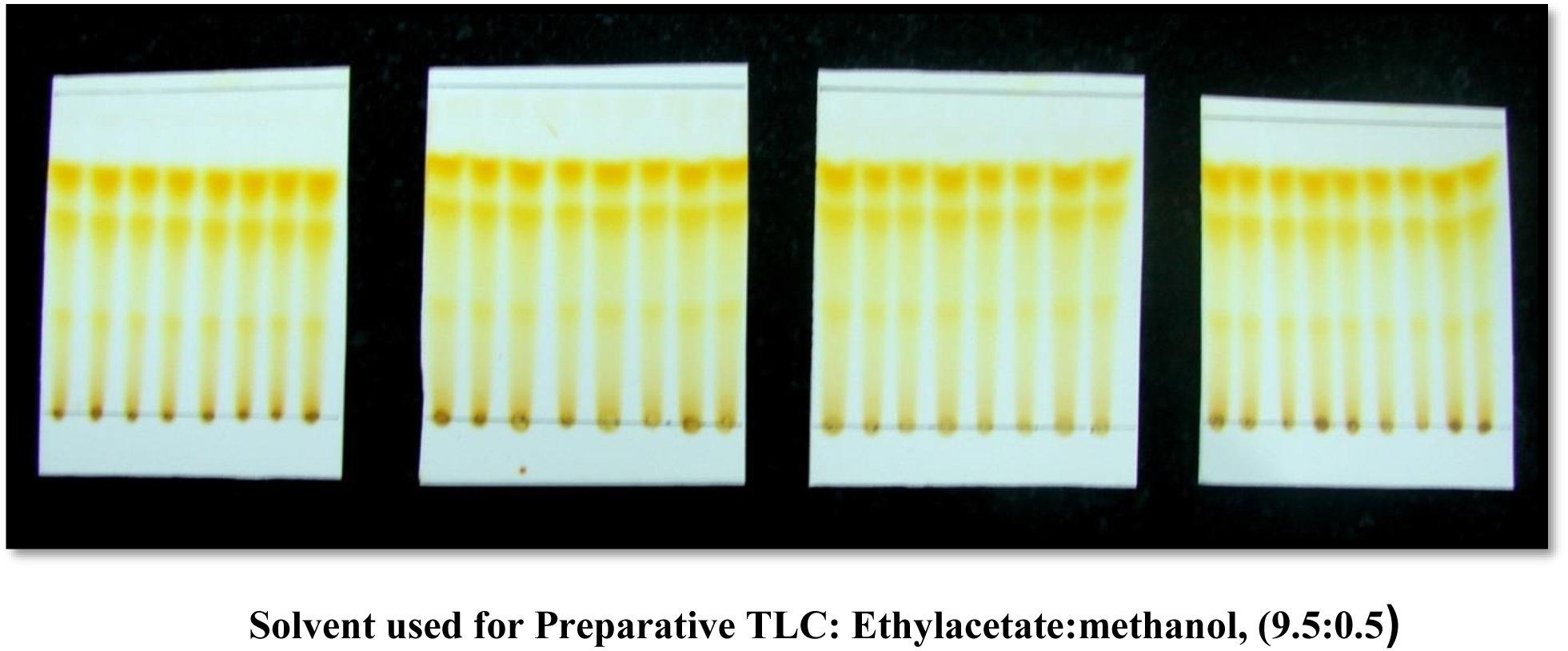
Purification of EA extract of *Streptomyces* sp. R2 by Preparative Thin-Layer Chromatography Extraction and Isolation. Purification of compounds were performed by preparative thin layer chromatography (TLC) using Merck silica gel 60 (GF254) pre coated aluminium (6x8 cm size) plates.The crude pigment was purified by using preparative thin layer chromatography commercially available pre coated silica gel chromatography sheets (6×8 cm size) were used. To find out the best solvent system to separate the crude compound, the solvents were used in different proportions, among all solvent systems used, Ethylacetate: methanol (9.5:0.5) showed good separation. The crude pigment (2 g) was dissolved in 5 mL of ethyl acetate. With the help of capillary tube, the sample was spotted at the bottom of silica gel coated sheet (6 x 8 cm) and then it was placed in the developing 100 mL beaker containing mobile phase (Ethyl acetate/ Methanol, 9.5:0.5) 5 mL, covered with the watch glass in order to prevent the evaporation of the solvents. The solvent was allowed to run till it reaches about half a centimetre below the top of the plate. After running, the 200 sheets were kept at room temperature for the complete drying of the plate. Spots on TLC were detected under UV light (254 and 365 nm) and by spraying with concentrated H2SO4 followed by heating at 105 °C for 5 min. After drying, the yellow pigment spot was scrapped, mixed with ethyl acetate and filtered using funnel fitted with what man filtered paper and Ethyl acetate was evaporated to dryness under vacuum to afford the pure compound Transitmucin R1 (10 mg), R 2 (10 mg), R3 (5 mg). Rf value of the spot separated on the TLC plate was determined. The solvent system Ethyl acetate: methanol (9.5:0.5) was found to have good separation with single spot when compared to all the solvent systems used for TLC.

**Figure S3.**
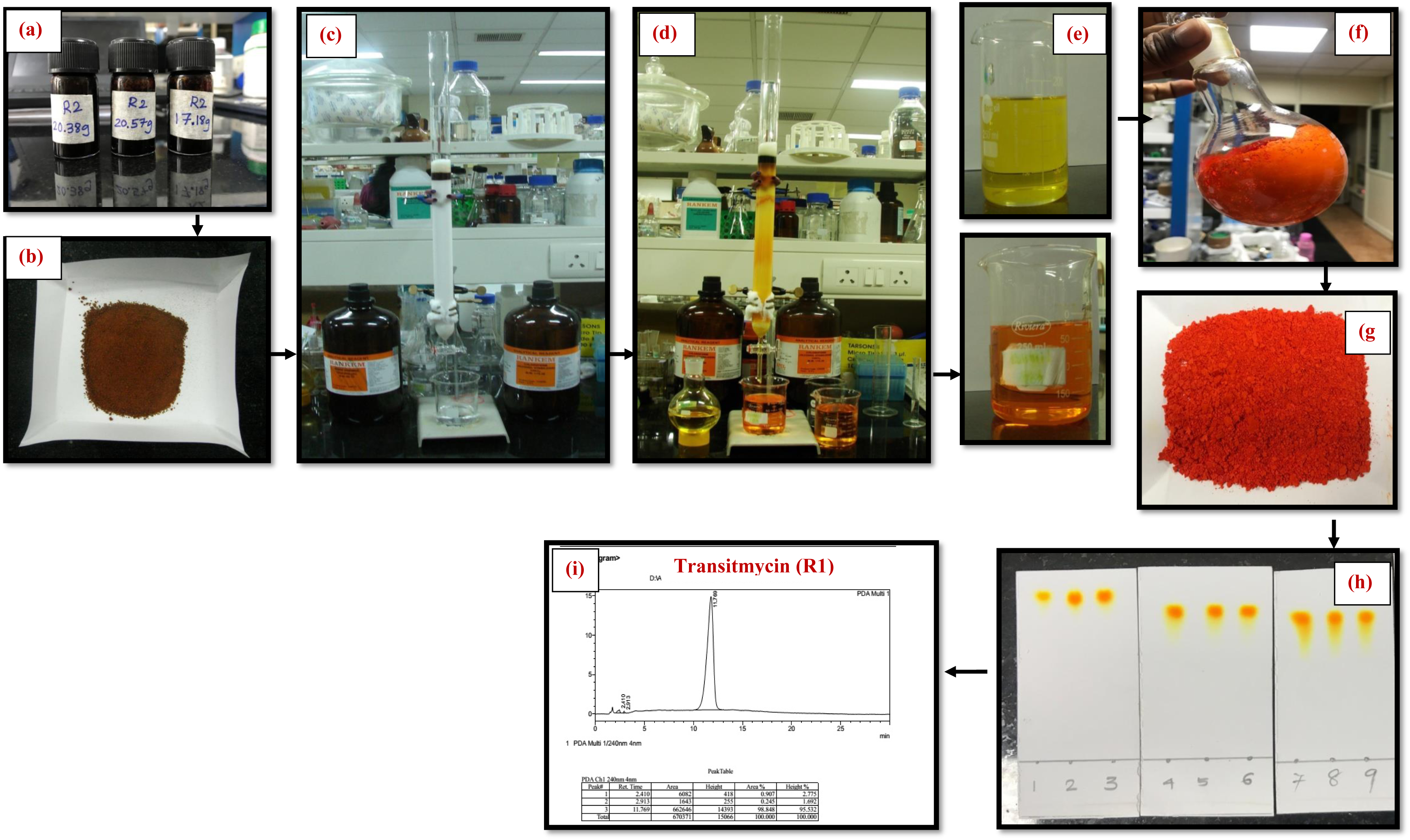
Scheme of purification of EA extract of *Streptomyces* sp. R2 by column chromatography Column chromatography was carried out on Neutral Alumina (230-400 mesh) and Column size: (id 30m × 90 cm).The crude ethyl acetate extract *Streptomyces* sp (R2) was purified using column chromatography packed with neutral alumina using a gradient of 1% **Methanol/Chloroform mixture (CH3OH/ CHCl3) was used as the eluent.** Fractions were collected and concentrated under vacuum to afford pure compounds. **The desired product was monitored in a TLC with pre coated alumina sheet silica.** The isolated compounds were obtained Transitmycin (R1) (200 mg), R2 (100 mg), R3 (50 mg). **(a)** Crude (b) Crude with alumina (c) Before elution (d) After elution (e) solution form (f) solid form (g) After drying (h) TLC (i) R**P-**H**P**LC

**Figure S4.**
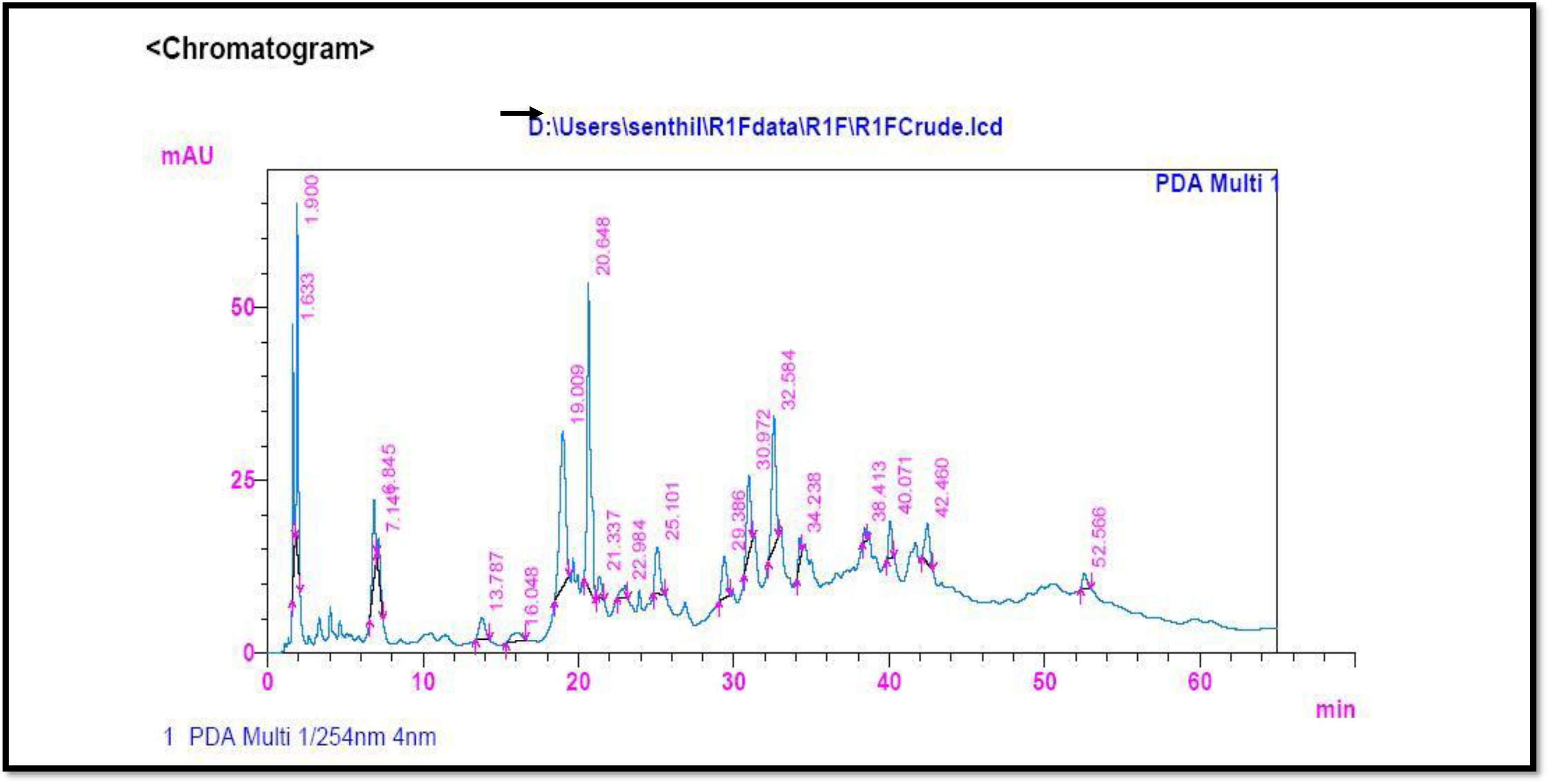
RP HPLC of the crude ethyl acetate extract of *Streptomyces* sp. R2 (Batch II) Analytical HPLC condition: Luna 5u C_18_ (2) 100 (150 X 4.6 mm) Solvent system: A: B (35: 65 v/v); flow rate 2 ml/min, 254 nm Solvent A: Acetonitrile; Solvent B: Water, Detection: PDA, Injection volume: 20µl, Column Temperature: 30 ^0^C

**S-Table 3.** RP-HPLC Retention time of R1, R2, R3.

|  |  |  |
| --- | --- | --- |
| a | Ret. Time of R1 | 5.807 min |
| b | Ret. Time of R1 | 3.062 min |
| c | Ret. Time of R2 | 1.100 min |

**Figure S5.**
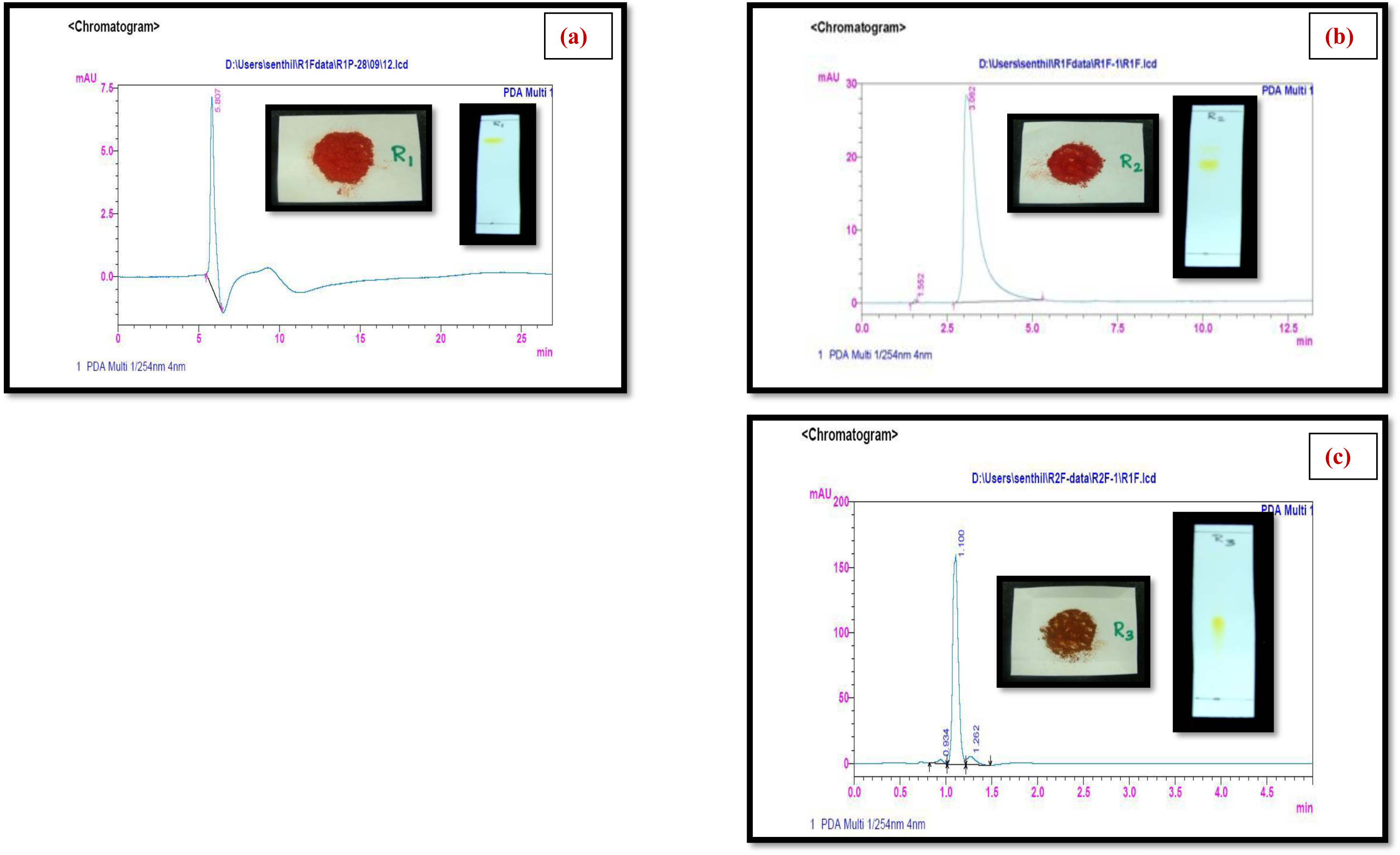
Purity of Transitmycin R1, R2, R3 were checked by RP HPLC method.

**Figure S6:**
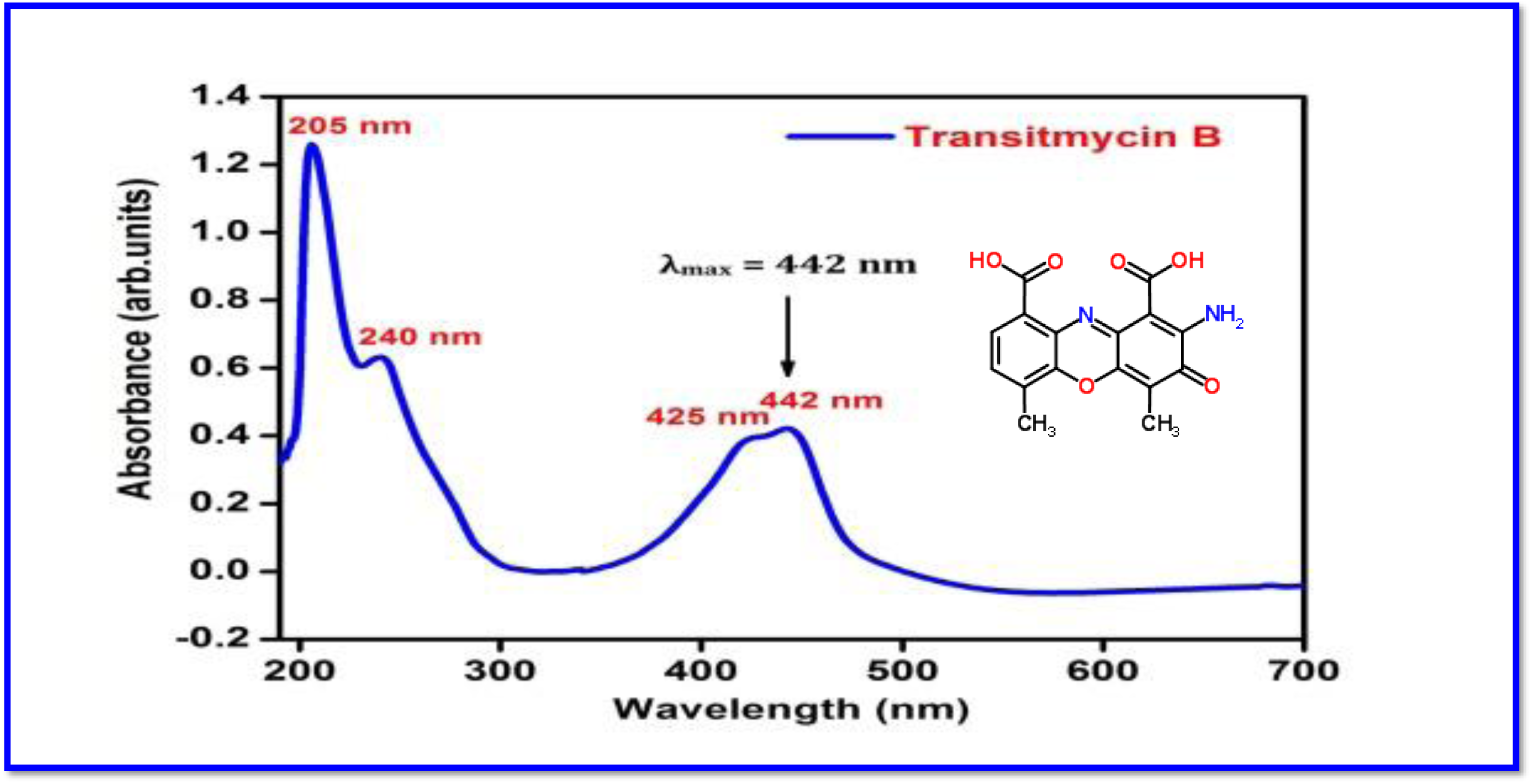
UV/Vis Spectrum of R2 in methanol [UV: (MeOH) λ _max,_ (log ε) 205, (1.25), 240 (0.63), 425 (0.39), 442(0.42) nm].

**Figure S7.**
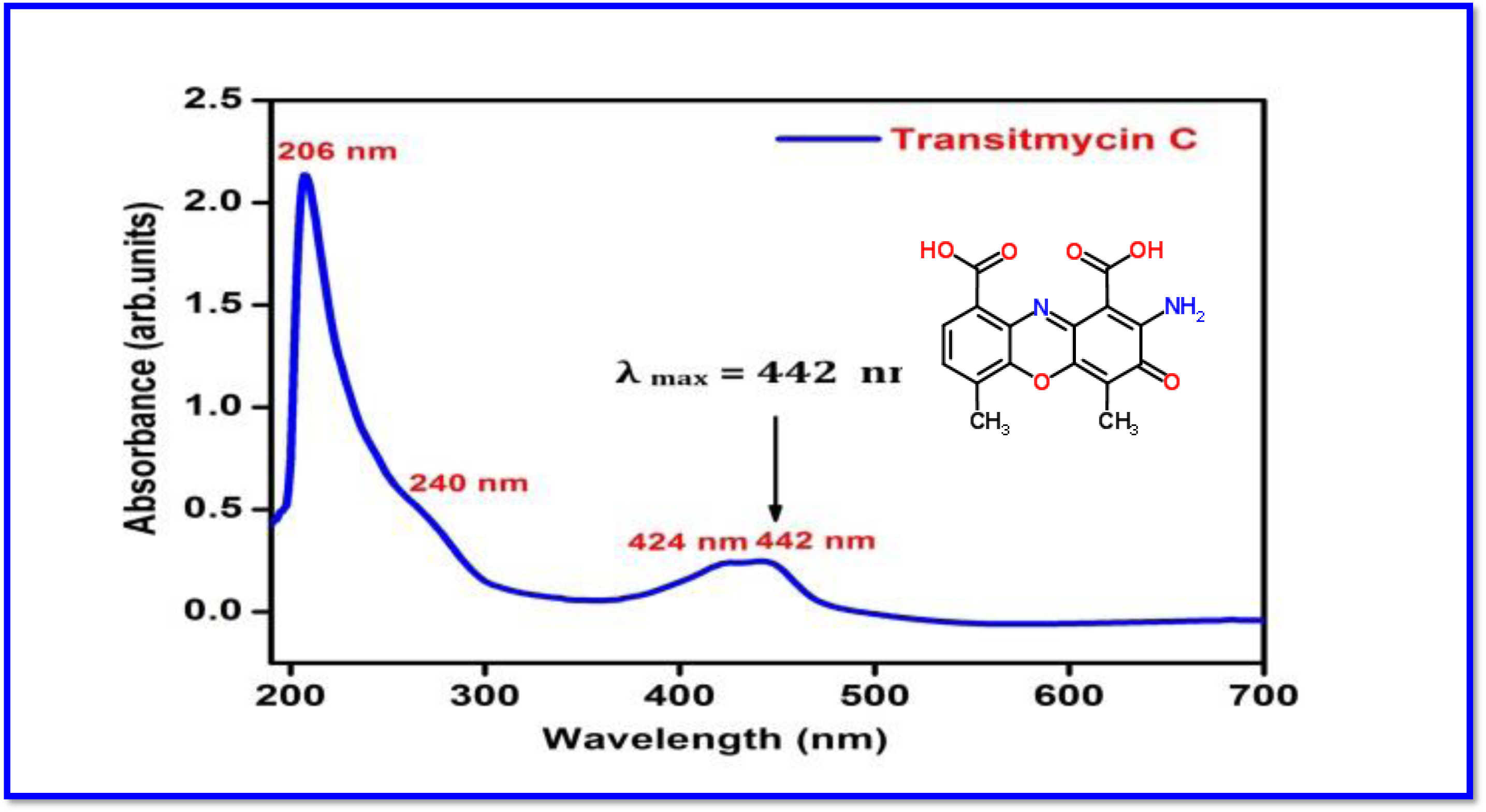
UV/Vis Spectrum of R3 in methanol [UV (MeOH) λ _max_ (log ε) 206 (1.90), 240 (0.69) 424 (0.191), 442.2 (0.19) nm].

**Figure S8.**
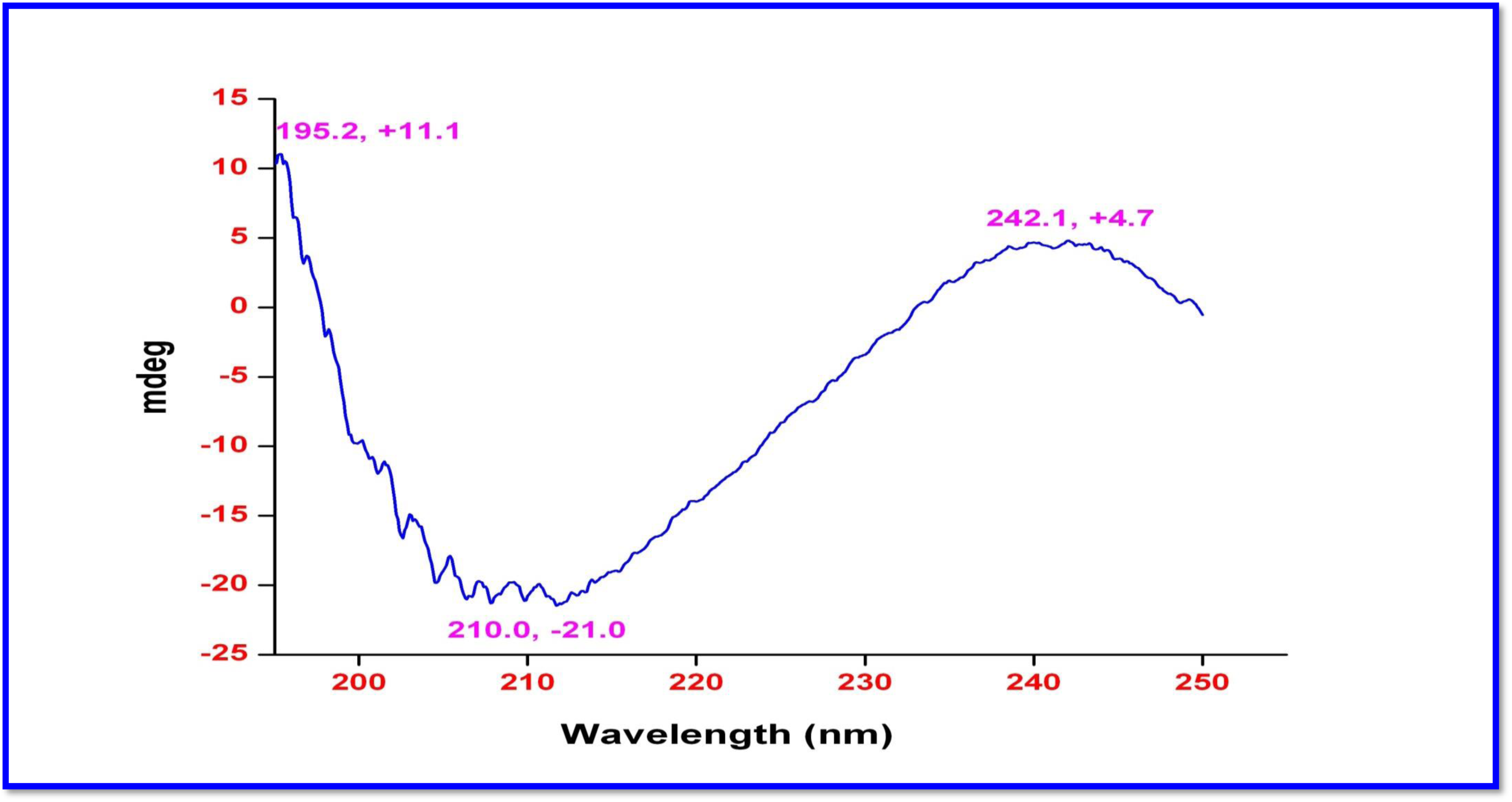
Circular Dichroism Spectrum of Transitmycin (R1) in methanol CD: [MeOH, [nm], (mdeg)]: λ_max_(Δε) 195 (+11.1), 210 (- 21.0), 242 (+4.7)

**Figure S9.**
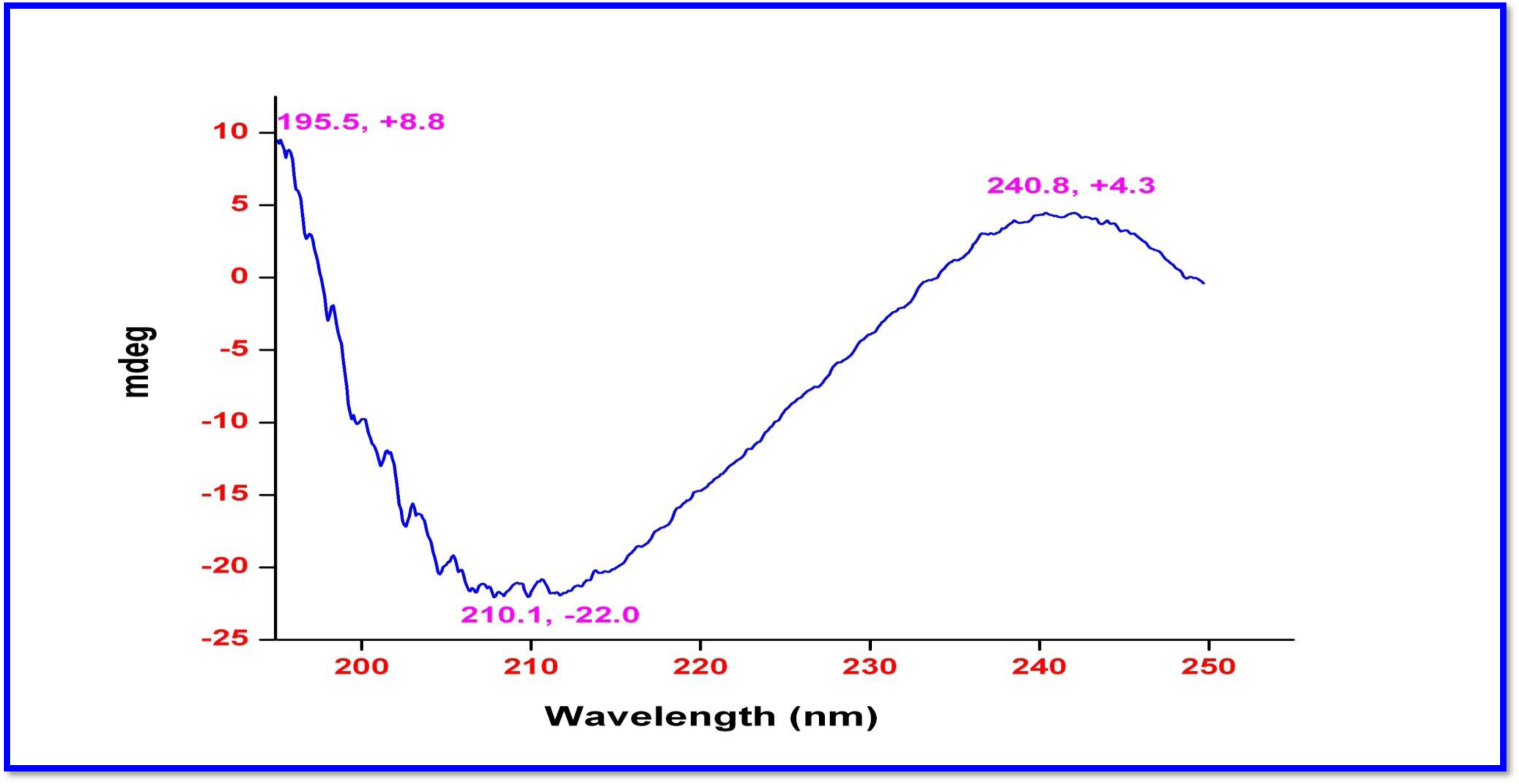
Circular Dichroism Spectrum of R2 in methanol CD: [MeOH, [nm], (mdeg)]: λ_max,_ (Δε) 195 (+8.8), 210 (-22.0), 240 (+4.3) nm.

**Figure S10.**
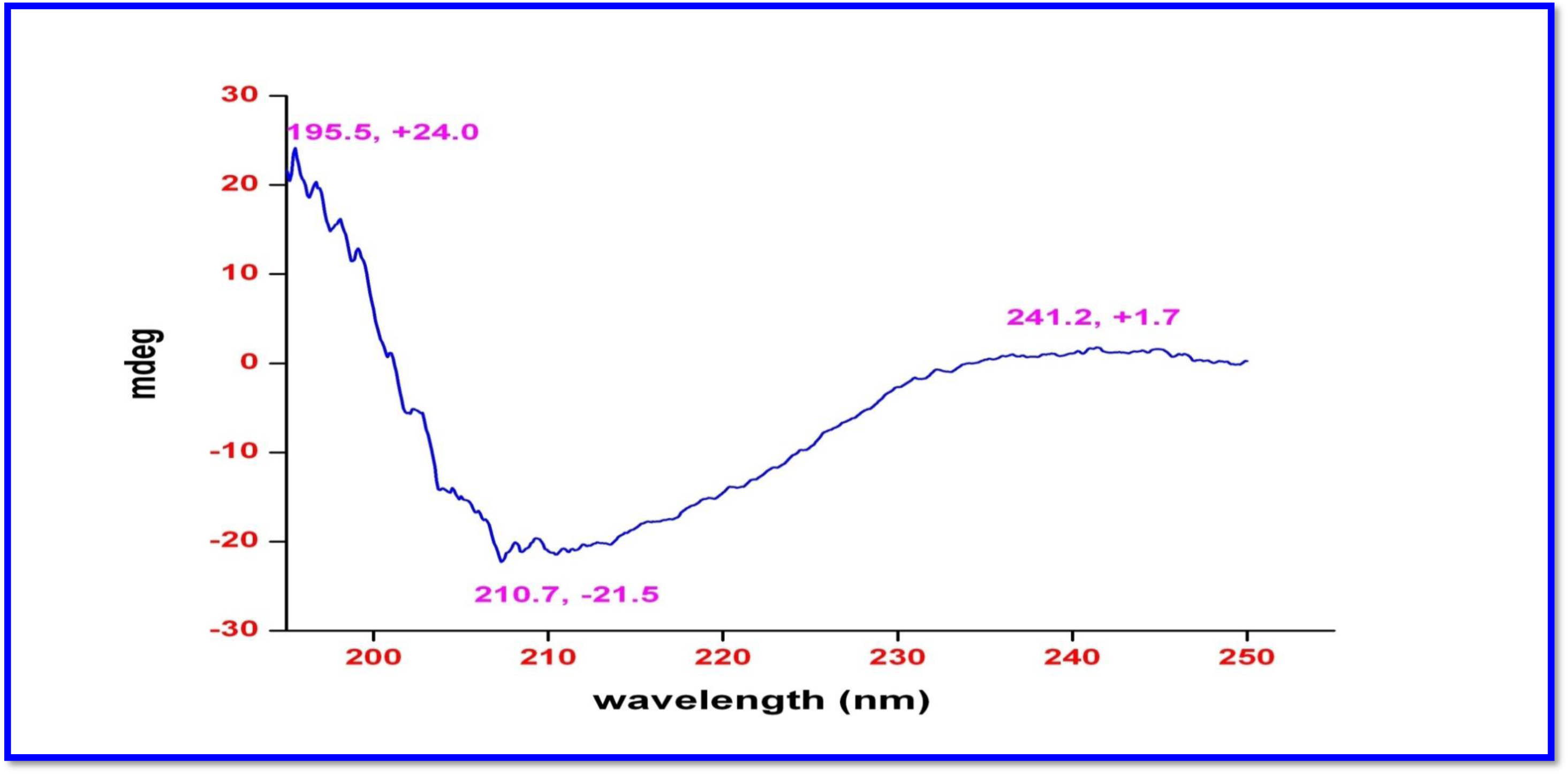
Circular Dichroism Spectrum of R3 in methanol CD: [MeOH, [nm], (mdeg)] λ_max_(Δε) 195 (+24.0), 210 (-21.5), 241 (+1.7)

**Figure S11.**
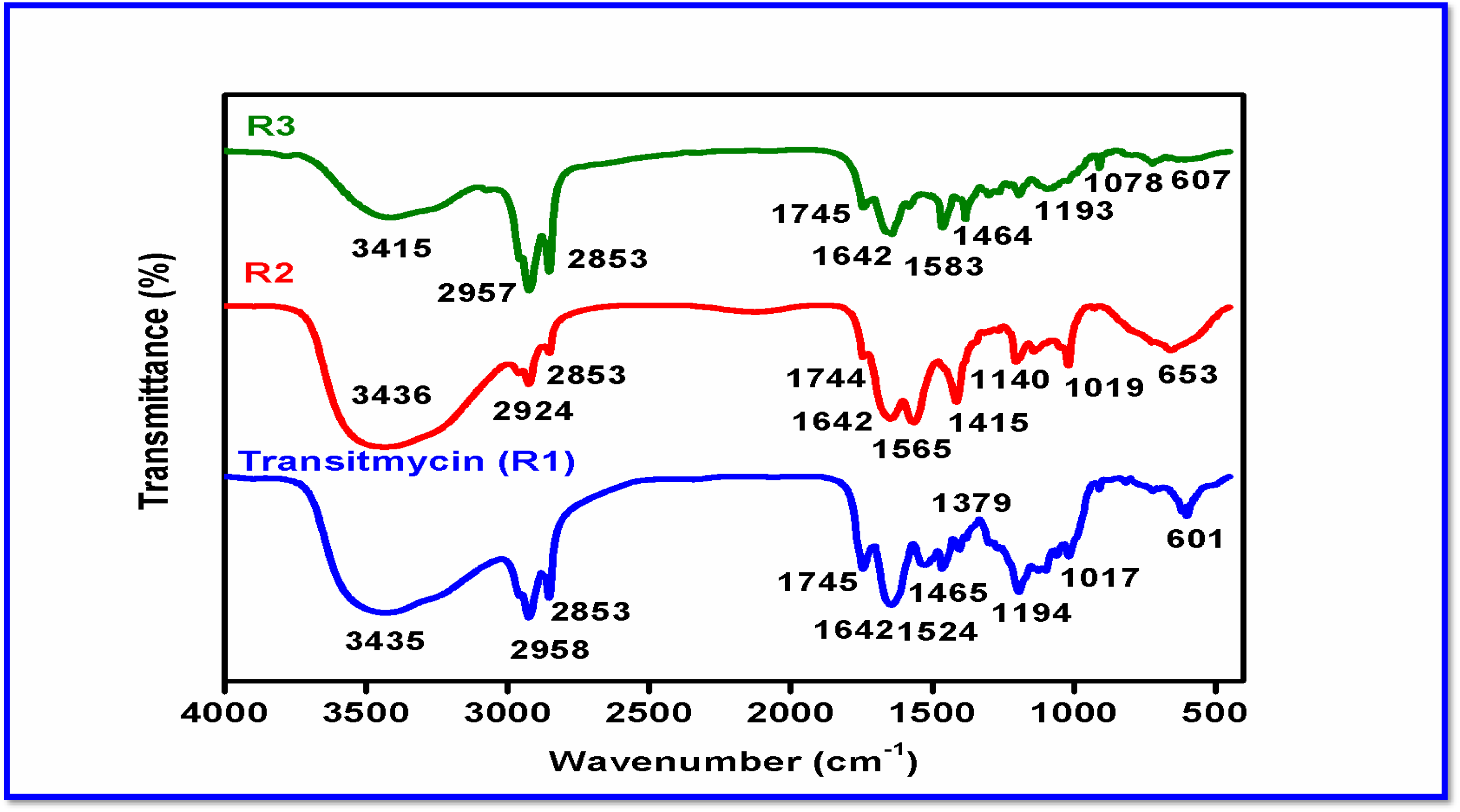
FT-IR Spectrum of Transitmycin (R1), R2, R3.

**Figure S12.**
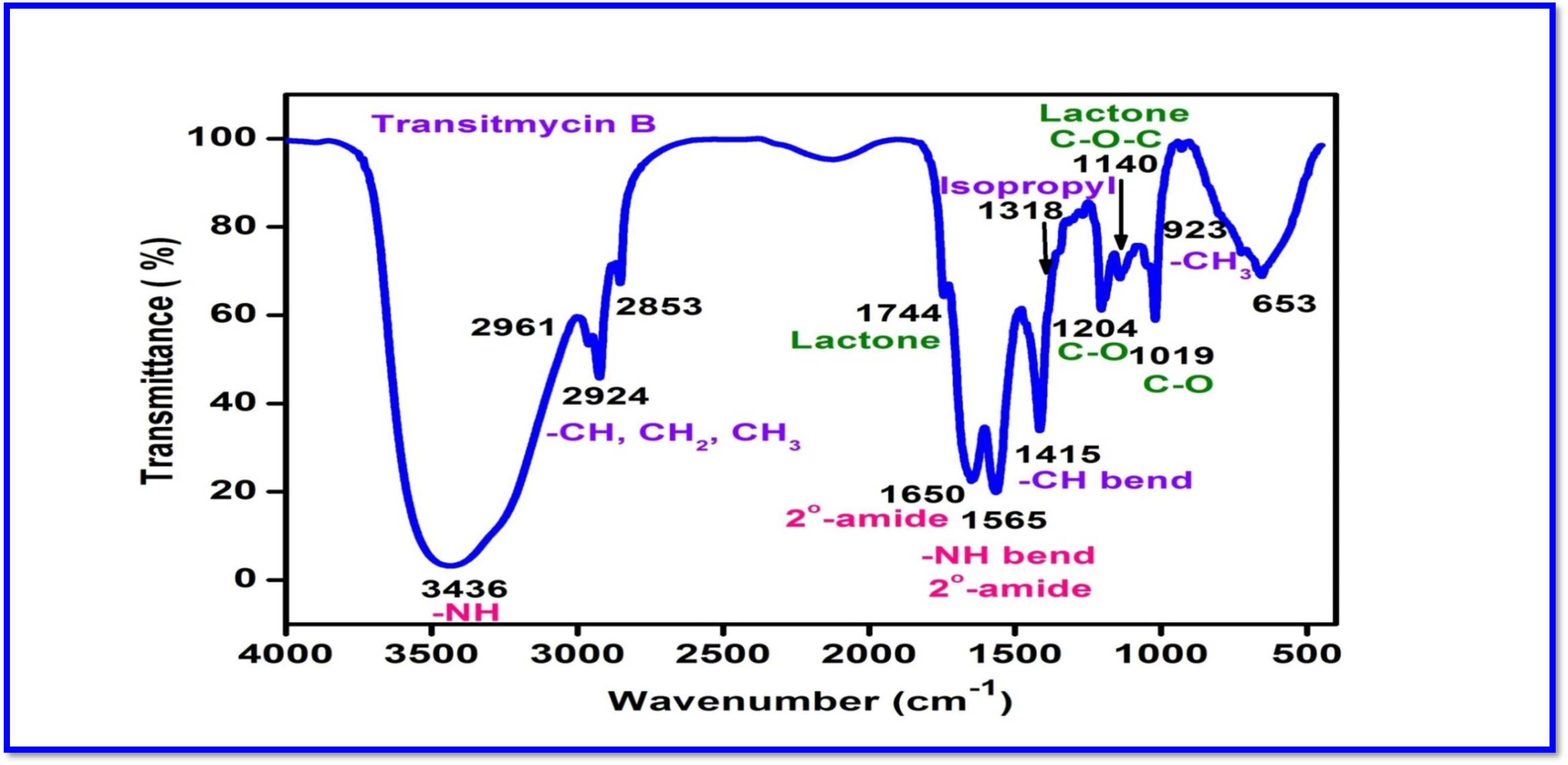
FT-IR Spectrum of R2.

**Figure S13.**
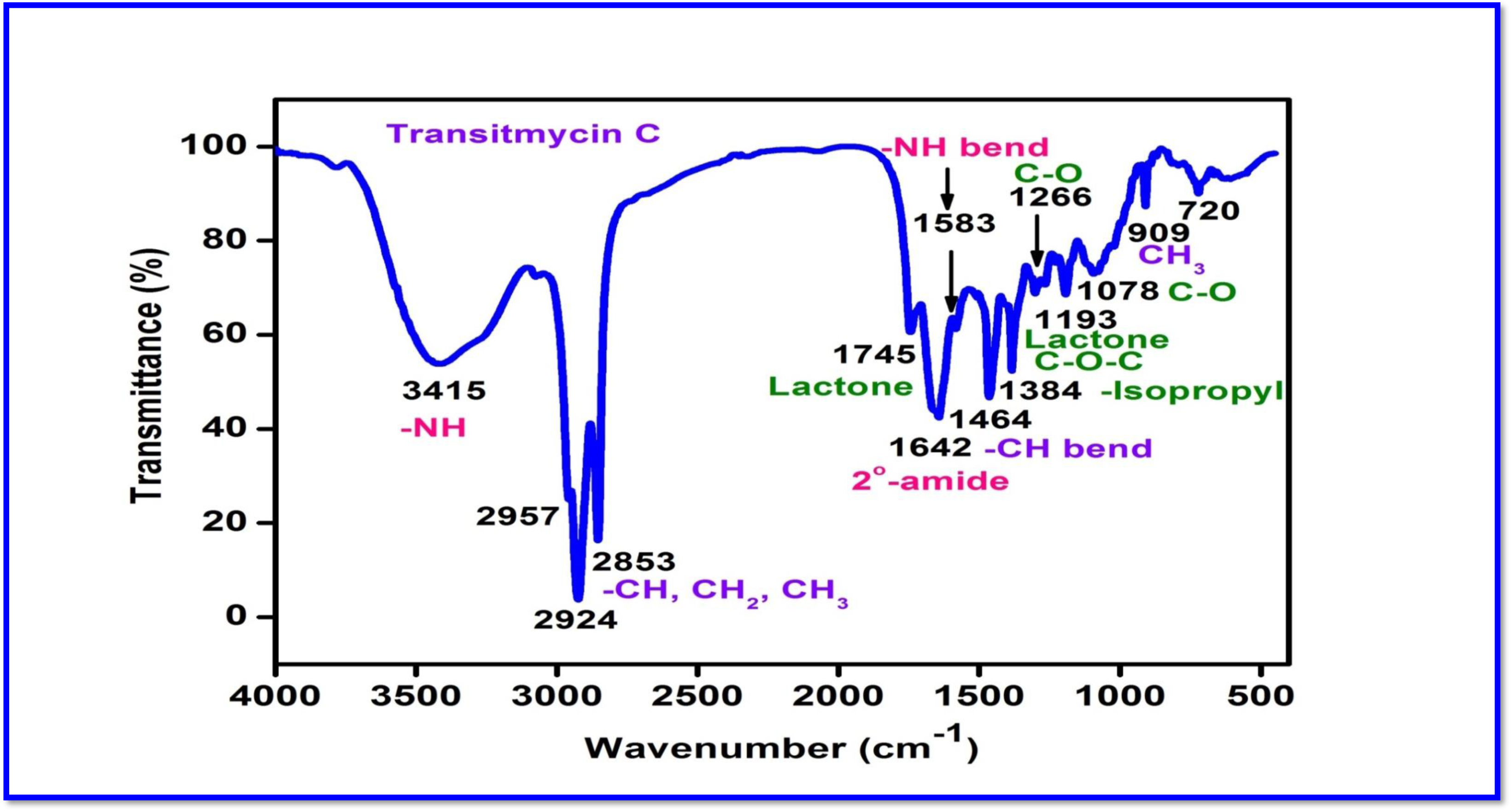
FT-IR Spectrum of R3.

**Table S4.** IR Absorption Frequencies of Functional Groups Present in Transitmycin (R1), R2, R3.

|  |  |
| --- | --- |
| **R1** | IR (KBr $\text{cm}^{-1}$), 3435 $\text{cm}^{-1}$ for NH, 2958, 2924 $\text{cm}^{-1}$ (m, -CH str, asym, $\text{CH}_3$ and $\text{CH}_2$), 2872 $\text{cm}^{-1}$, 2853 $\text{cm}^{-1}$ (m, -CH str, sym, $\text{CH}_3$ and $\text{CH}_2$), 1746 $\text{cm}^{-1}$ (s, C=O str, lactone ring), 1642 $\text{cm}^{-1}$ (s, -C=O str, $2^0$ amide), 1524, 1503 (m, -NH bend, $2^0$ amide), 1466 (m, CH bend (scissoring), $\text{CH}_2$), 1379 $\text{cm}^{-1}$ (s, -CH bend, isopropyl group), 1268 (s, C-O str, ester), 1194 (C-O-C of lactone), 1099, 1059, 1017 (s, C-O or C-N), 720, 712, 694, 689 (s, -CH bend, oop, aromatic ring), 909 (w, $\text{CH}_3$ rocking). |
| **R2** | IR (KBr $\text{cm}^{-1}$), 3436 $\text{cm}^{-1}$ for NH, 2961, 2924 $\text{cm}^{-1}$ (m, -CH str, asym, $\text{CH}_3$ and $\text{CH}_2$), 2872 $\text{cm}^{-1}$, 2853 $\text{cm}^{-1}$ (m, -CH str, sym, $\text{CH}_3$ and $\text{CH}_2$), 1744 $\text{cm}^{-1}$ (s, C=O str, lactone ring), 1650 $\text{cm}^{-1}$ (s, -C=O str, $2^0$ amide), 1565, 1503 (m, -NH bend, $2^0$ amide), 1415 (m, CH bend (scissoring), $\text{CH}_2$), 1318 $\text{cm}^{-1}$ (s, -CH bend, isopropyl group), 1268, 1204 (s, C-O str, ester), 1140 (C-O-C of lactone), 1045, 1019 (s, C-O or C-N), 720, 712, 653 (s, -CH bend, oop, aromatic ring), 929 (w, $\text{CH}_3$ rocking). |
| **R3** | IR (KBr $\text{cm}^{-1}$), 3415 $\text{cm}^{-1}$ for NH, 2957, 2924 $\text{cm}^{-1}$ (m, -CH str, asym, $\text{CH}_3$ and $\text{CH}_2$), 2872 $\text{cm}^{-1}$, 2853 $\text{cm}^{-1}$ (m, -CH str, sym, $\text{CH}_3$ and $\text{CH}_2$), 1745 $\text{cm}^{-1}$ (s, C=O str, lactone ring), 1642 $\text{cm}^{-1}$ (s, -C=O str, $2^0$ amide), 1583, 1507 (m, -NH bend, $2^0$ amide), 1464 (m, CH bend (scissoring), $\text{CH}_2$), 1384, 1301 $\text{cm}^{-1}$ (s, -CH bend, isopropyl group), 1266, 1228 (s, C-O str, ester), 1193 (C-O-C of lactone), 1093, 1078, 1017 (s, C-O or C-N), 909 (w, $\text{CH}_3$ rocking), 720, 712, 694, 689 (s, -CH bend, oop, aromatic ring). |

**Figure S14.**
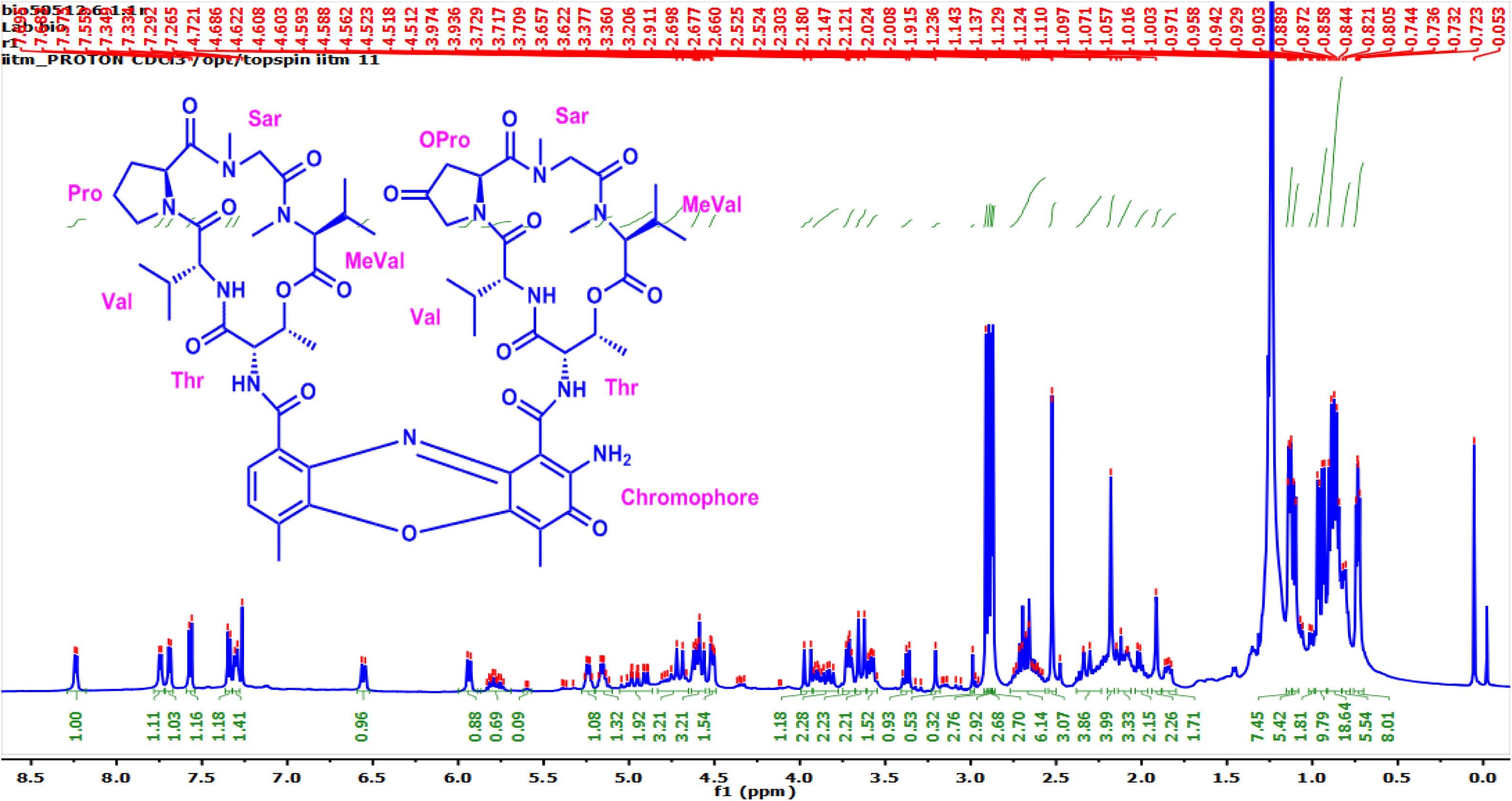
^1^H-NMR (500 MHz, CDCl_3_) Spectrum of Transitmycin (R1)

**Figure S15.**
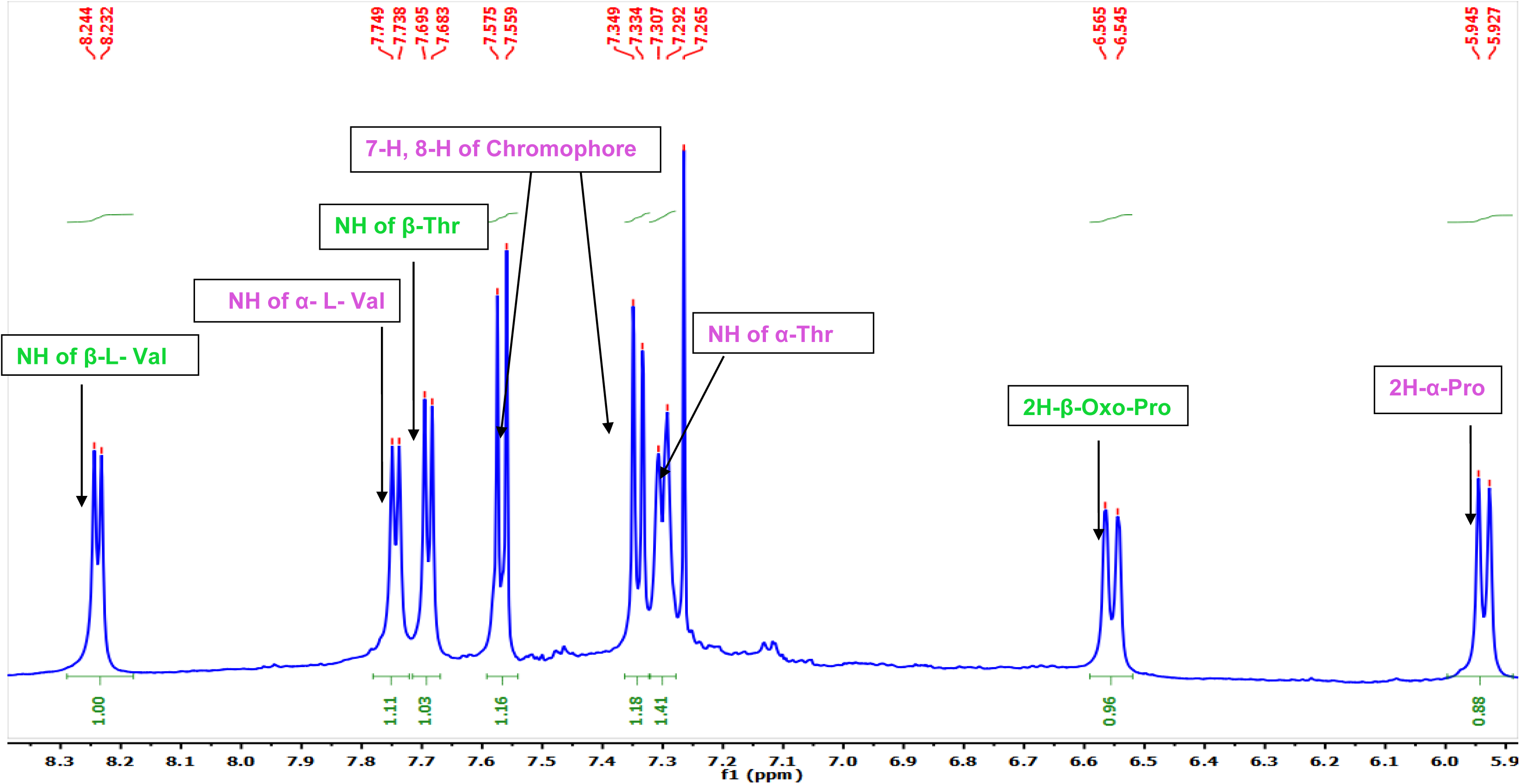
Expansion of ^1^H-NMR (500 MHz, CDCl_3_) Spectrum of Transitmycin (R1)

**Figure S16.**
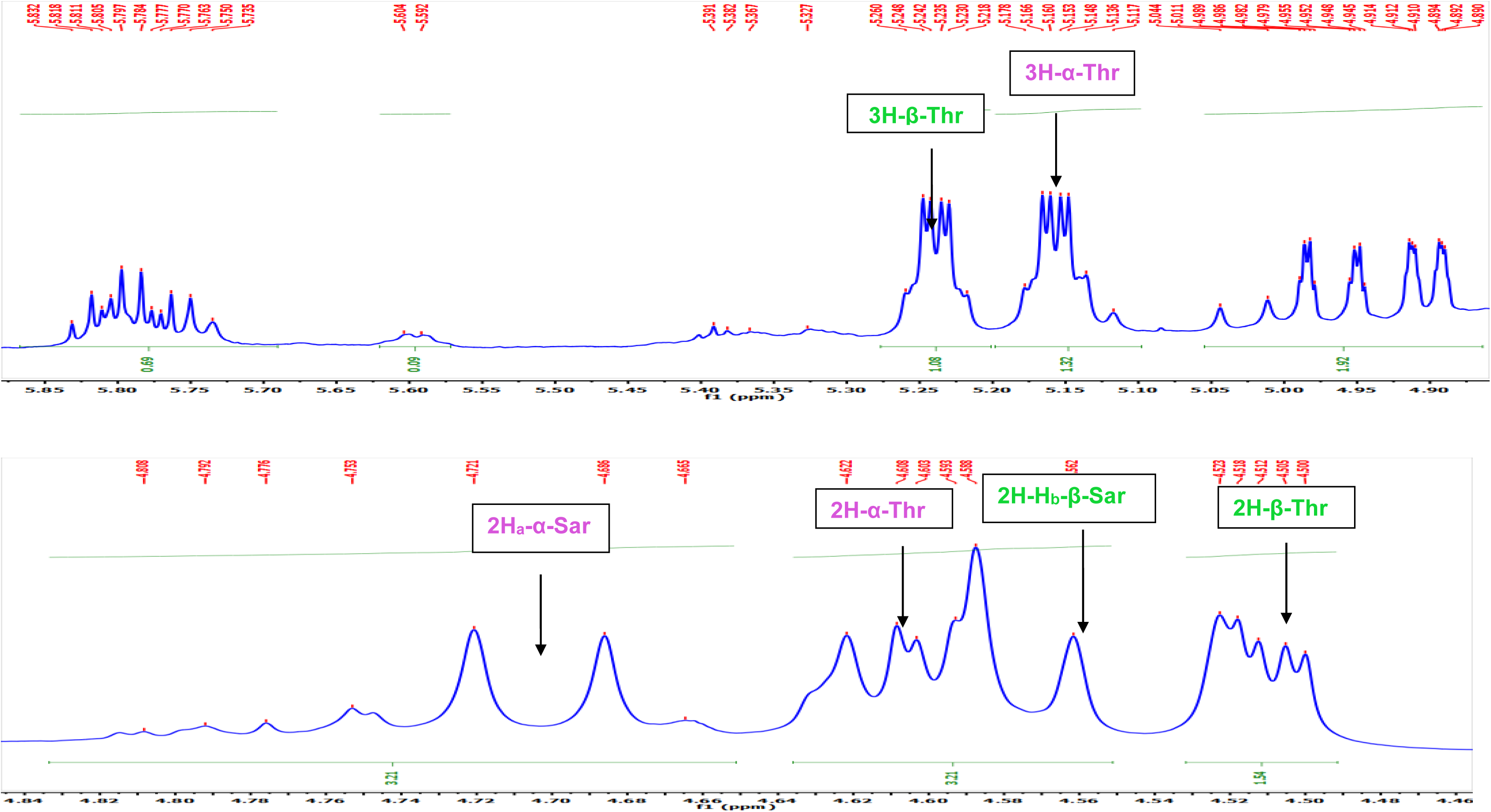
Expansion of ^1^H-NMR (500 MHz, CDCl_3_) Spectrum of Transitmycin (R1)

**Figure S17.**
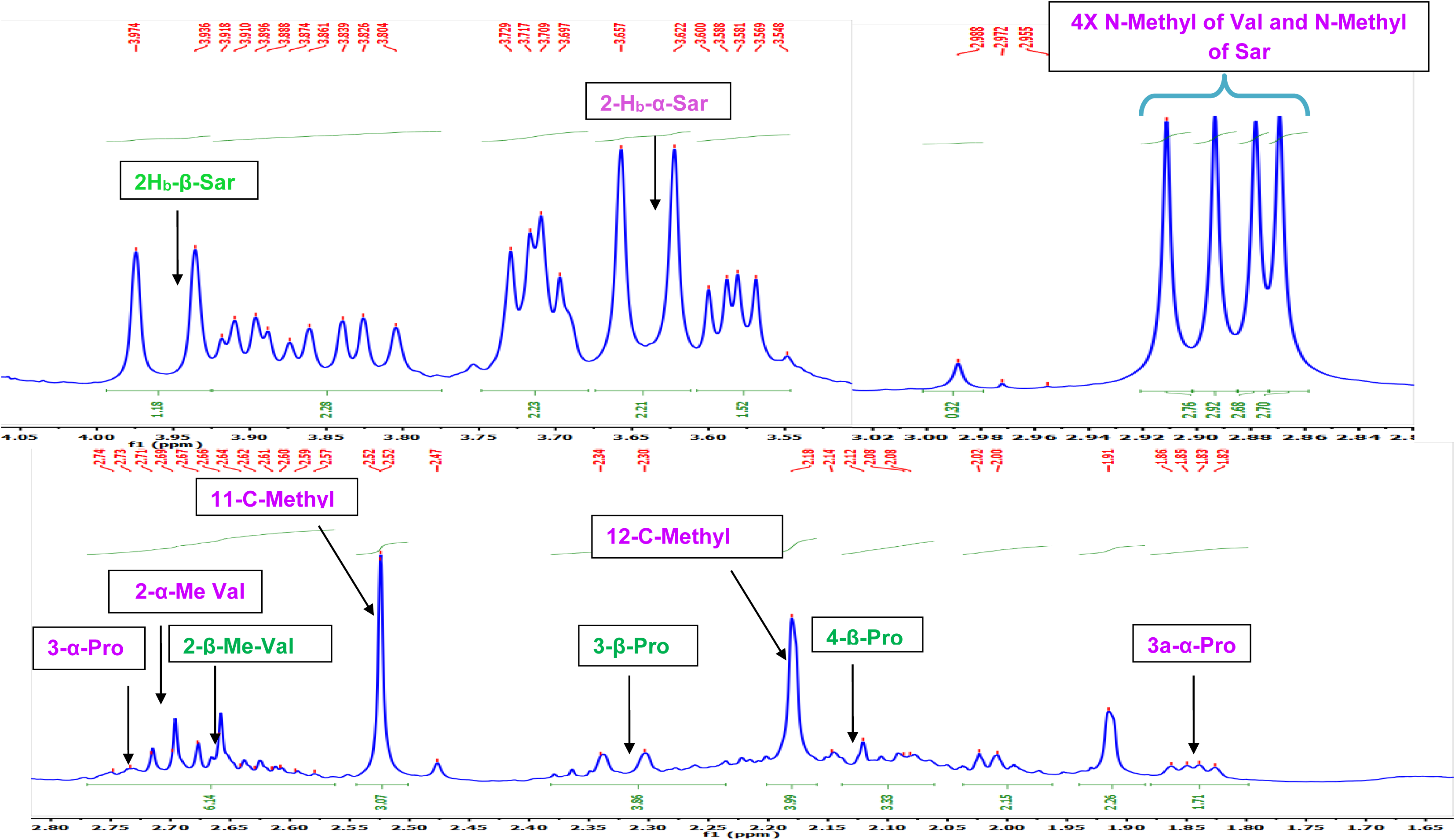
Expansion of ^1^H-NMR (500 MHz, CDCl_3_) Spectrum of Transitmycin (R1)

**Figure S18.**
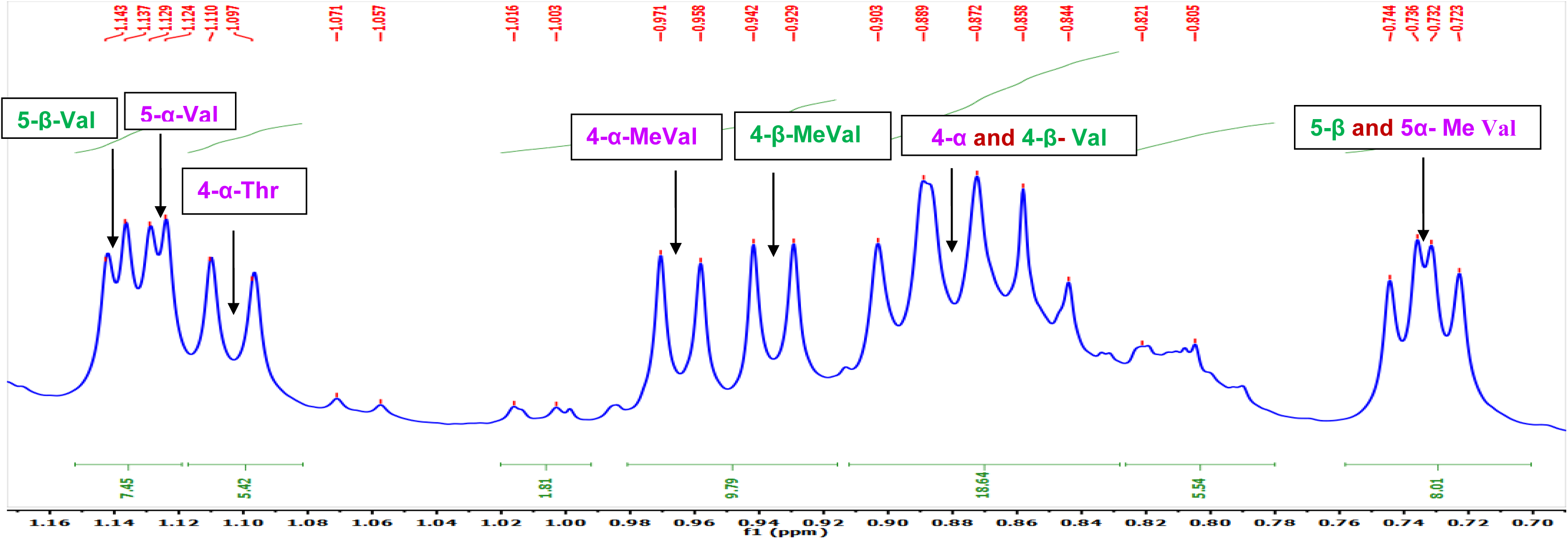
Expansion of ^1^H-NMR (500 MHz, CDCl_3_) Spectrum of Transitmycin (R1)

**Figure S19.**
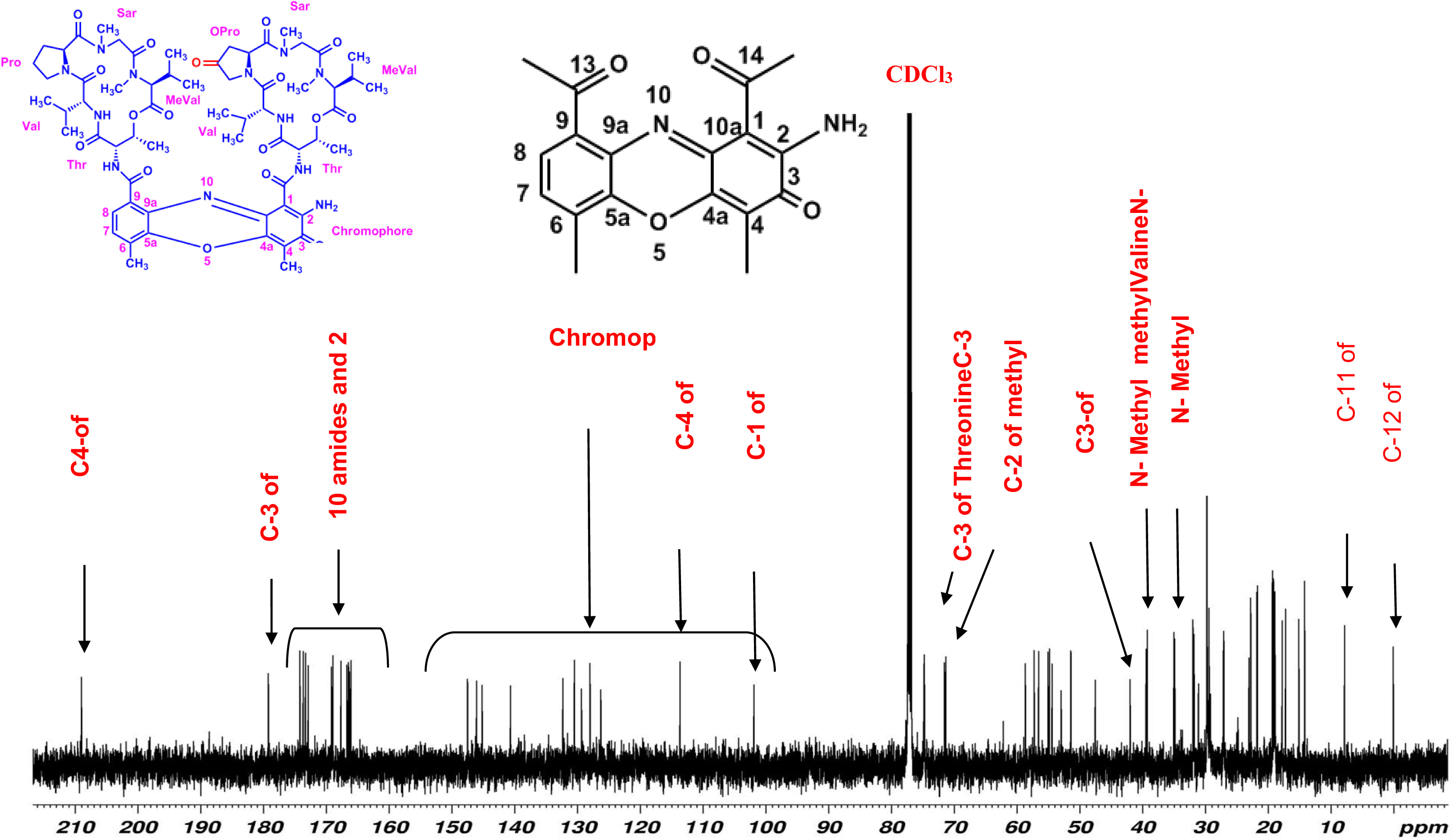
^13^C-NMR (125 MHz, CDCl3) Spectrum of Transitmycin (R1)

**Figure S20.**
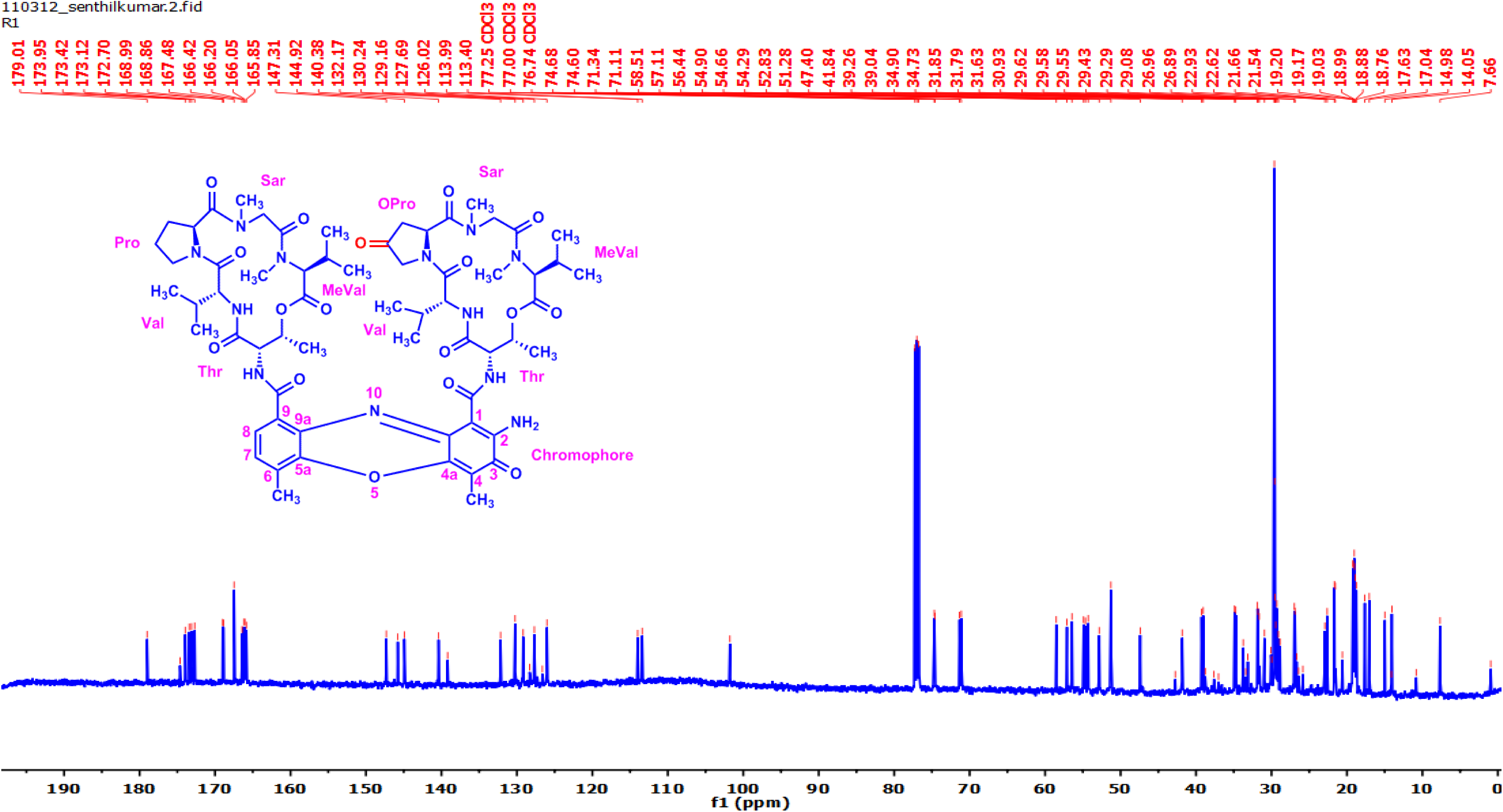
^13^C-NMR (125 MHz, CDCl_3_) Spectrum of Transitmycin (R1)

**Figure S21.**
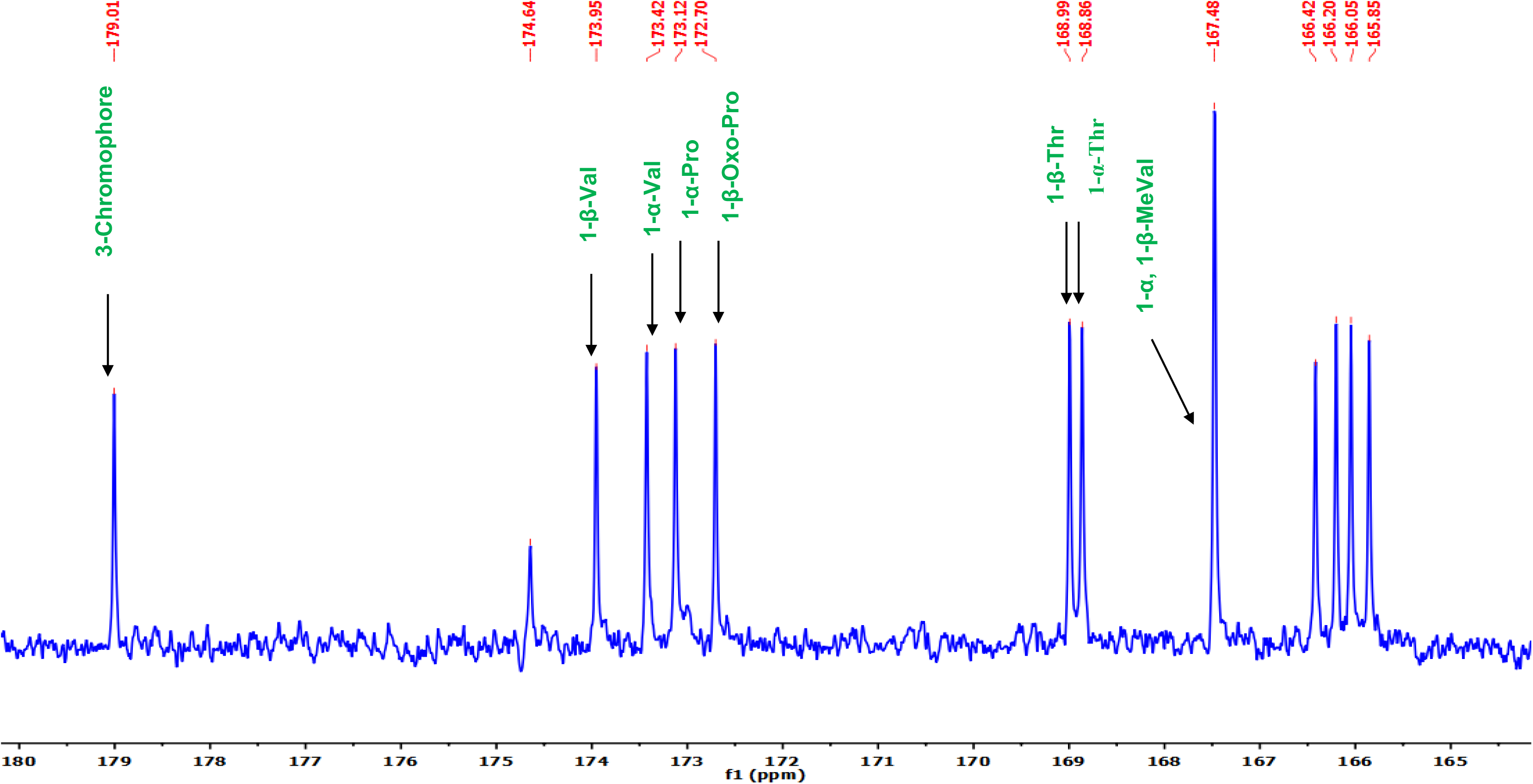
^13^C-NMR (125 MHz, CDCl3) Spectrum of Transitmycin (R1)

**Figure S22.**
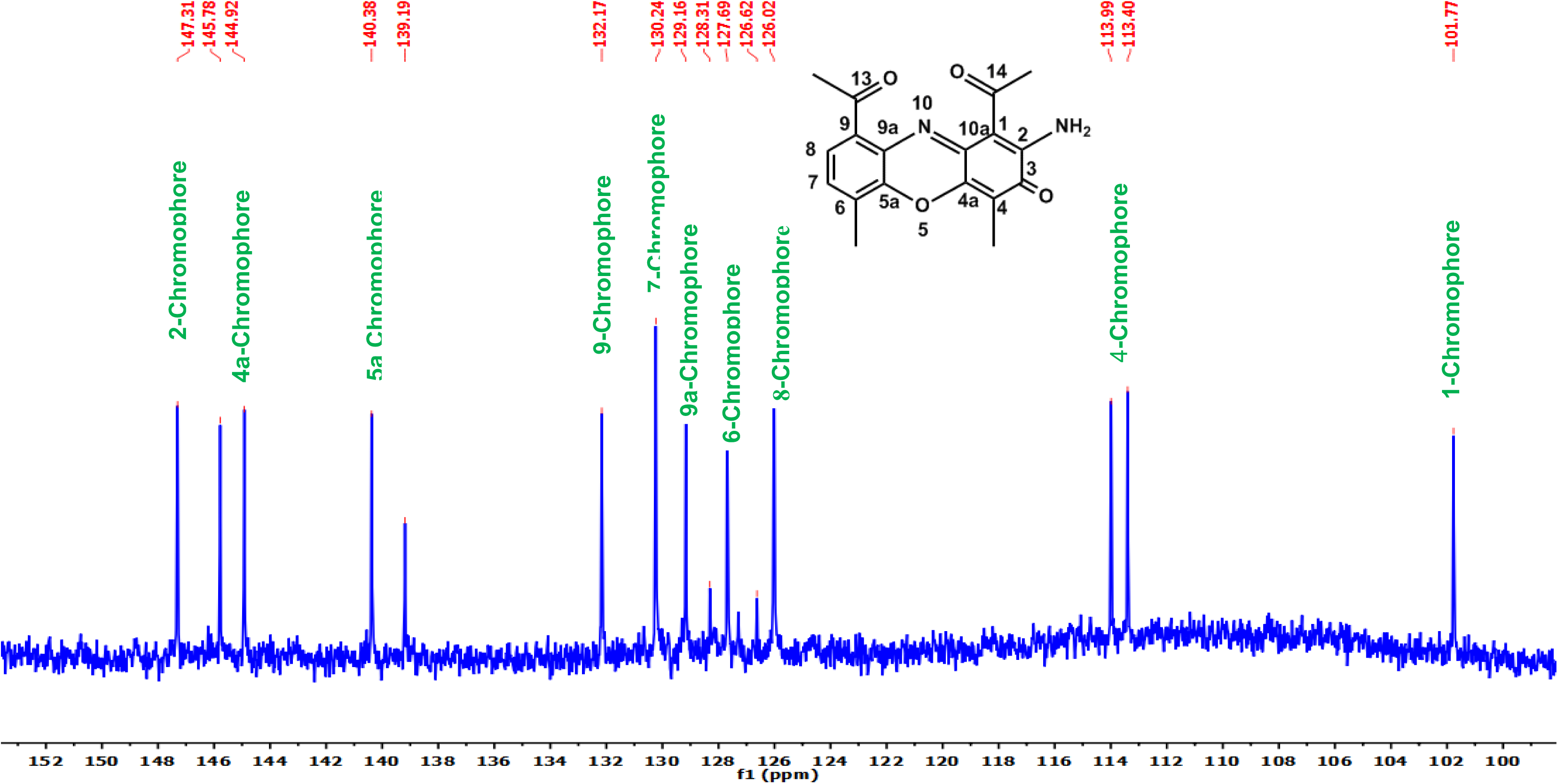
^13^C-NMR (125 MHz, CDCl3) Spectrum of Transitmycin (R1)

**Figure S23.**
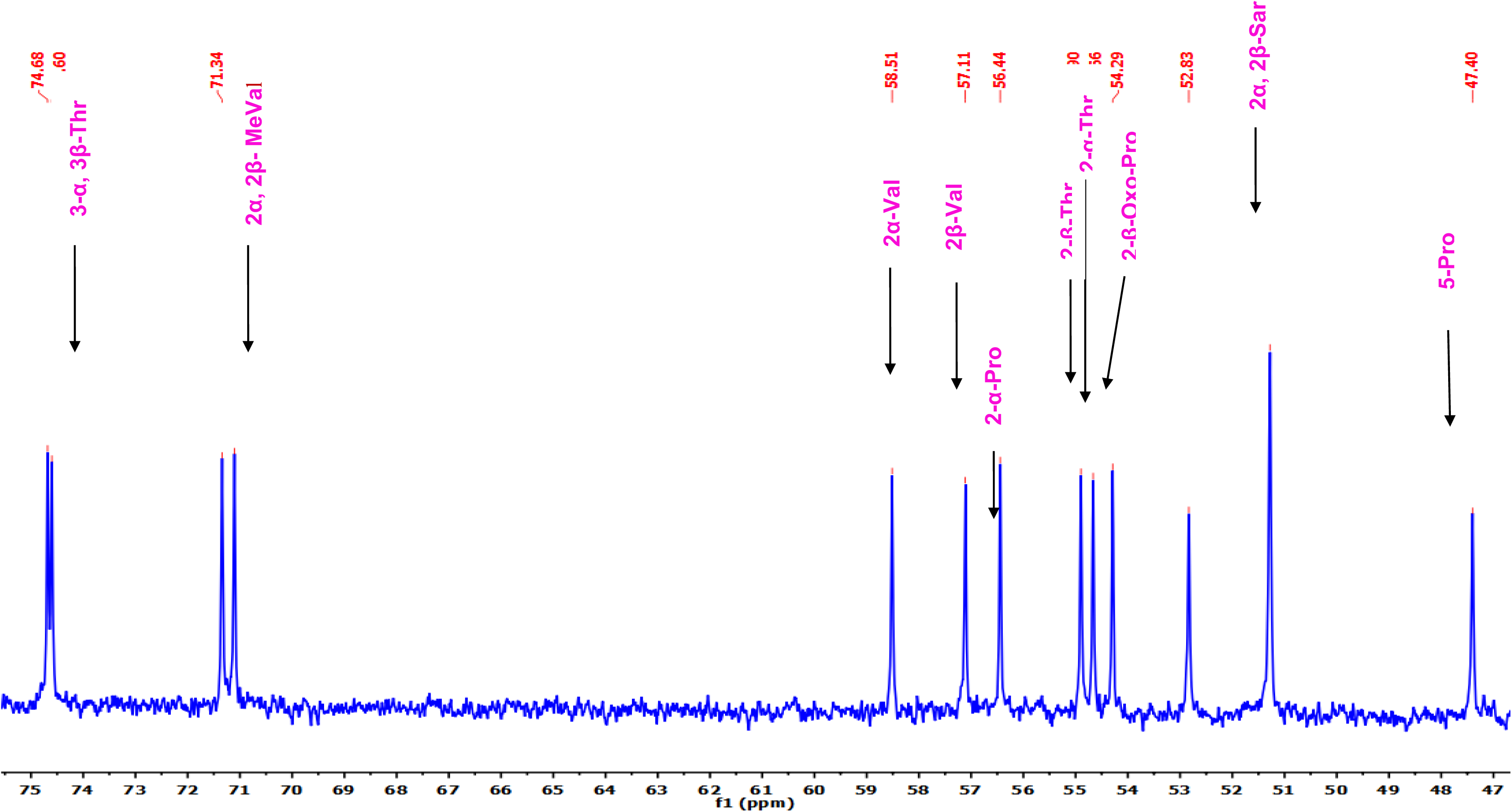
^13^C-NMR (125 MHz, CDCl3) Spectrum of Transitmycin (R1)

**Figure S24.**
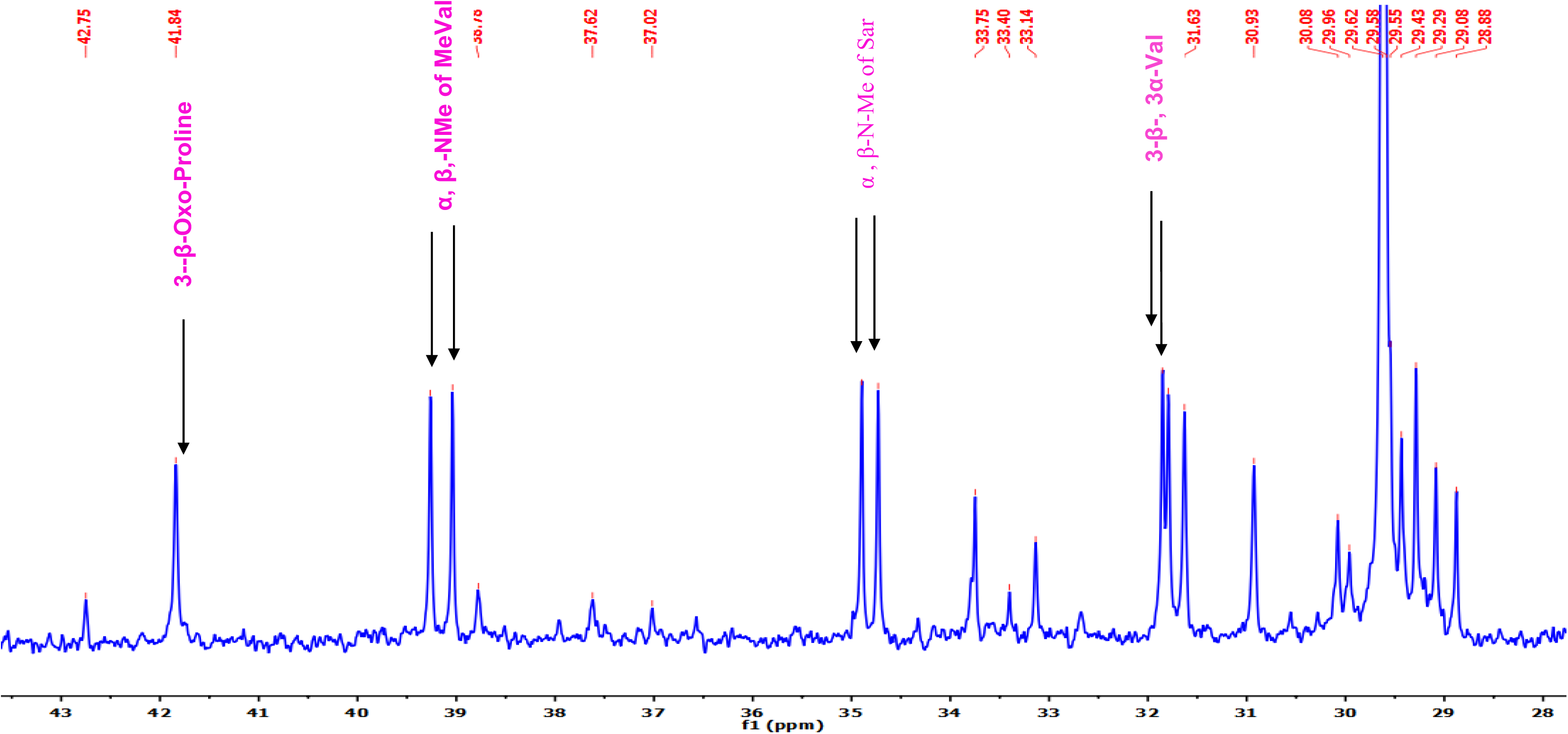
^13^C-NMR (125 MHz, CDCl3) Spectrum of Transitmycin (R1)

**Figure S25.**
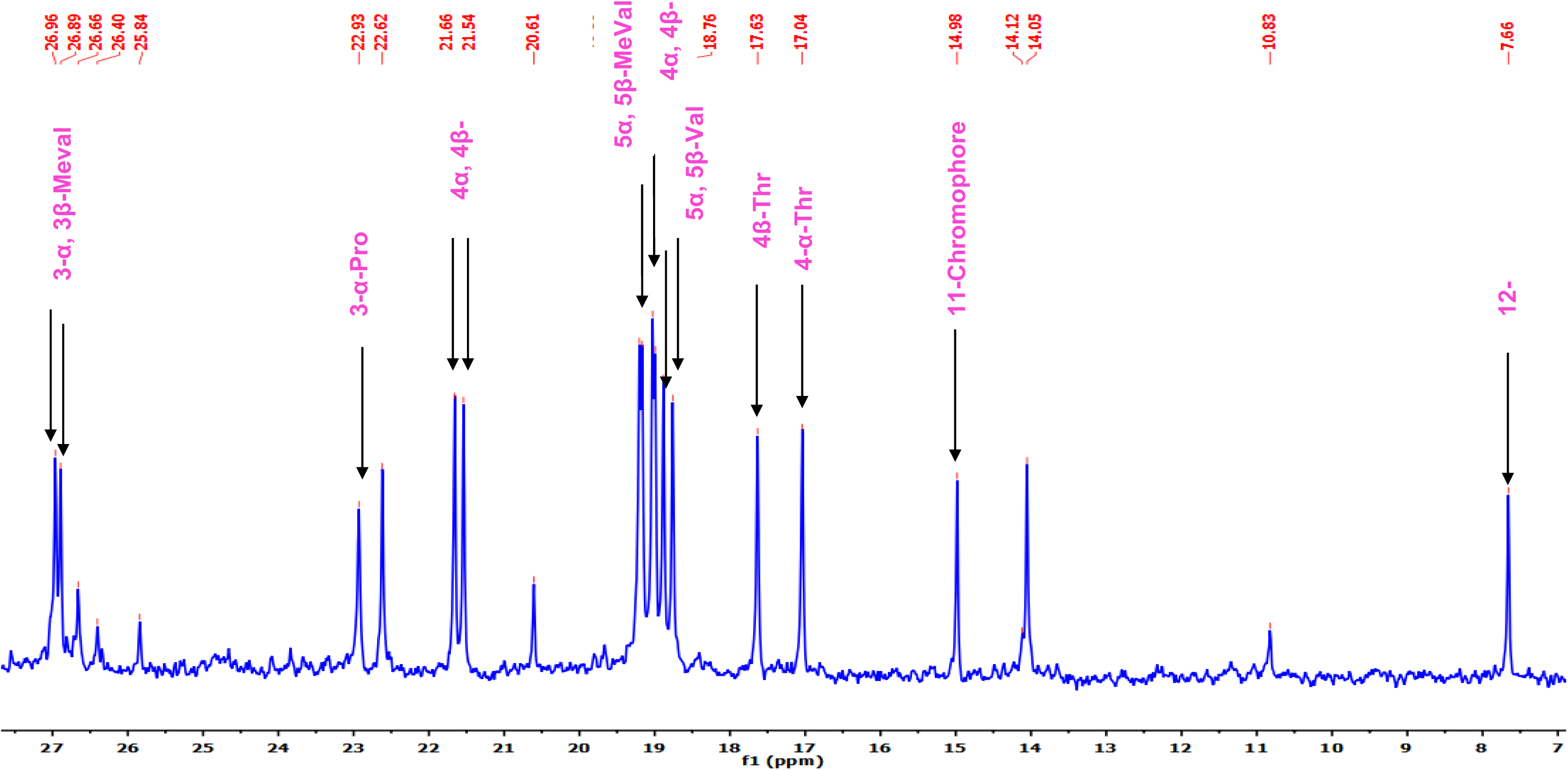
^13^C-NMR (125 MHz, CDCl3) Spectrum of Transitmycin (R1)

**Figure S26.**
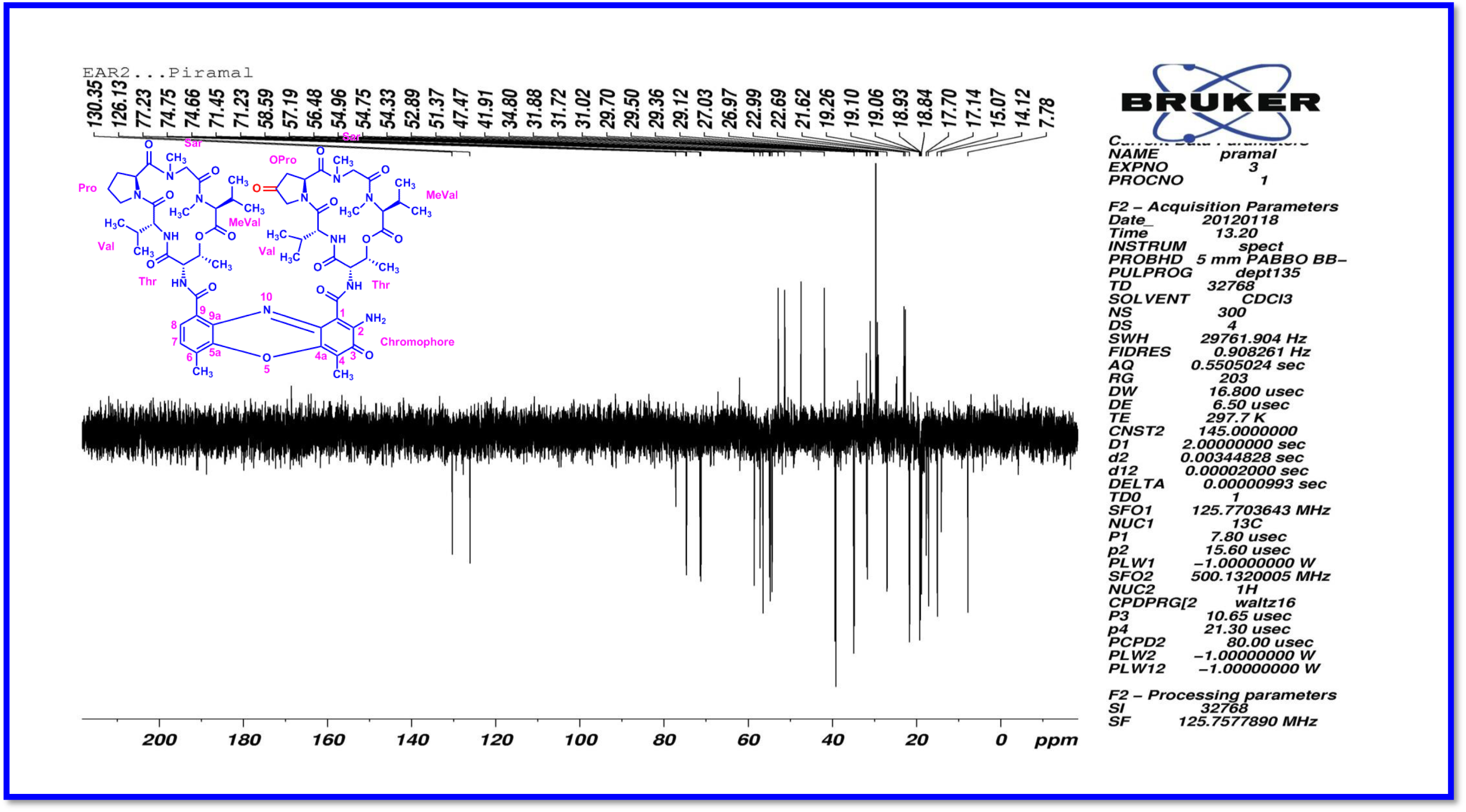
DEPT135 (125 MHz, CDCl_3_) Spectrum of Transitmycin (R1)

**Figure S27.**
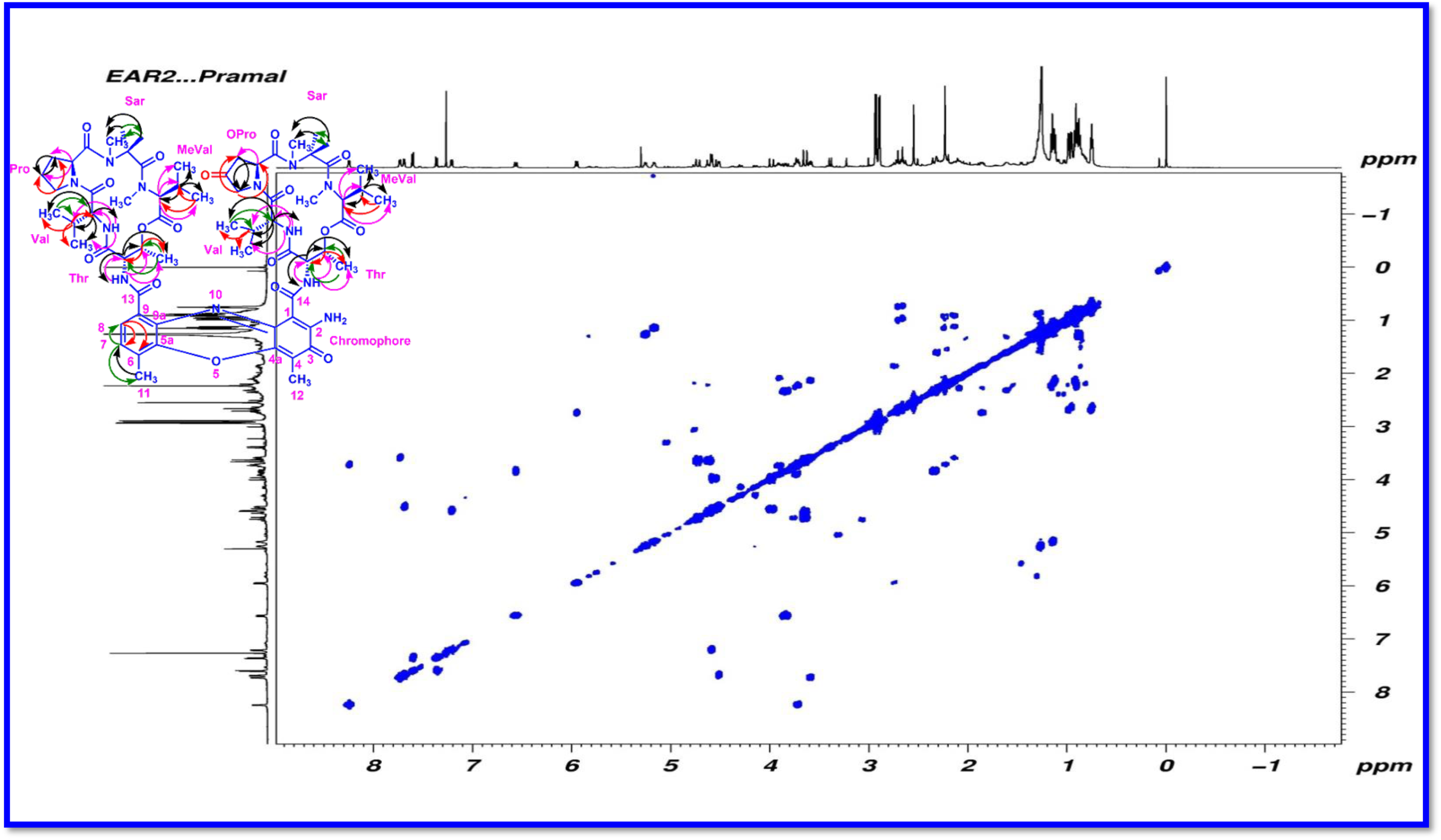
COSY (500 MHz, CDCl3) Spectrum of Transitmycin (R1)

**Figure S28.**
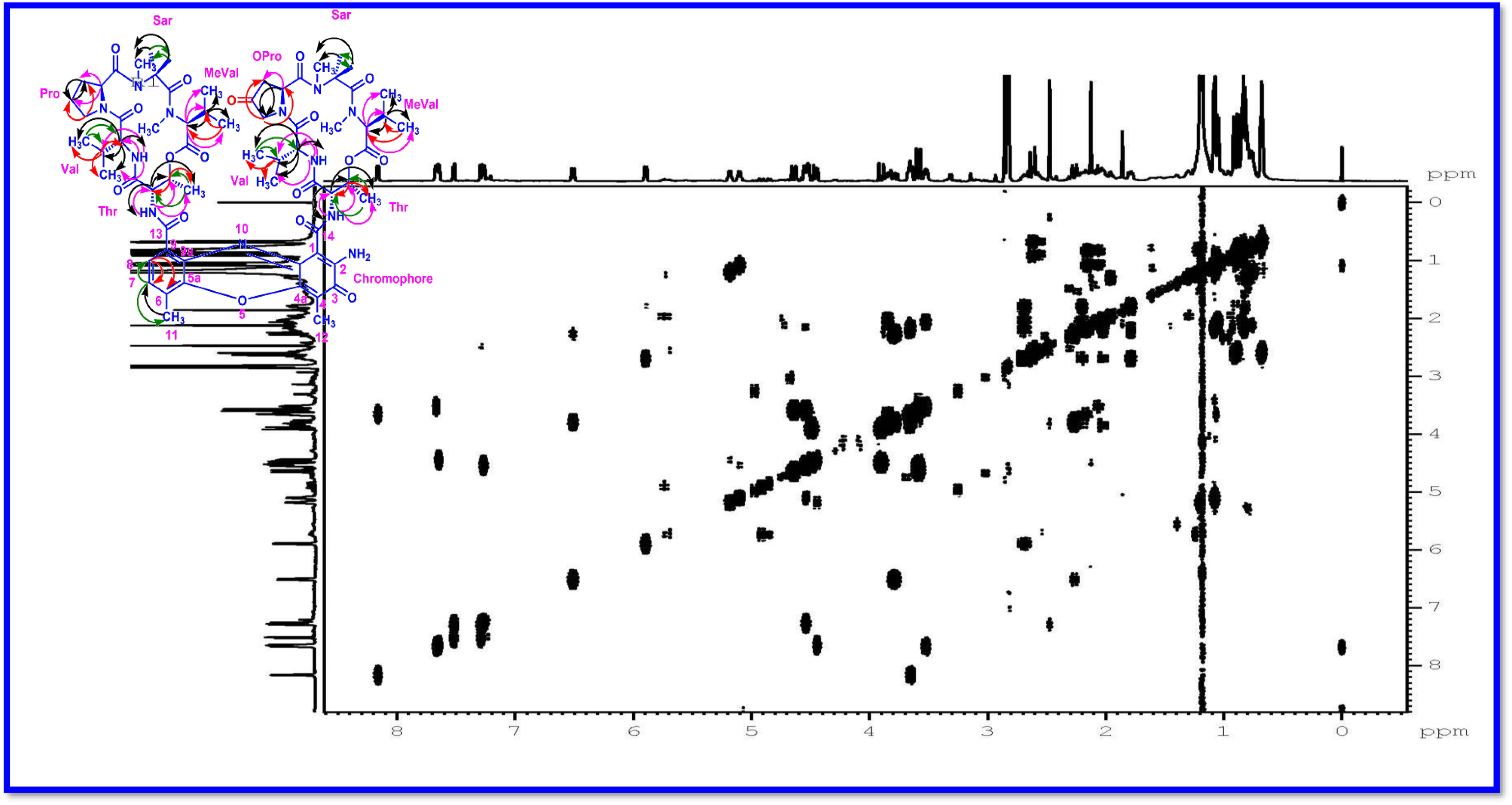
DQF-COSY (500 MHz, CDCl3) Spectrum of Transitmycin (R1)

**Figure S29.**
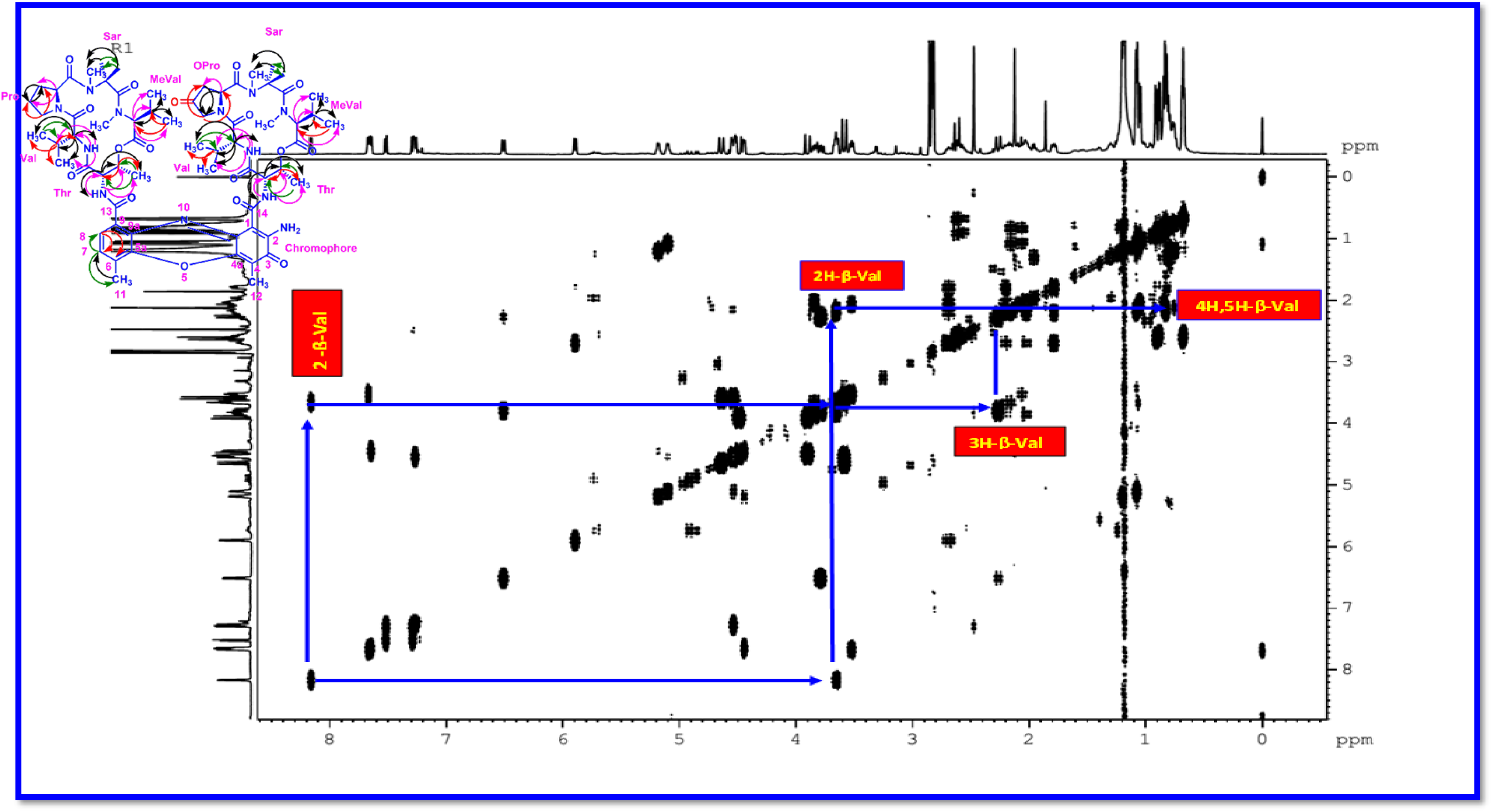
Expansion of DQF-COSY (500 MHz) Spectrum of Transitmycin (R1)

**Figure S30.**
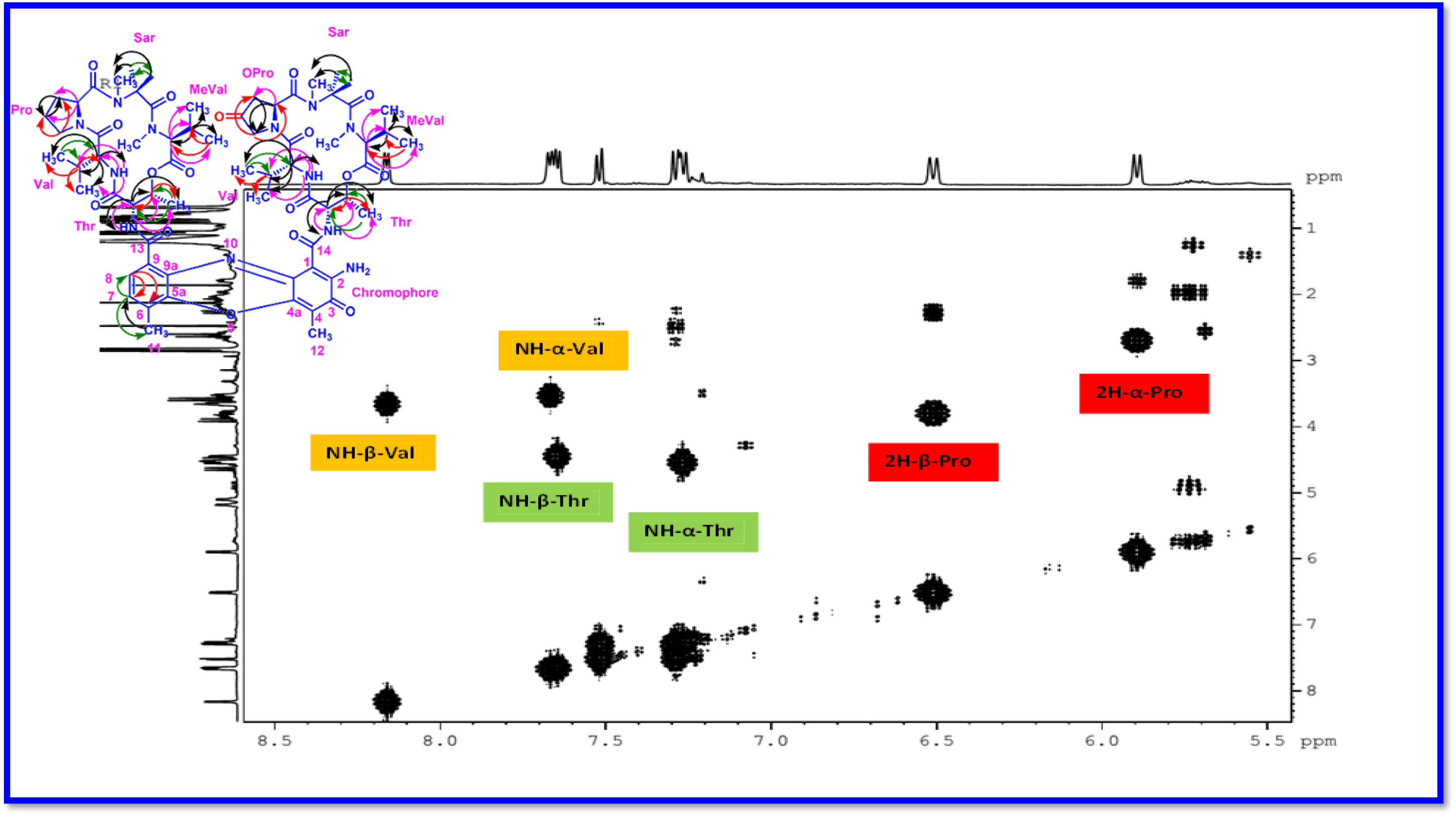
Expansion of DQF-COSY (500 MHz) Spectrum of Transitmycin (R1)

**Figure S31.**
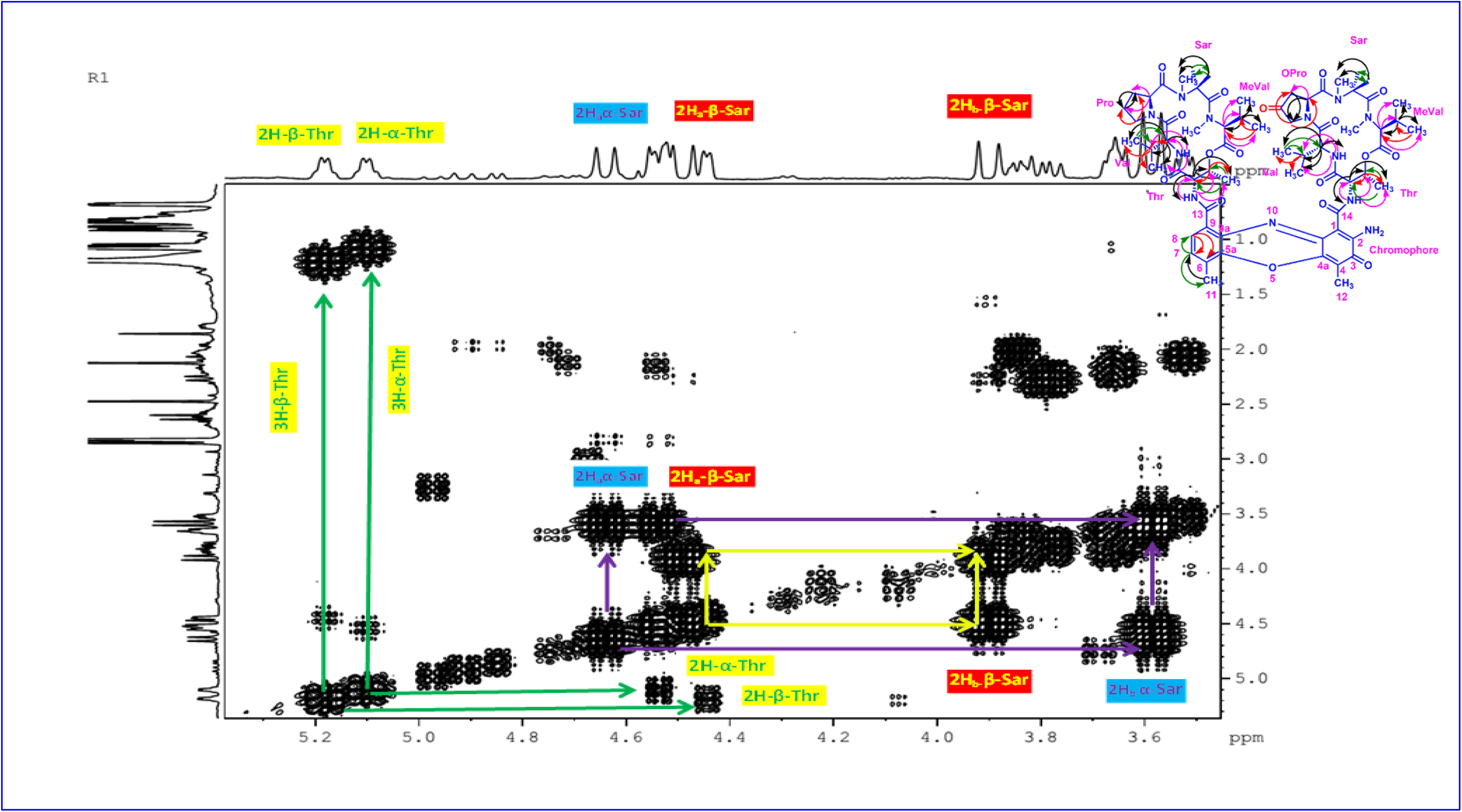
Expansion of DQF-COSY (500 MHz) Spectrum of Transitmycin (R1)

**Figure S32.**
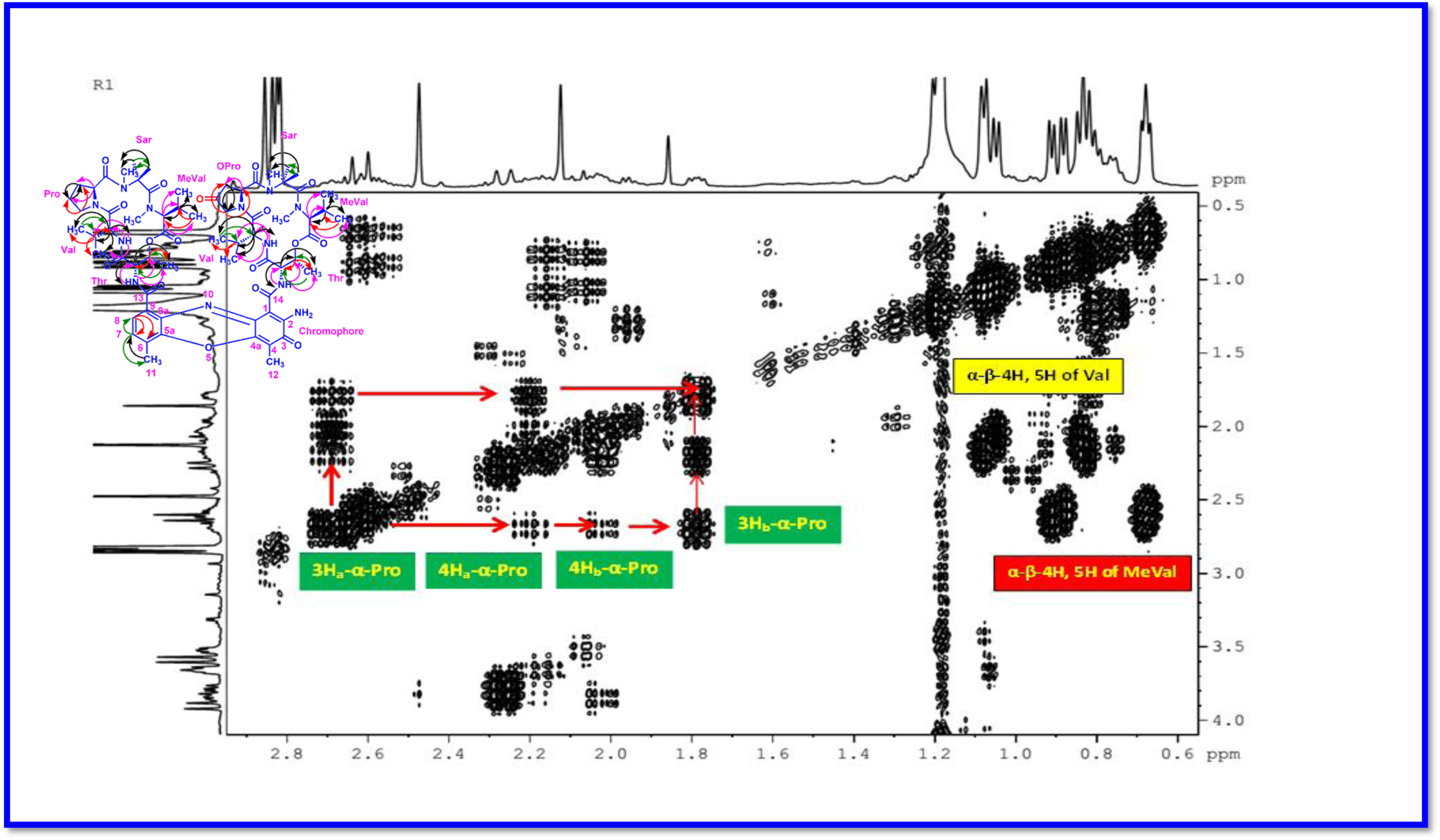
Expansion of DQF-COSY (500 MHz) Spectrum of Transitmycin (R1)

**Figure S33.**
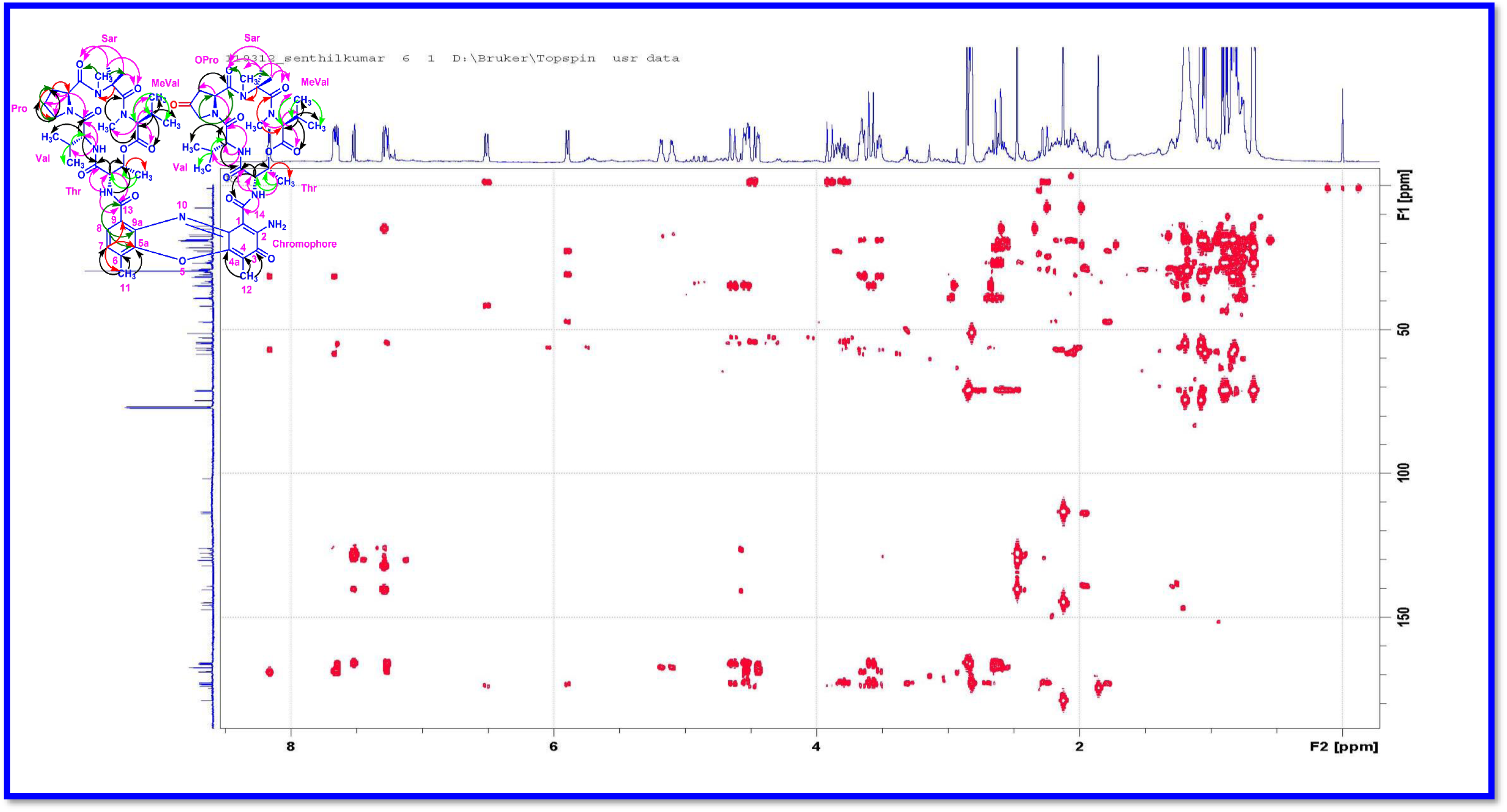
HMBC (500 MHz) Spectrum of Transitmycin (R1)

**Figure S34.**
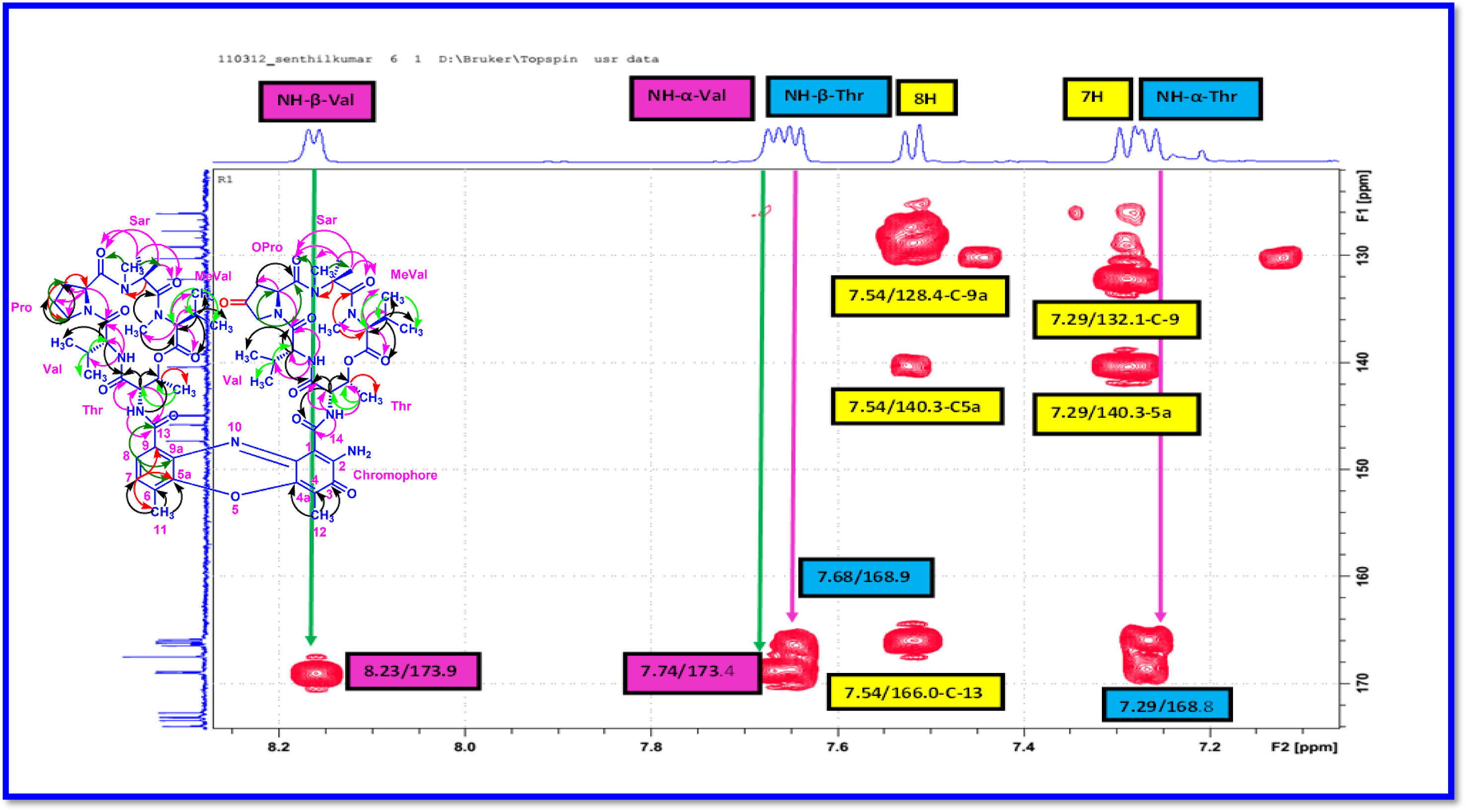
Expansion of HMBC (500 MHz, CDCl_3_) Spectrum of Transitmycin (R1)

**Figure S35.**
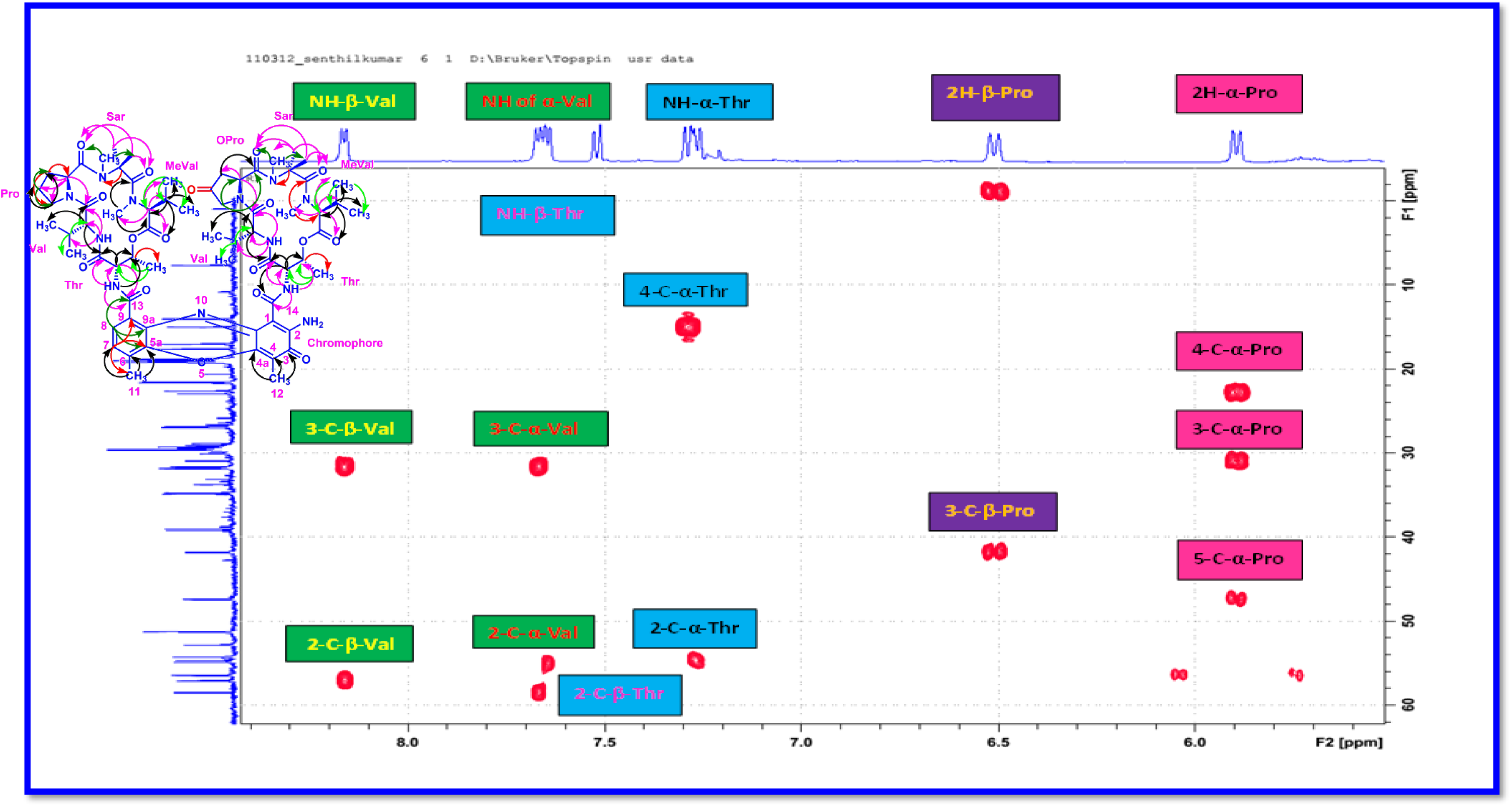
Expansion of HMBC (500 MHz, CDCl_3_) Spectrum of Transitmycin (R1)

**Figure S36.**
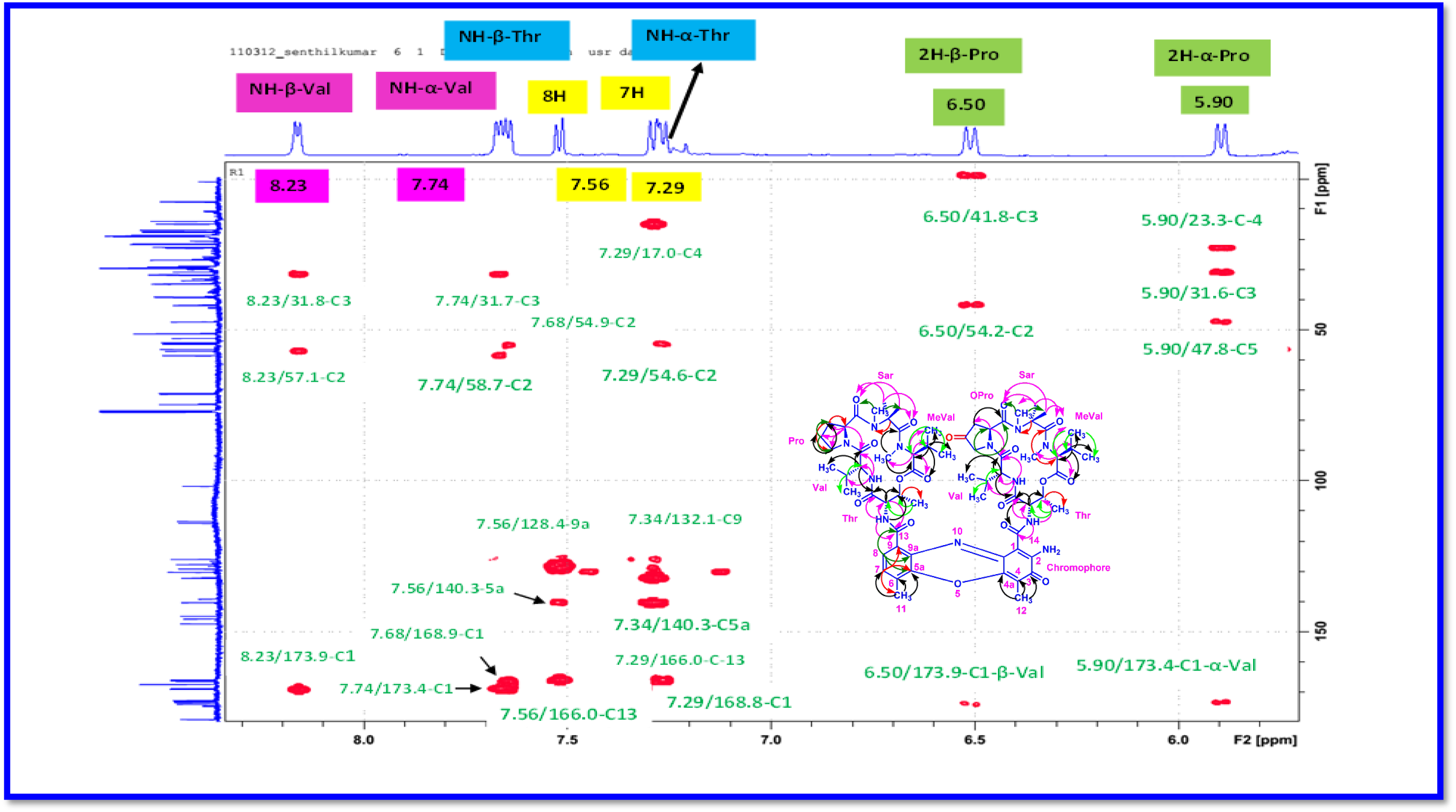
Expansion of HMBC (500 MHz, CDCl_3_) Spectrum of Transitmycin (R1)

**Figure S37.**
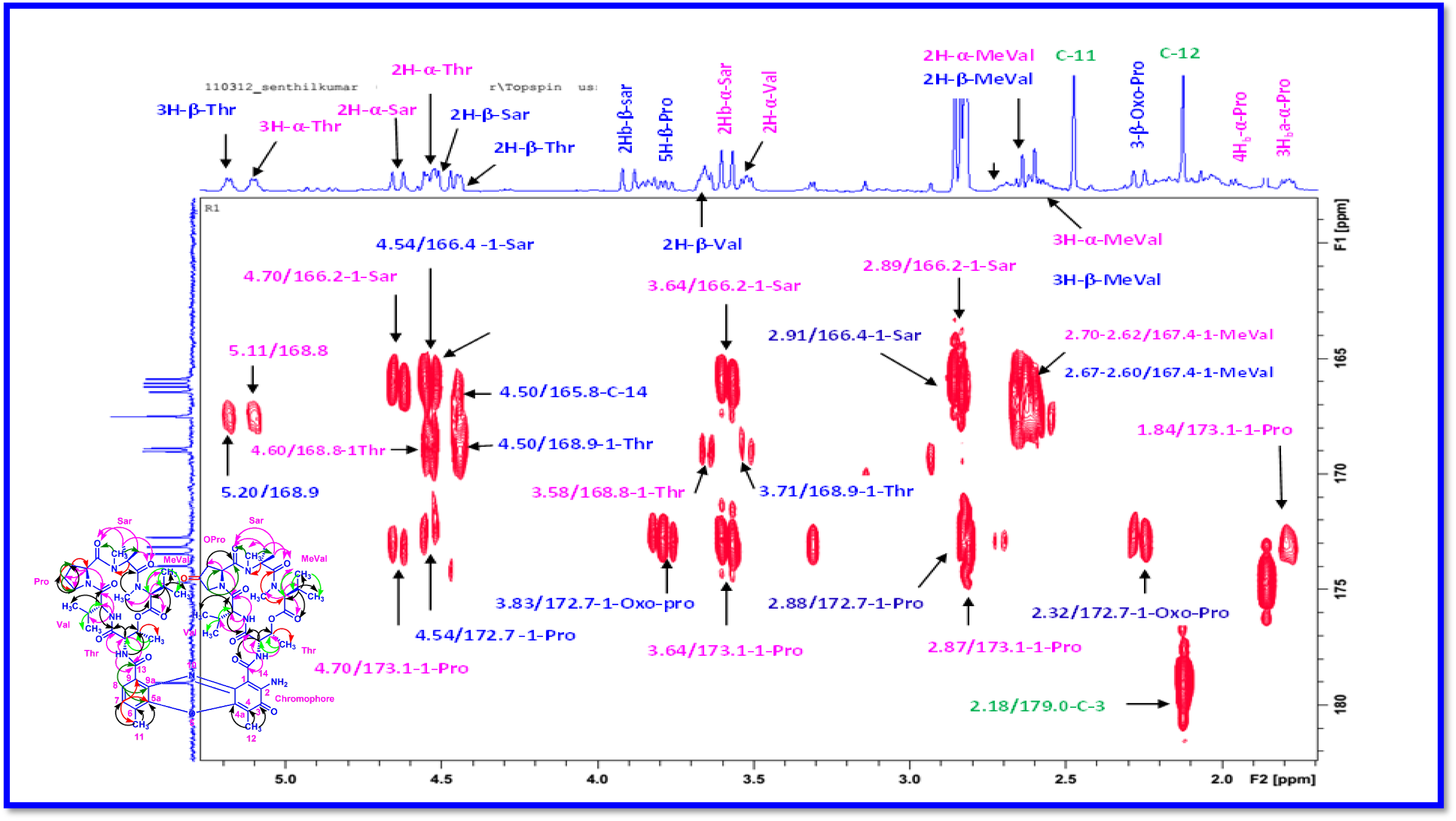
Expansion of HMBC (500 MHz, CDCl_3_) Spectrum of Transitmycin (R1)

**Figure S38.**
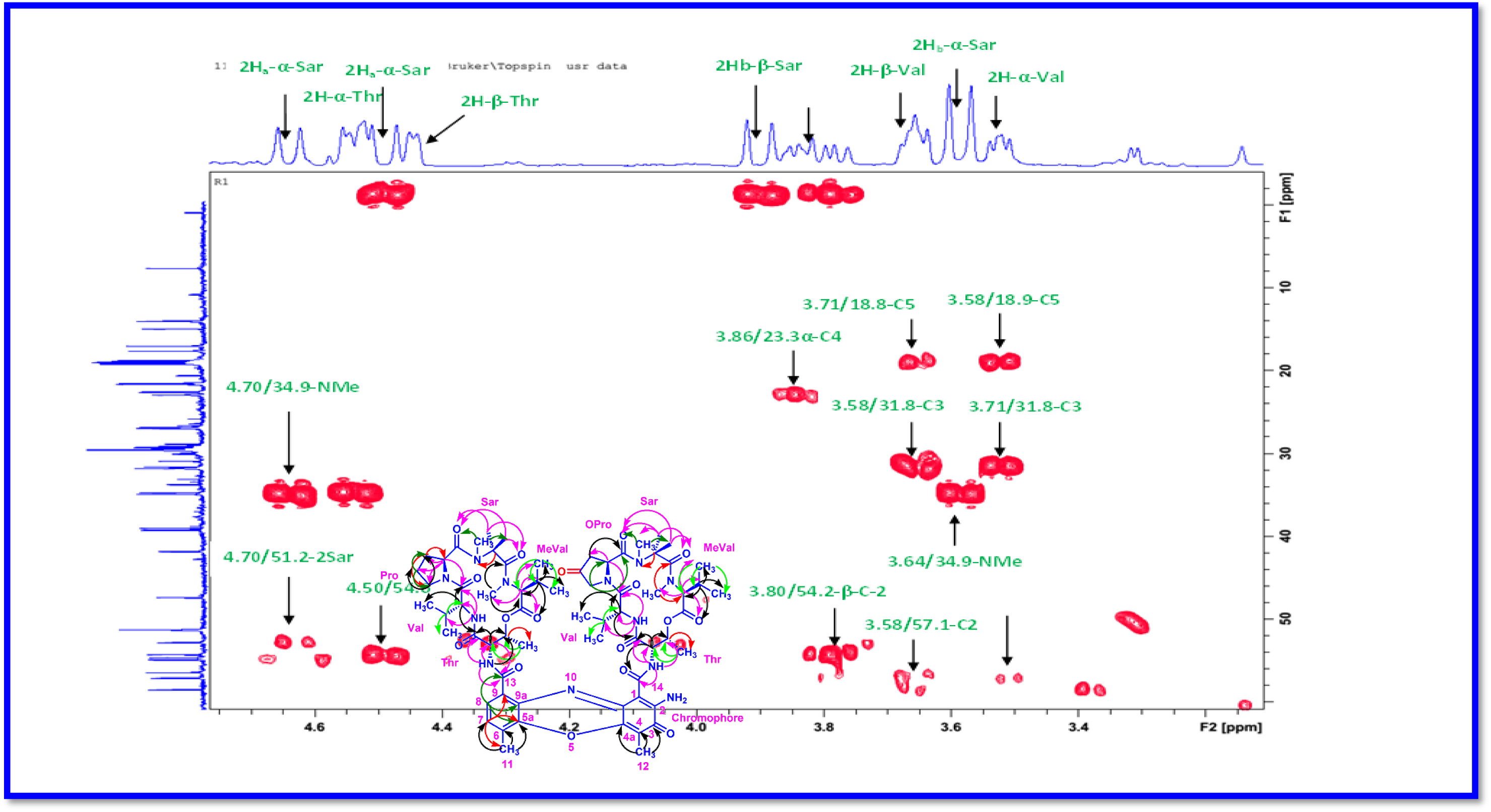
Expansion of HMBC (500 MHz, CDCl_3_) Spectrum of Transitmycin (R1)

**Figure S39.**
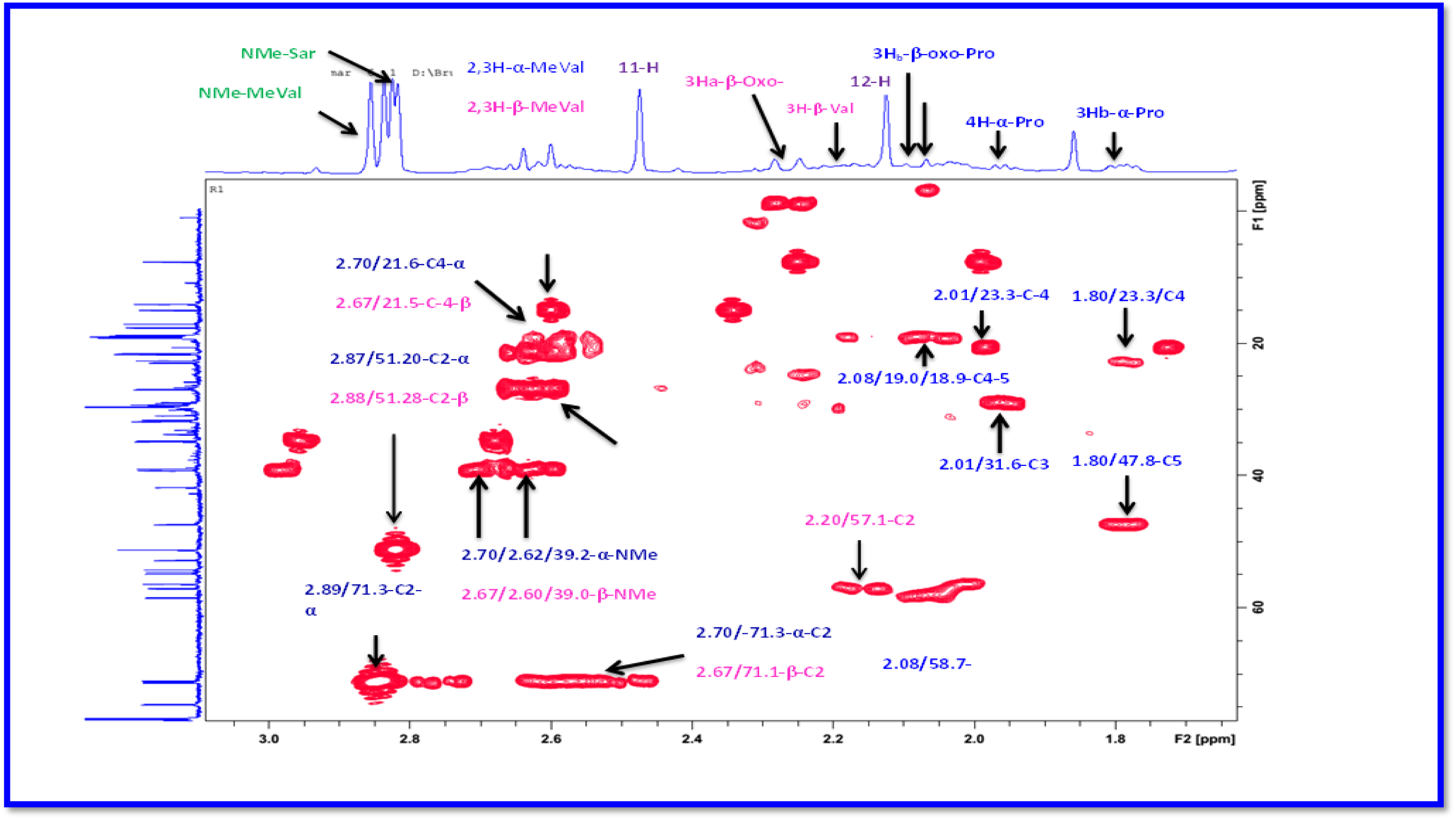
Expansion of HMBC (500 MHz, CDCl_3_) Spectrum of Transitmycin (R1)

**Figure S40.**
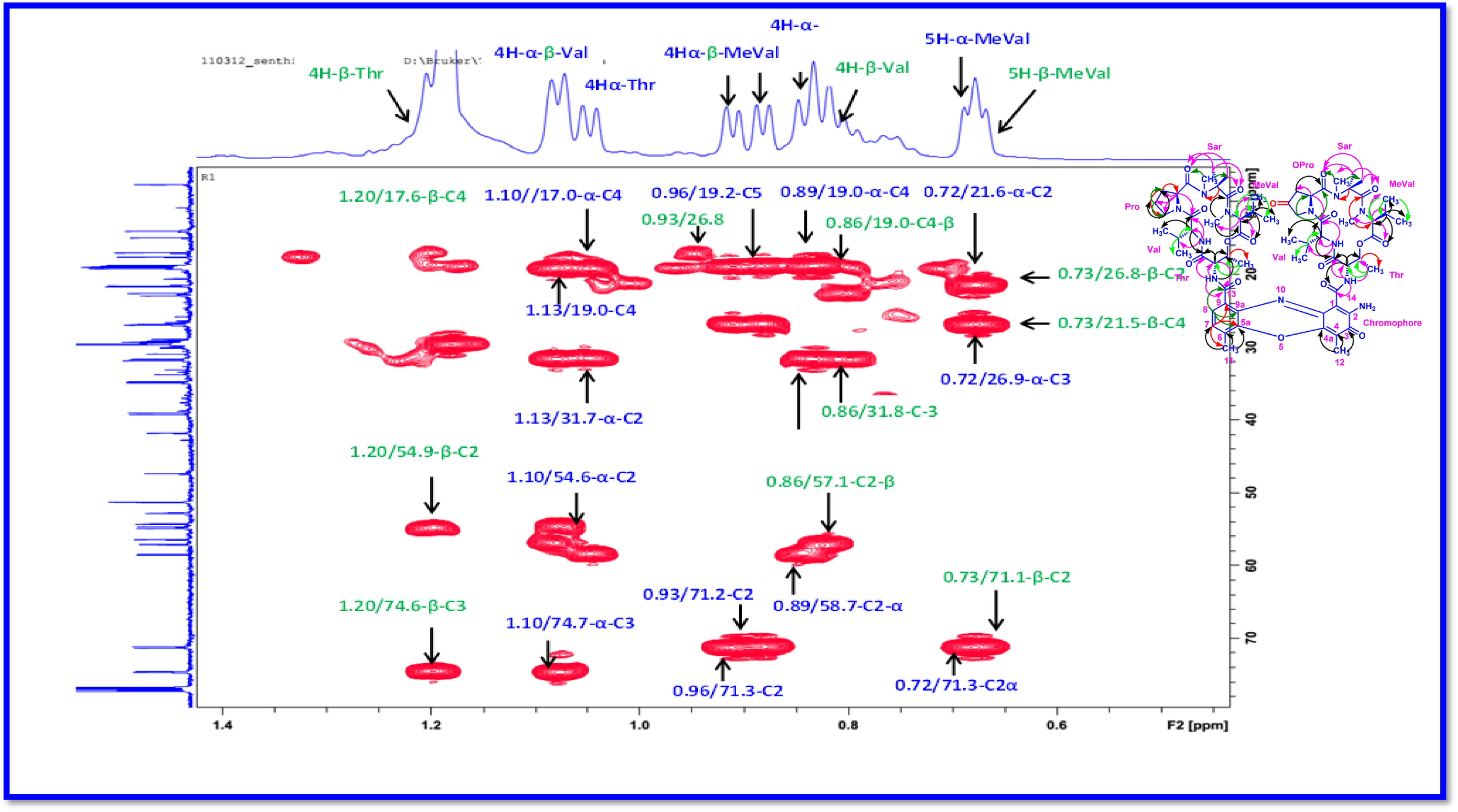
Expansion of HMBC (500 MHz, CDCl_3_) Spectrum of Transitmycin (R1)

**Figure S41.**
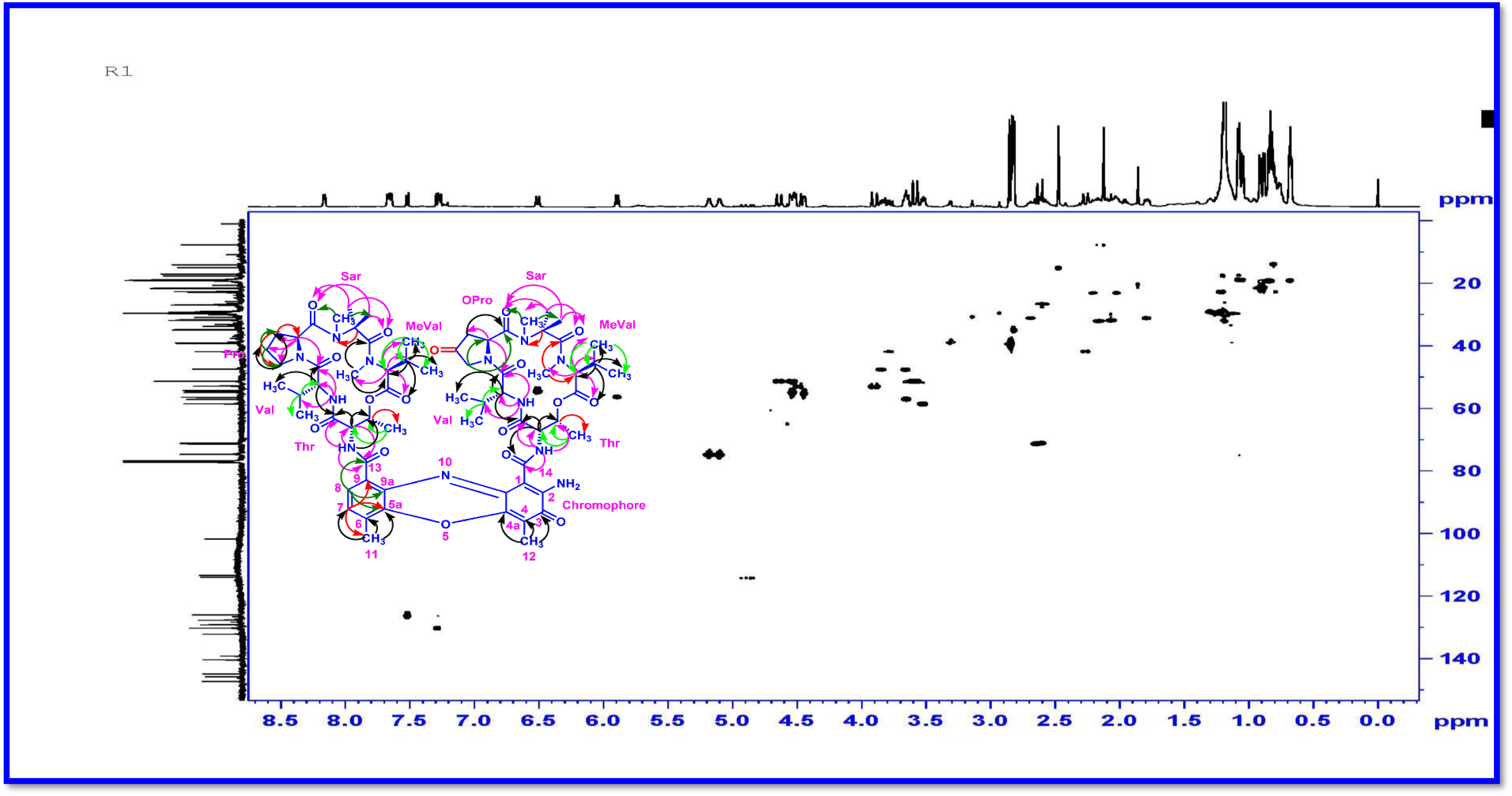
HSQC (500 MHz, CDCl_3_) Spectrum of Transitmycin (R1)

**Figure S42.**
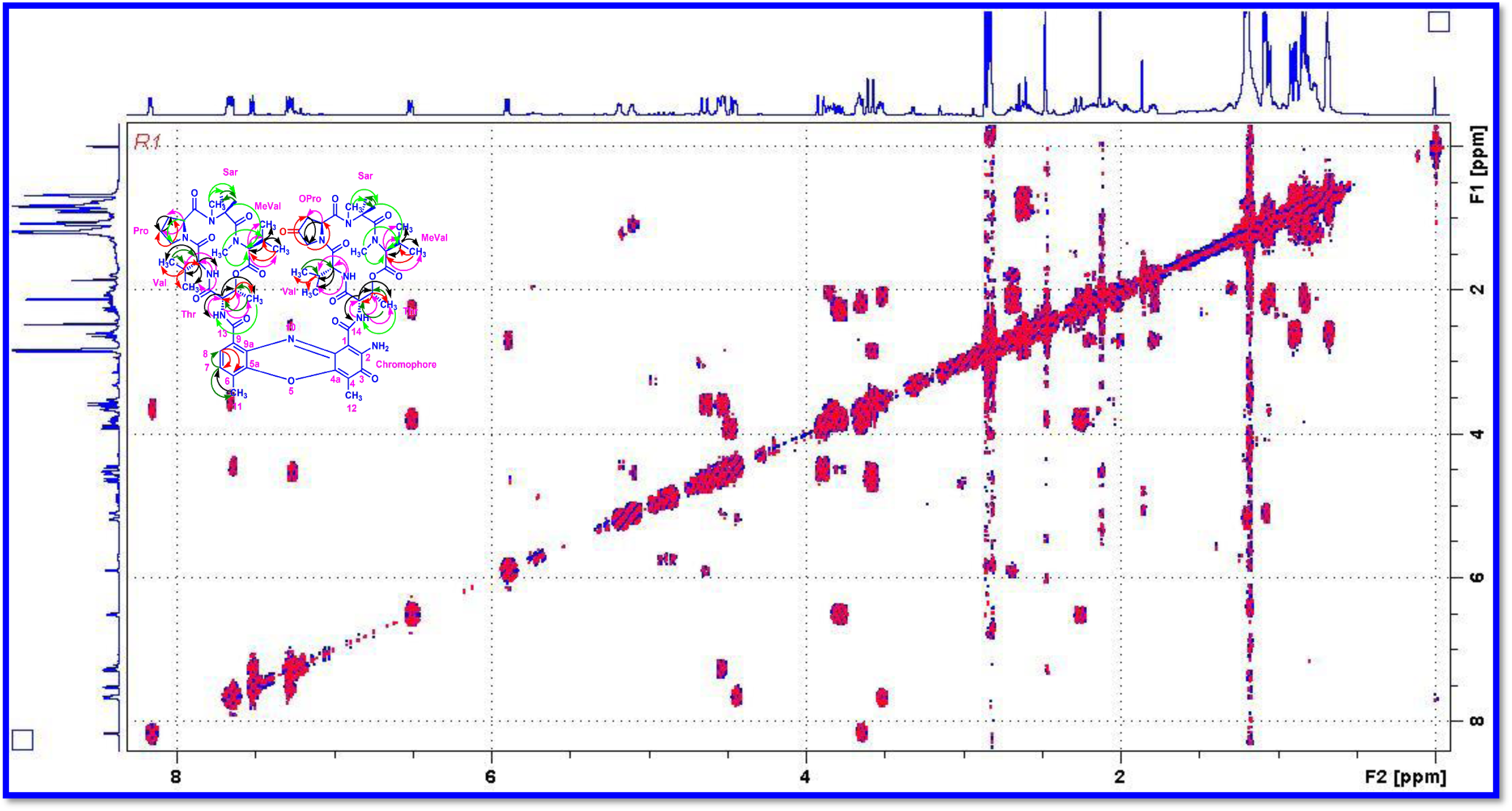
TOCSY (500 MHz, CDCl_3_) Spectrum of Transitmycin (R1)

**Figure S43.**
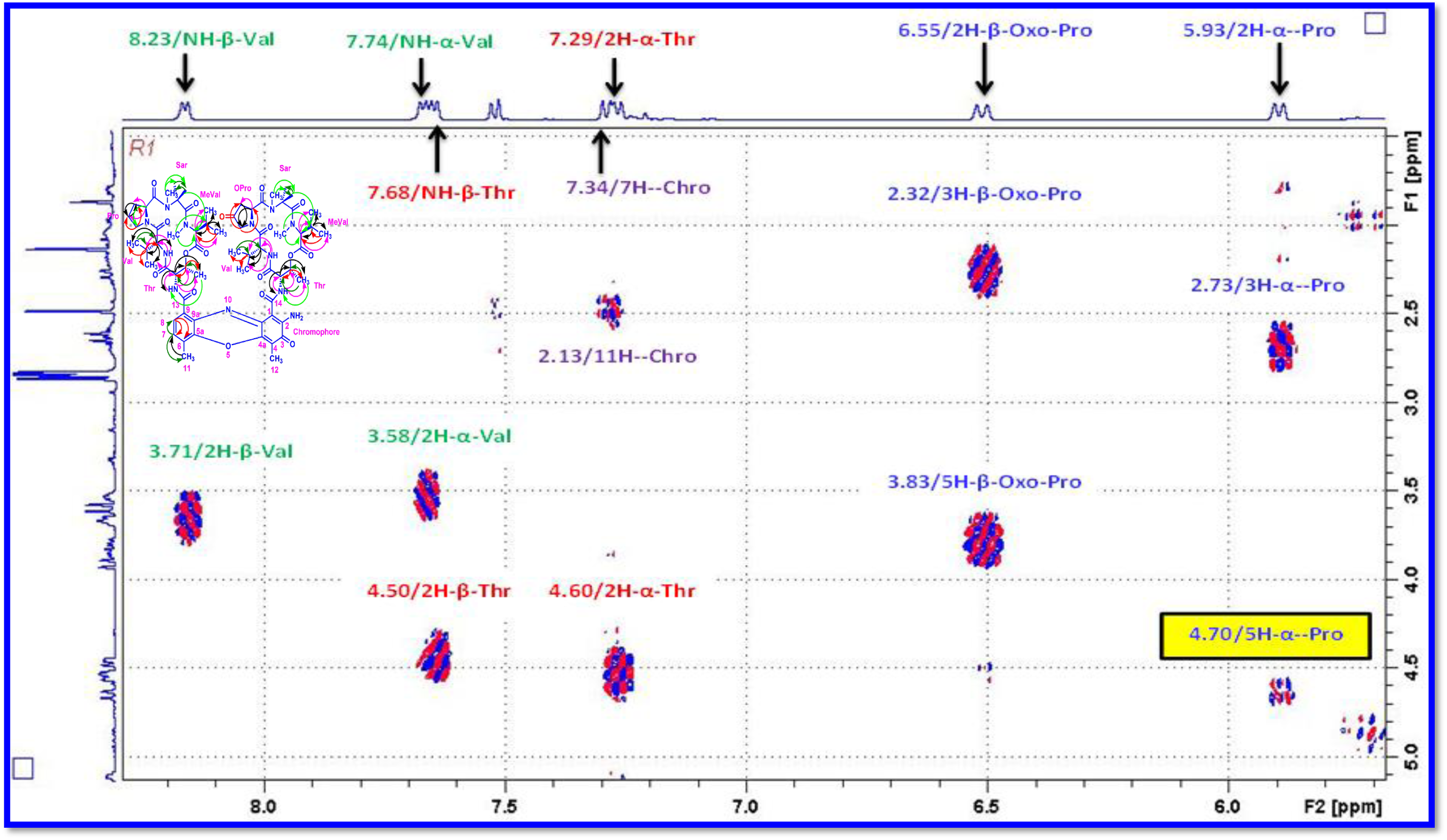
Expansion of TOCSY (500 MHz, CDCl_3_) Spectrum of Transitmycin (R1)

**Figure S44.**
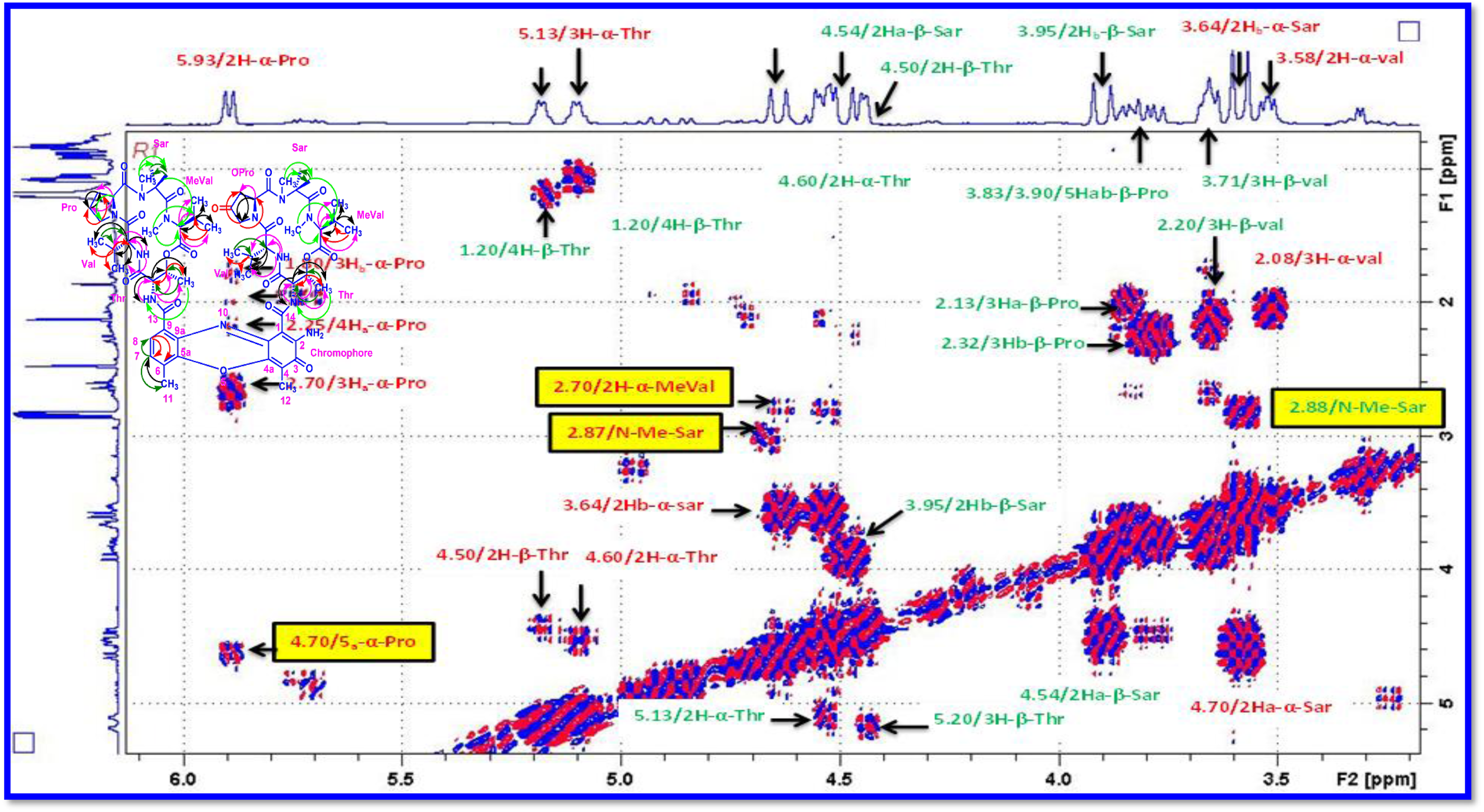
Expansion of TOCSY (500 MHz, CDCl_3_) Spectrum of Transitmycin (R1)

**Figure S45.**
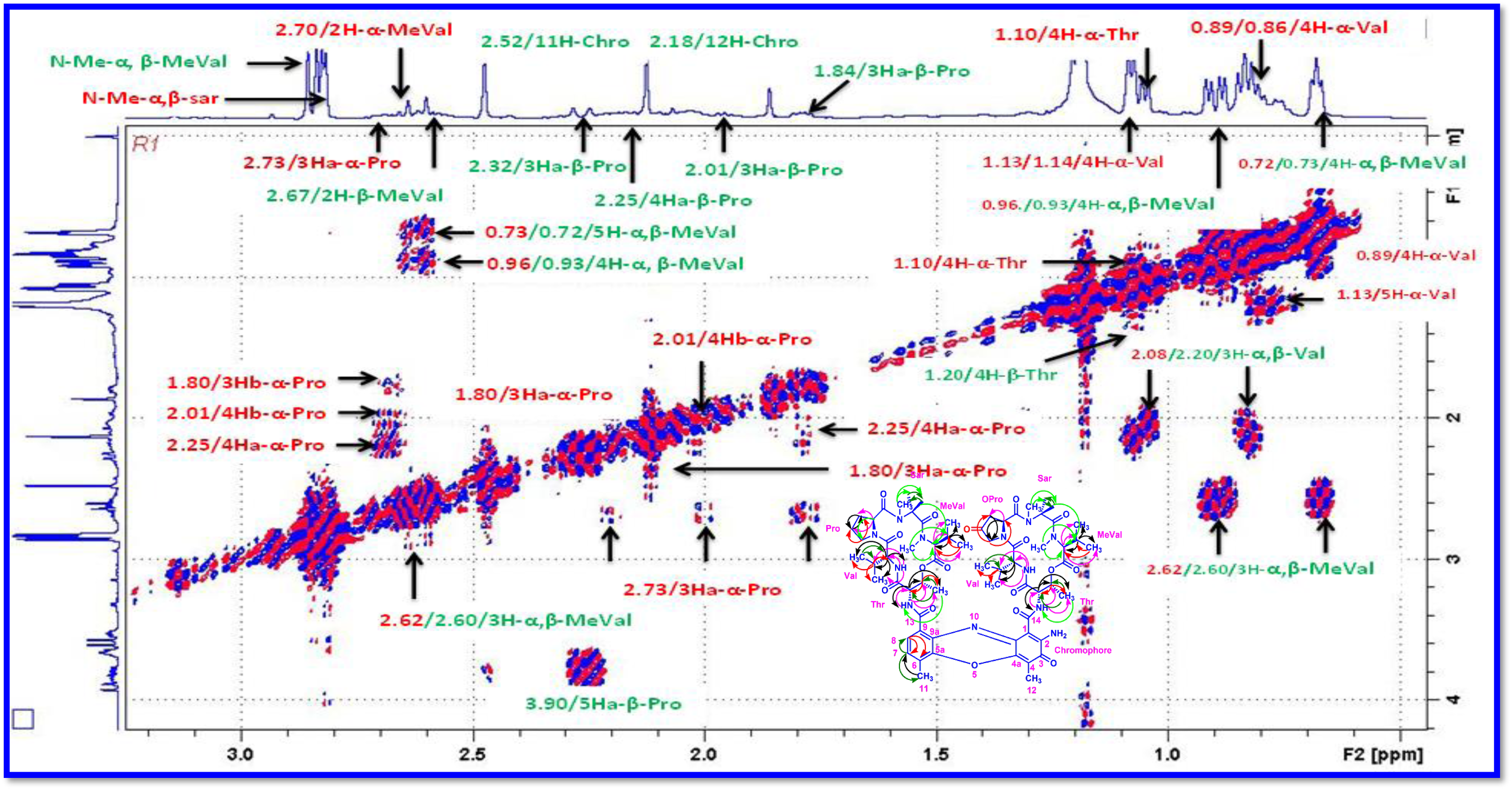
Expansion of TOCSY (500 MHz, CDCl_3_) Spectrum of Transitmycin (R1)

**Figure S46.**
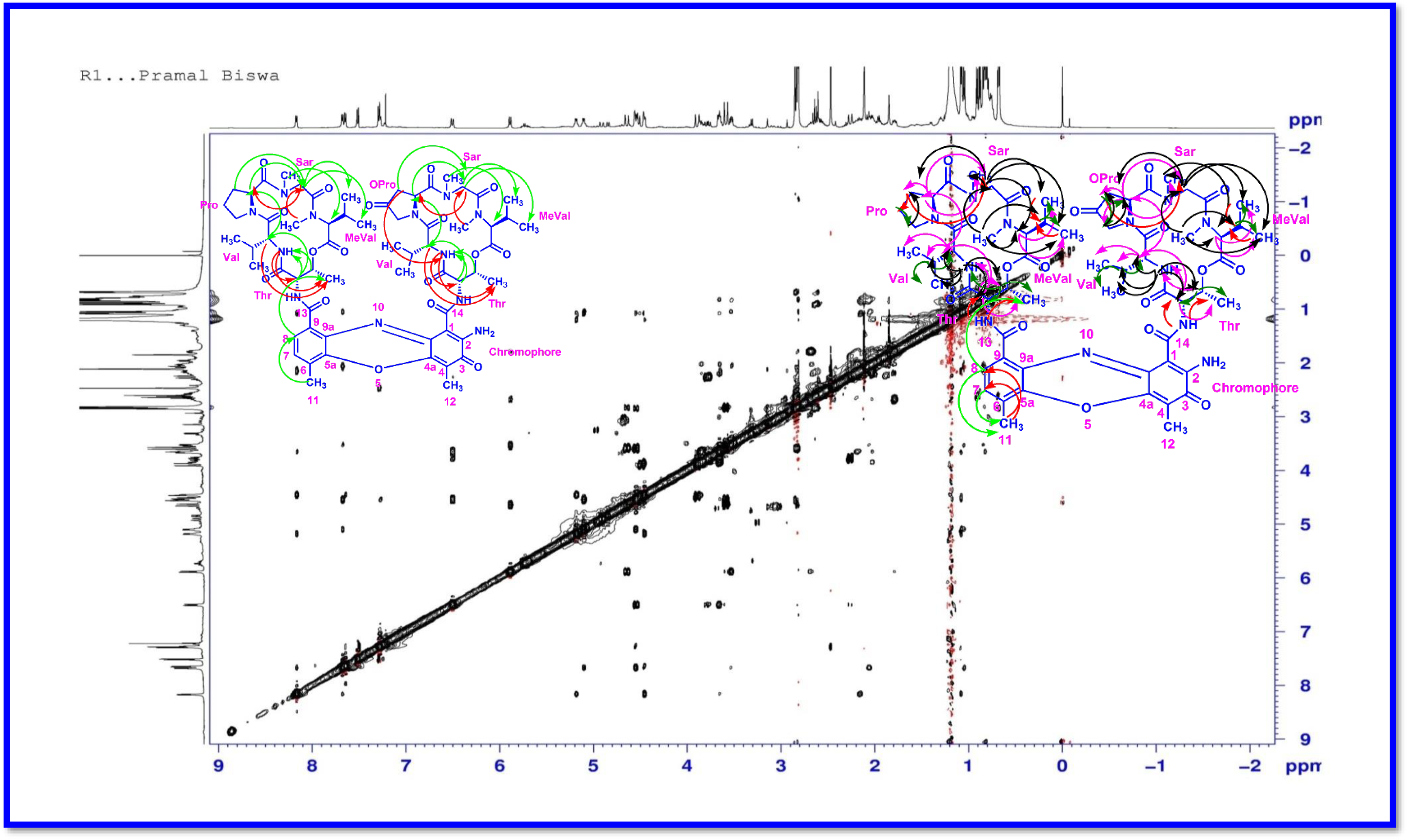
NOESY (500 MHz, CDCl_3_) Spectrum of Transitmycin (R1)

**Figure S47.**
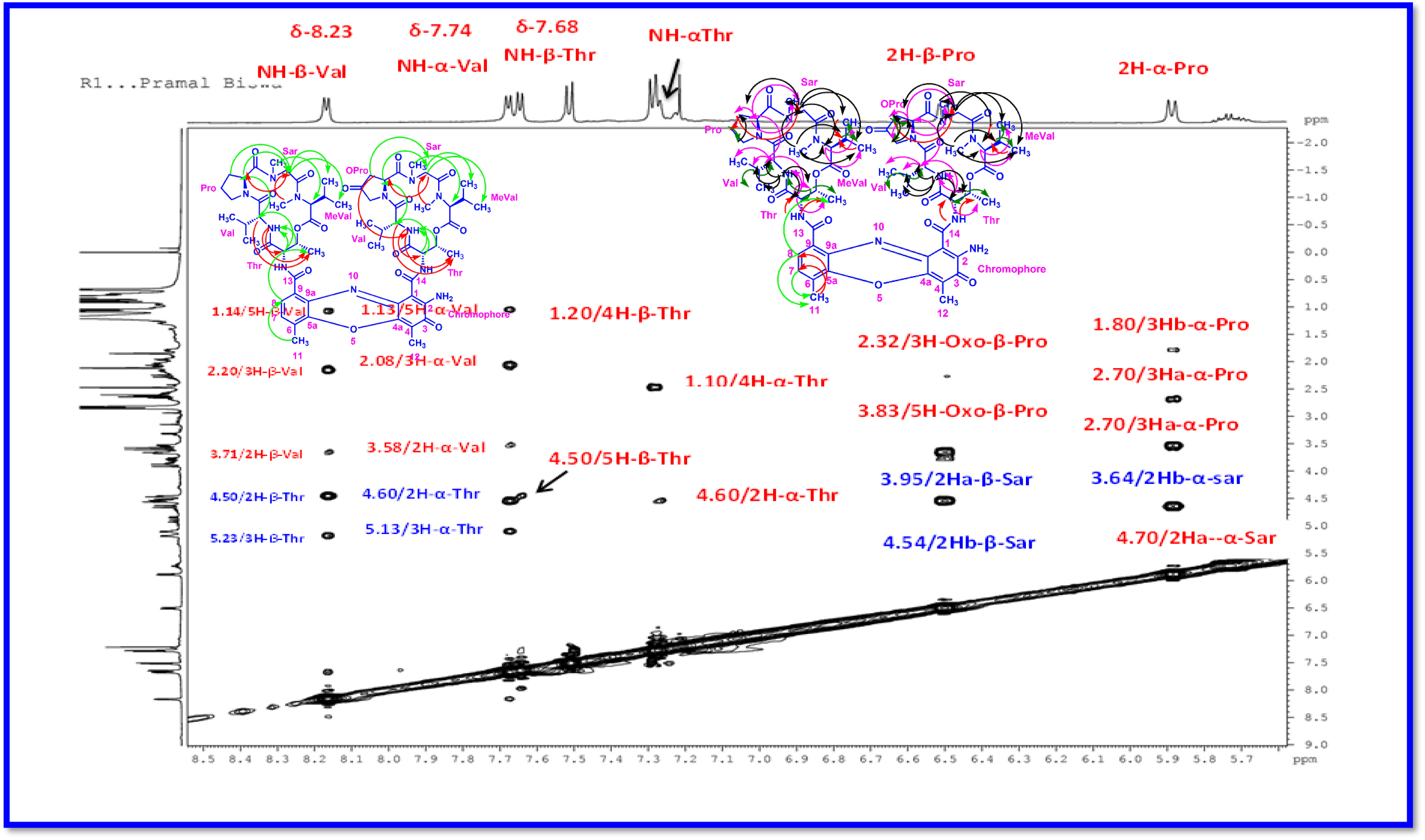
Expansion of NOESY (500 MHz, CDCl_3_) Spectrum of Transitmycin (R1)

**Figure S48.**
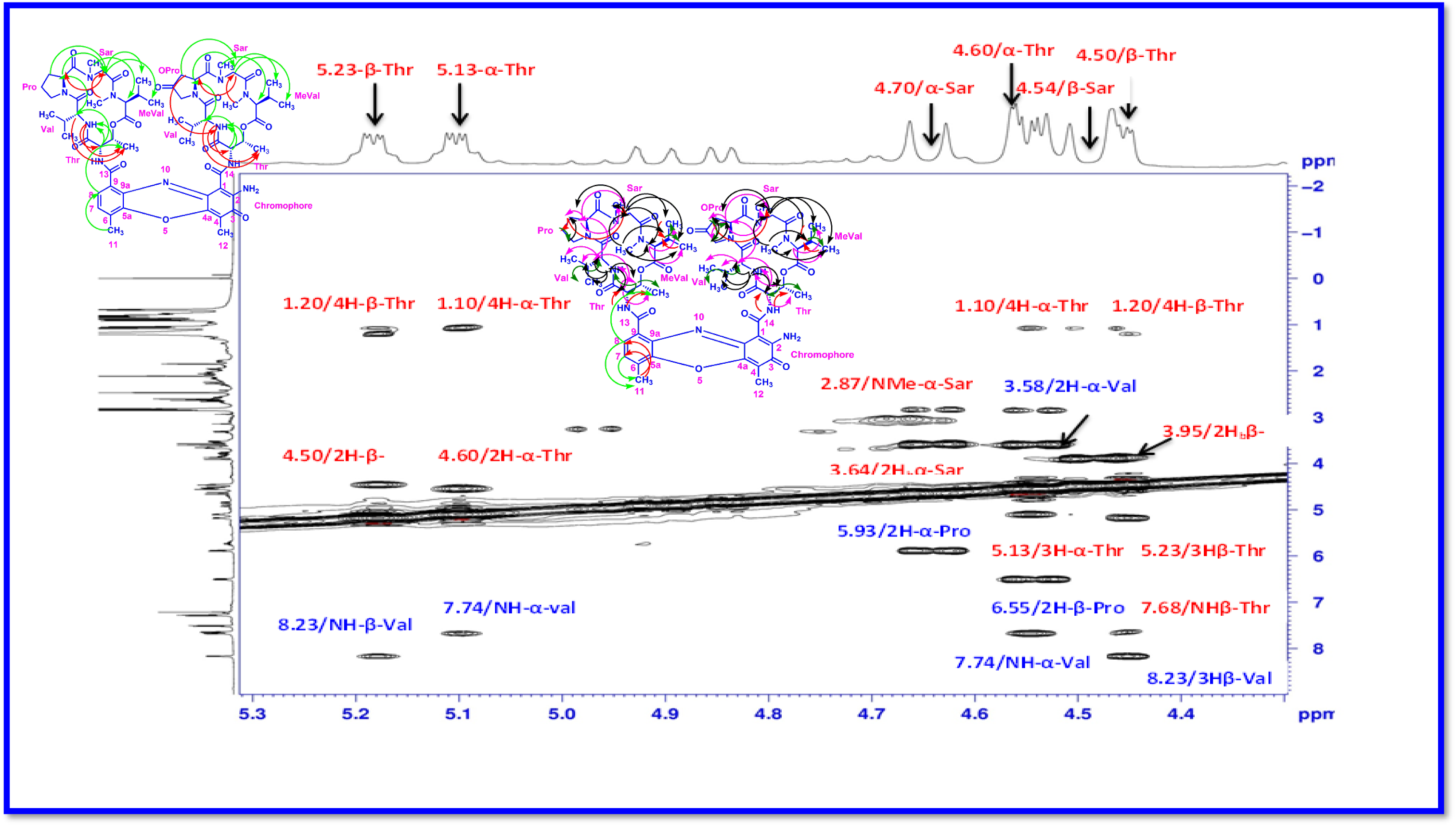
Expansion of NOESY (500 MHz, CDCl_3_) Spectrum of Transitmycin (R1)

**Figure S49.**
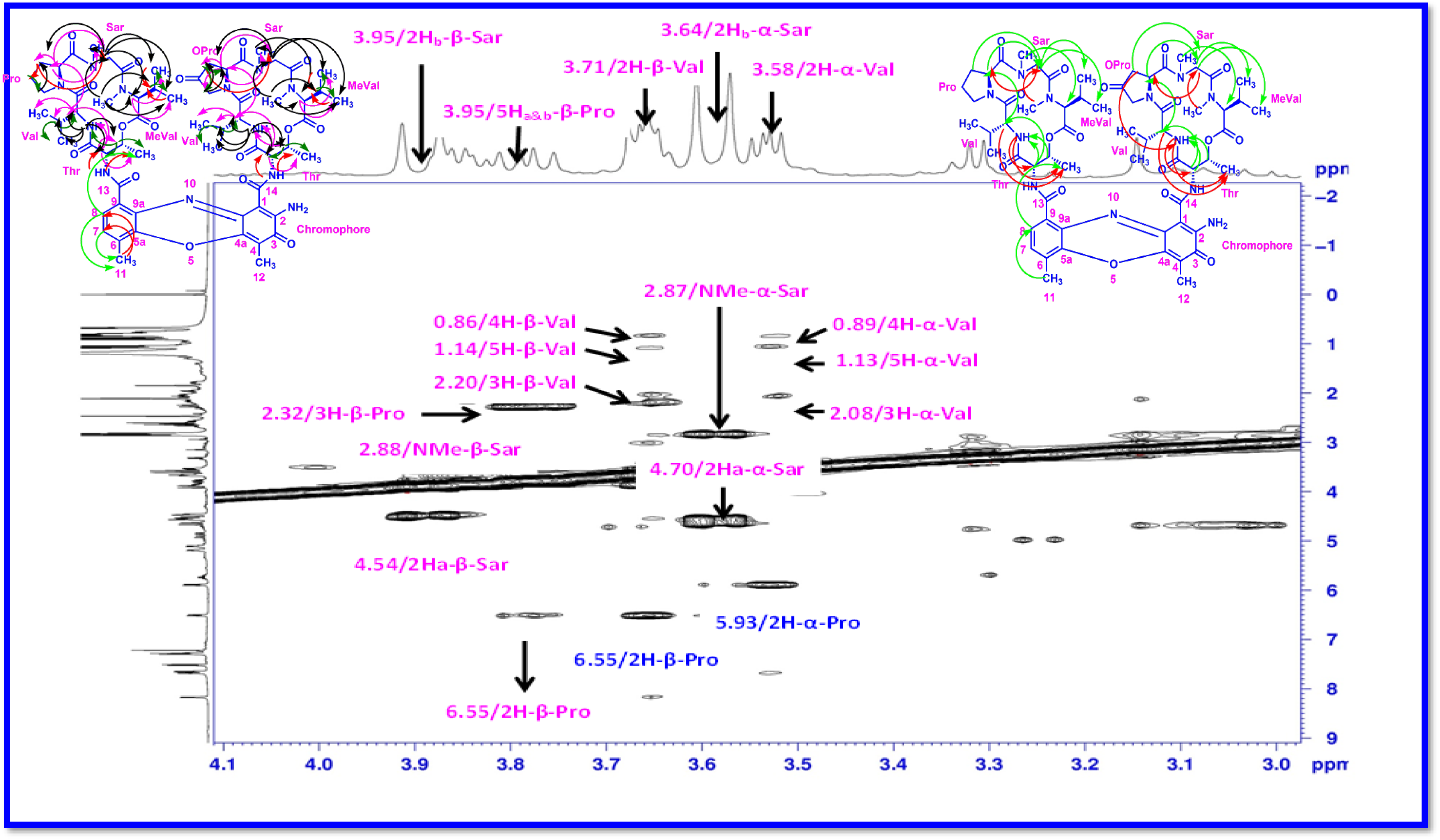
Expansion of NOESY (500 MHz, CDCl_3_) Spectrum of Transitmycin (R1)

**Figure S50.**
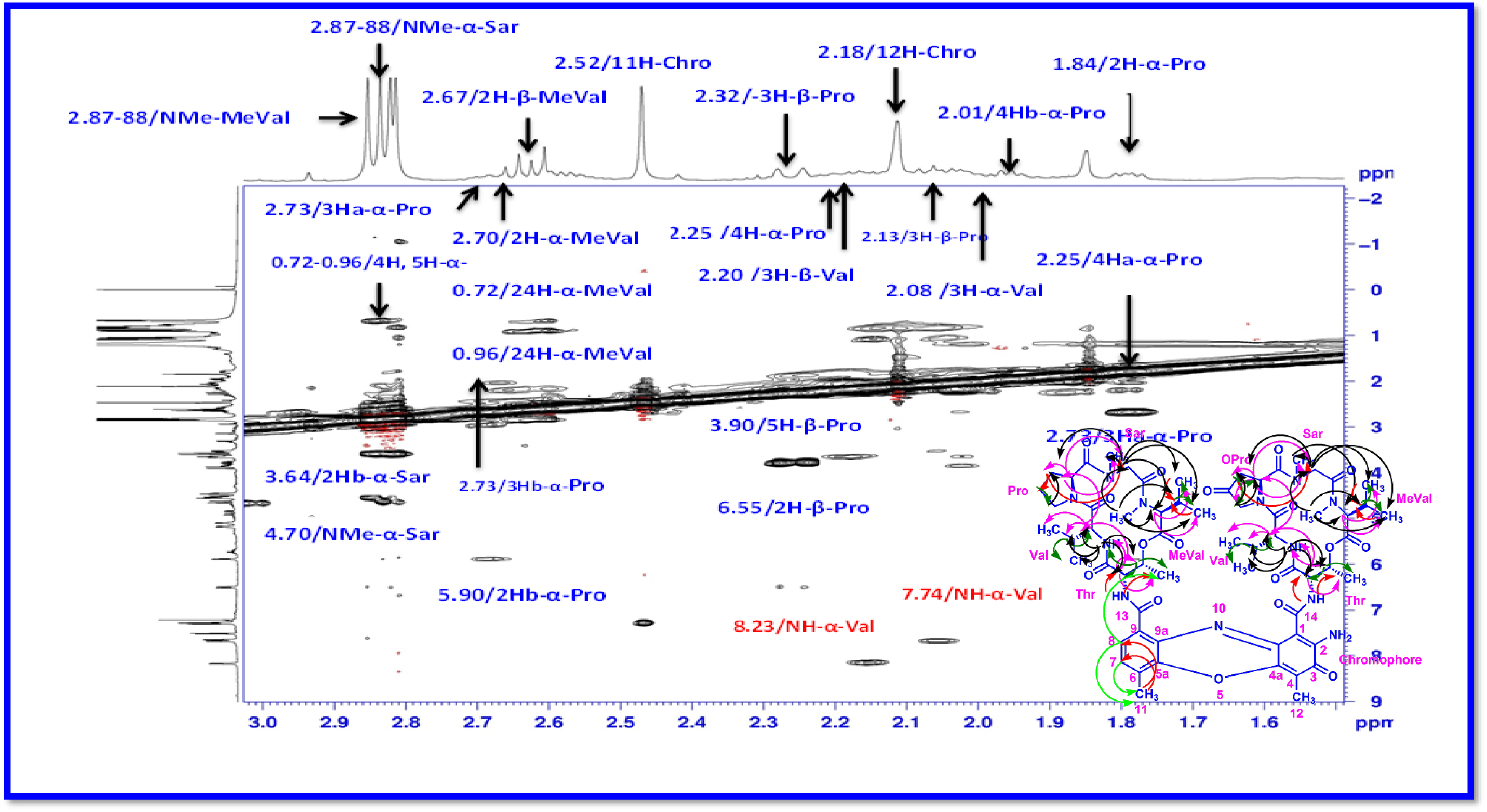
Expansion of NOESY (500 MHz, CDCl_3_) Spectrum of Transitmycin (R1)

**Figure S51.**
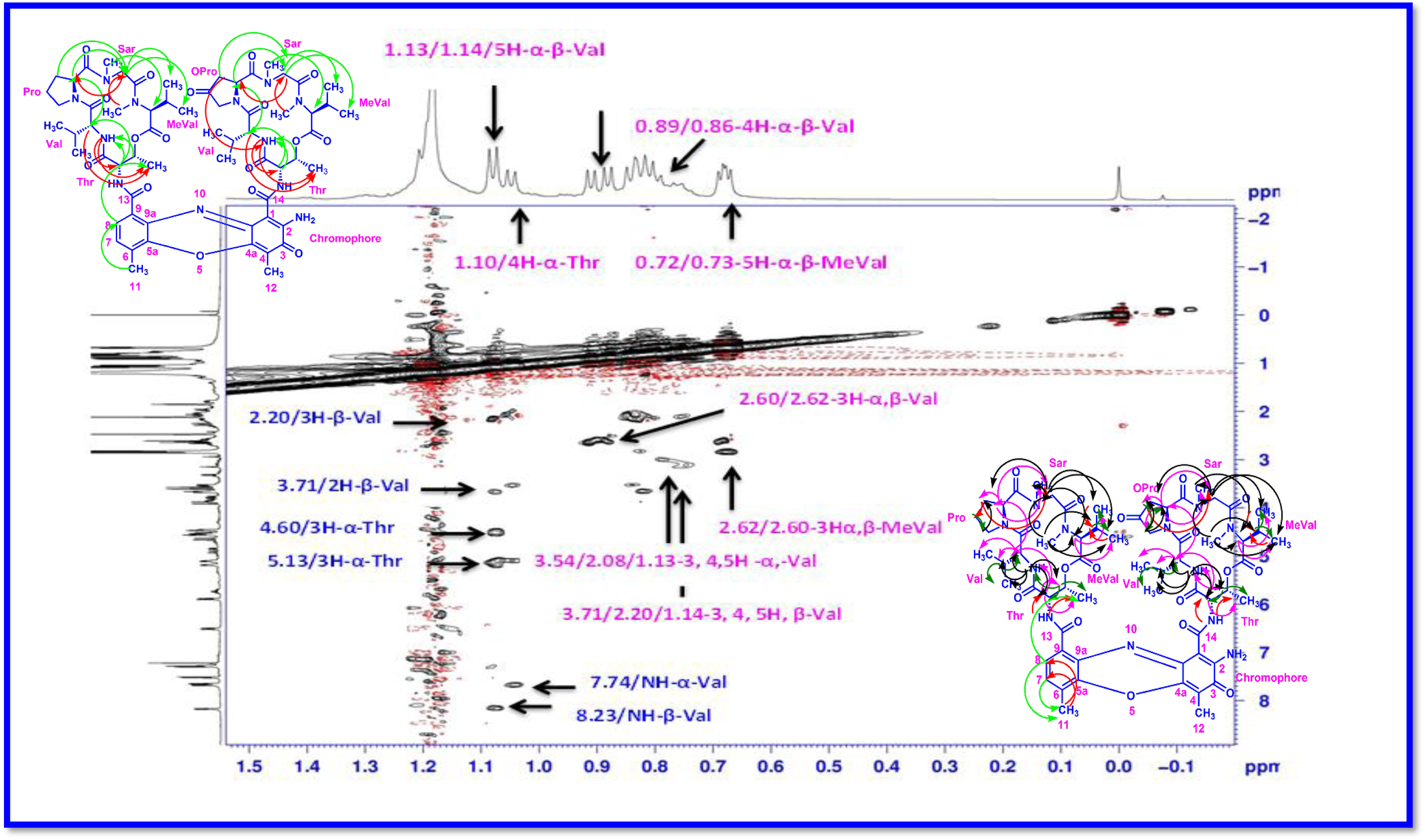
Expansion of NOESY (500 MHz, CDCl_3_) Spectrum of Transitmycin (R1)

**Figure S52.**
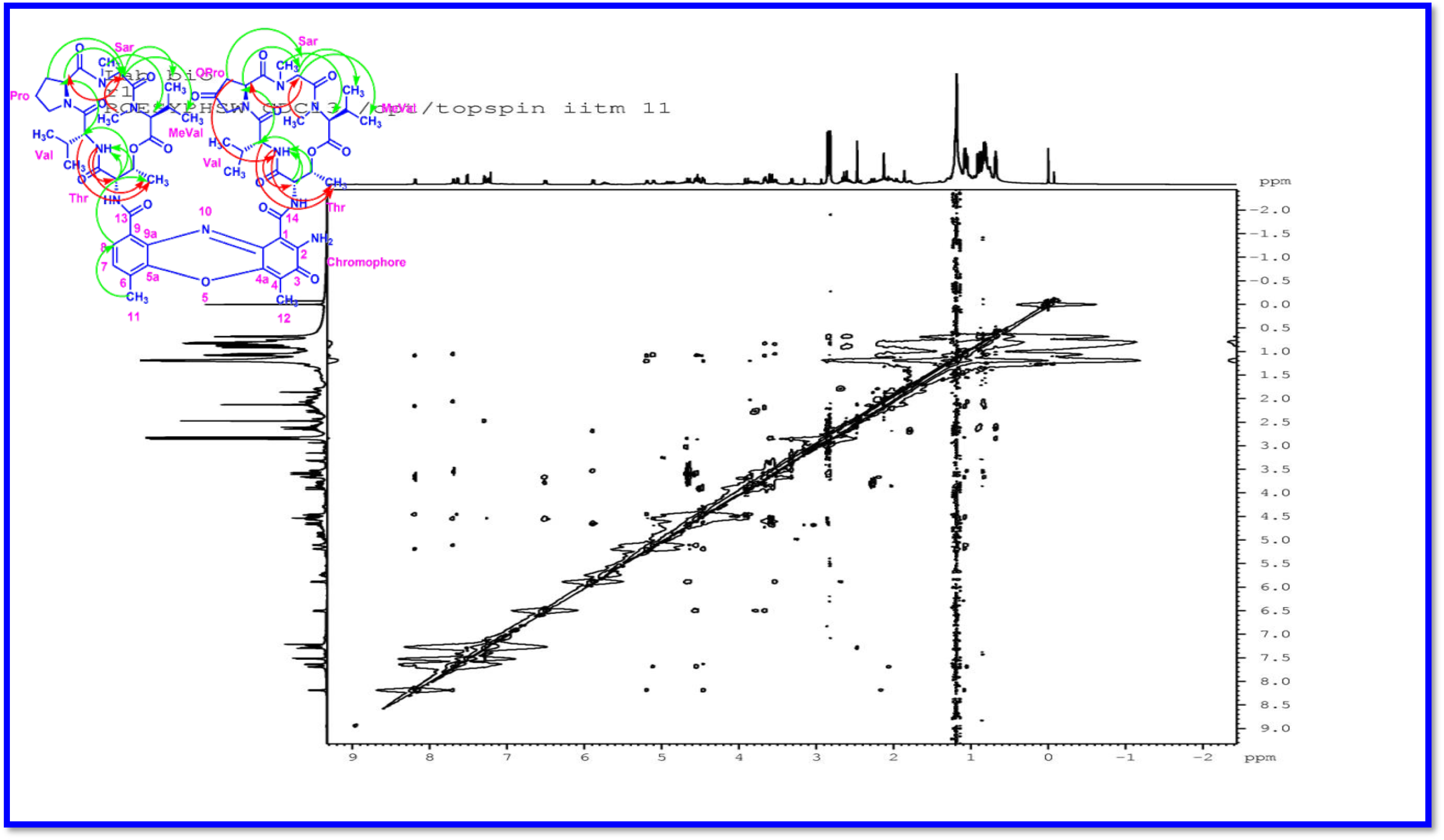
ROESY (500 MHz) Spectrum of Transitmycin (R1)

**Table S5:**
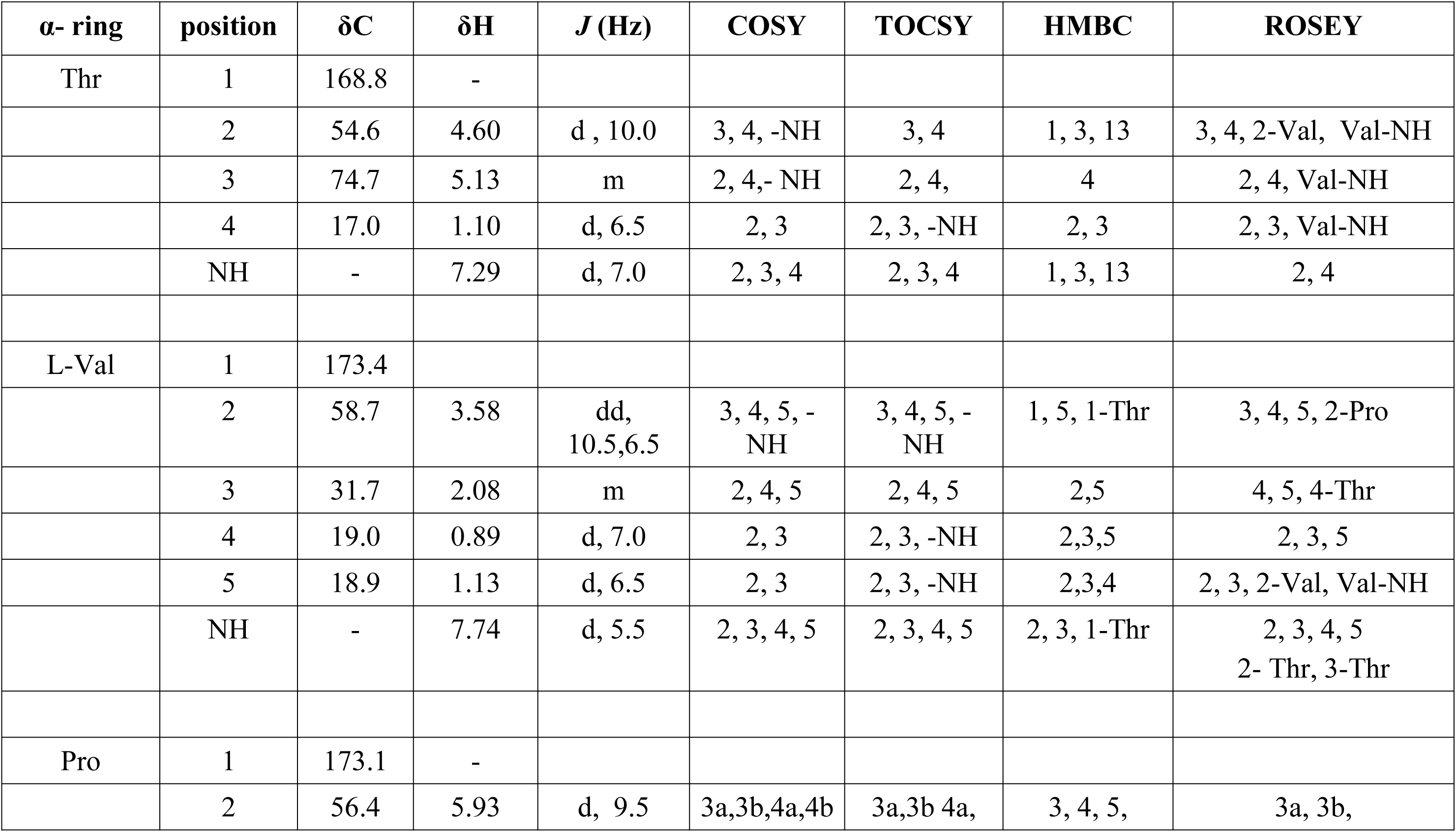
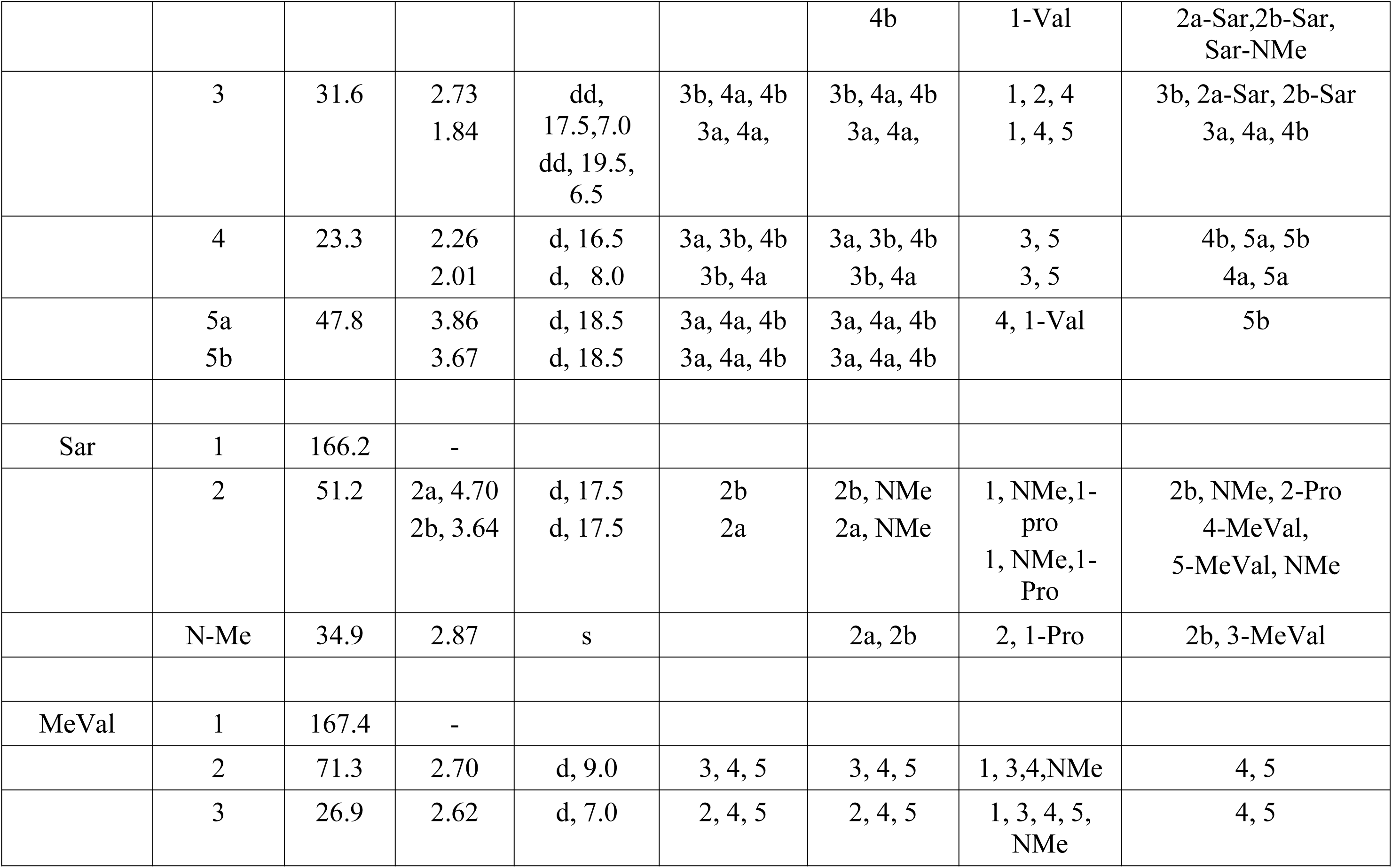
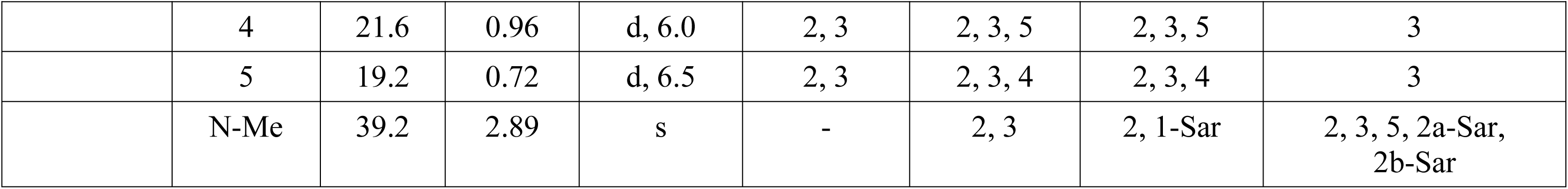
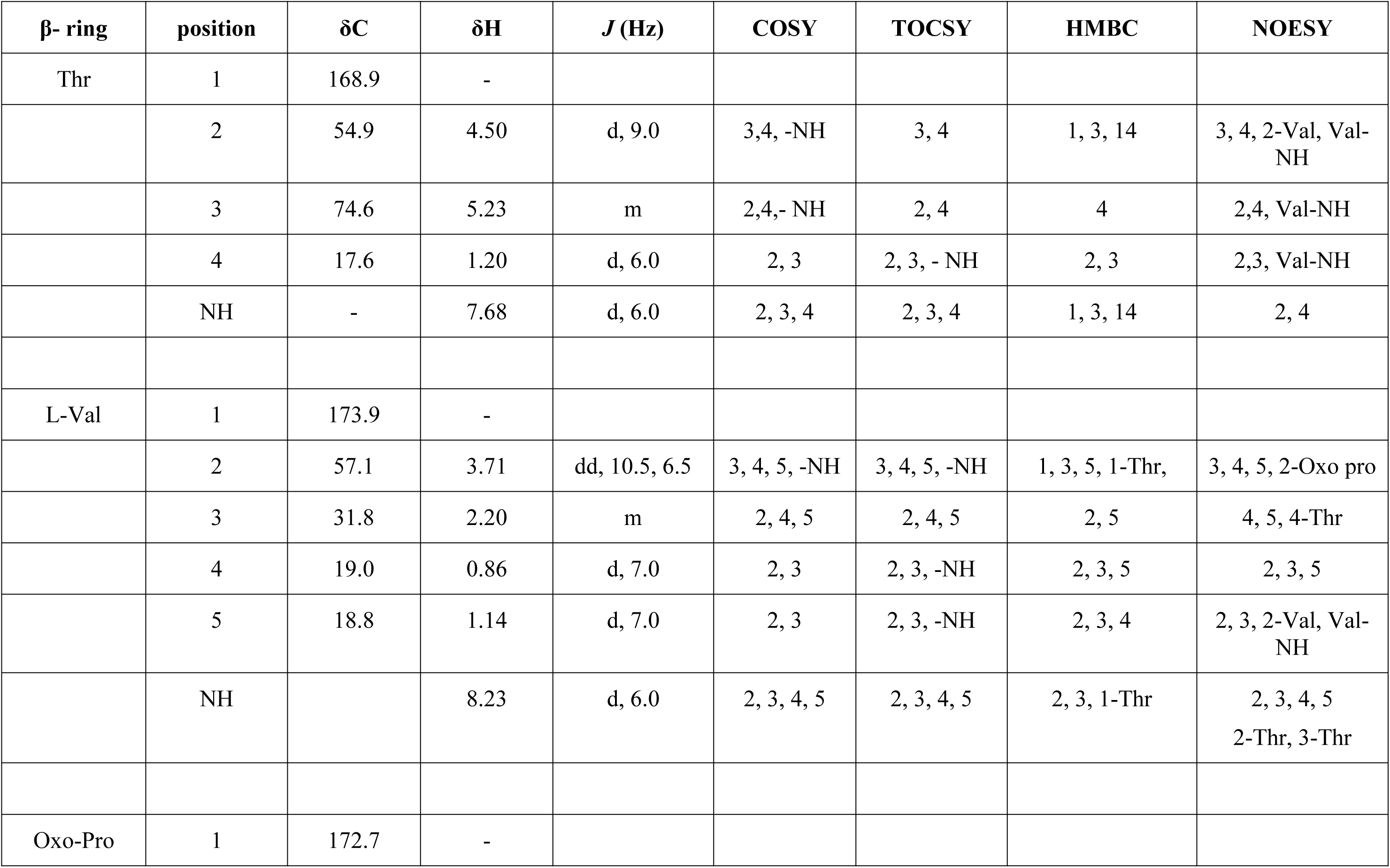
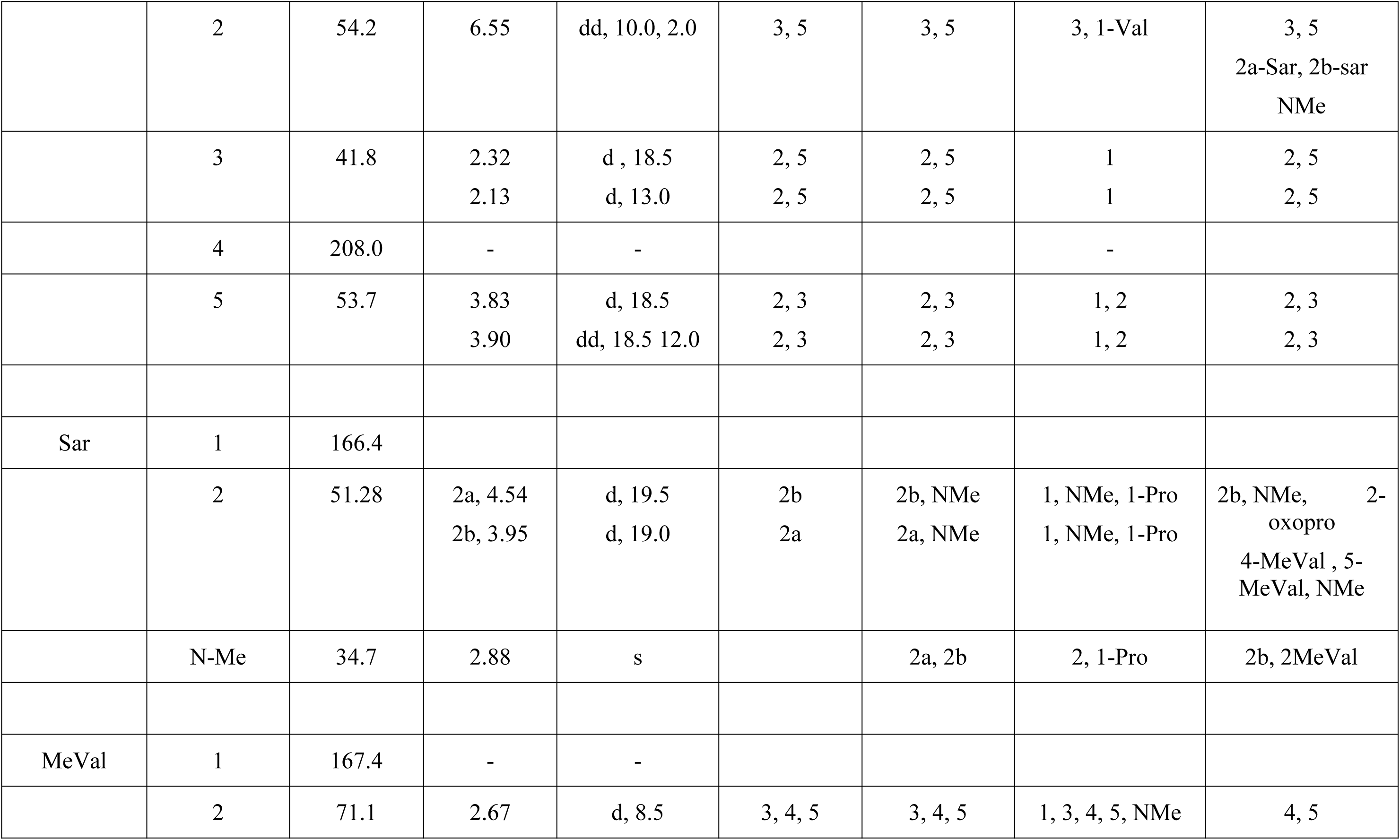
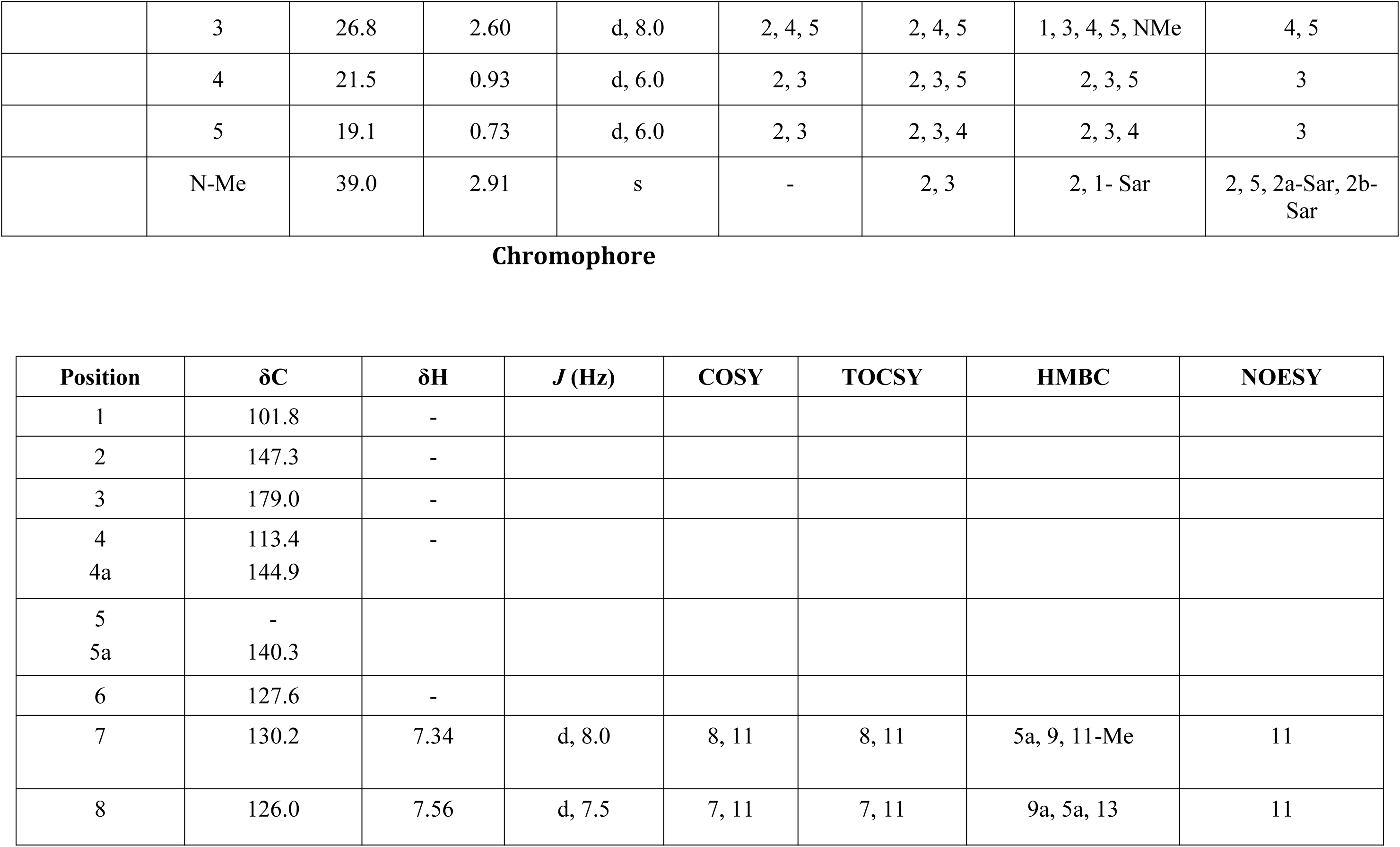
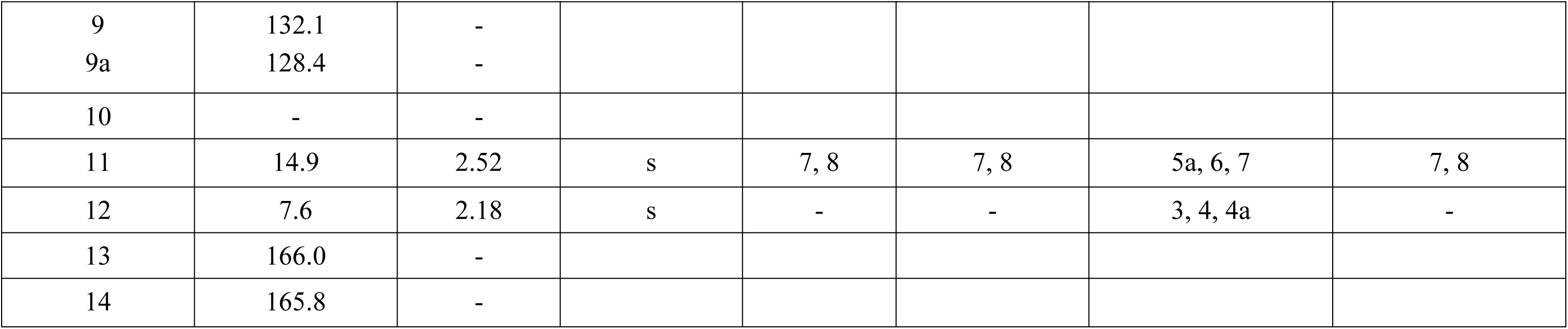
NMR data of Trasitmycin (R1) in CDCl_3_ (^1^H: 500 MHz ^13^C: 125 MHz)

**Figure S53.**
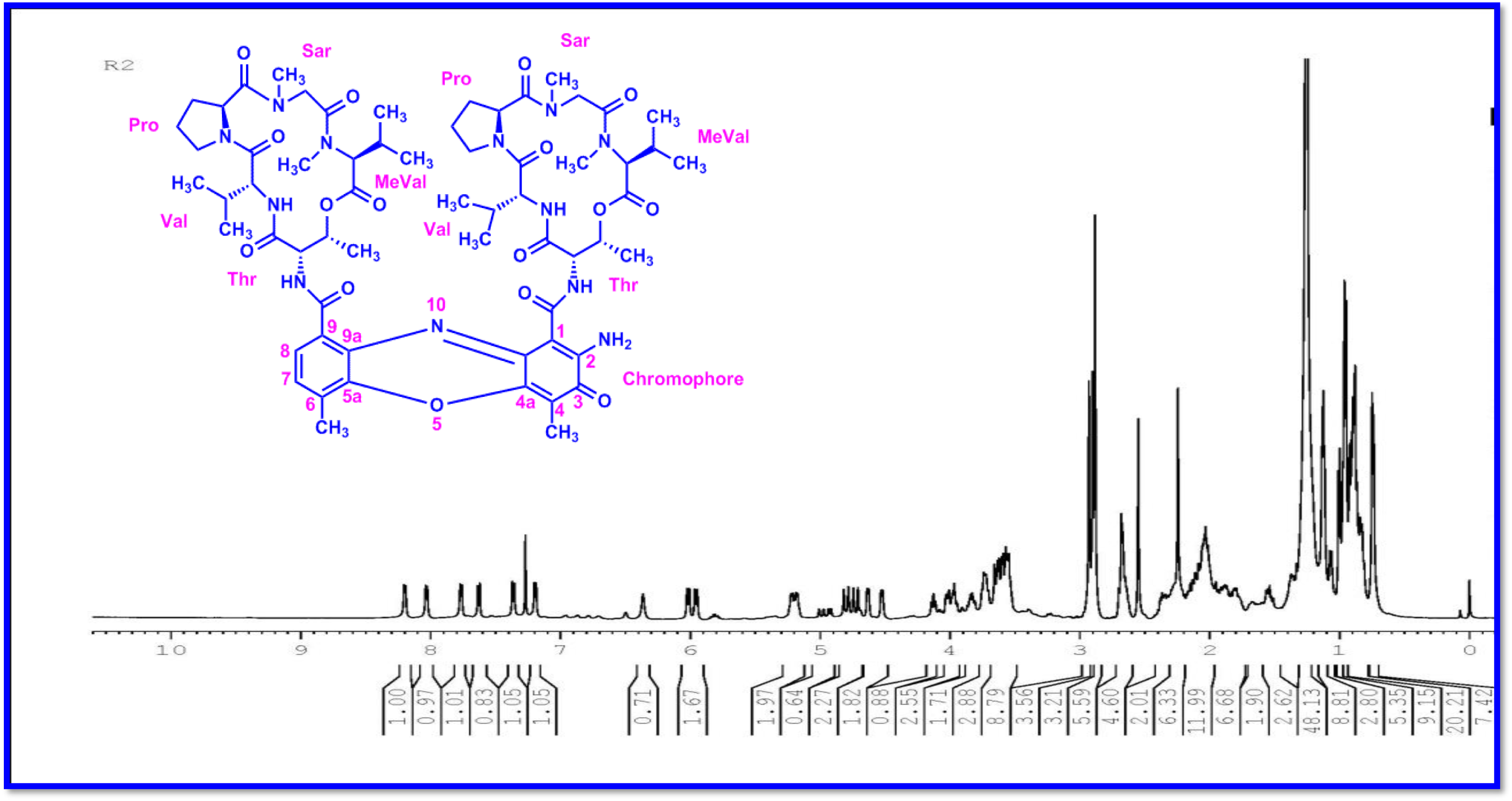
^1H^-NMR (500 MHz, CDCl_3_) Spectrum Of Compound R2.

**Figure S54.**
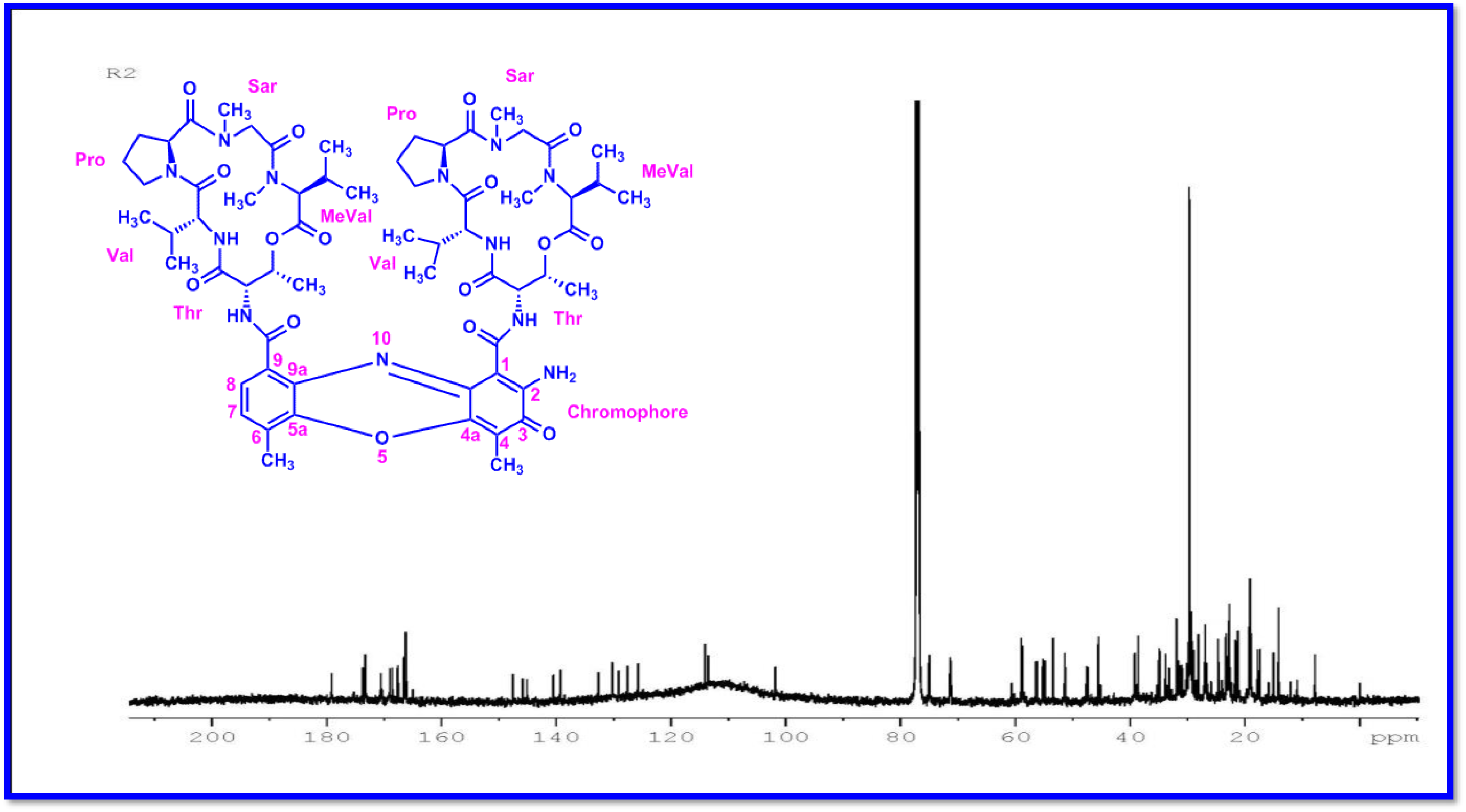
^13^ C NMR (125 MHz, CDCl_3_) Spectrum Of Compound R2.

**Figure S55.**
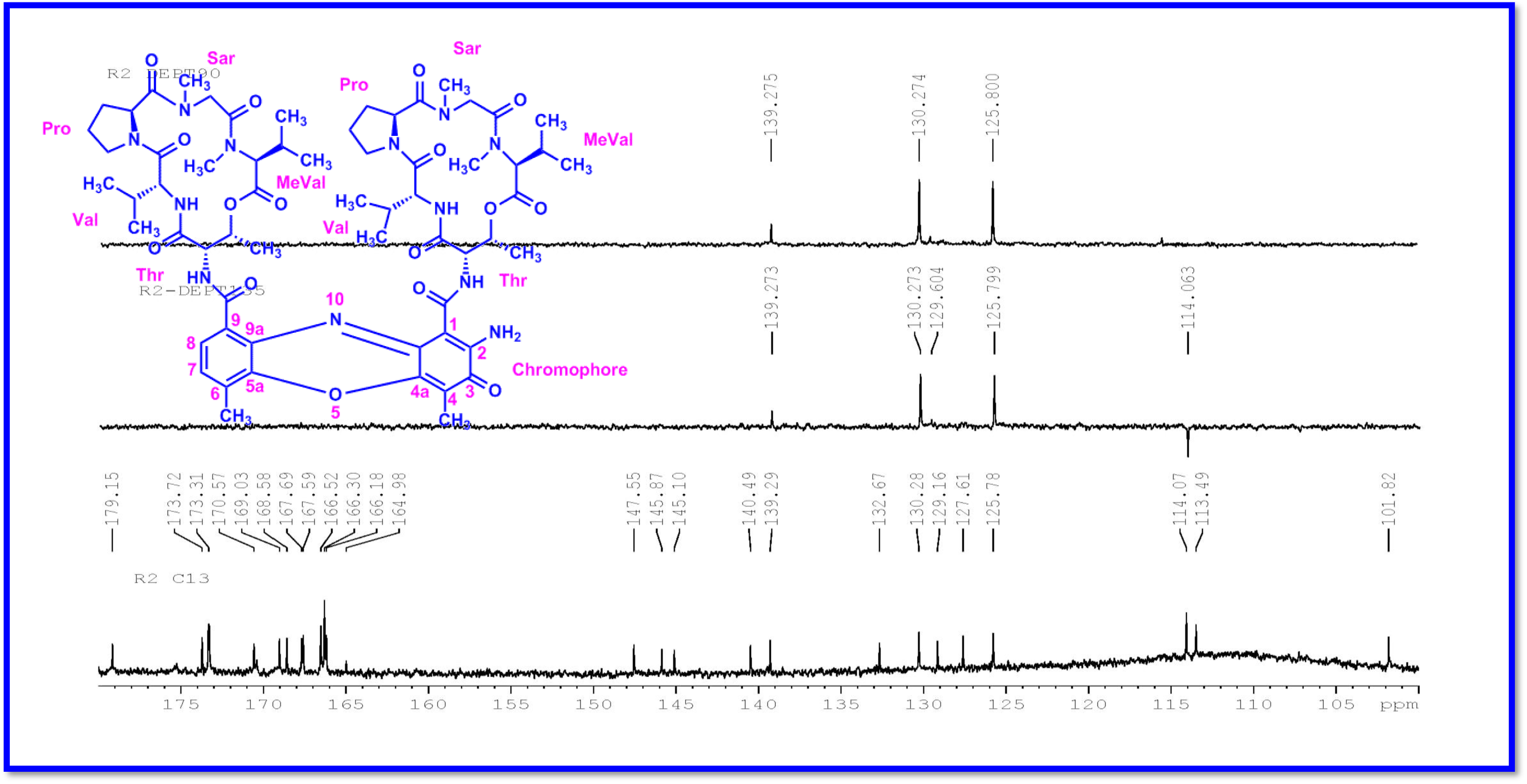
DEPT135&90 (125 MHz, CDCl_3_) Spectrum of Compound R2.

**Figure S56.**
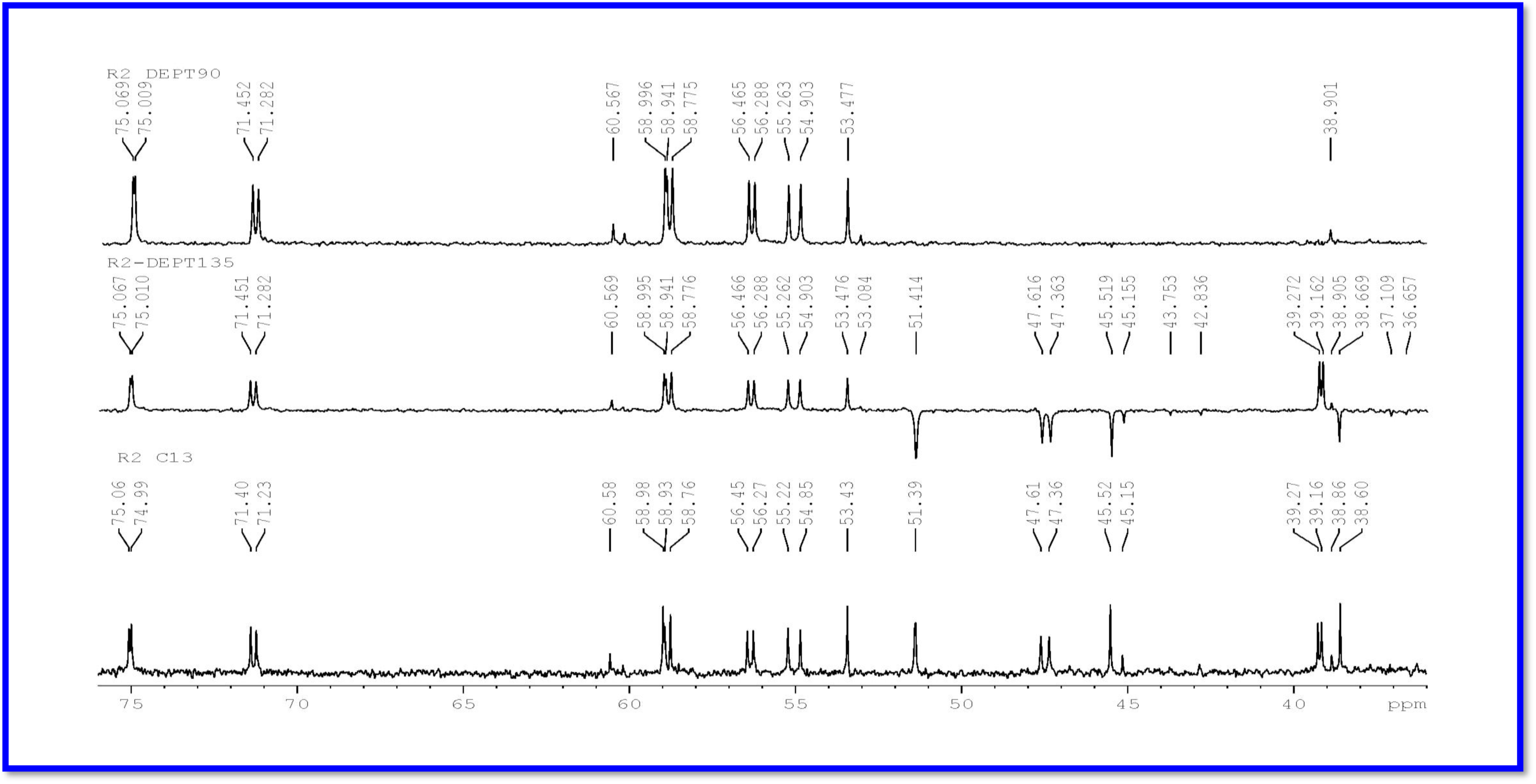
Expansion of DEPT135 &90 (125 MHz, CDCl_3_) Spectrum of Compound R2.

**Figure S57.**
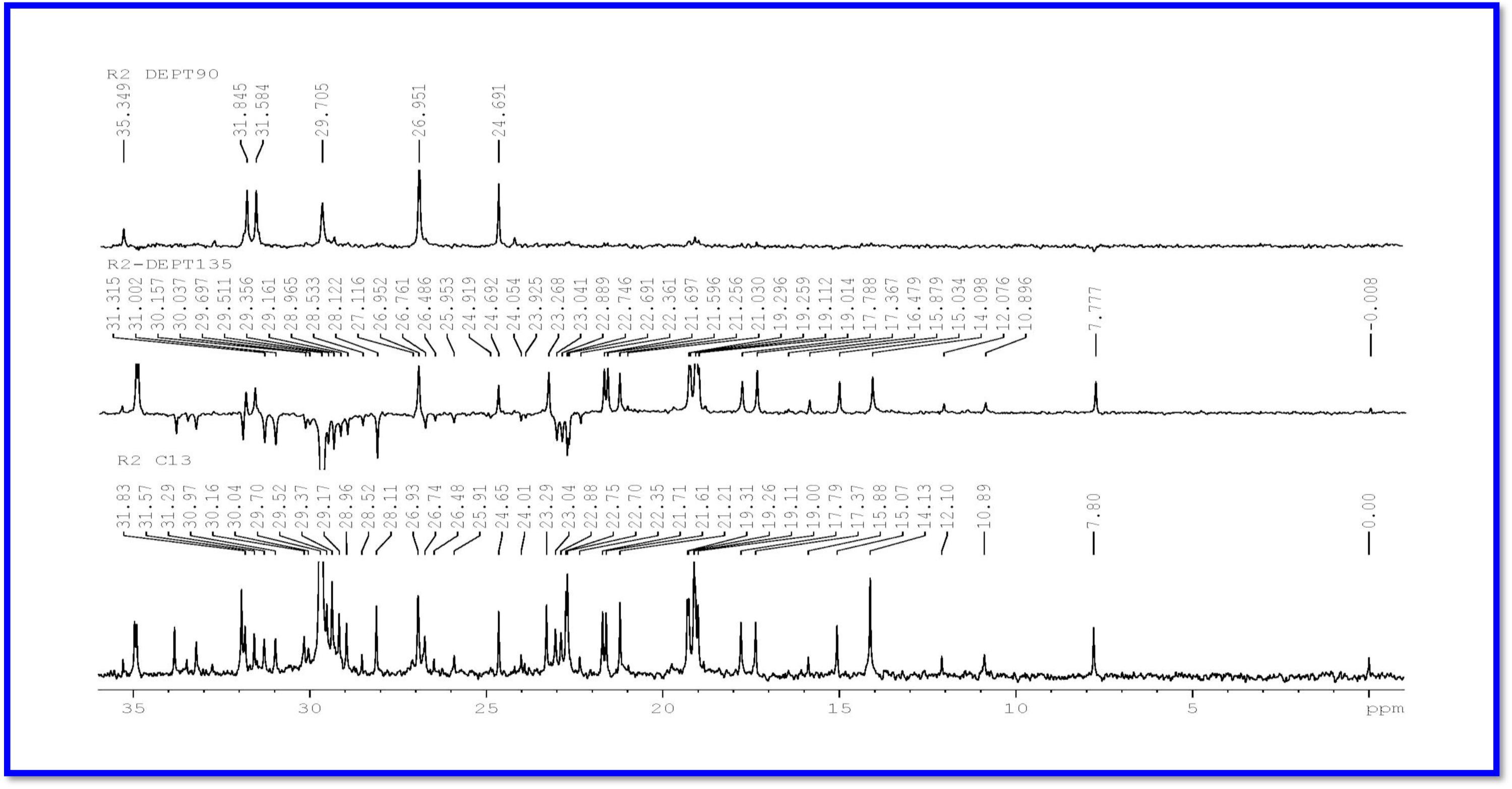
Expansion of DEPT135 &90 (125 MHz, CDCl_3_) Spectrum of Compound R2.

**Figure S58.**
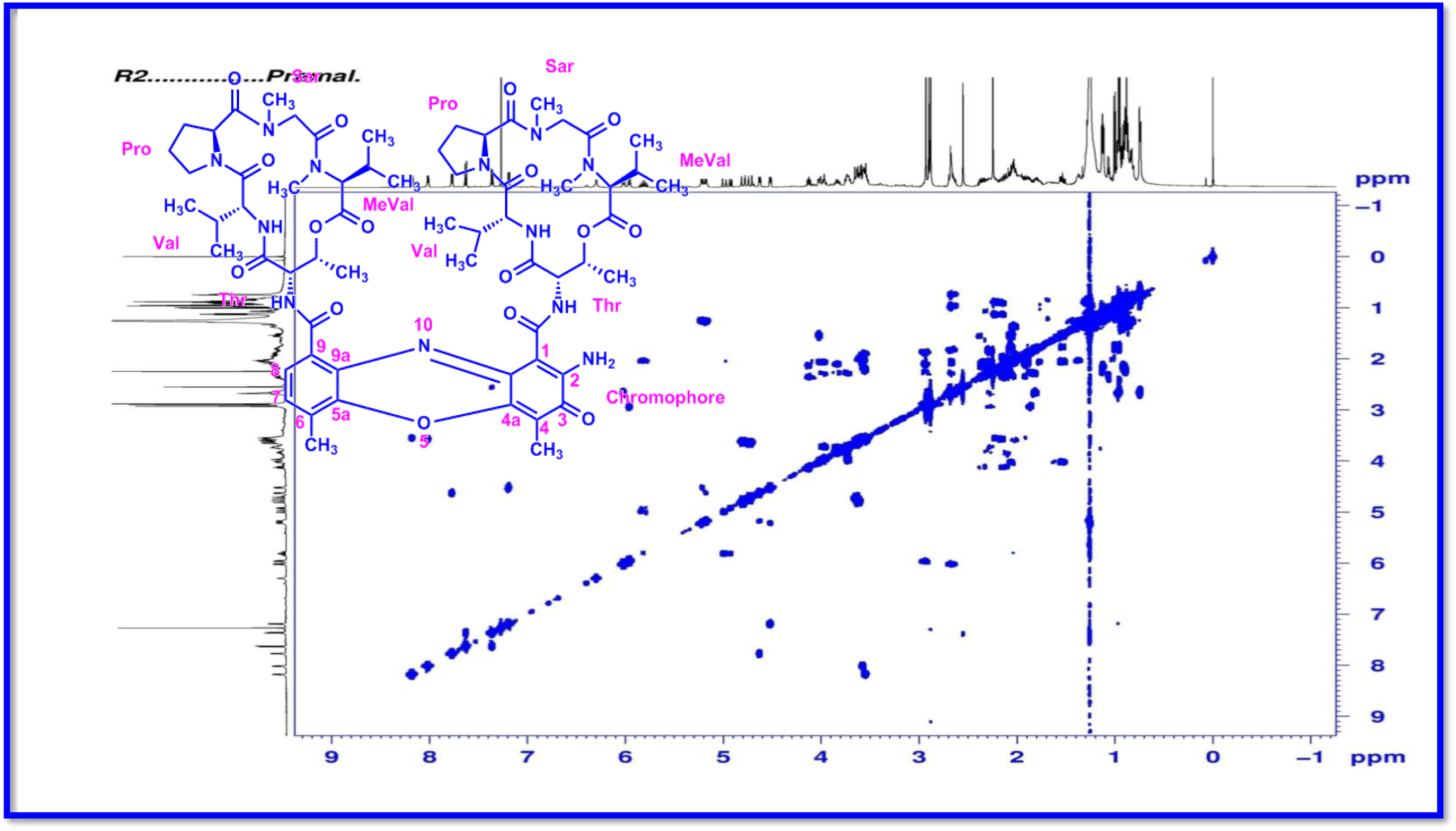
COSY (500 MHz) Spectrum of Compound R2.

**Figure S59.**
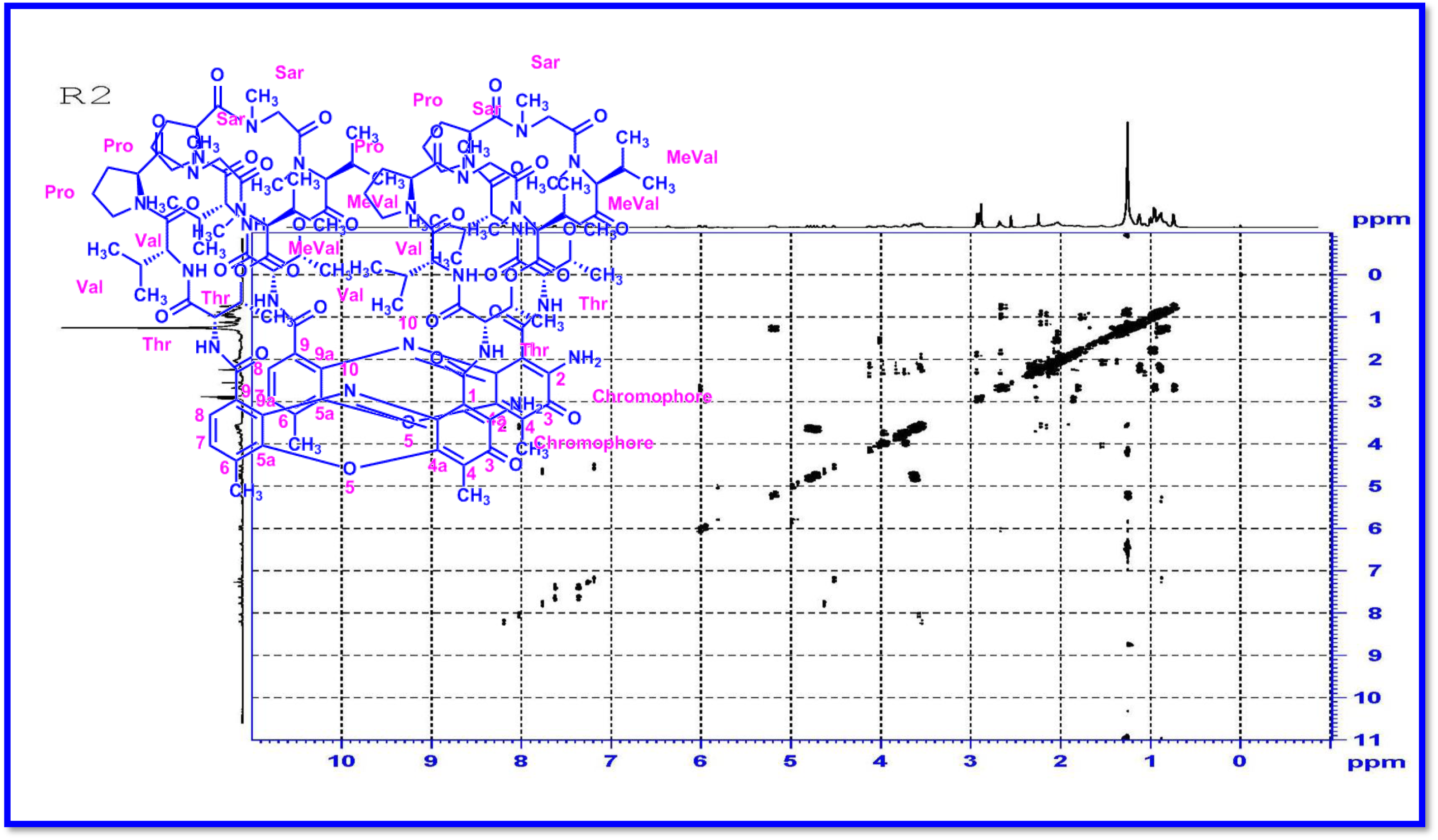
DQF-COSY (500 MHz) Spectrum of Compound R2.

**Figure S60.**
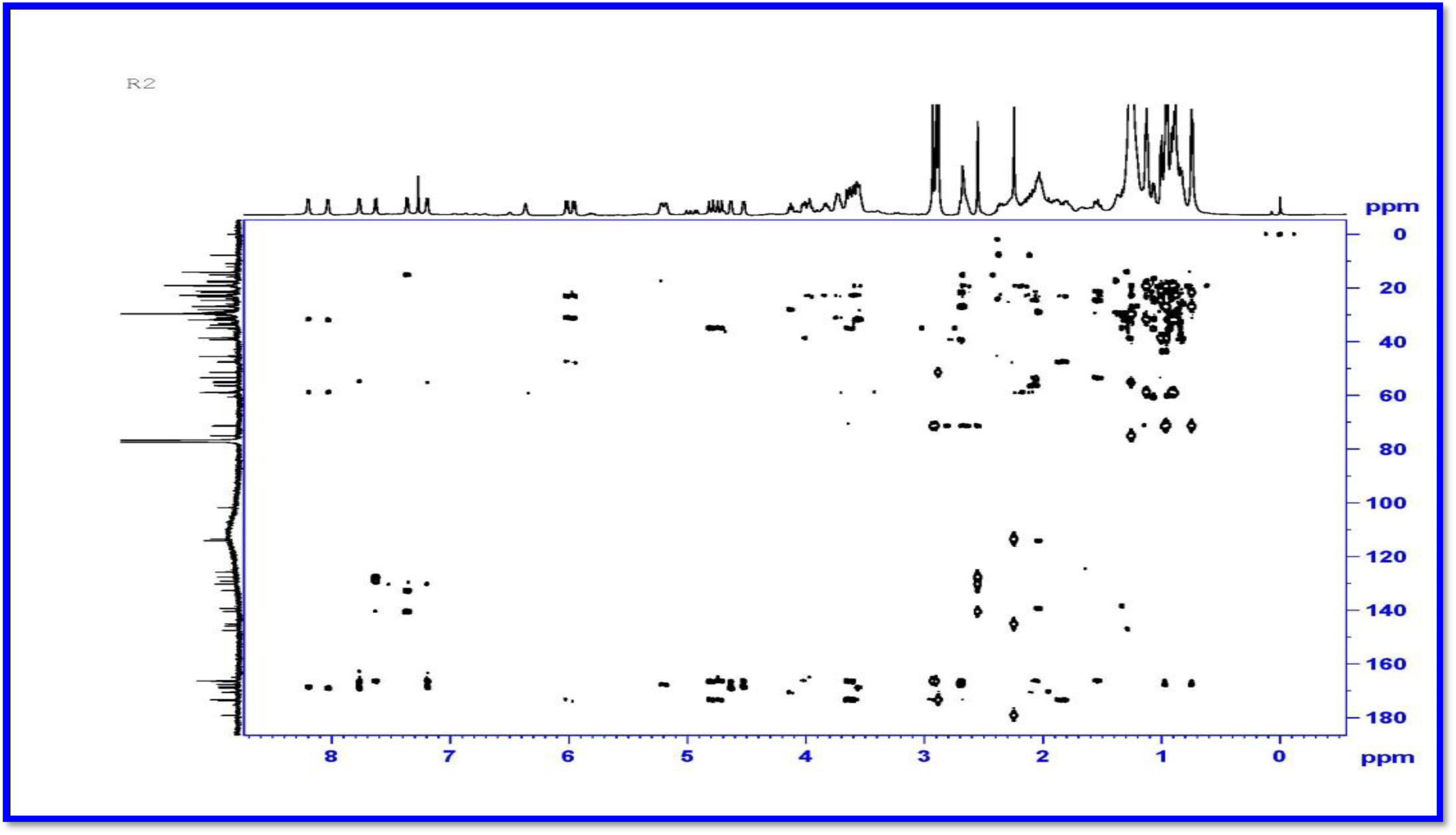
HMBC (500 MHz) Spectrum Compound of R2.

**Figure S61.**
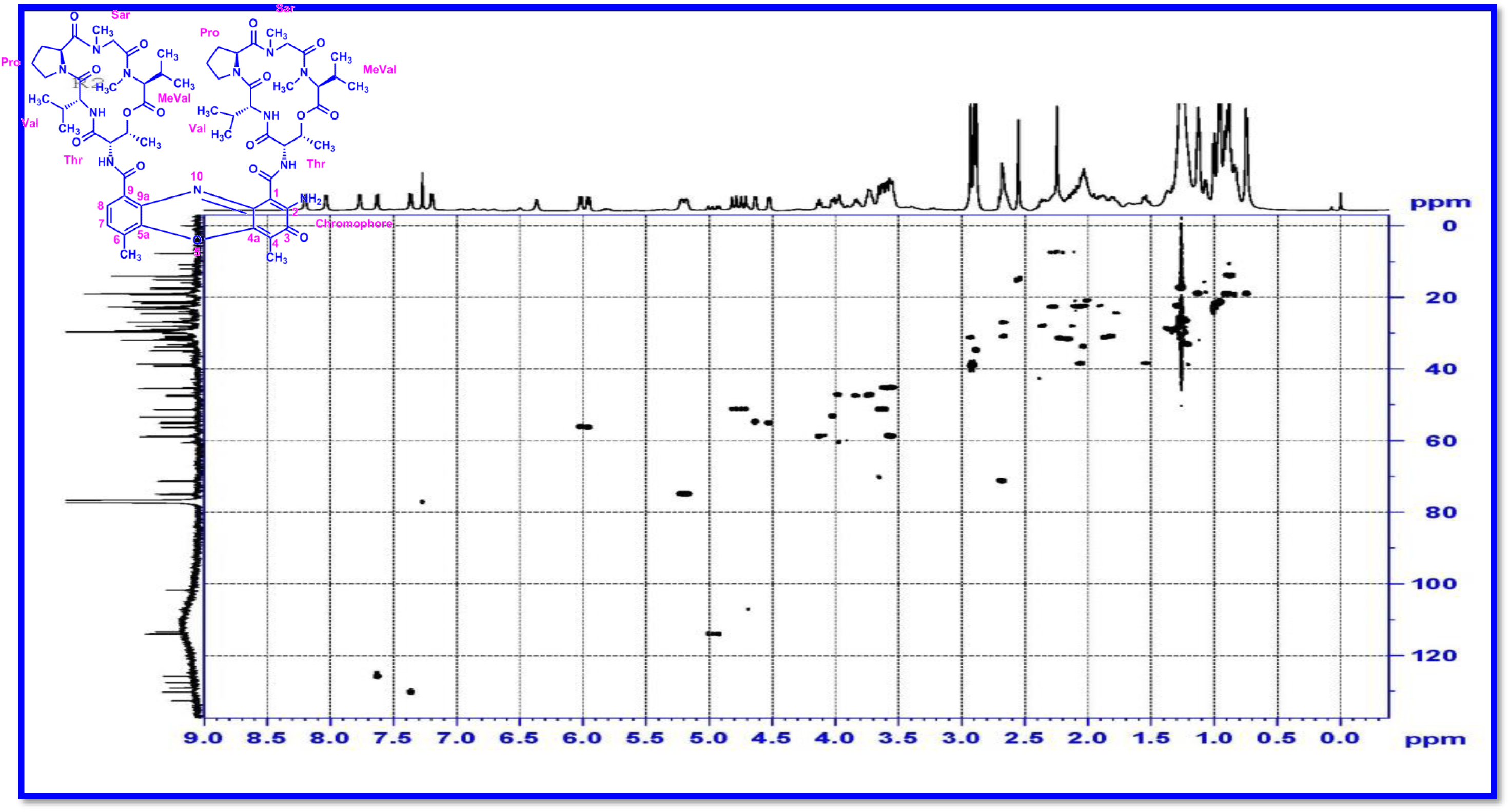
HSQC (500 MHz) Spectrum of Compound R2.

**Figure S62.**
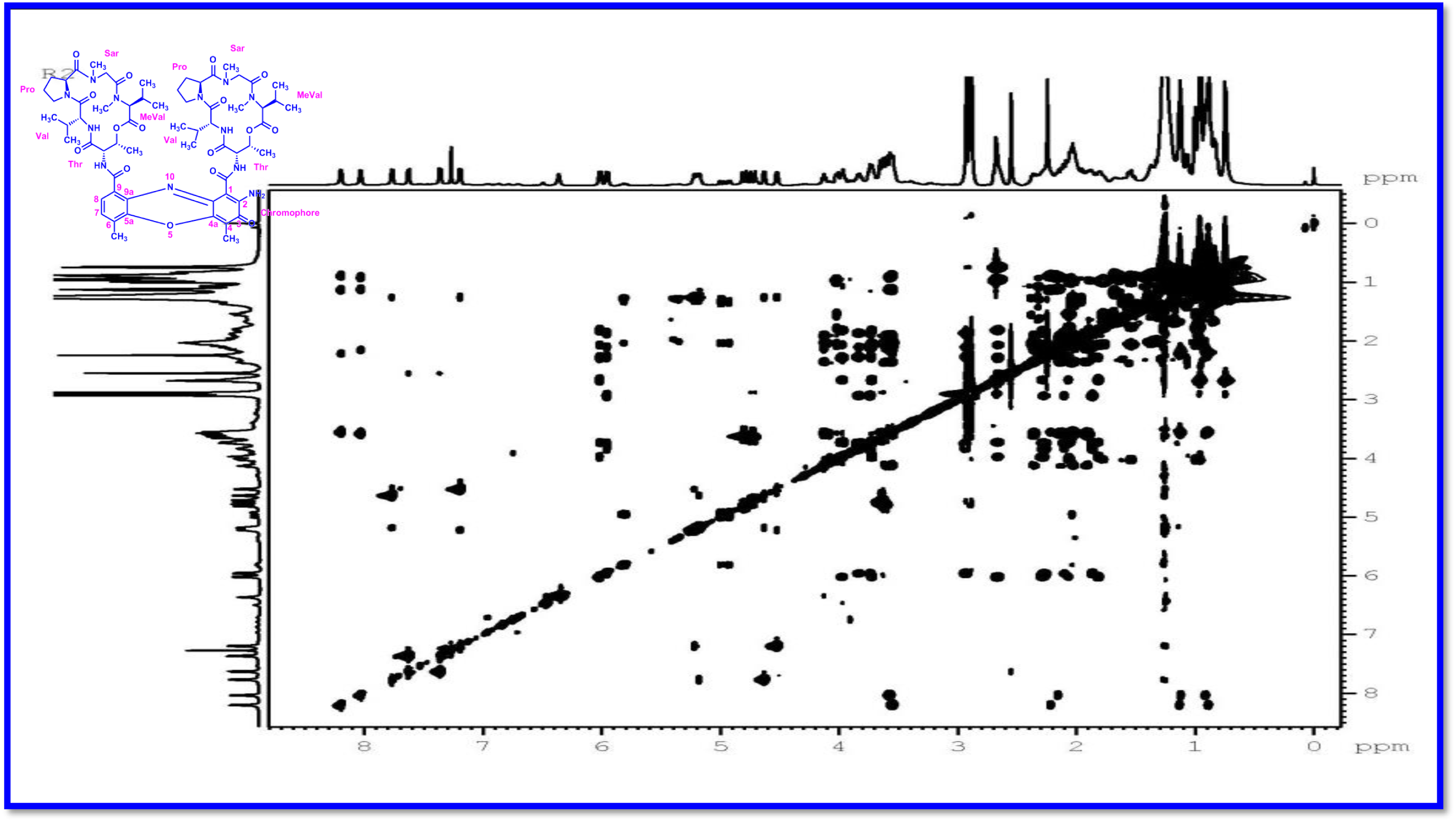
TOCSY (500 MHz) Spectrum of Compound R2.

**Figure S63.**
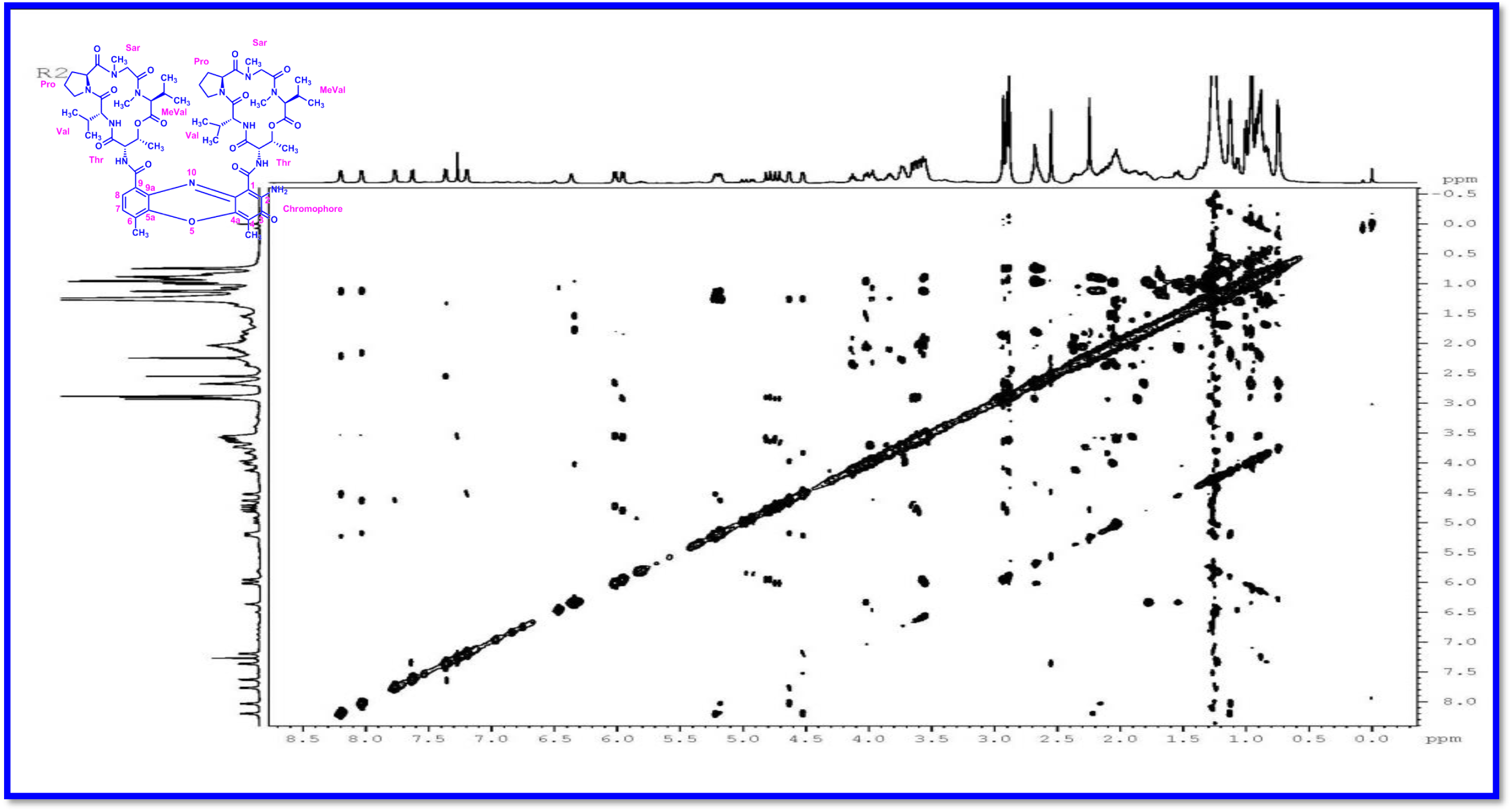
NOESY (500 MHz) Spectrum of Compound R2.

**Figure S64.**
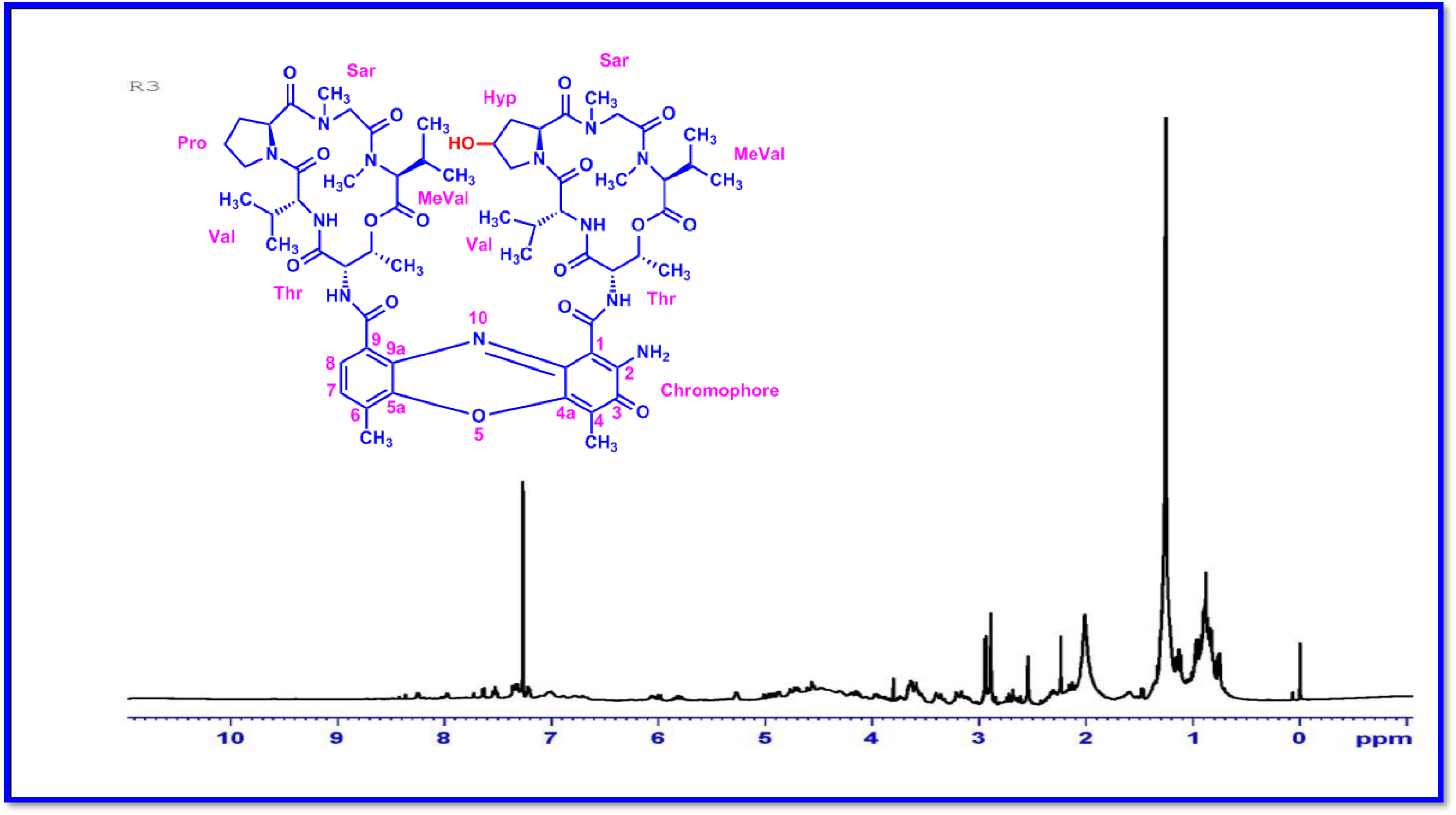
^1^H-NMR (500 MHz, CDCl_3_) Spectrum of Compound R3.

**Figure S65.**
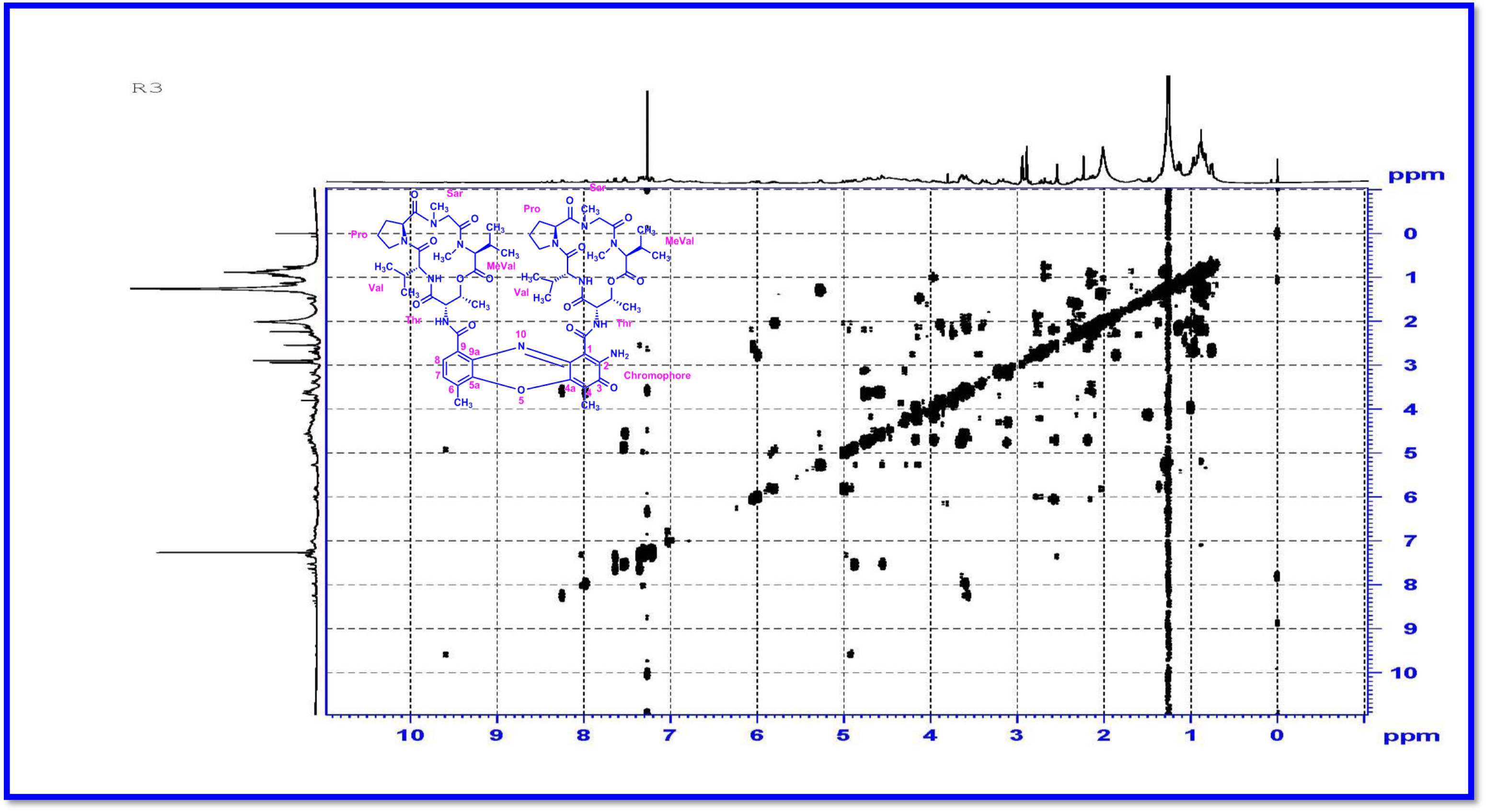
DQF-COSY (500 MHz, CDCl_3_) Spectrum of Compound R3.

**Figure S66.**
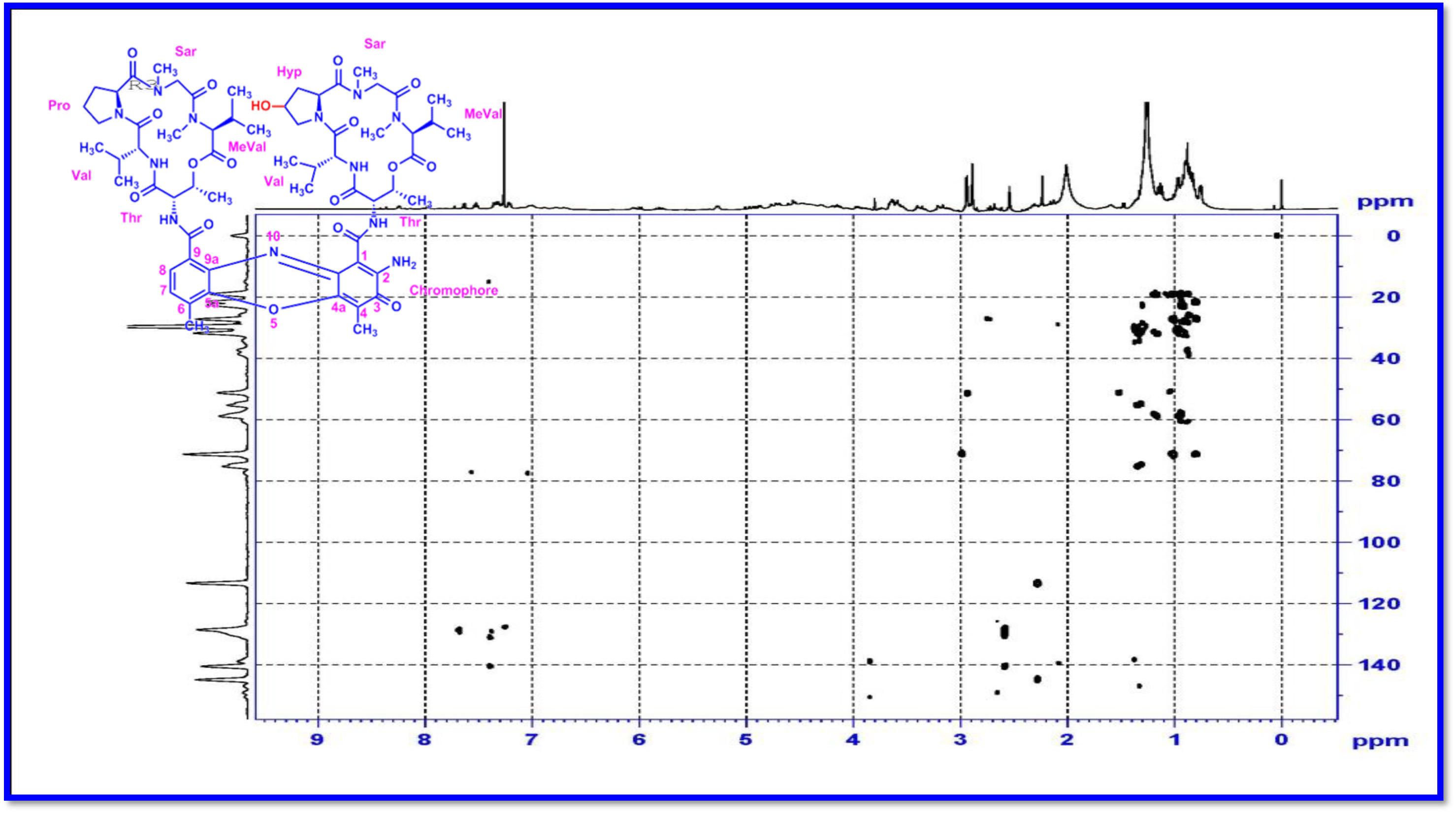
HMBC (500 MHz) Spectrum of Compound R3.

**Figure S67.**
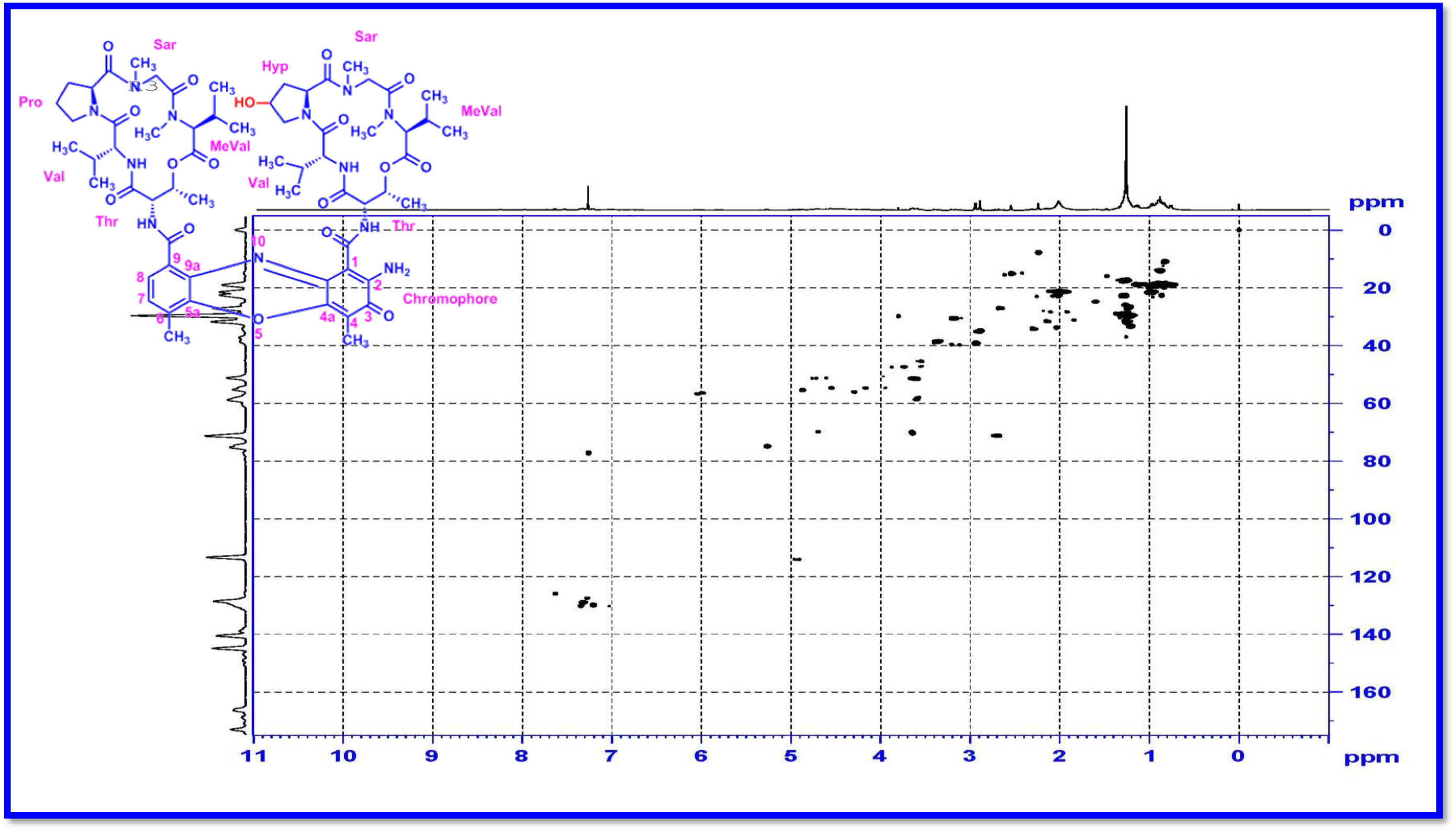
HSQC (500 MHz) Spectrum of Compound R3.

**Figure S68.**
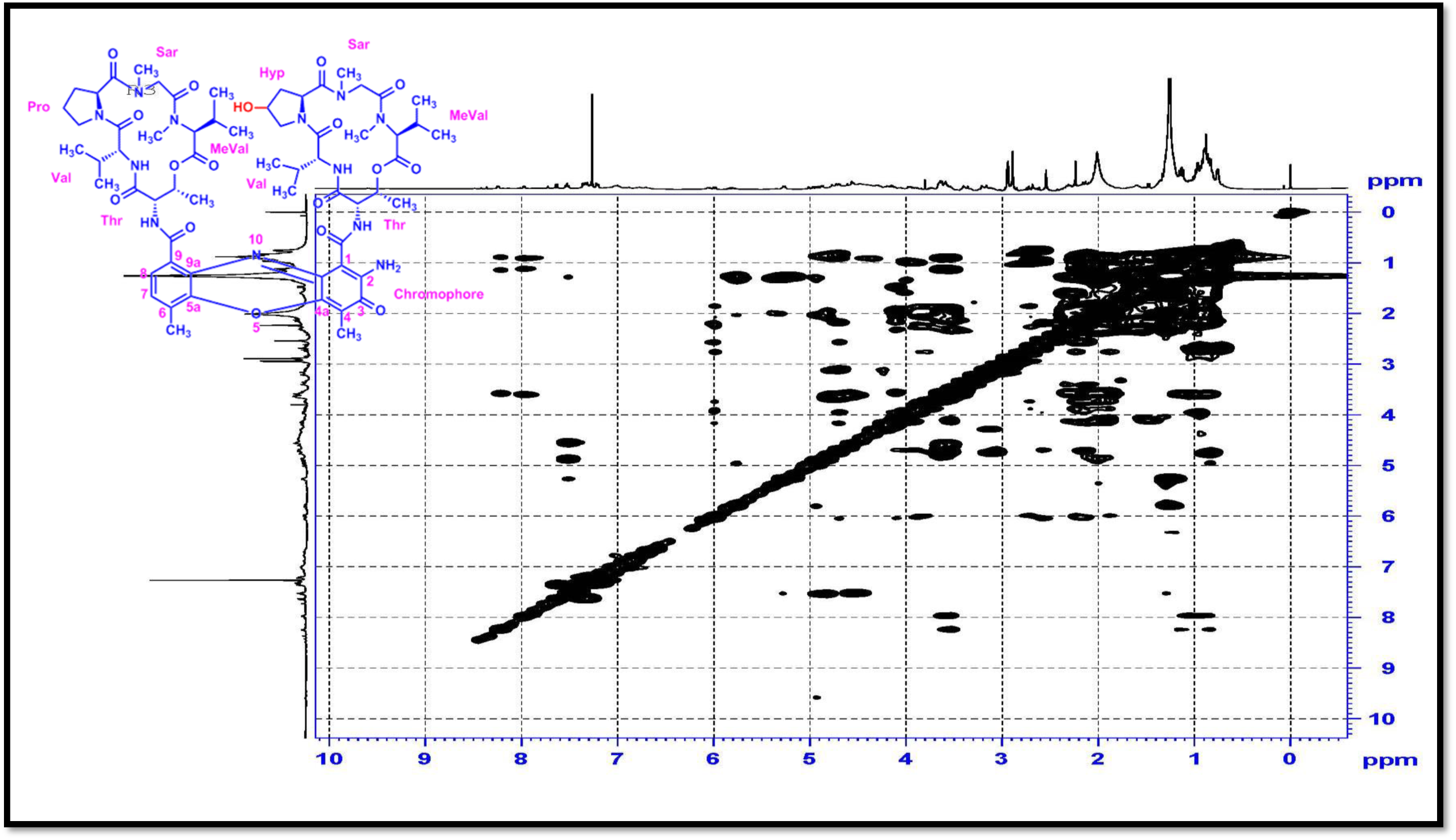
TOCSY (500 MHz, CDCl_3_) Spectrum of Compound R3.

**Figure S69.**
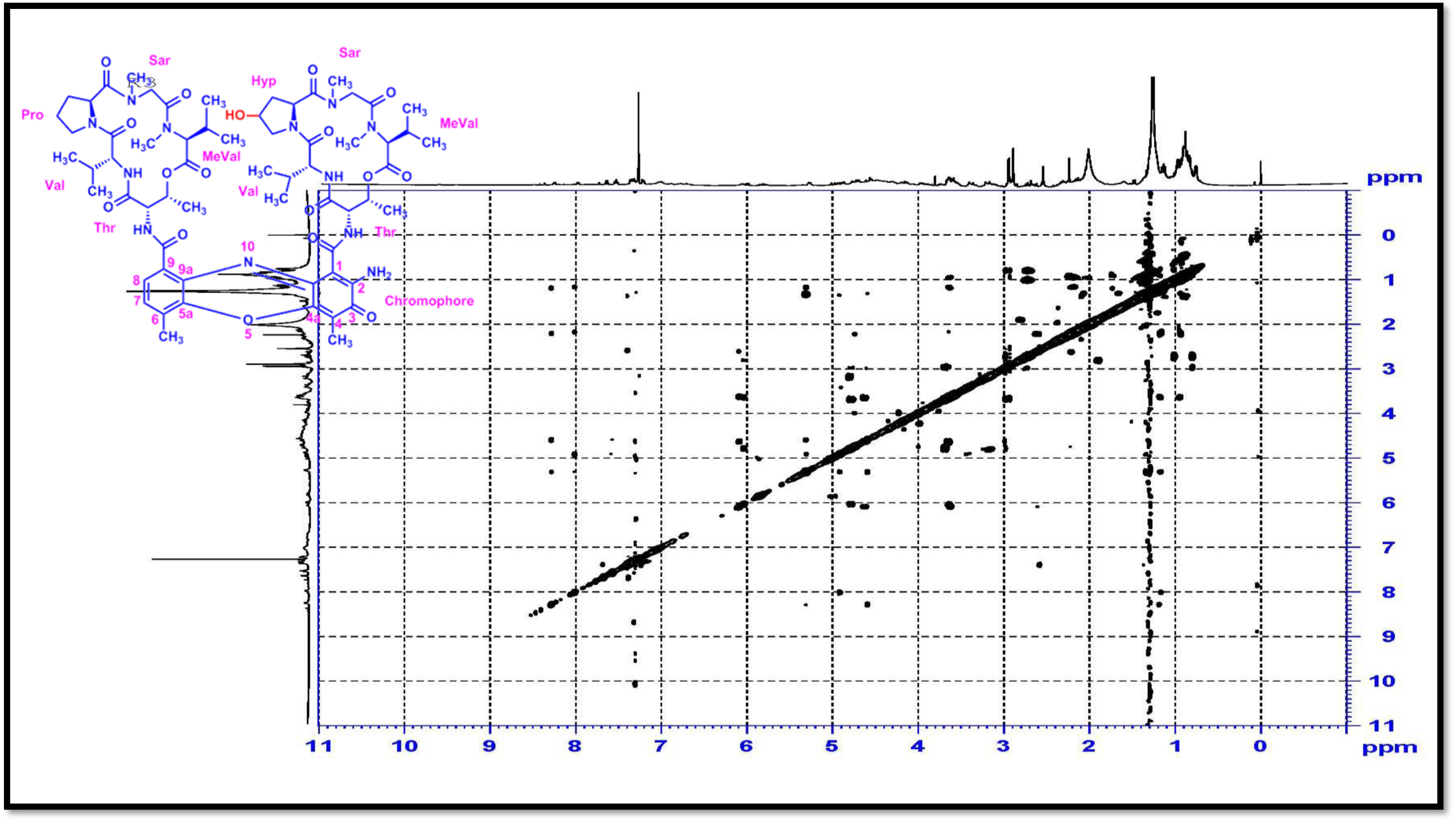
NOESY (500 MHz) Spectrum of Compound R3.

**Figure S70.**
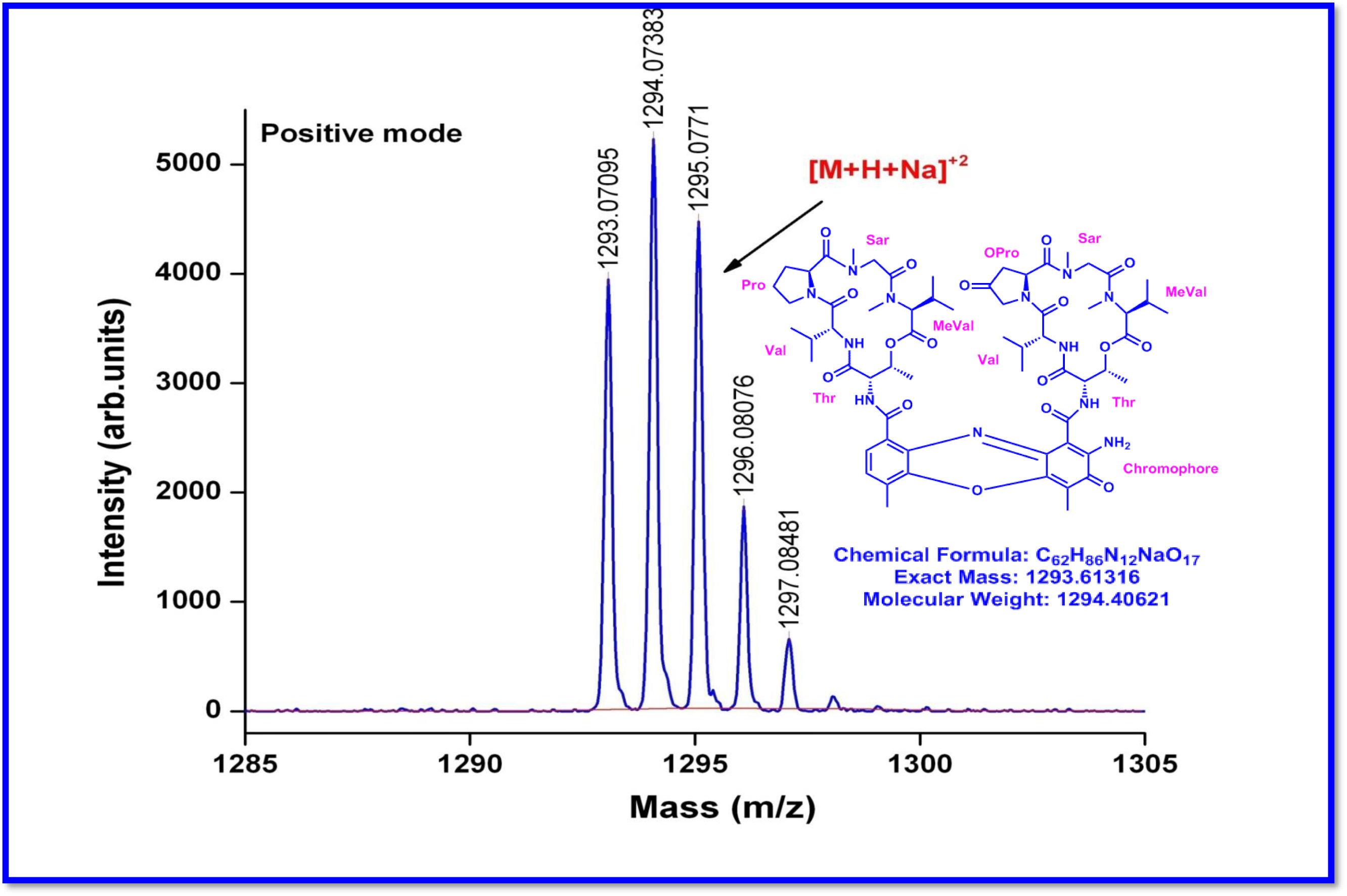
MALDI-TOF MS Spectrum of Transitmycin (R1) (Molecular ion peak) (positive mode)

**Figure S71.**
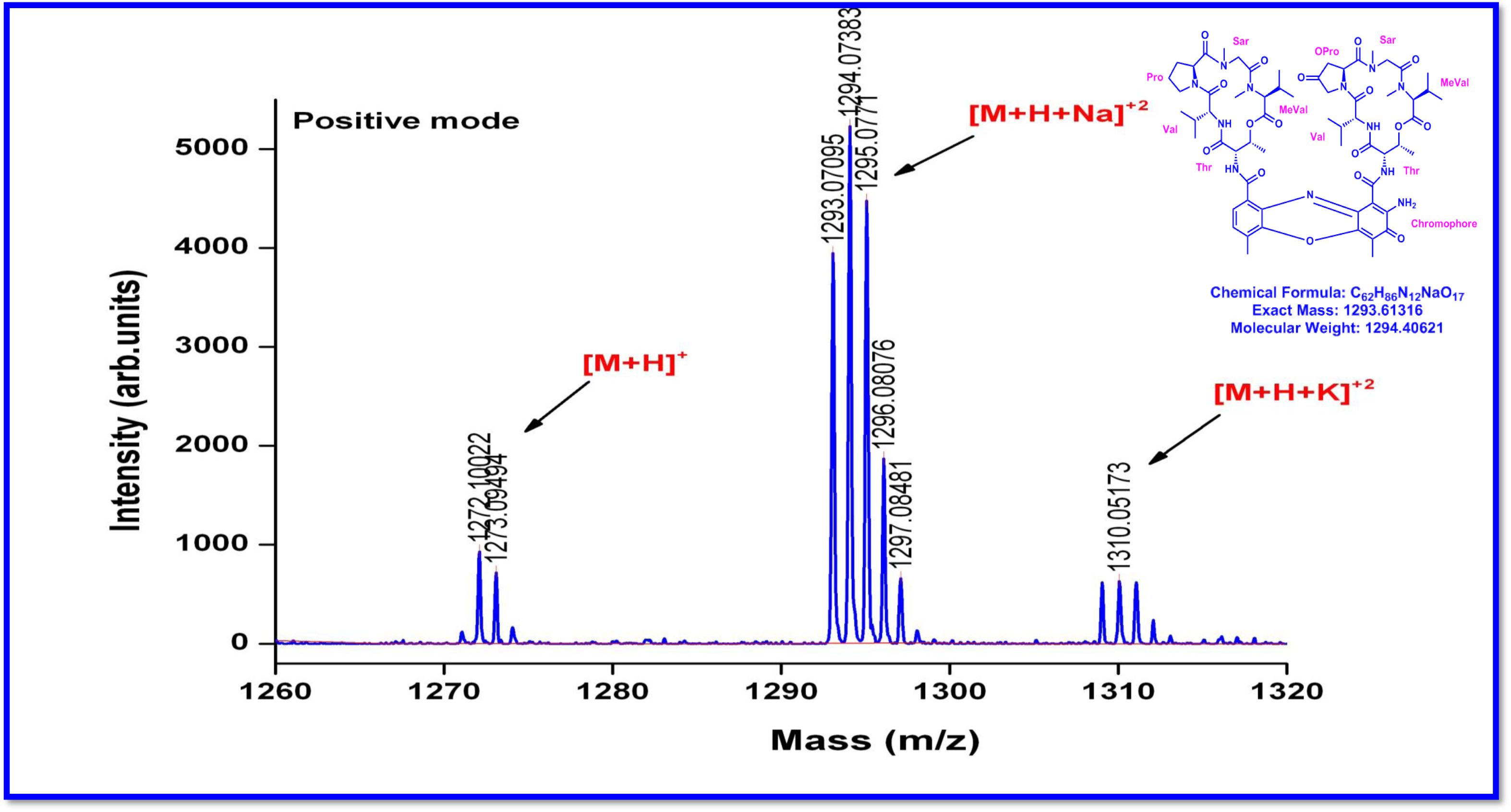
Expansion of MALDI-TOF MS Spectrum of Transitmycin (R1) (Molecular ion peak) (positive mode)

**Figure S72.**
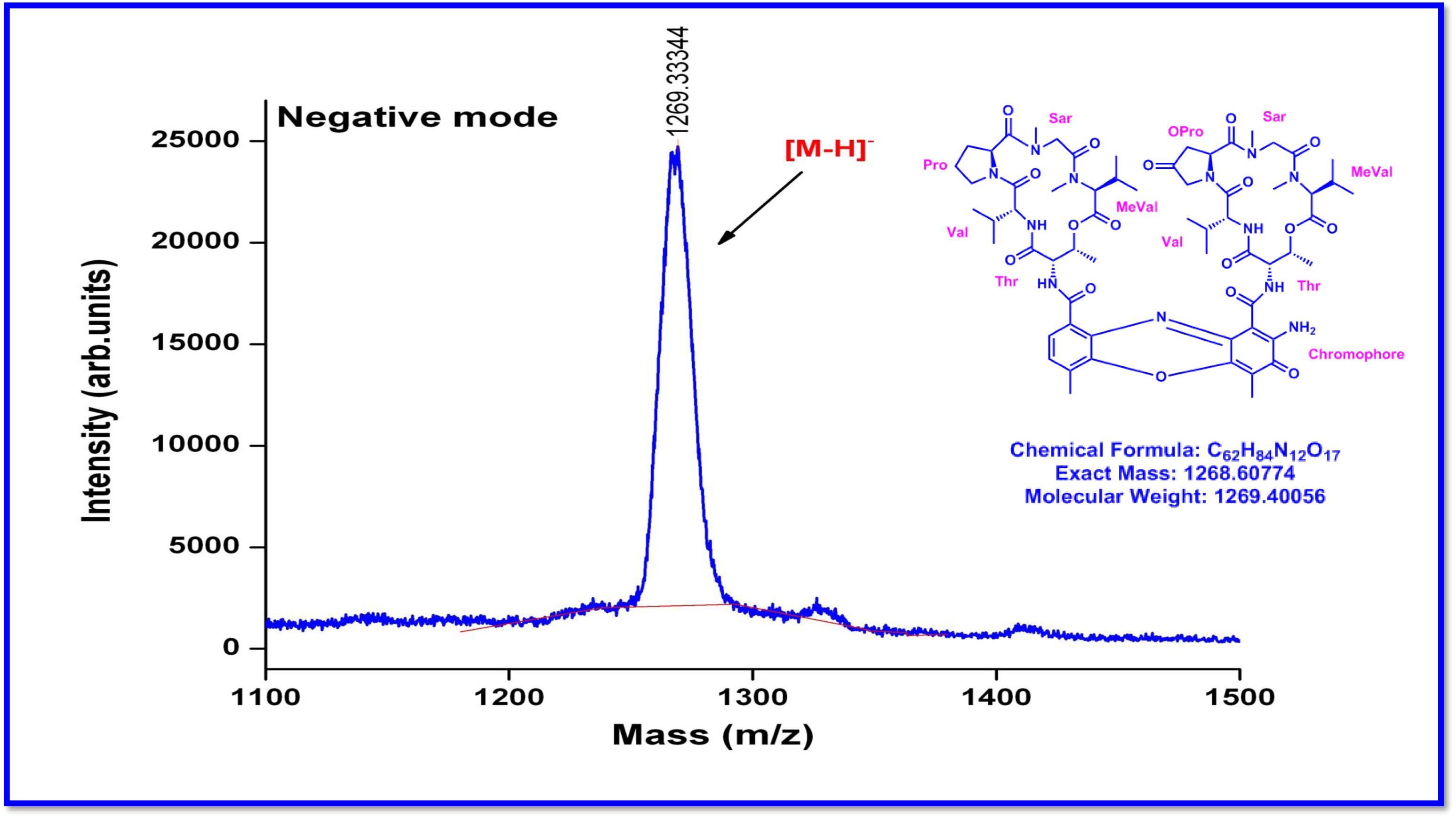
Expansion of MALDI-TOF MS Spectrum of Transitmycin (R1) (Molecular ion peak) (Negative mode)

**Figure S73.**
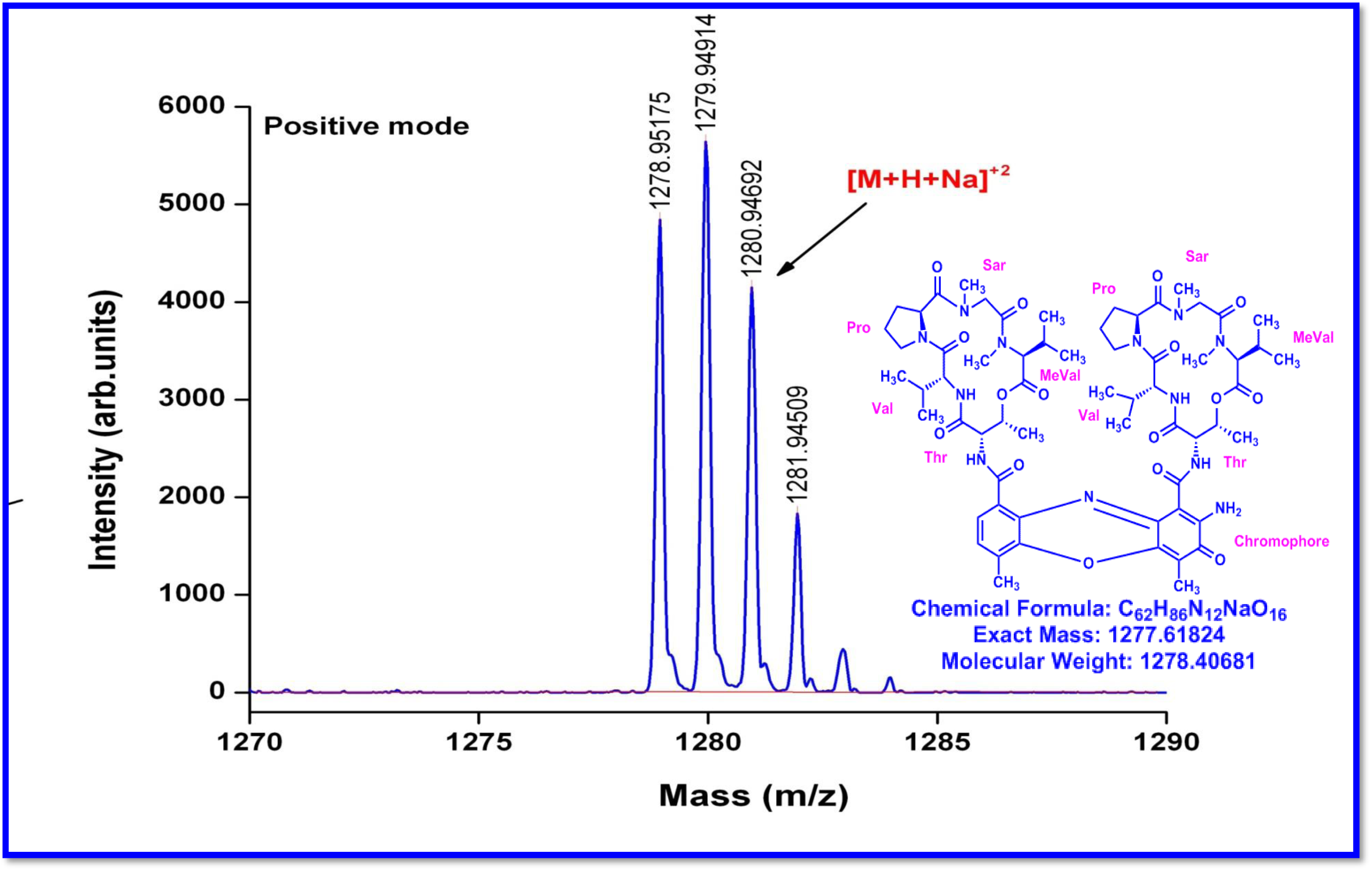
MALDI-TOF MS Spectrum of R2 (Molecular ion peak) (positive mode)

**Figure S74.**
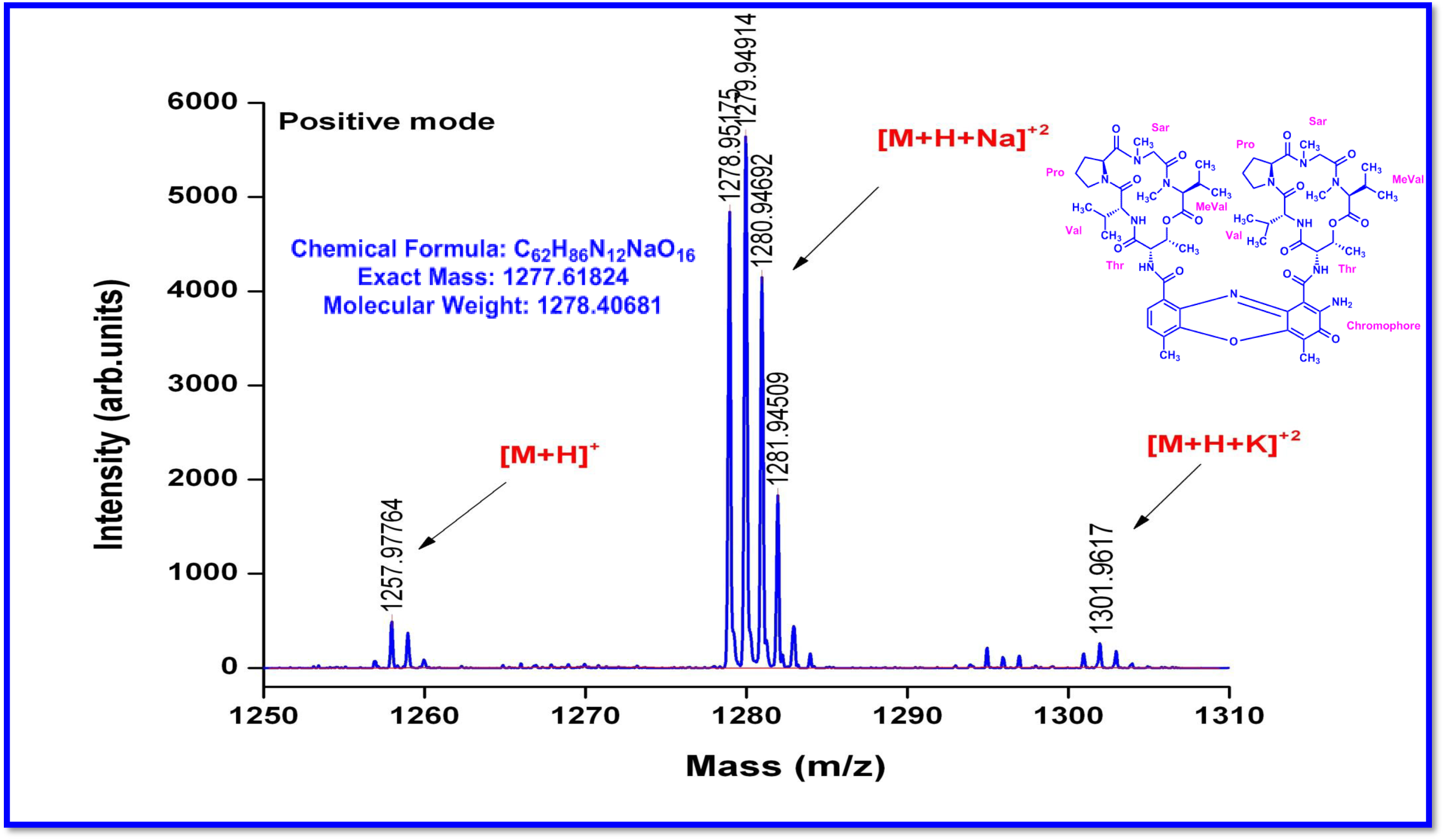
Expansion of MALDI-TOF MS Spectrum of R2 (Molecular ion peak) (positive mode)

**Figure S75.**
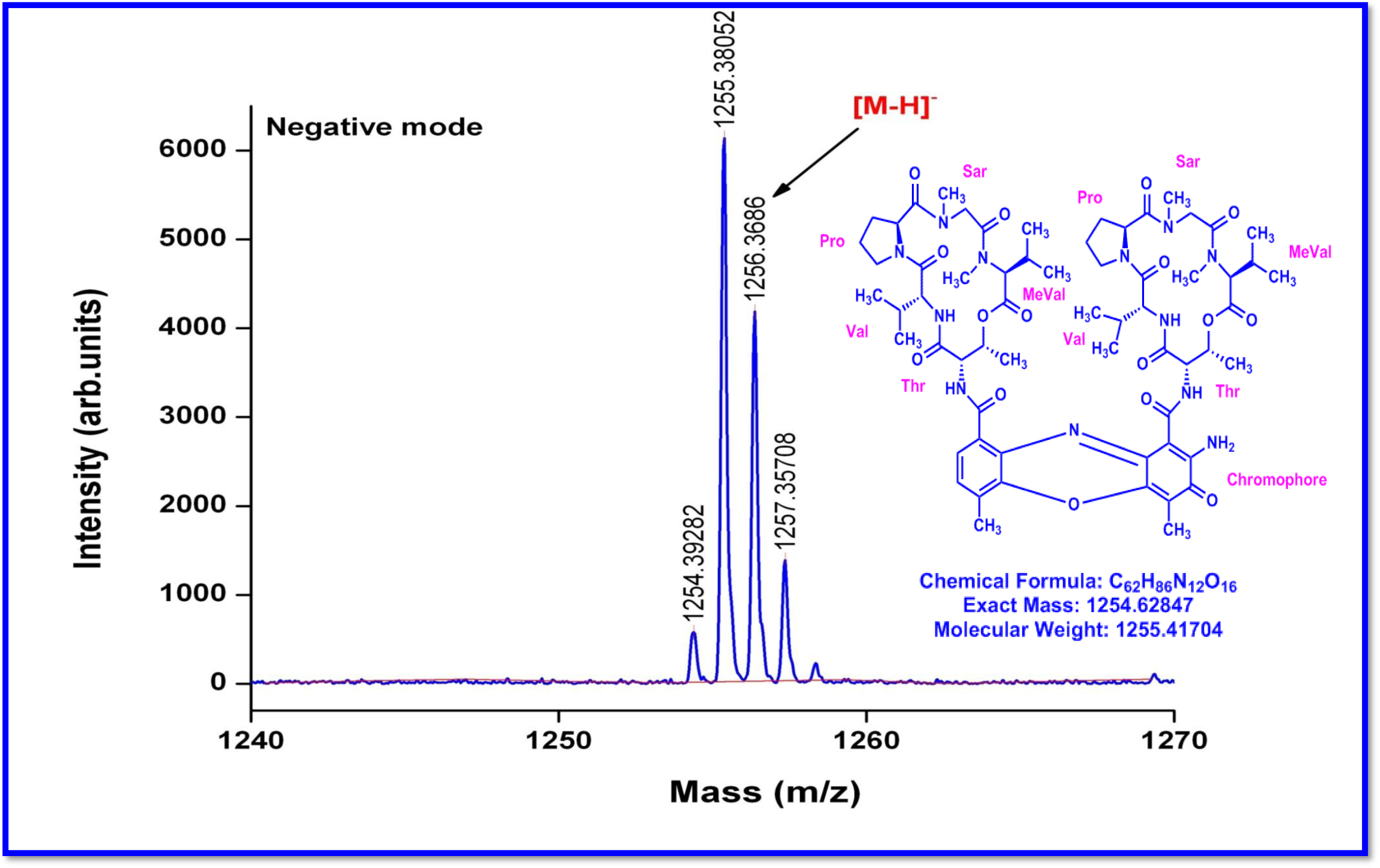
Expansion of MALDI-TOF MS Spectrum of R2 (Molecular ion peak) (Negative mode)

**Figure S76.**
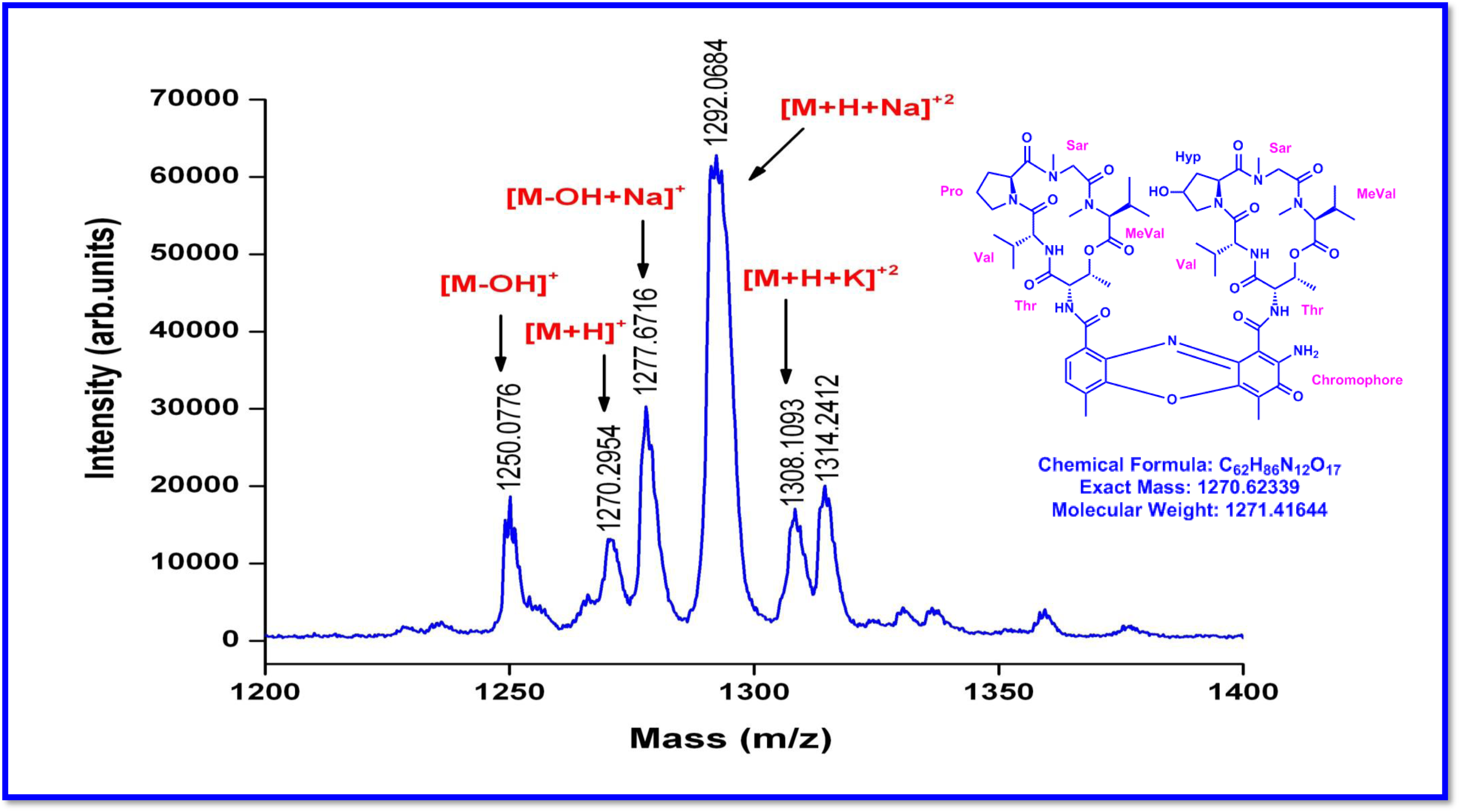
MALDI-TOF MS Spectrum of R3 (Molecular ion peak) (positive mode)

**Figure S77.**
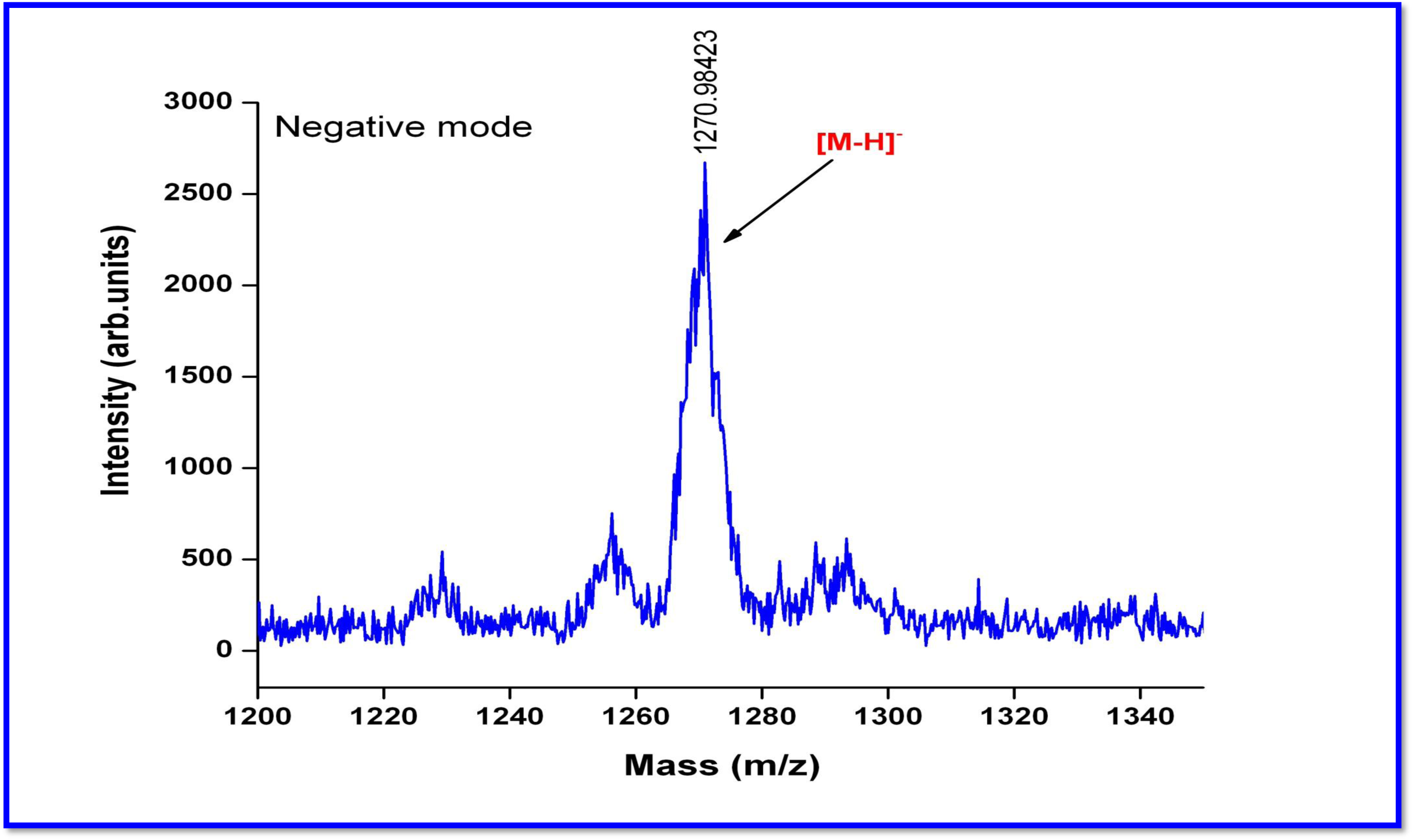
MALDI-TOF MS Spectrum of R3 (Molecular ion peak) (Negative mode)

**Figure S78.**
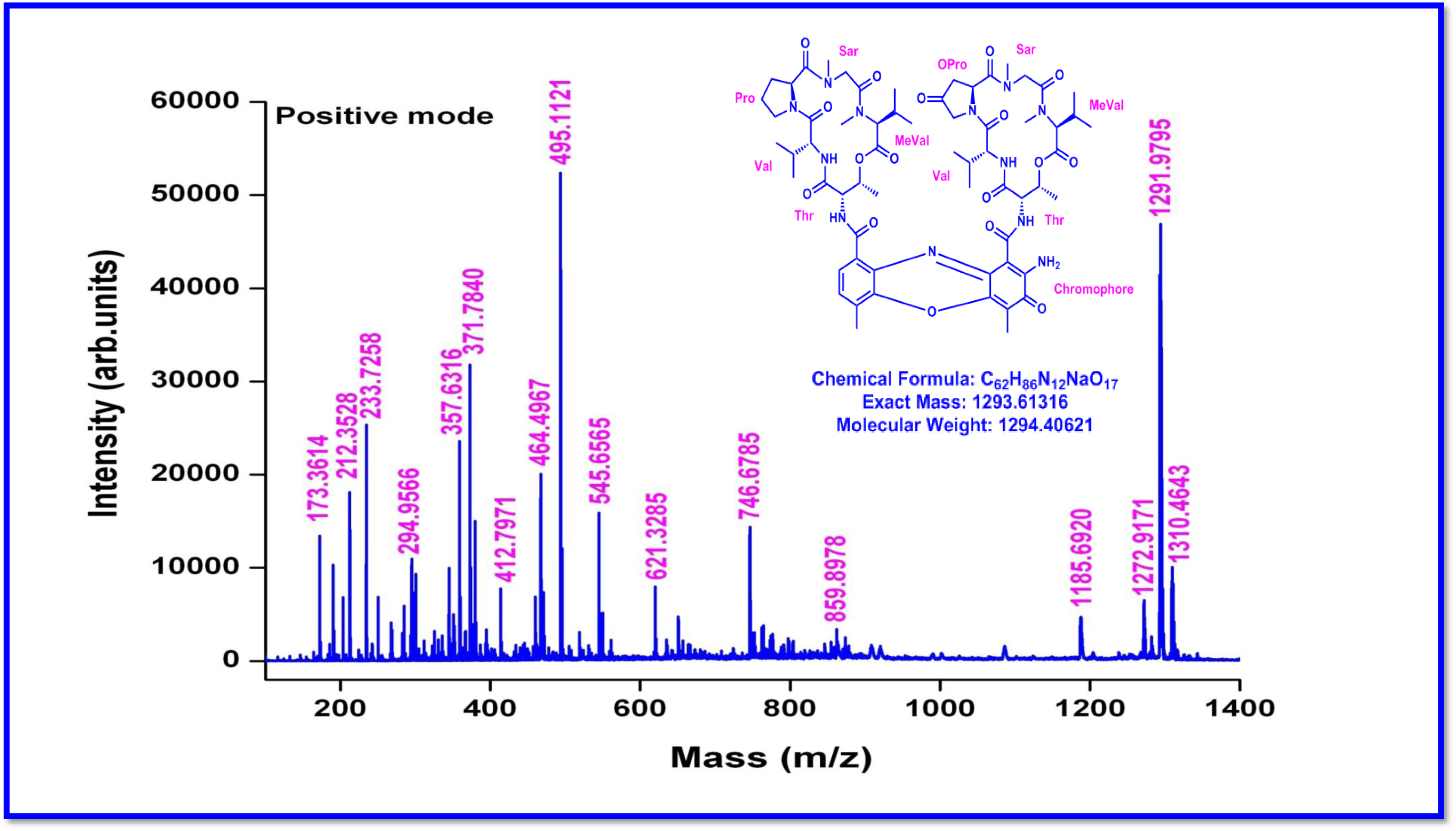
MALDI-TOF MS Spectrum of Transitmycin (R1) (Positive mode)

**Figure S79.**
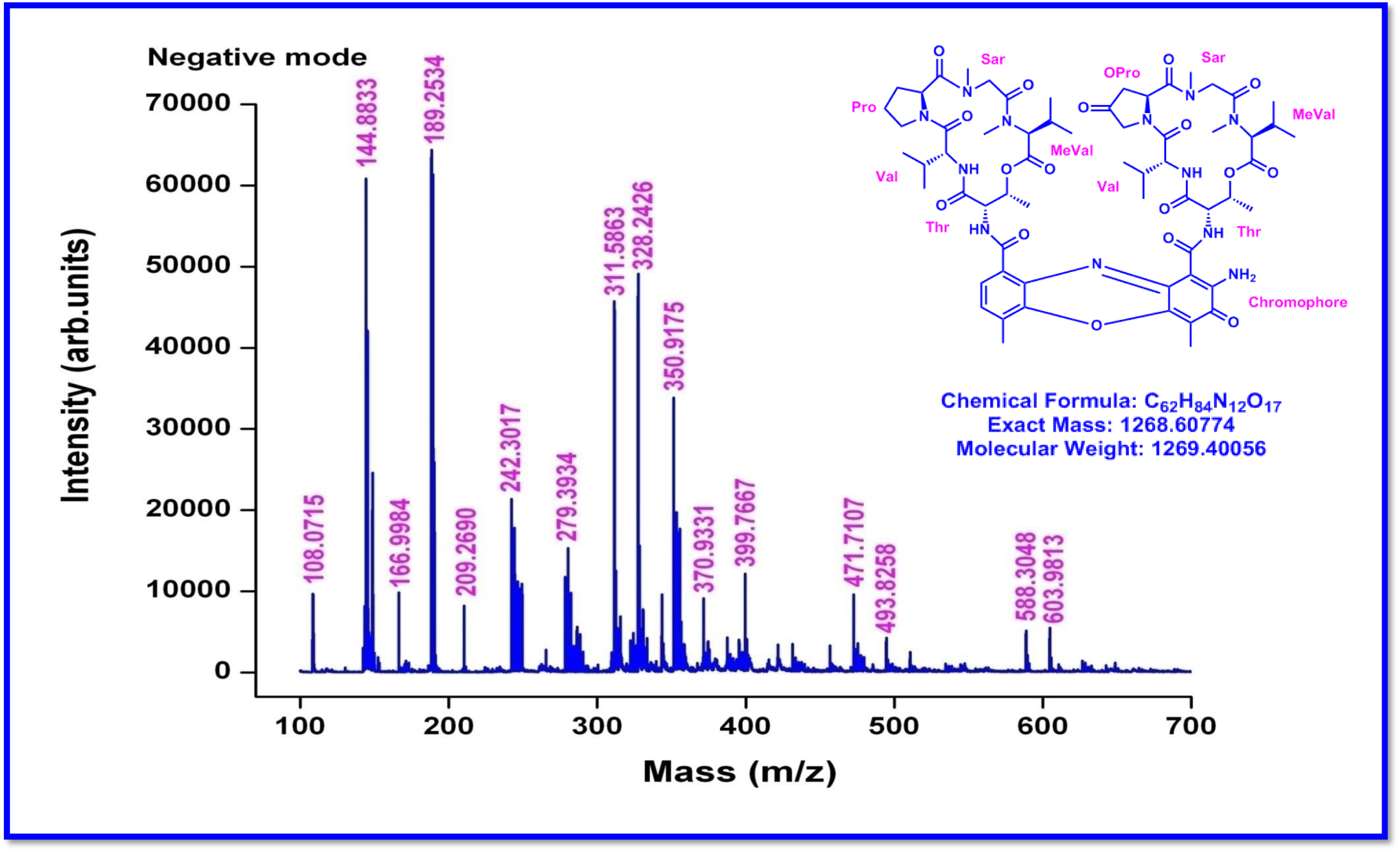
MALDI-TOF MS Spectrum of Transitmycin (R1) (Negative mode)

**Table S6.**
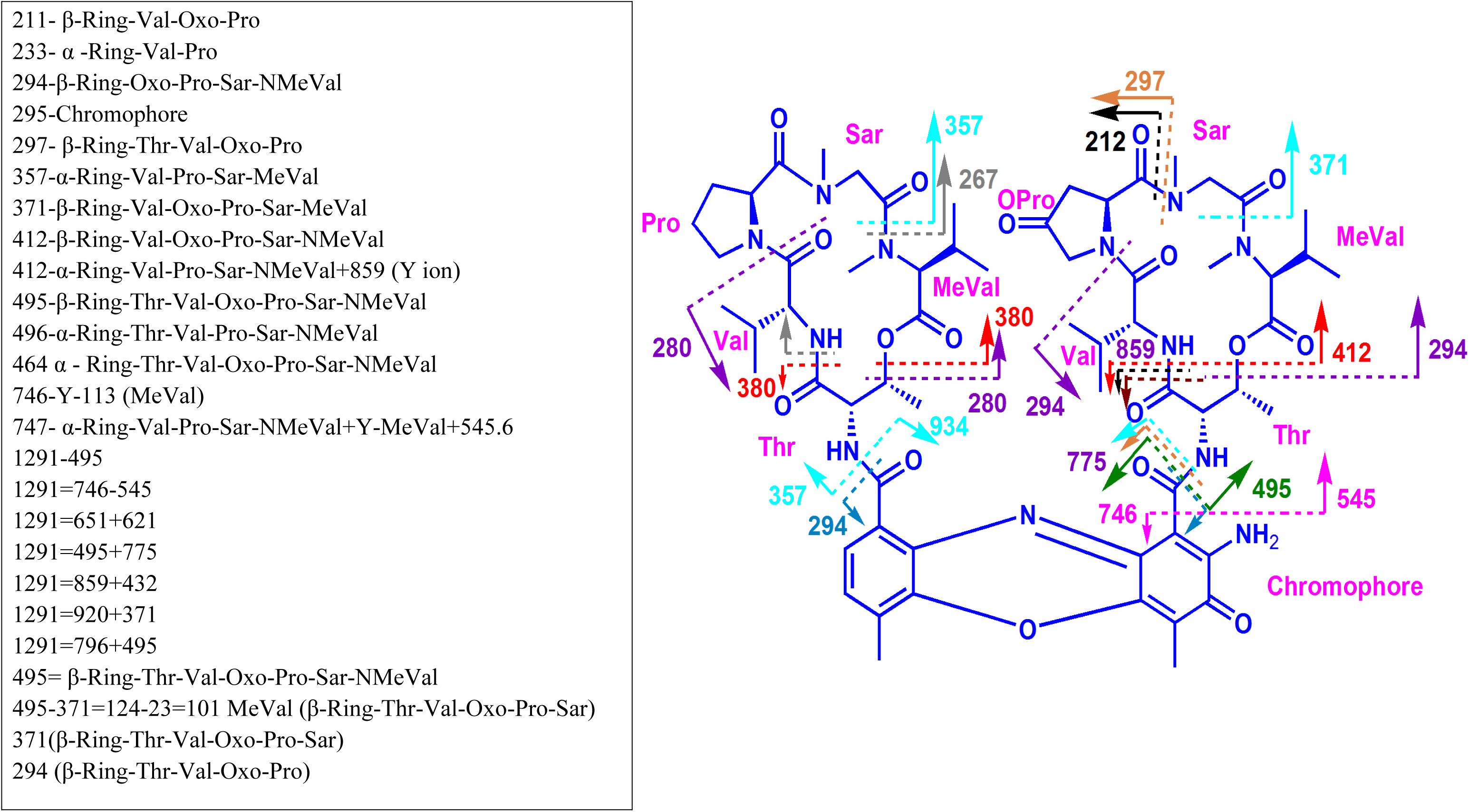
MALDI-TOF MS Spectral Fragmentation of Transitmycin (R1) (Positive mode)

**Figure S80.**
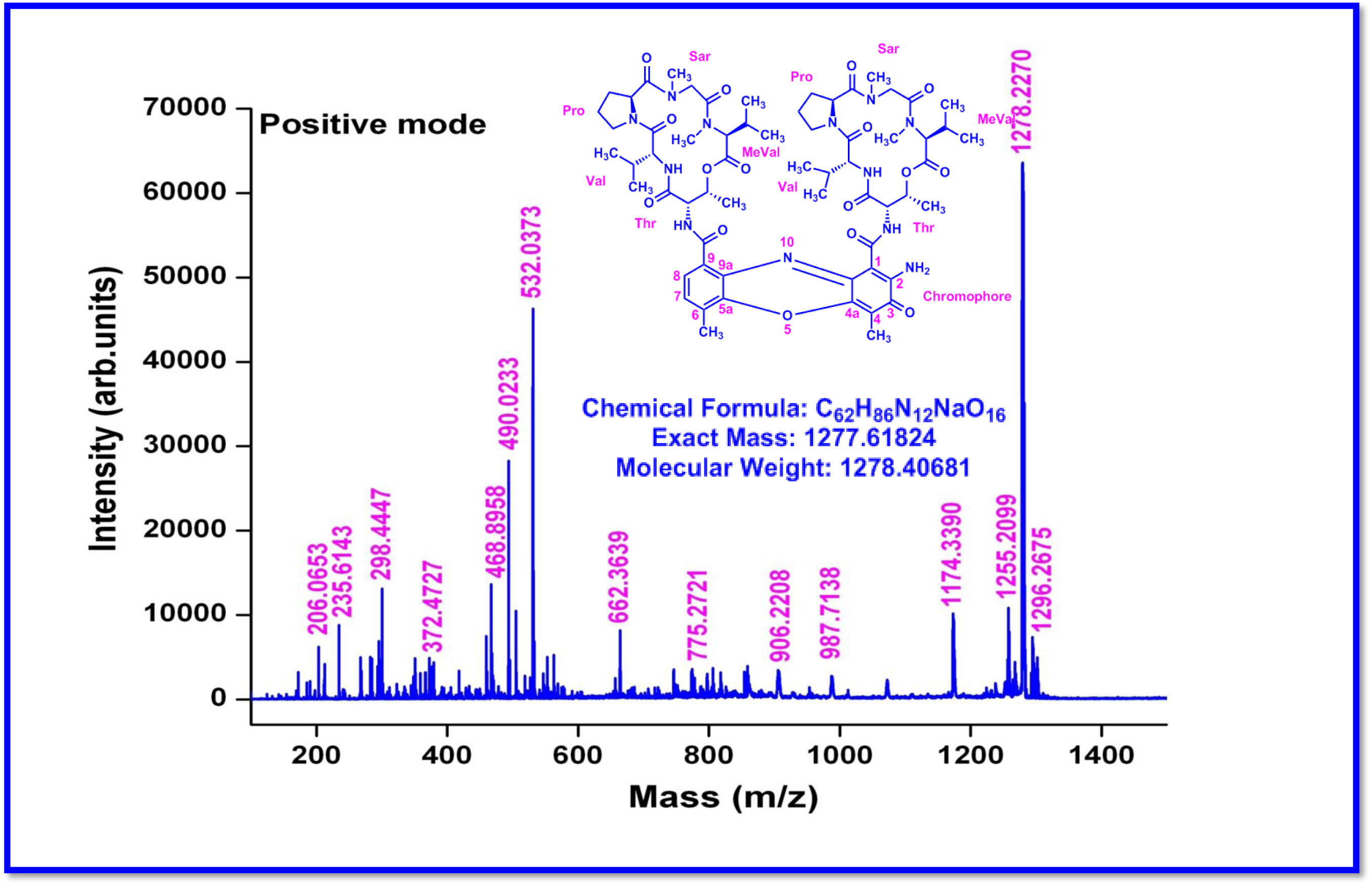
MALDI-TOF MS Spectrum of R2 (Positive mode)

**Figure S81.**
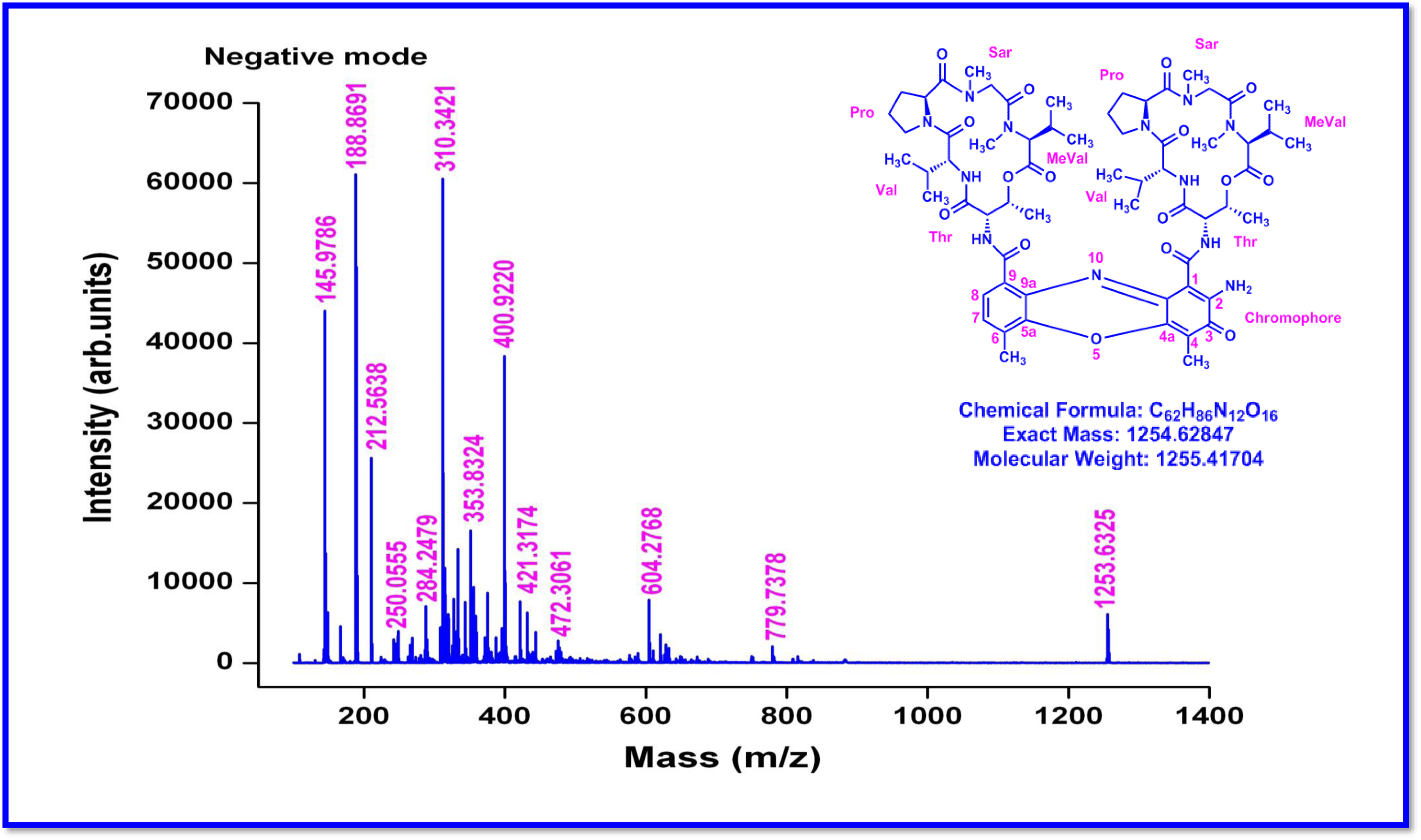
MALDI-TOF MS Spectrum of R2 (Negative mode)

**Figure S82.**
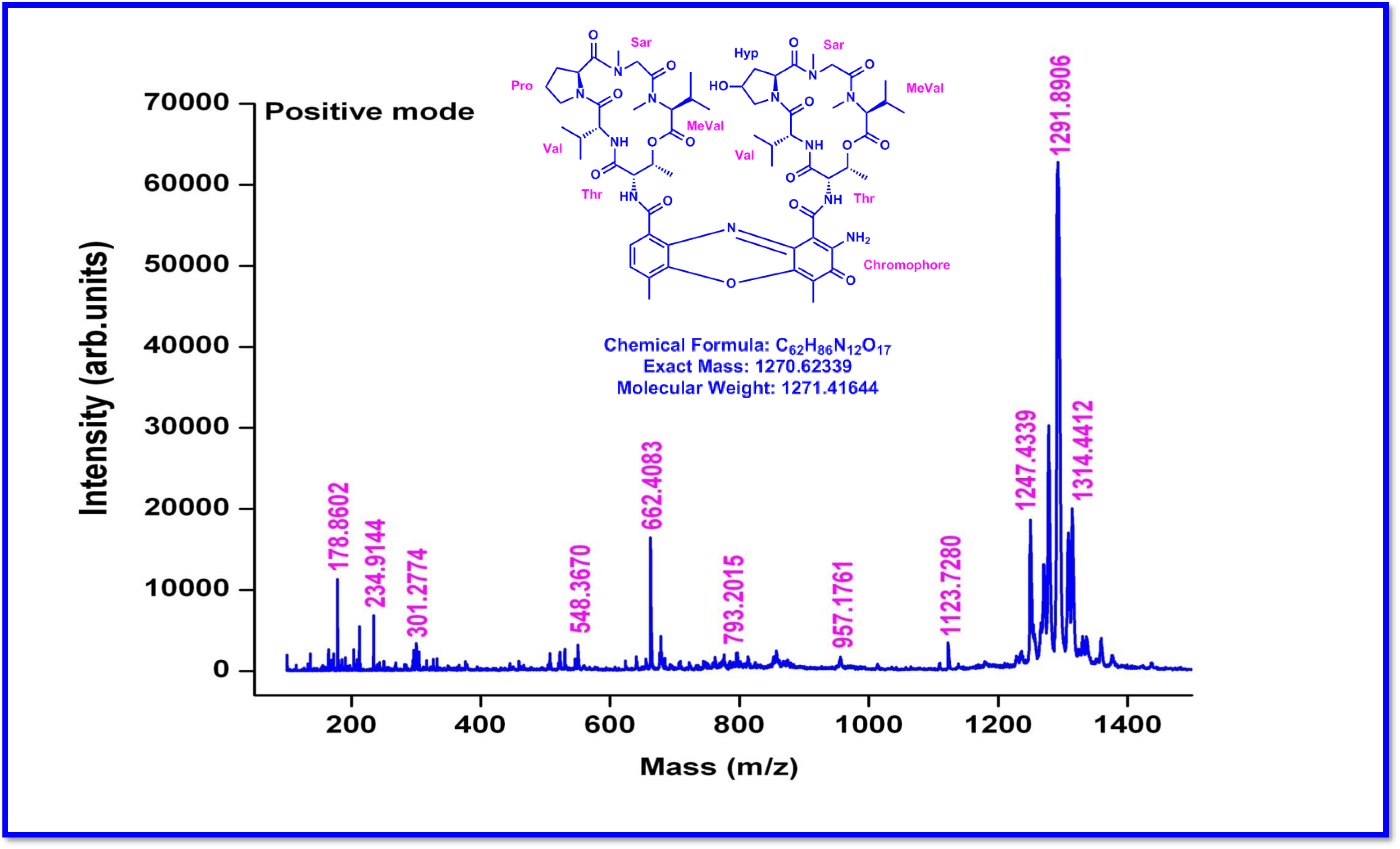
MALDI-TOF MS Spectrum of R3 (Positive mode)

**Figure S83.**
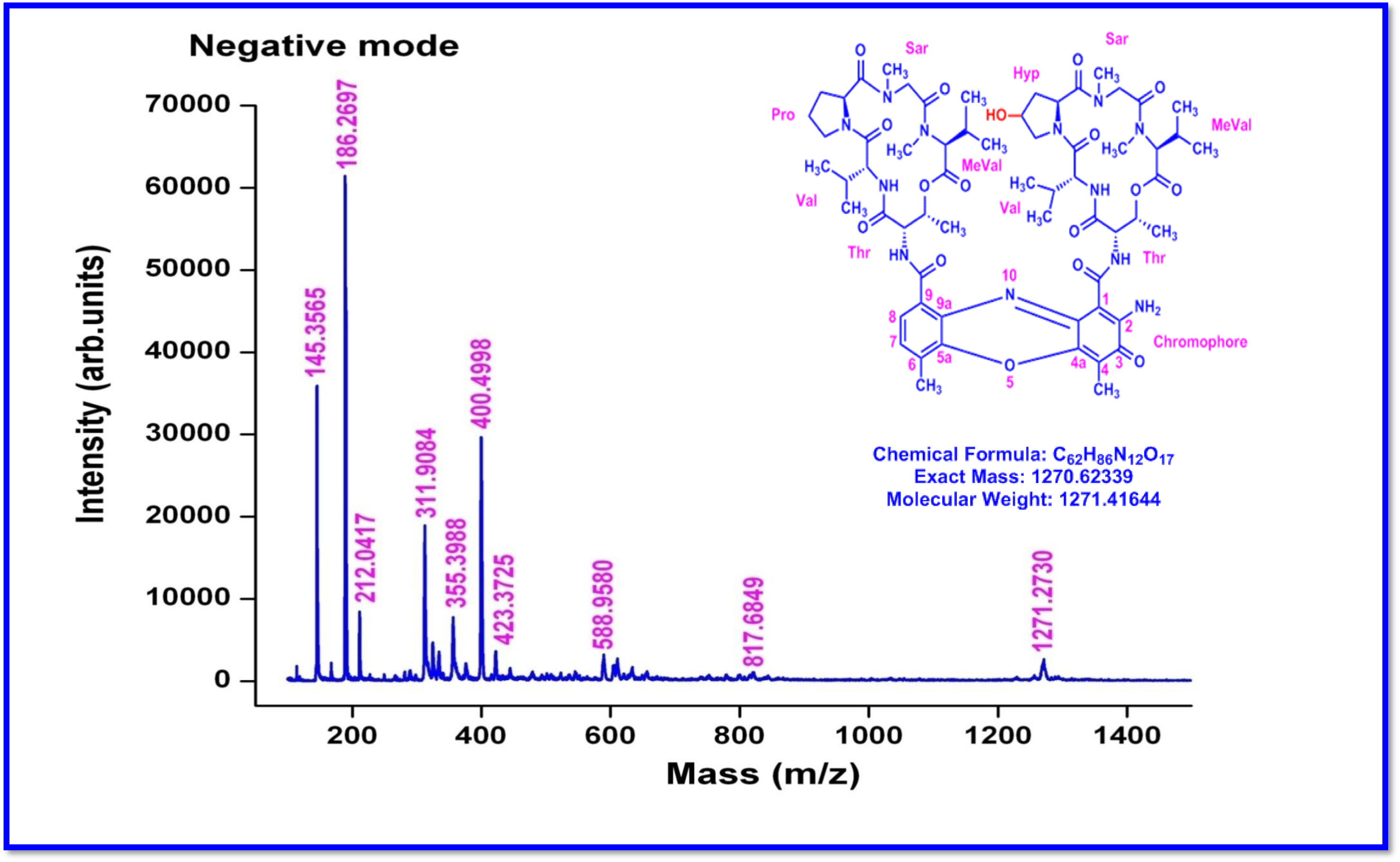
MALDI-TOF MS Spectrum of R3 (Negative mode)

**Figure S84.**
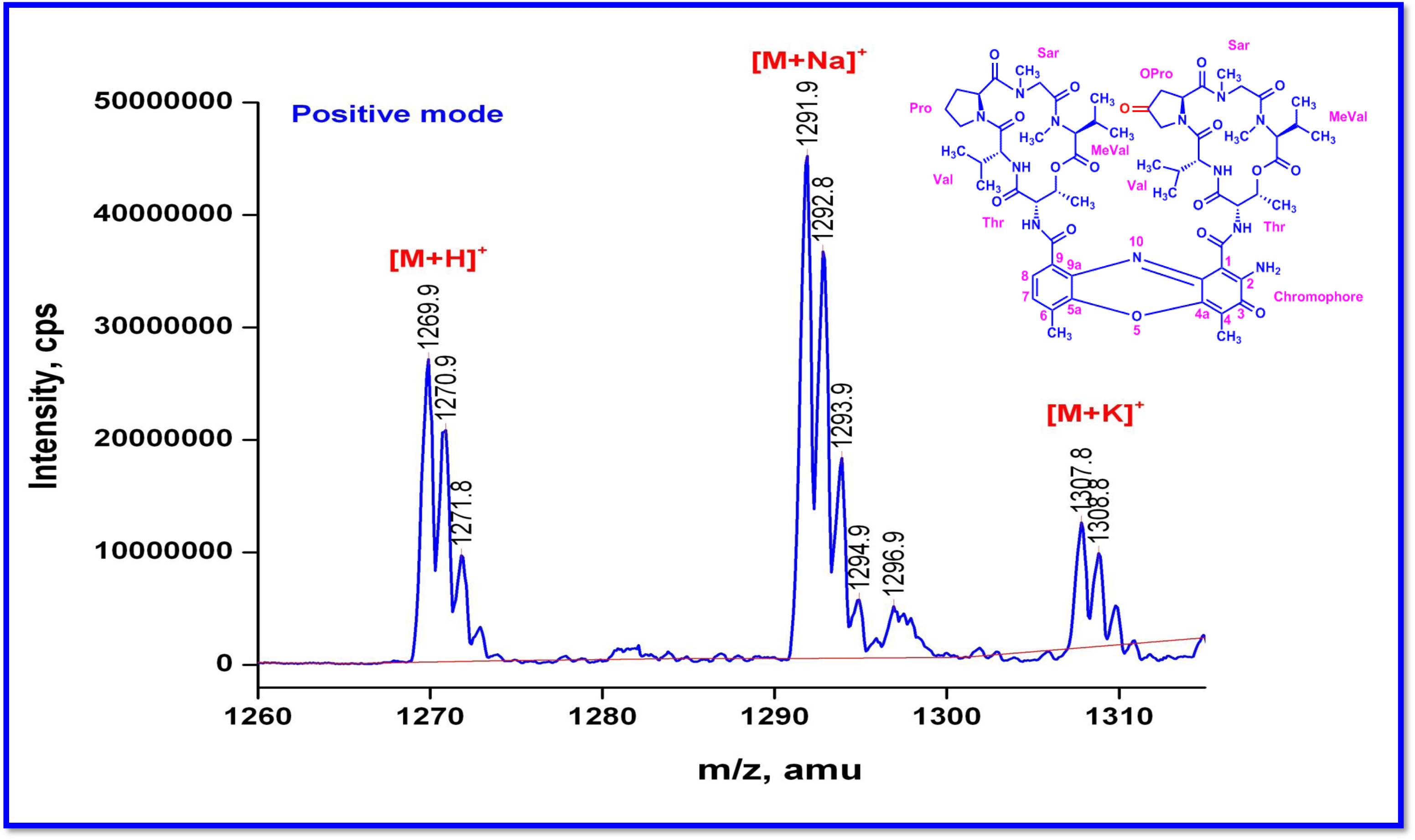
QTRAP MS/MS Transitmycin (R1) (Molecular ion peak) (Positive mode)

**Figure S85.**
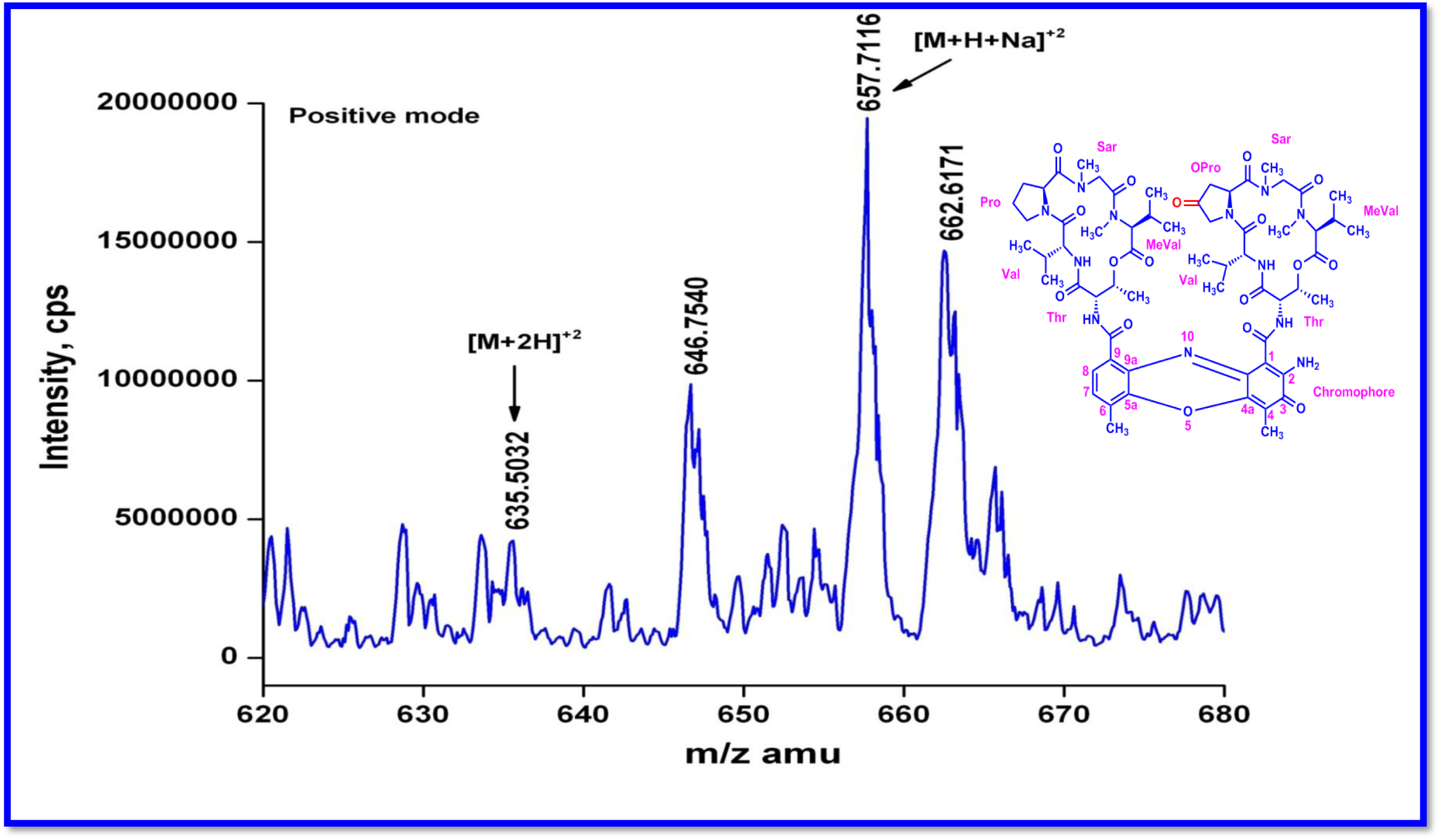
QTRAP MS/MS Transitmycin (R1) (Molecular ion peak [M+H]^2+^ (Positive mode)

**Figure S86.**
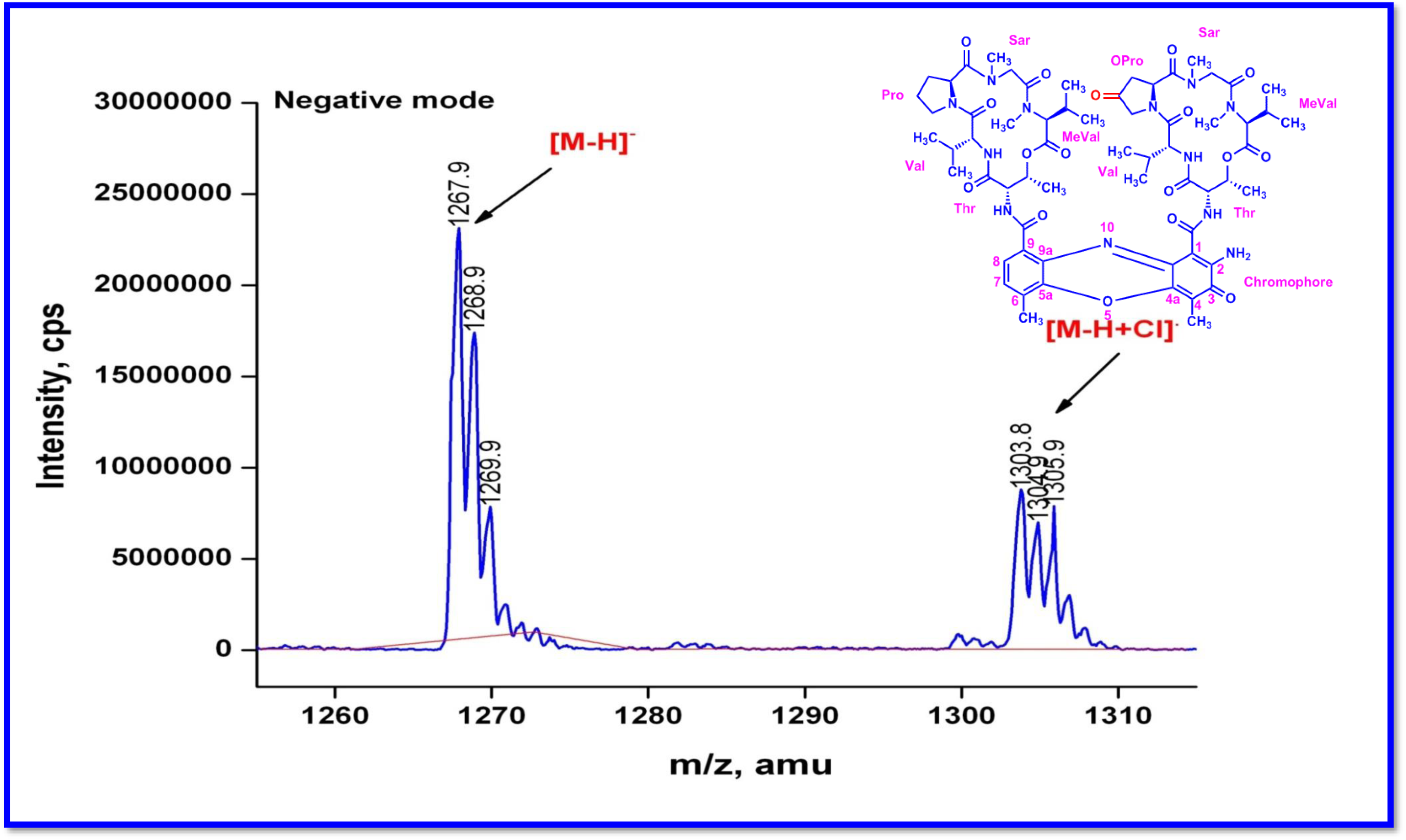
QTRAP MS/MS of Transitmycin (R1) (Molecular ion peak) (Negative mode)

**Figure S87.**
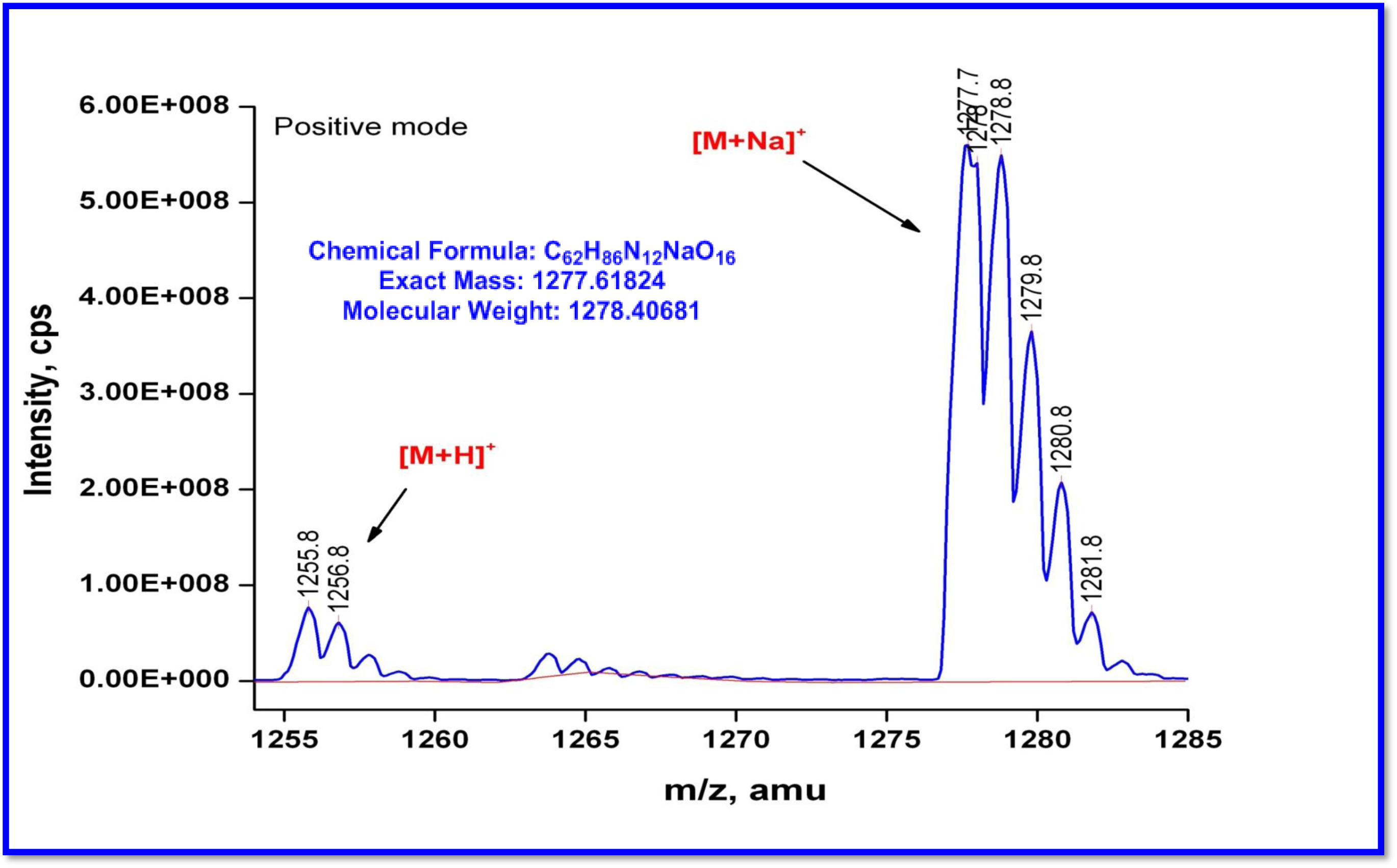
QTRAP MS/MS of R2 (Molecular ion peak) ((Positive mode)

**Figure S88.**
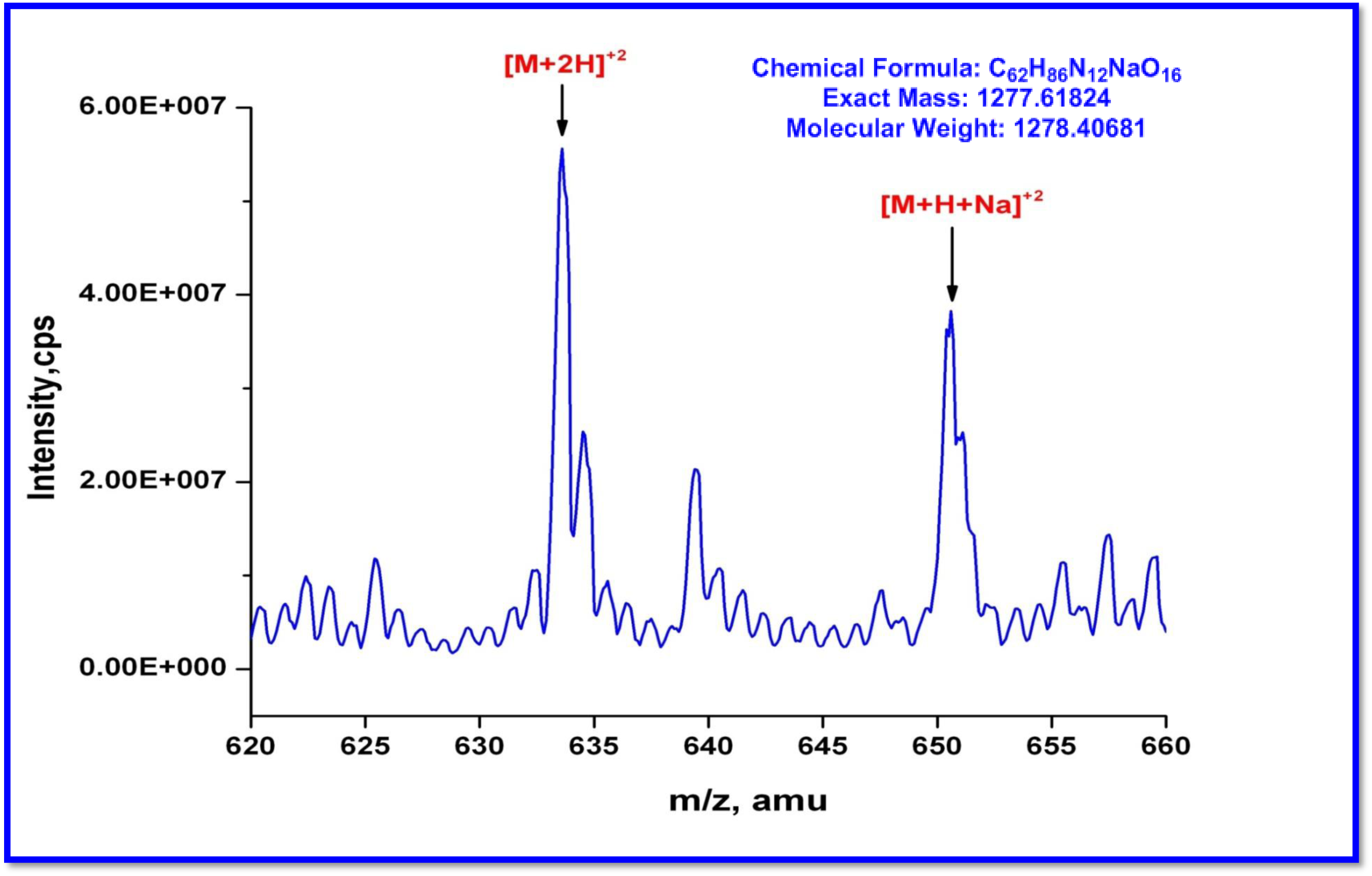
QTRAP MS/MS of R2 [M+H]^2+^ ion peak (Positive mode)

**Figure S89.**
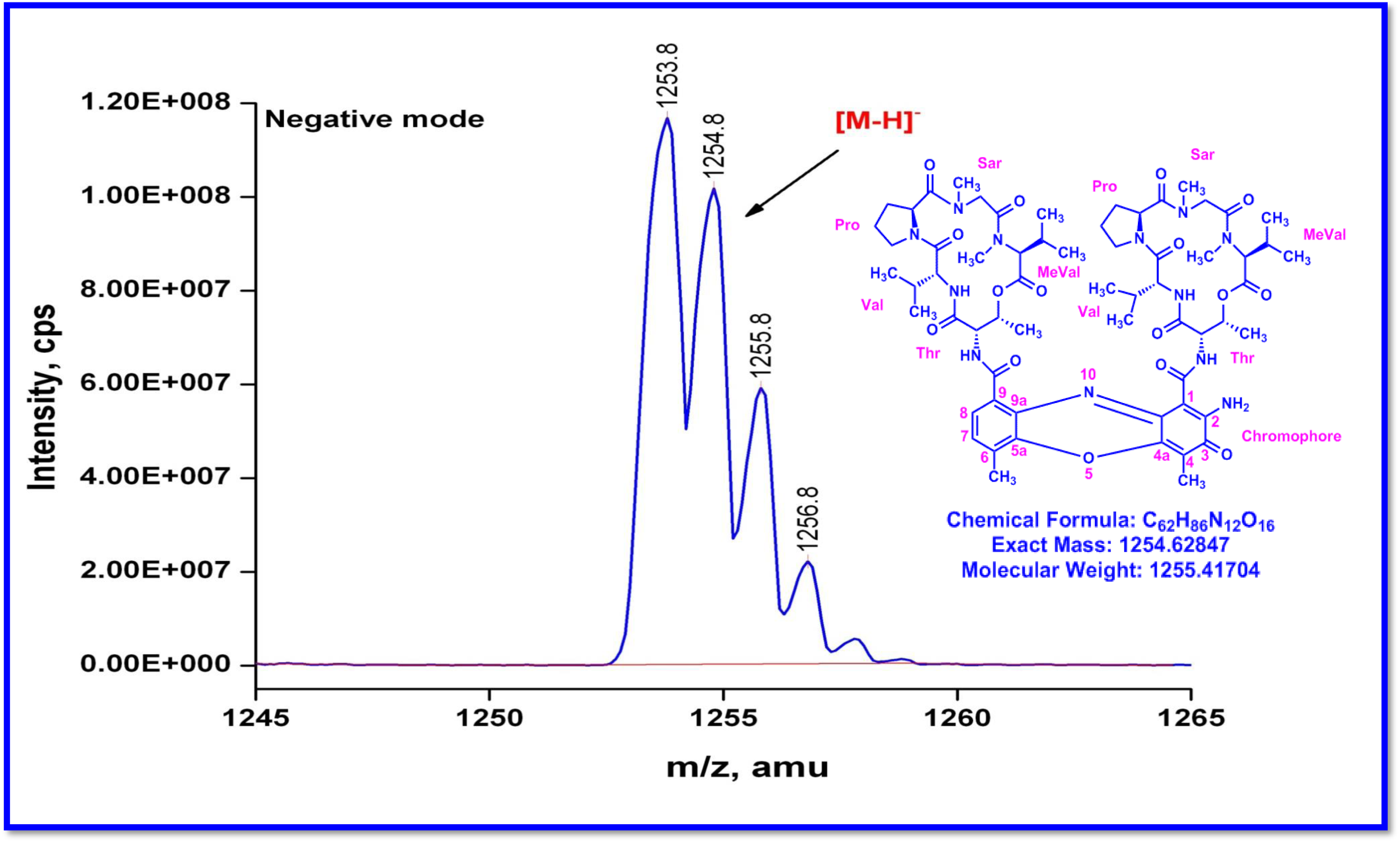
QTRAP MS/MS of R2 (Molecular ion peak (Positive mode)

**Figure S90.**
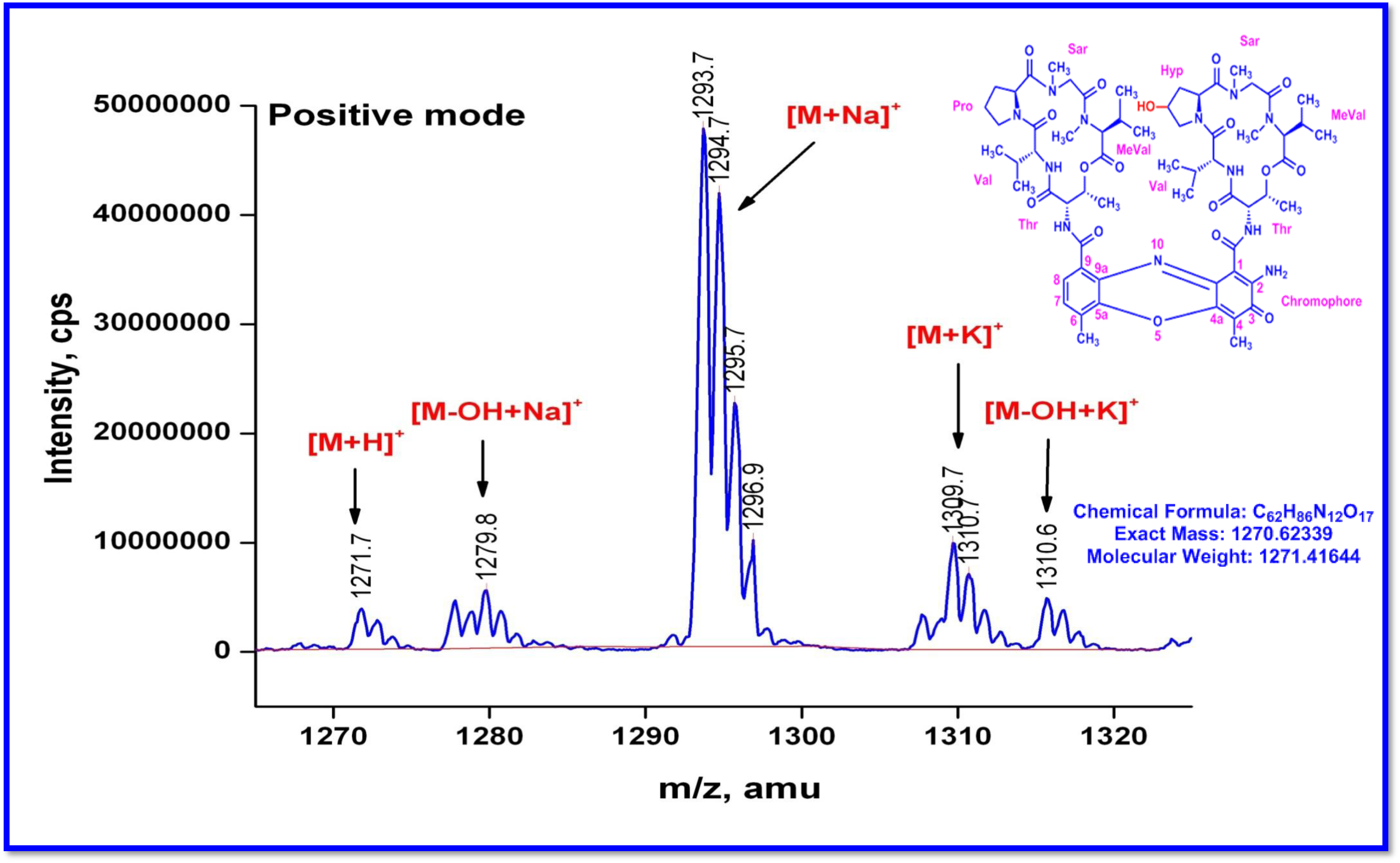
QTRAP MS/MS of R3 Molecular ion peak (Positive mode)

**Figure S91.**
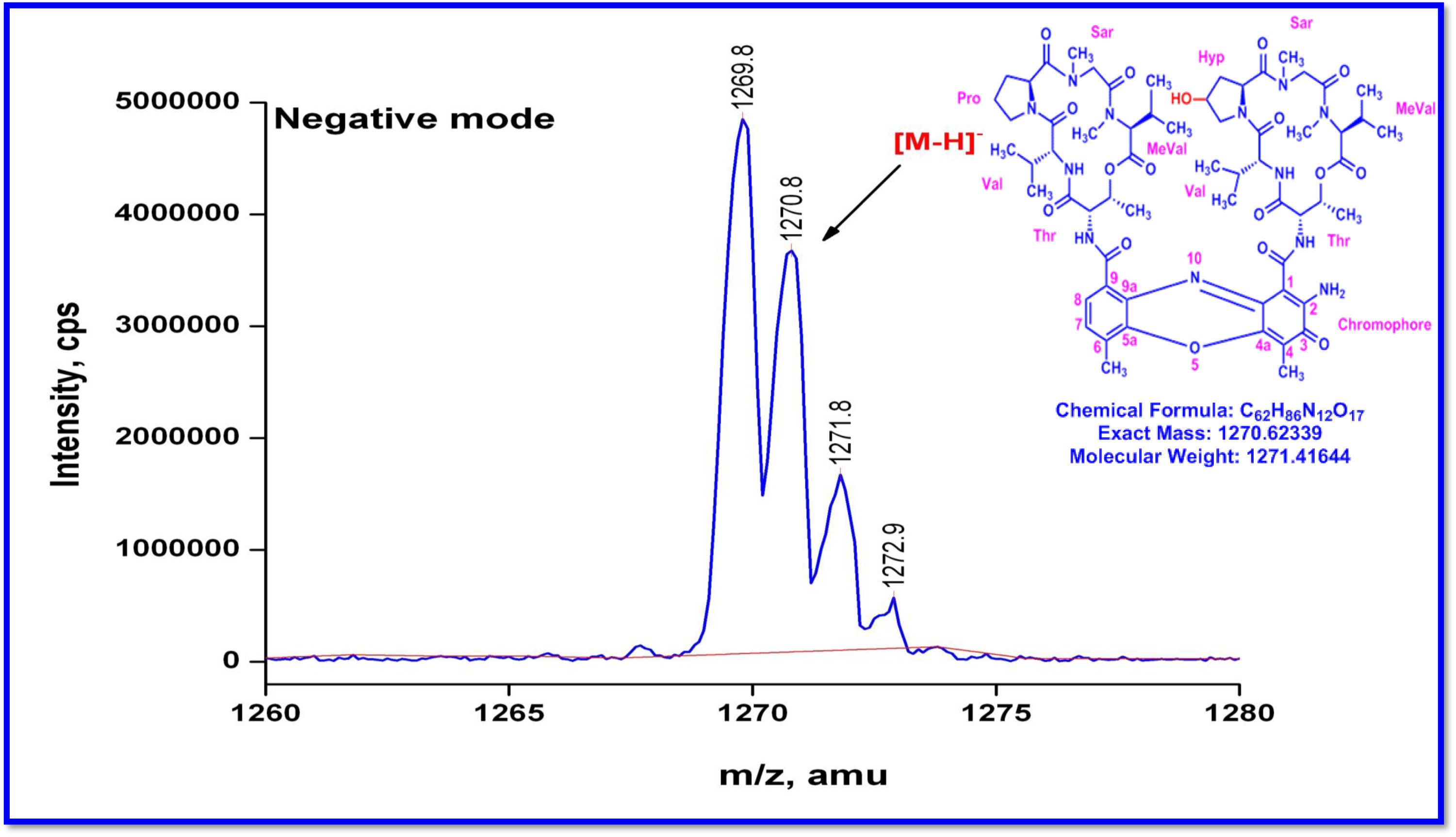
QTRAP MS/MS of R3 (Molecular ion peak (Negative mode)

**Figure S92.**
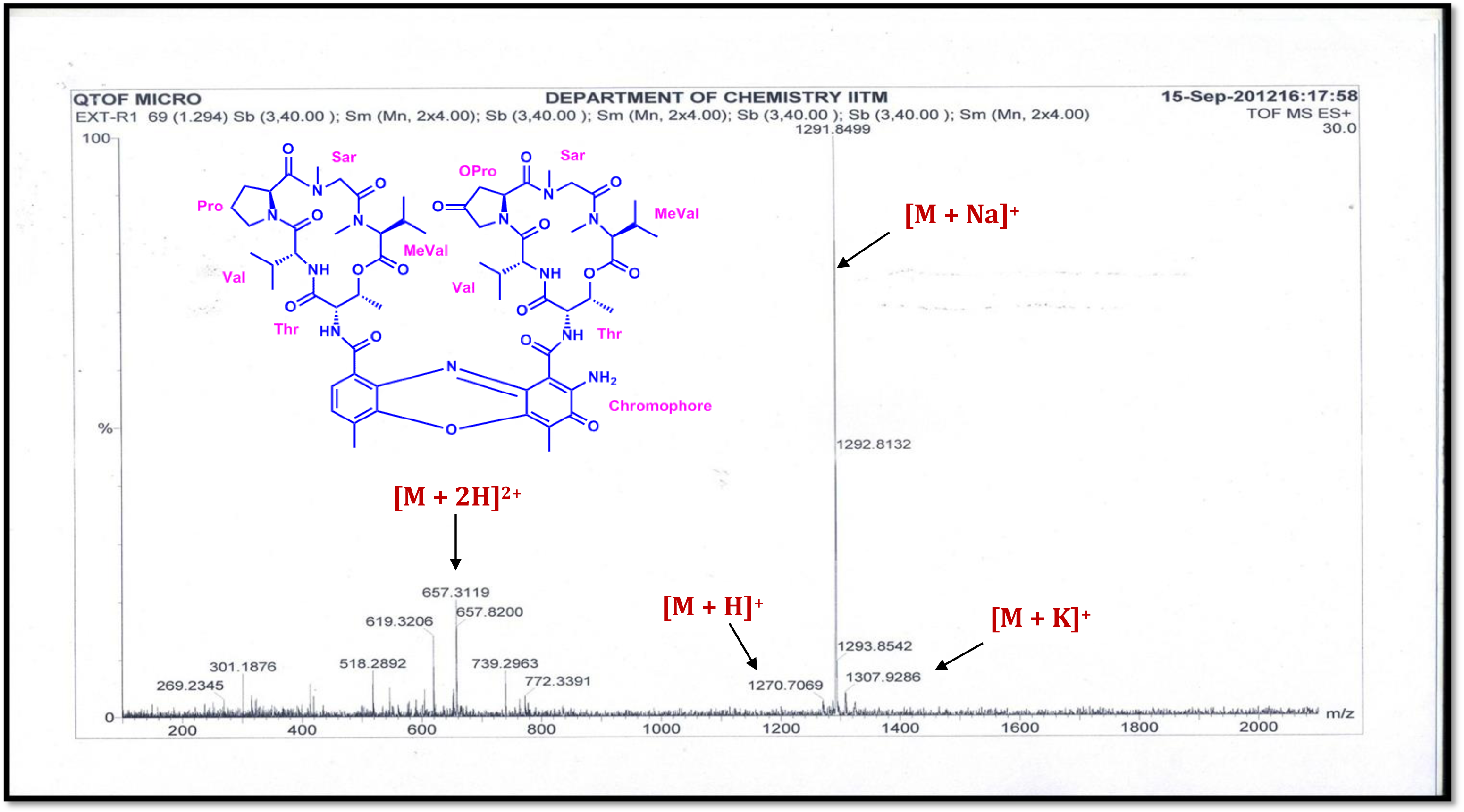
HR-ESI-MS Spectrum of Transitmycin (R1) (Positive mode)

**Figure S93.**
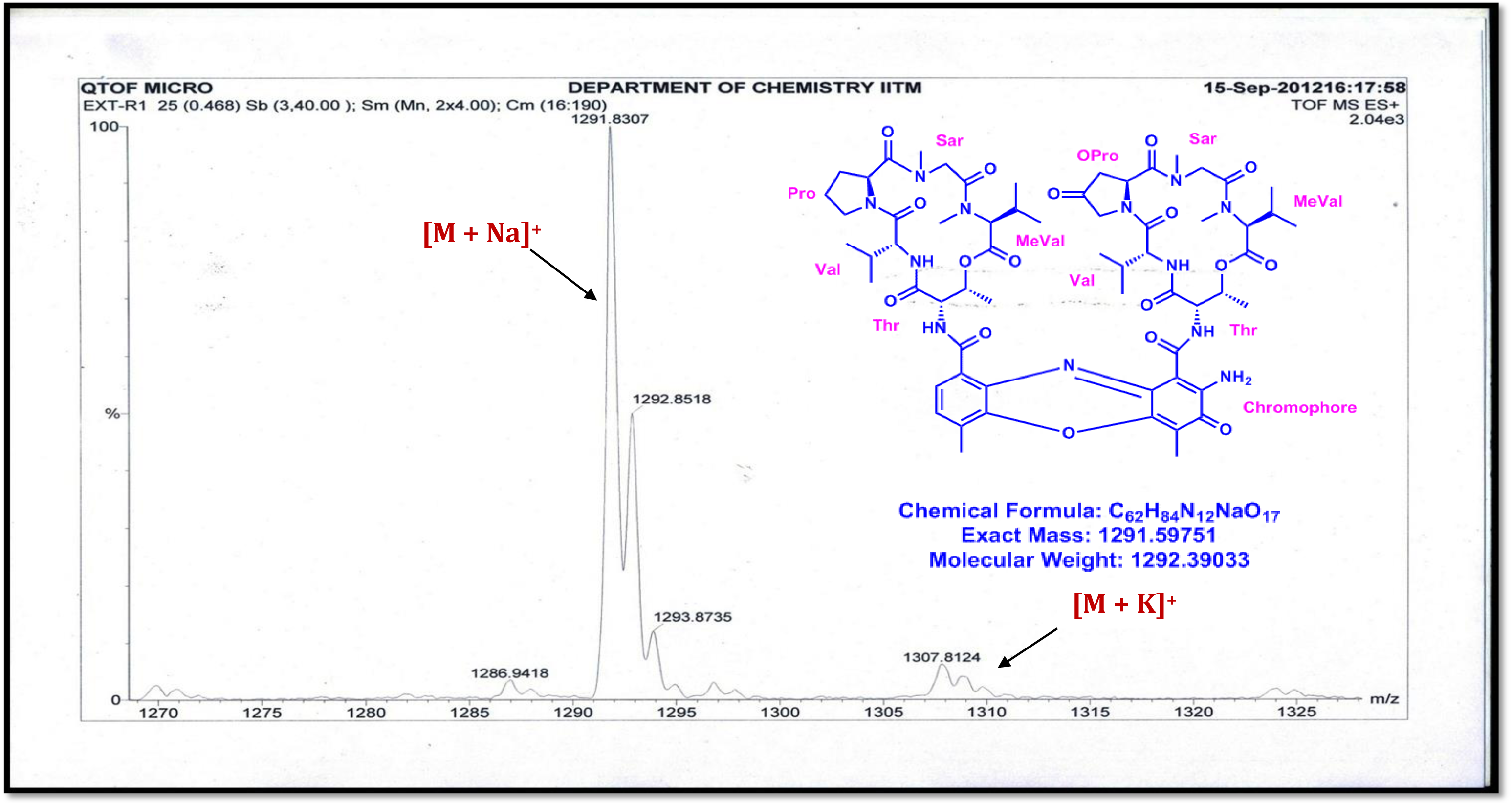
HR-ESI-MS Spectrum of Transitmycin (R1) (Molecular ion Peak) (Positive mode)

**Figure S94.**
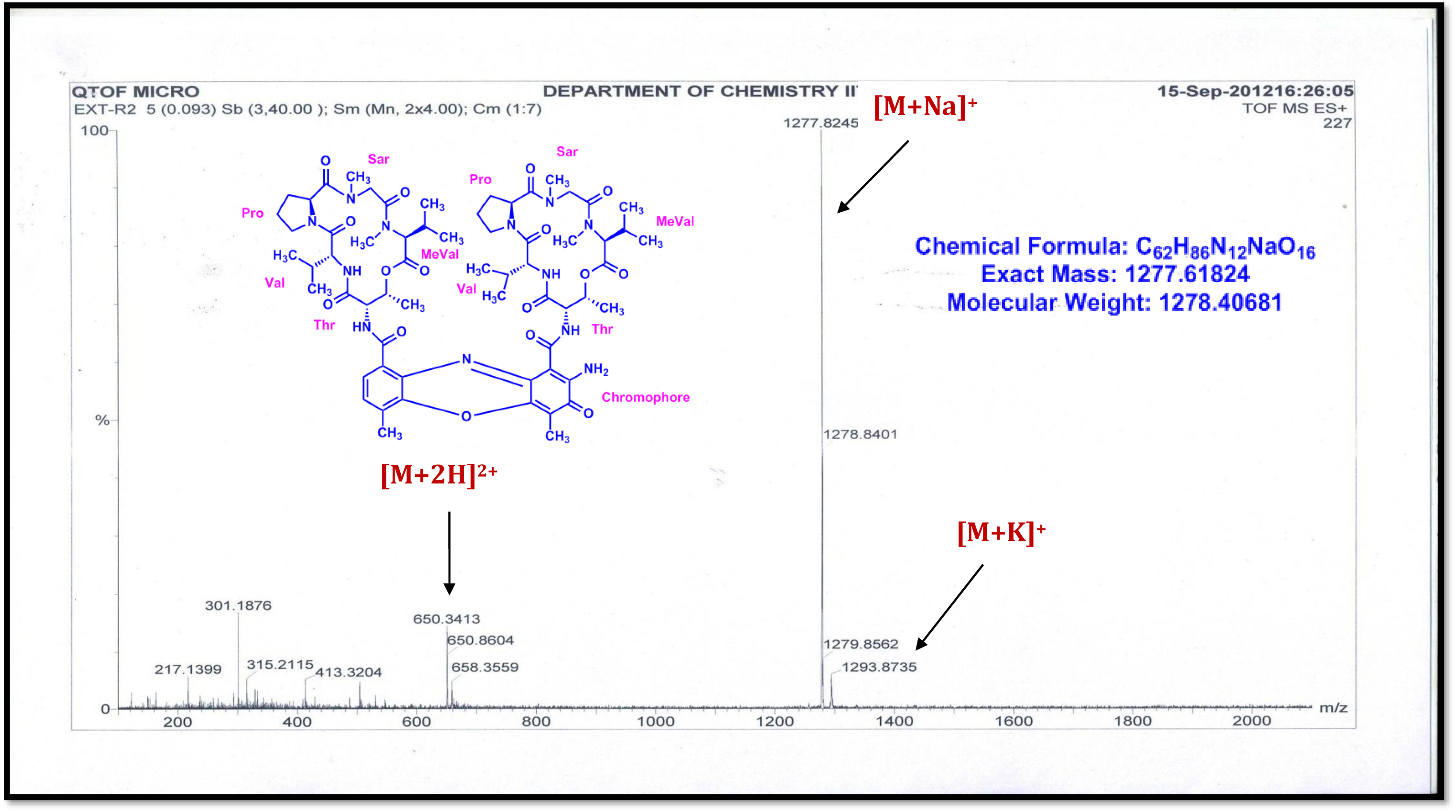
HR-ESI-MS Spectrum of R2 (Positive mode)

**Figure S95.**
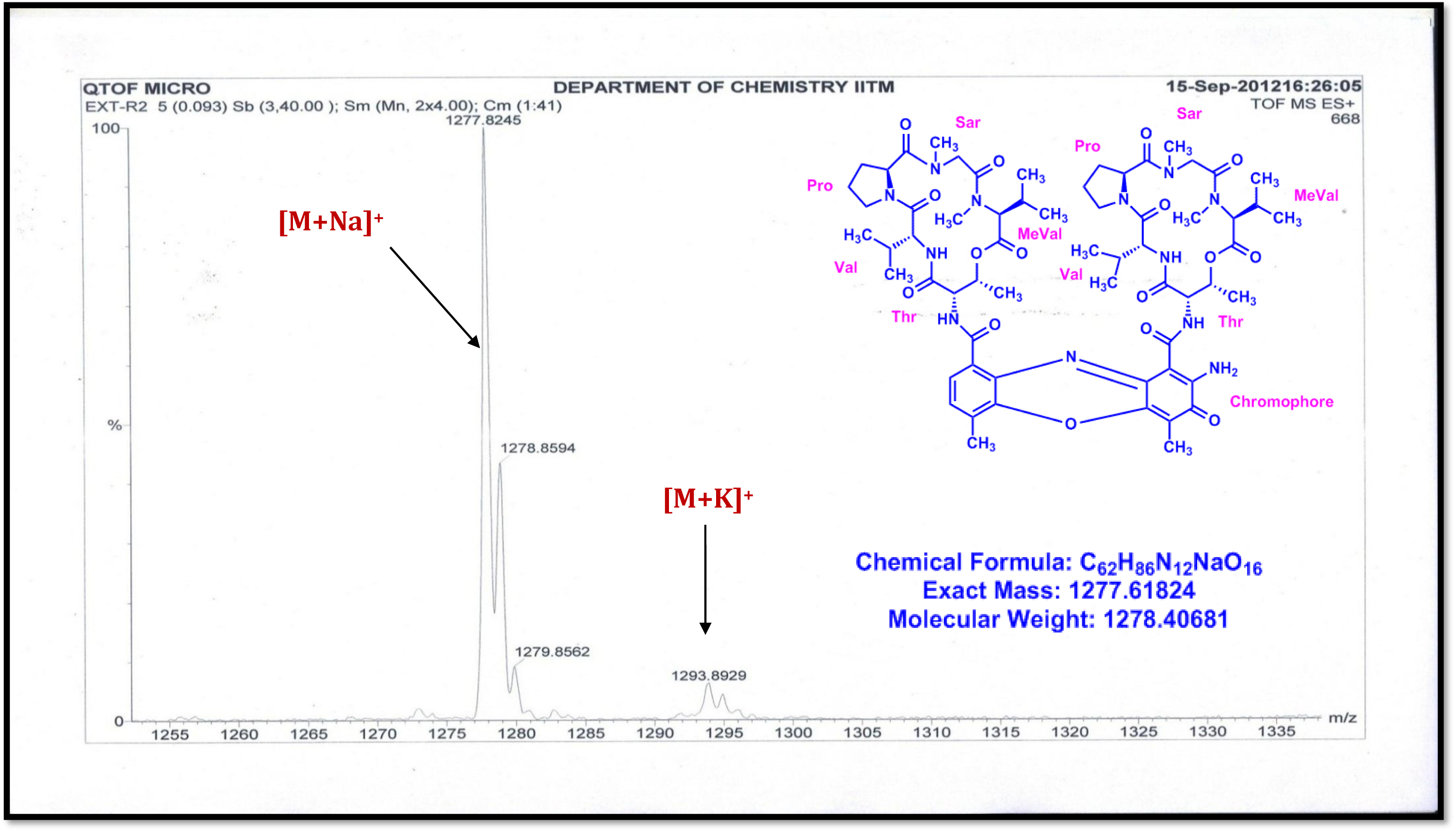
HR-ESI-MS Spectrum of R2 (Molecular ion Peak) (Positive mode)

**Figure S96.**
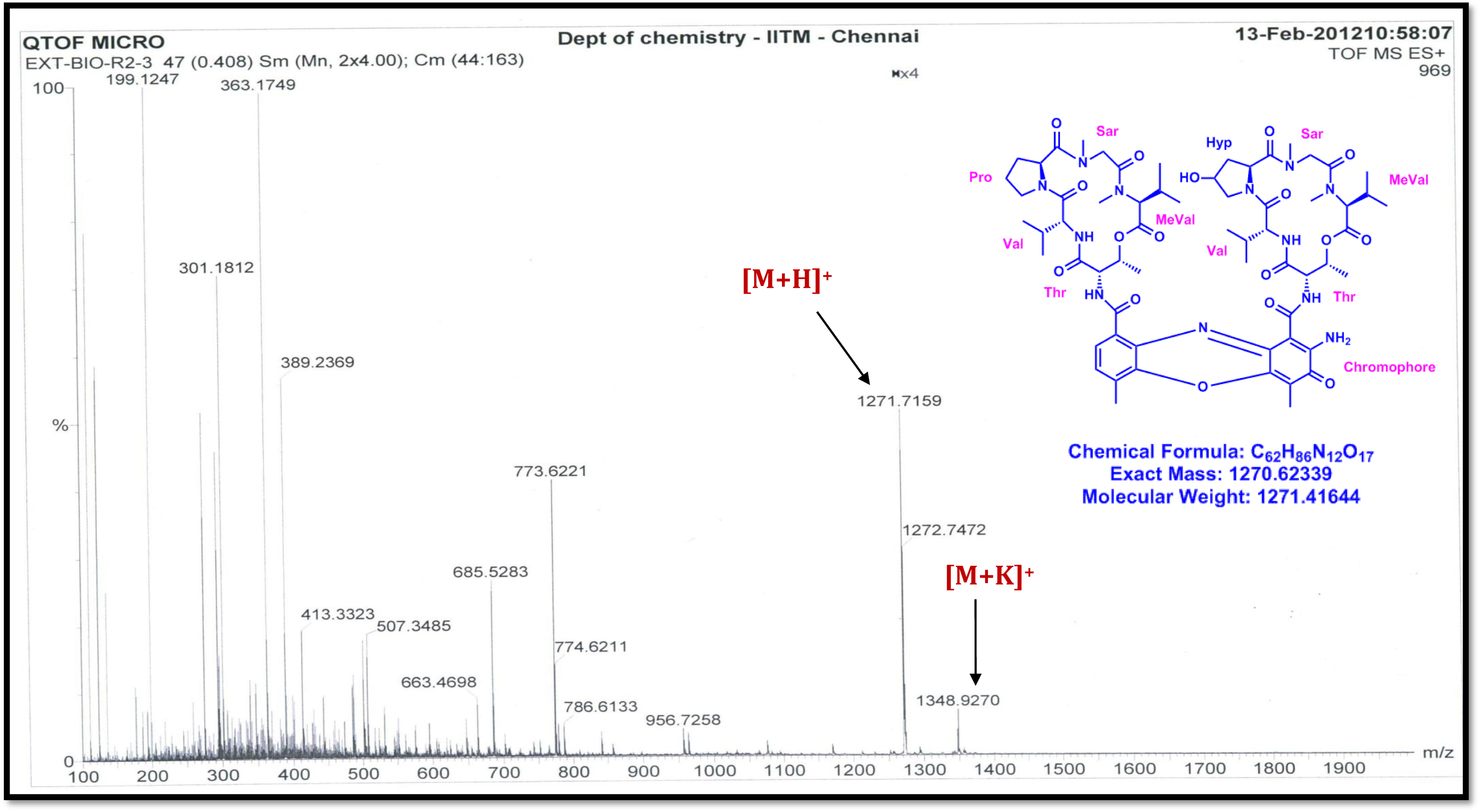
HR-ESI-MS Spectrum of R3 (Positive mode)

**Figure S97.**
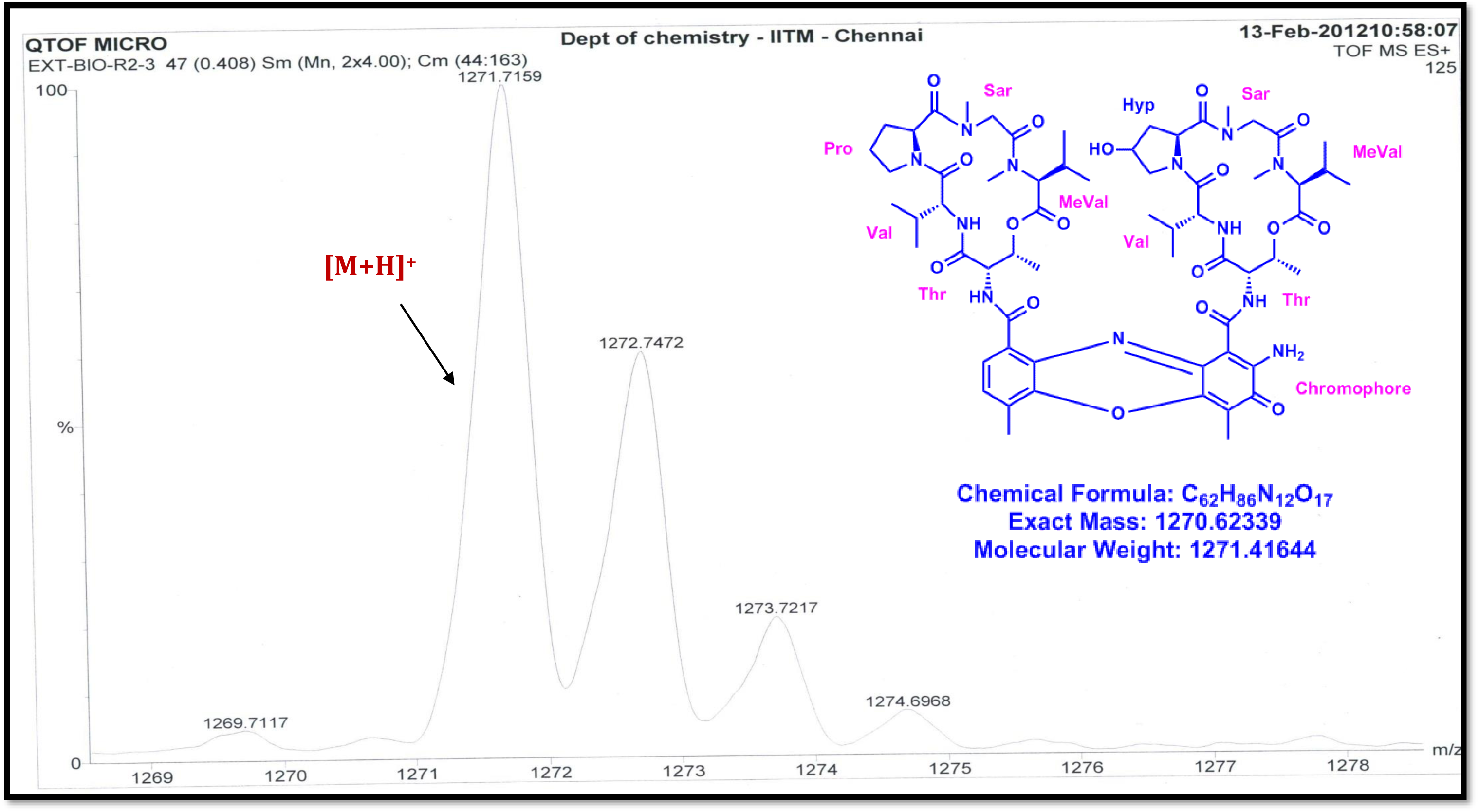
HR-ESI-MS Spectrum of R3 (Molecular ion Peak) (Positive mode)

**Figure S98.**
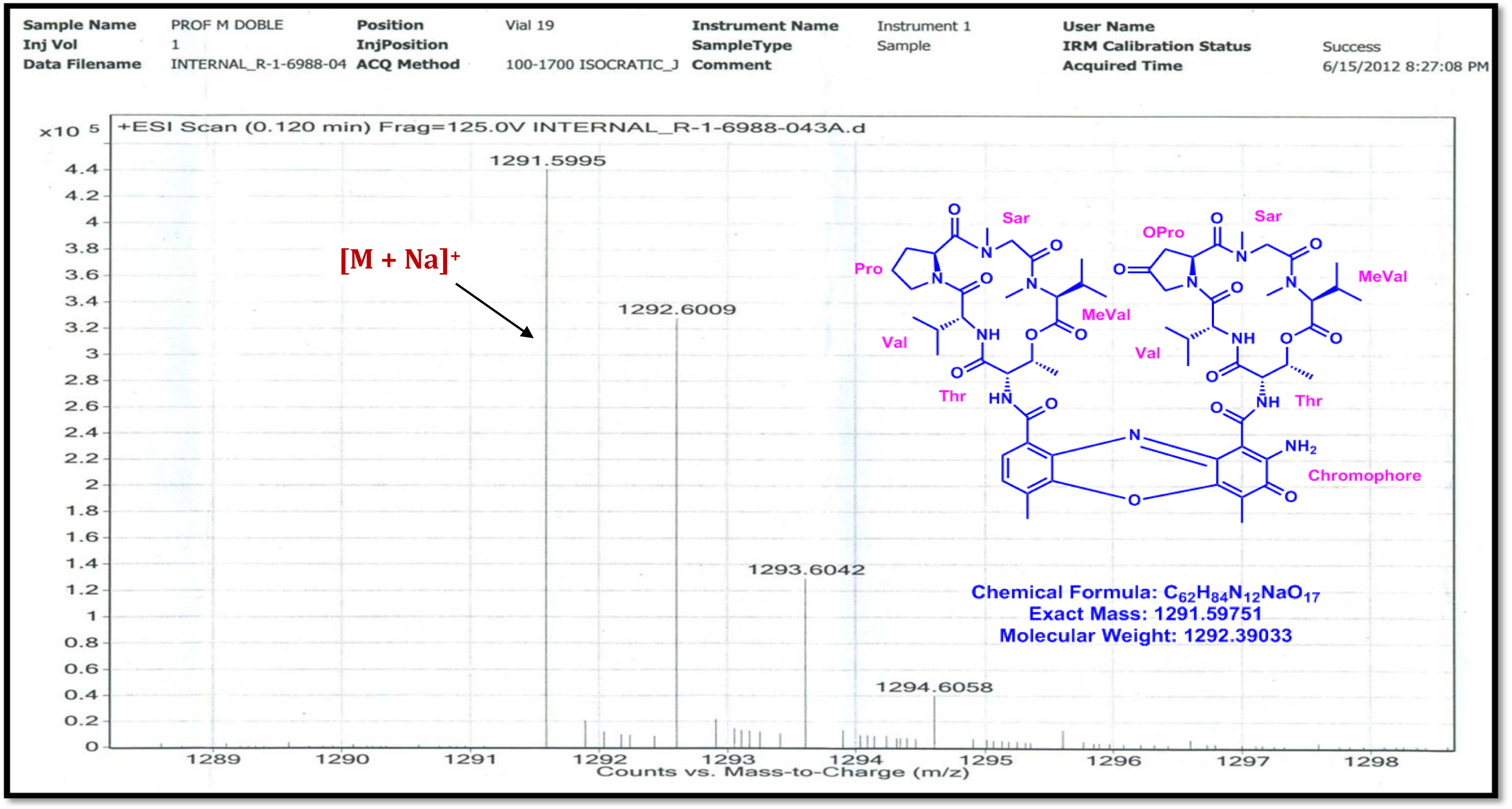
LC-ESI-MS Spectrum of Transitmycin (R1) (Molecular ion peak) (Positive mode)

**Figure S99.**
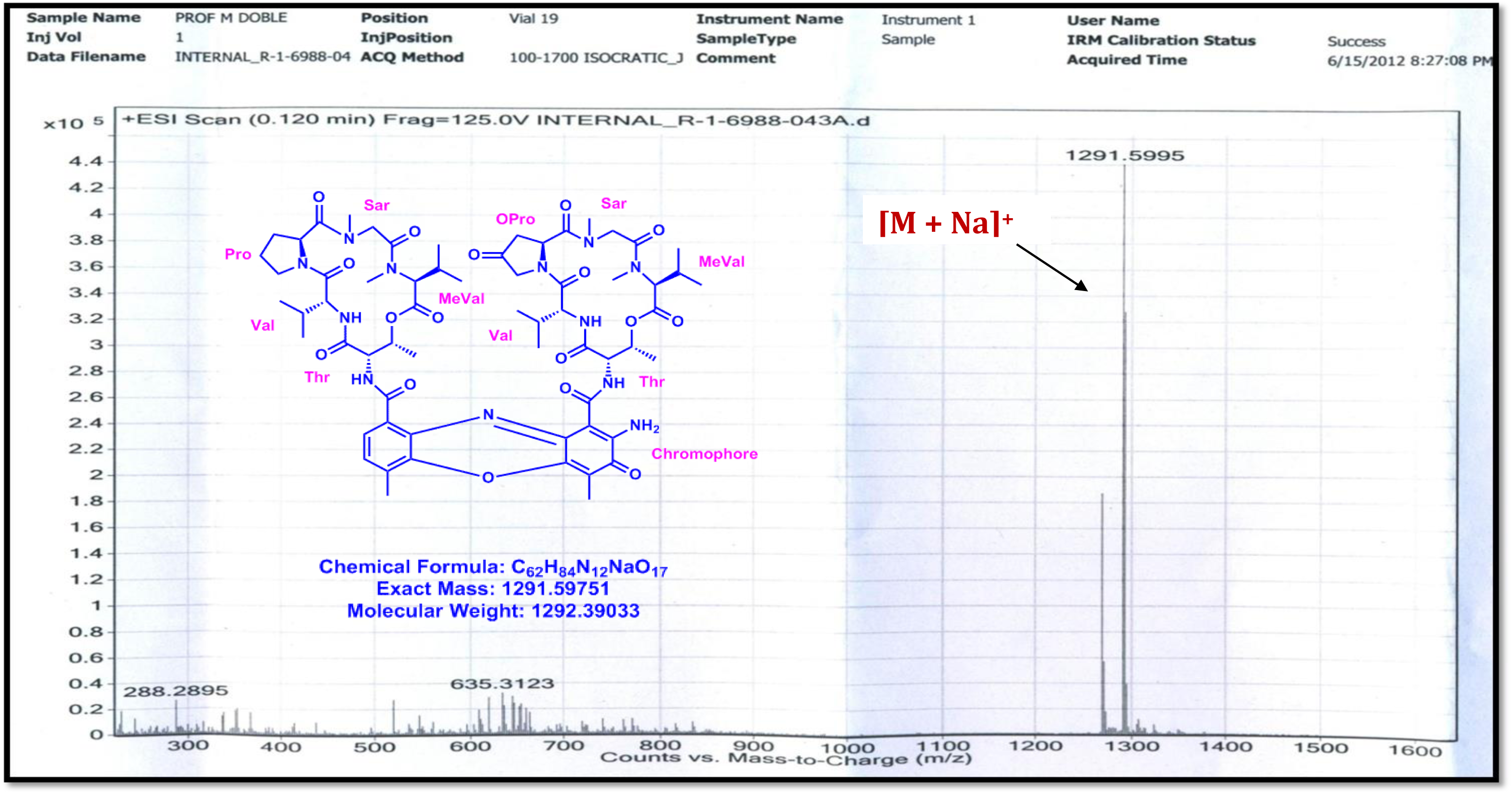
LC-ESI-MS Spectrum of Transitmycin (R1) (Positive mode)

**Figure S100.**
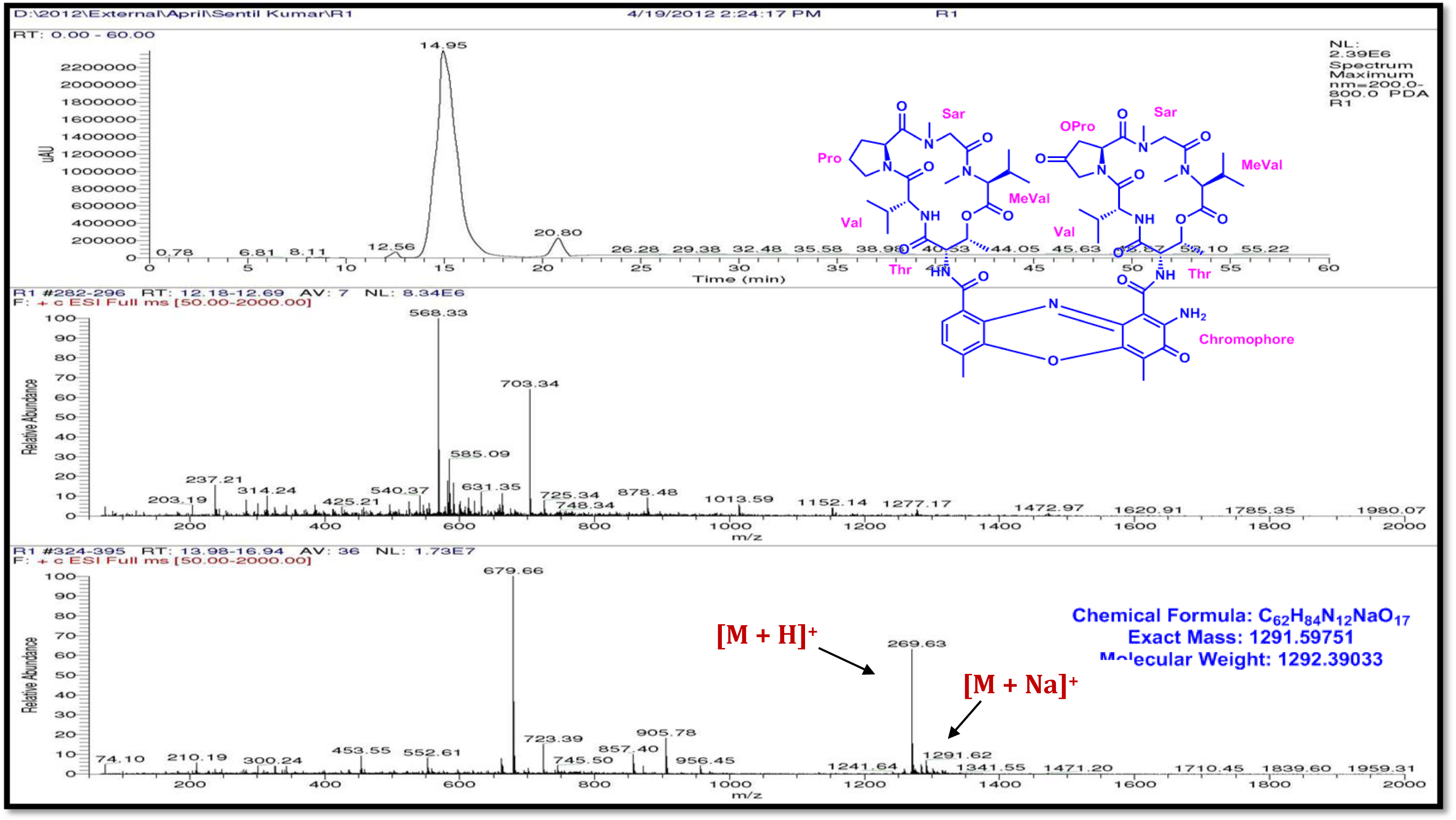
LC-ESI-MS Spectrum of Trasitmycin (R1) (Positive mode)

**Figure S101.**
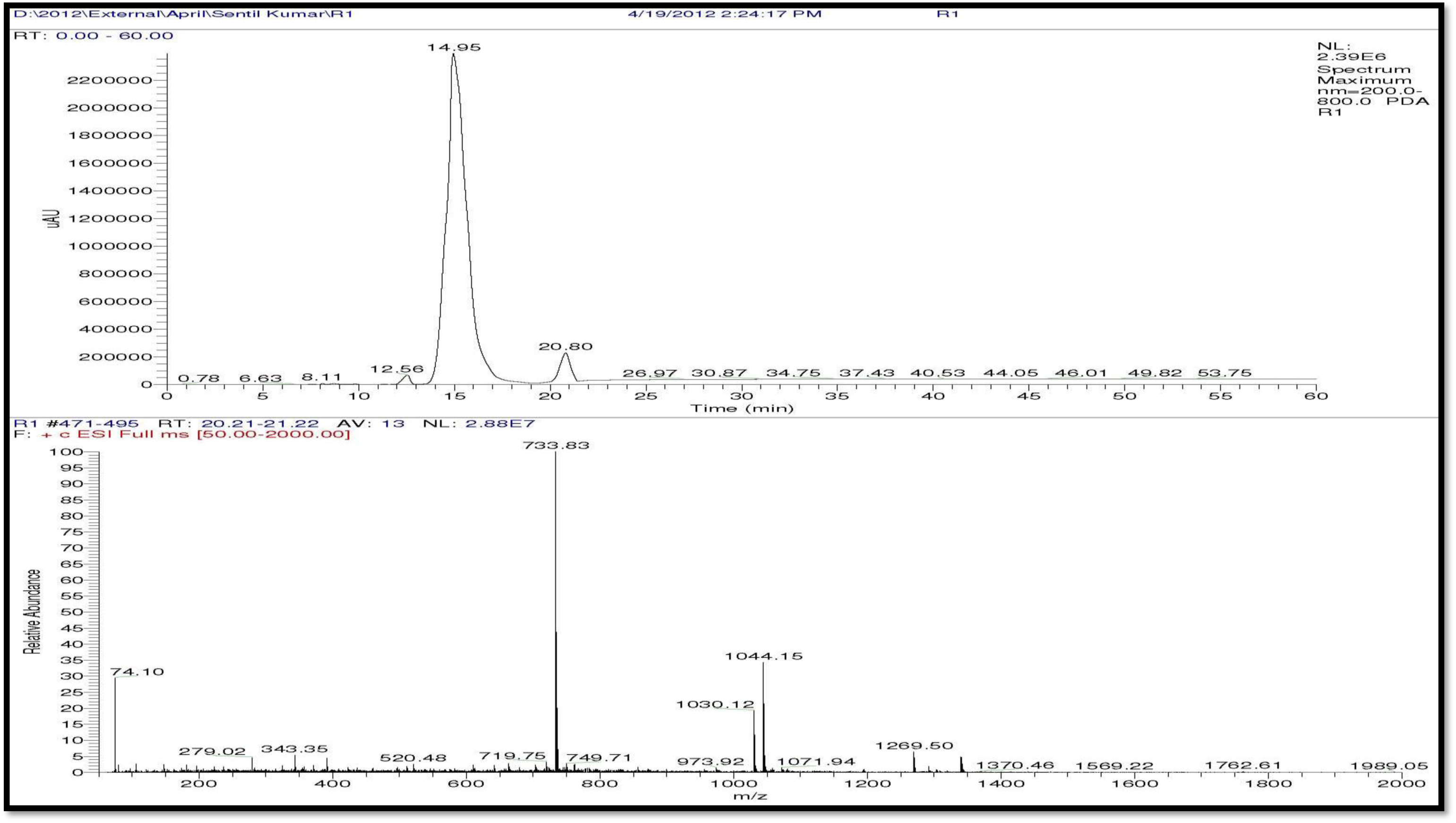
LC-ESI-MS Spectrum of Trasitmycin (R1) (Positive mode)

**Figure S102.**
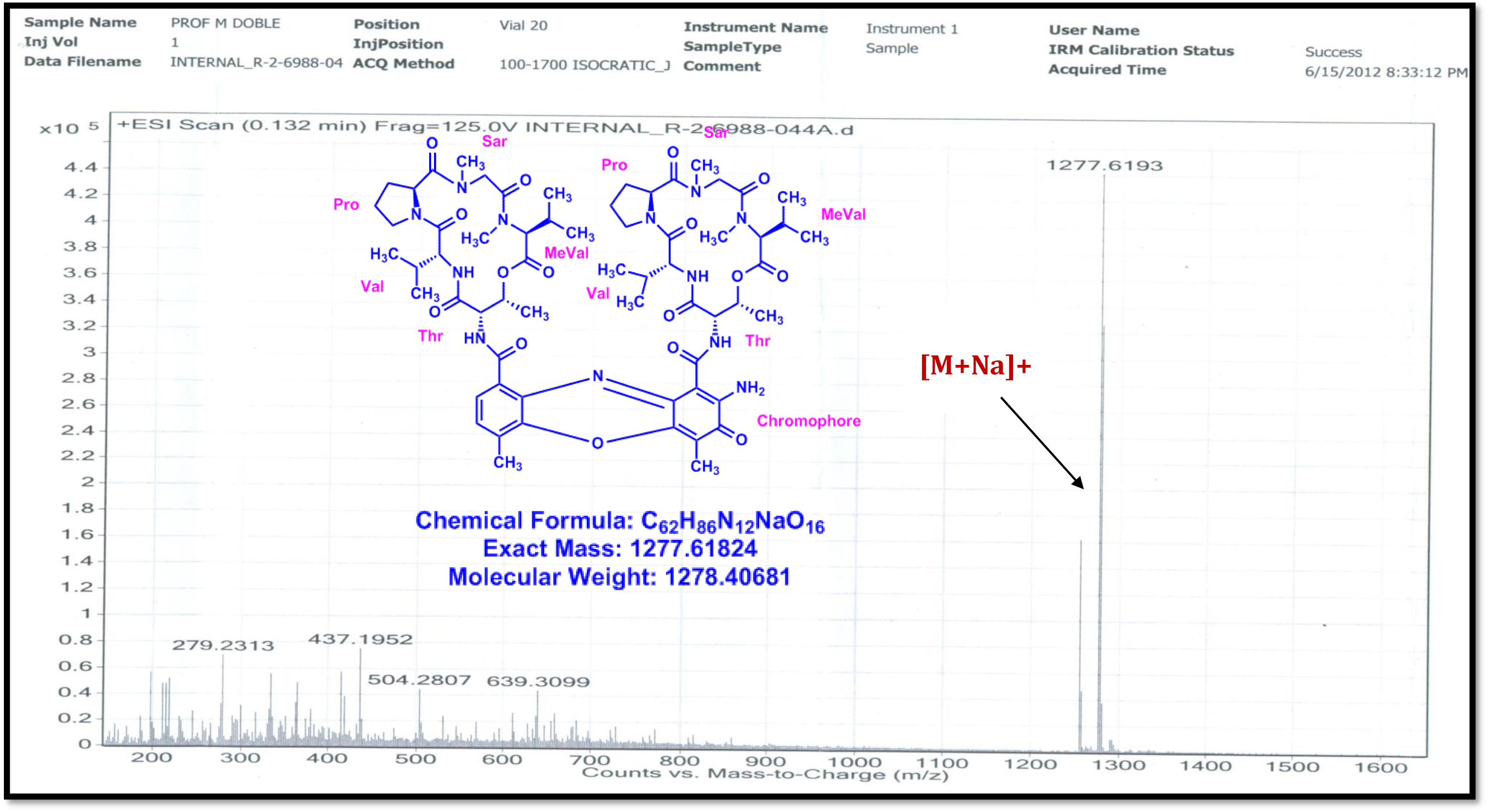
LC-ESI-MS Spectrum of R2 (Positive mode)

**Figure S103.**
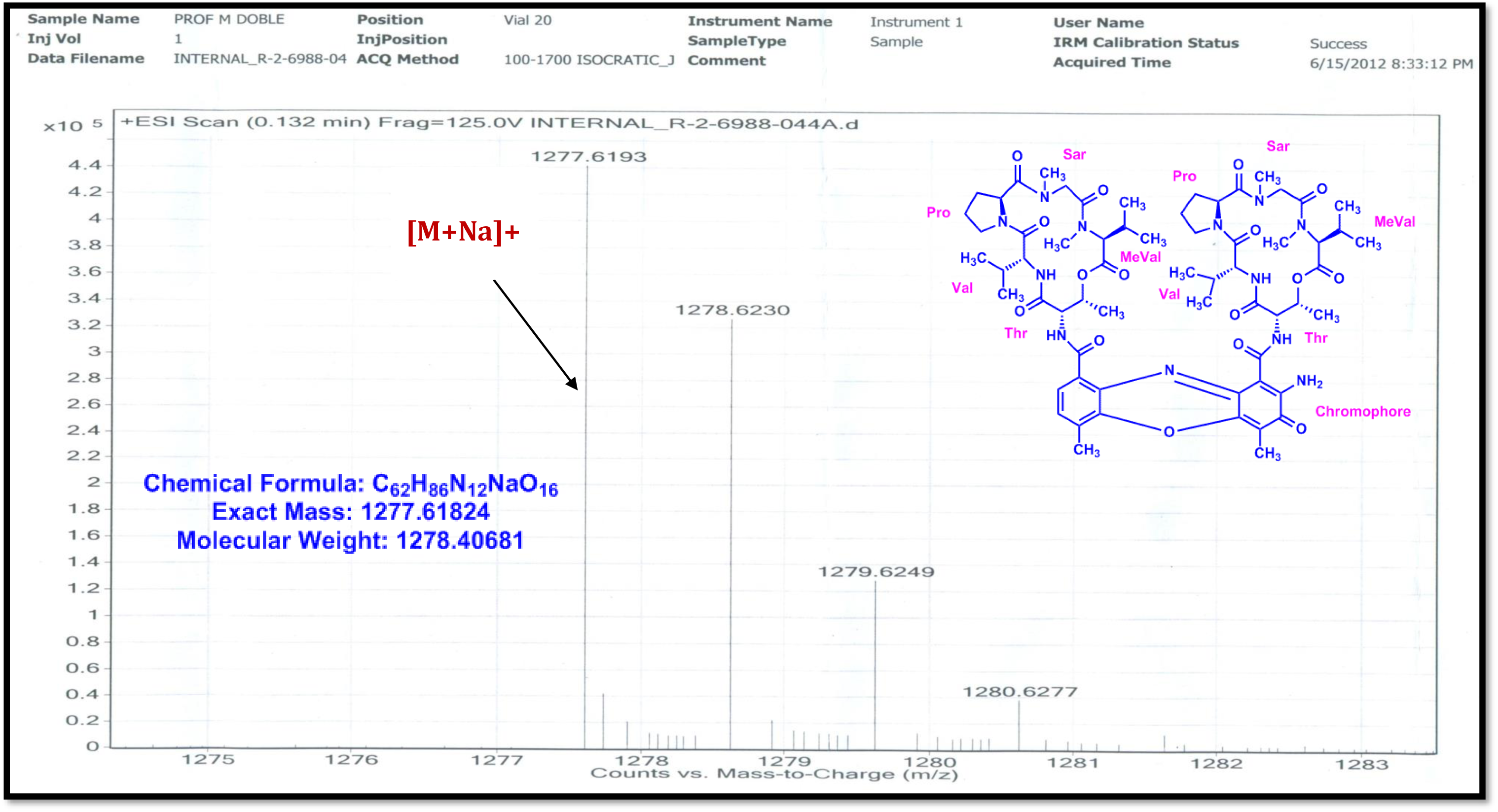
LC-ESI-MS Spectrum of R2 (Molecular ion) (Positive mode)

**Figure S104.**
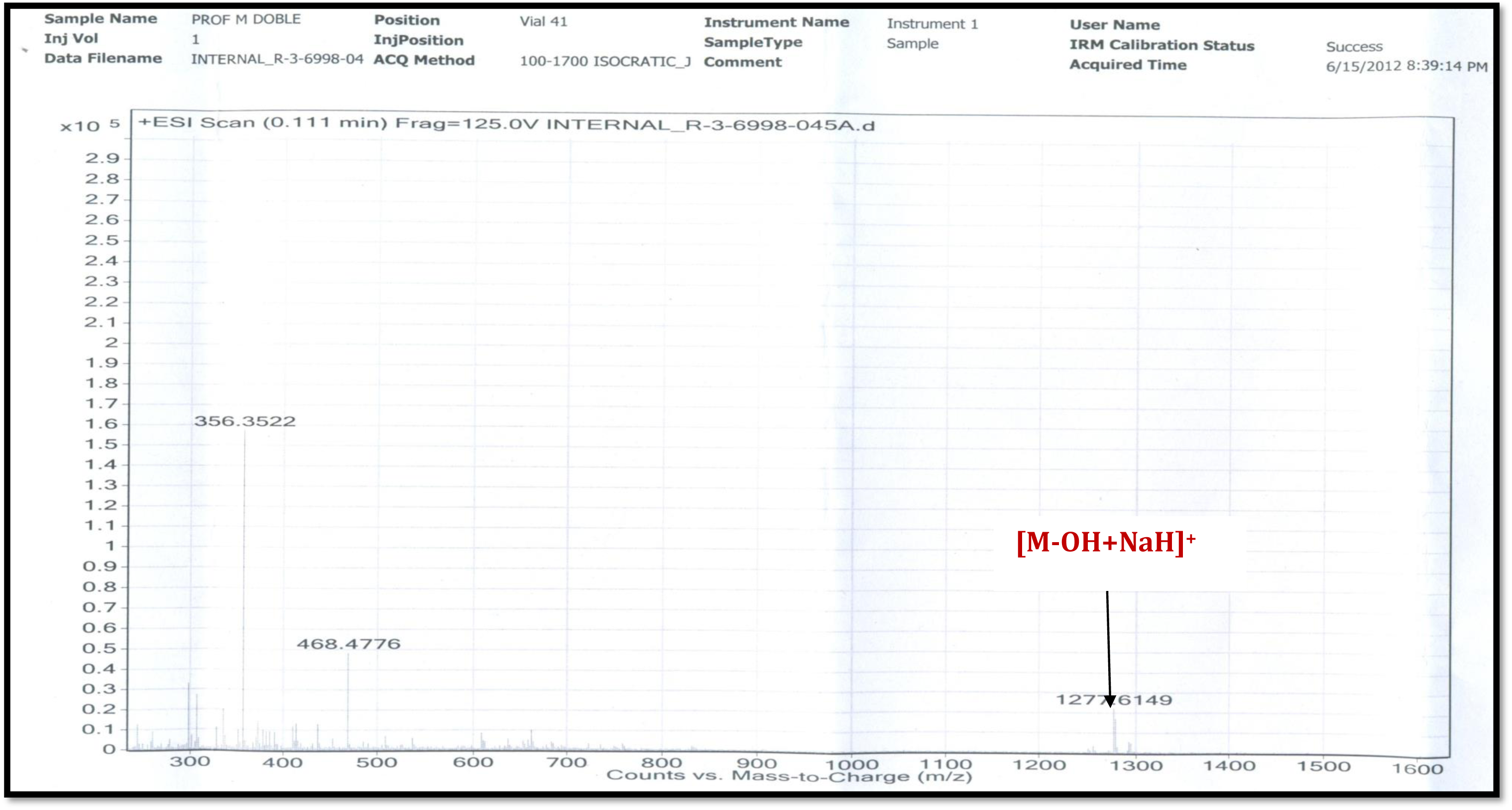
LC-ESI-MS Spectrum of R3 (Positive mode)

**Figure S105.**
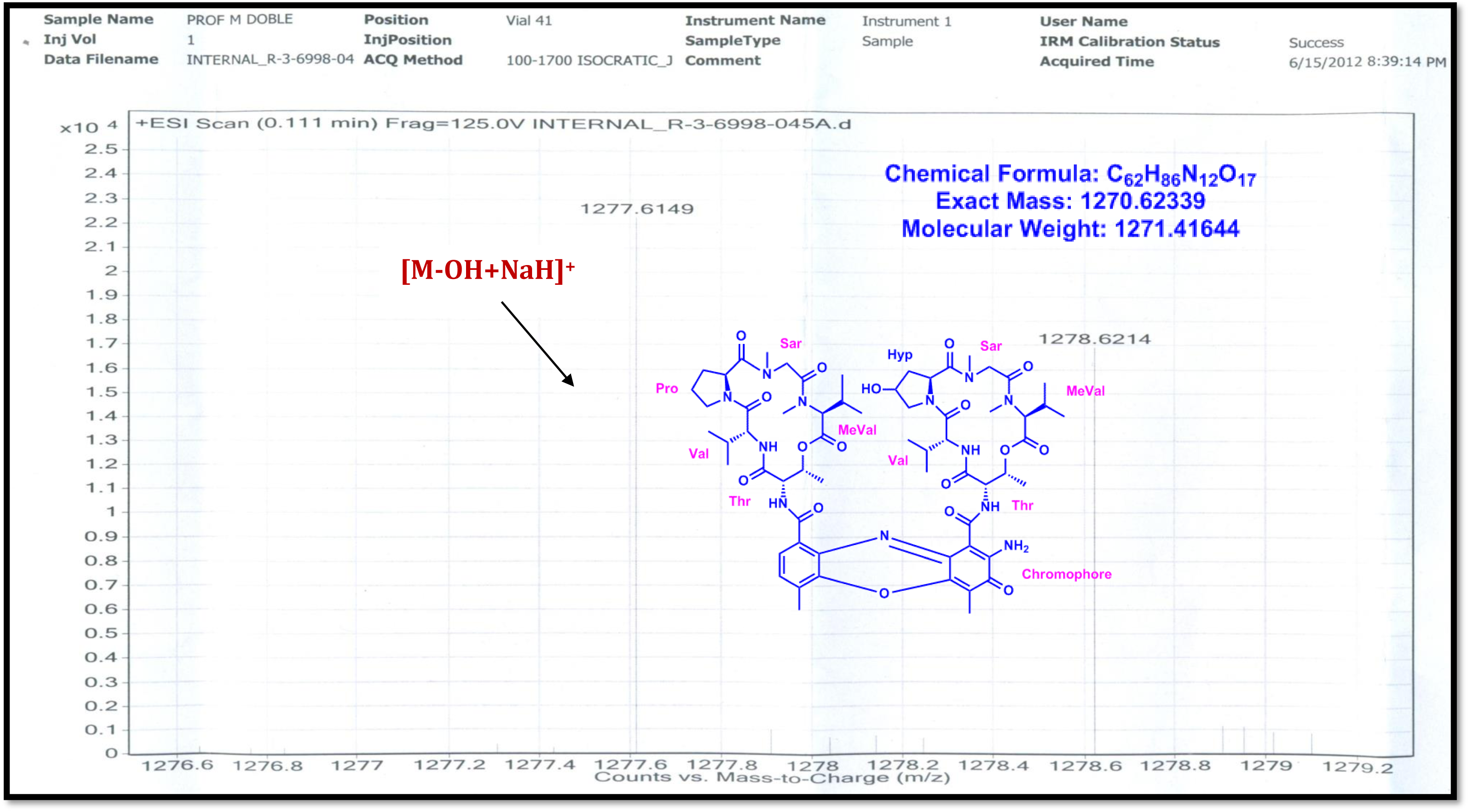
LC-ESI-MS Spectrum of R3 (Positive mode)

**Figure S106.**
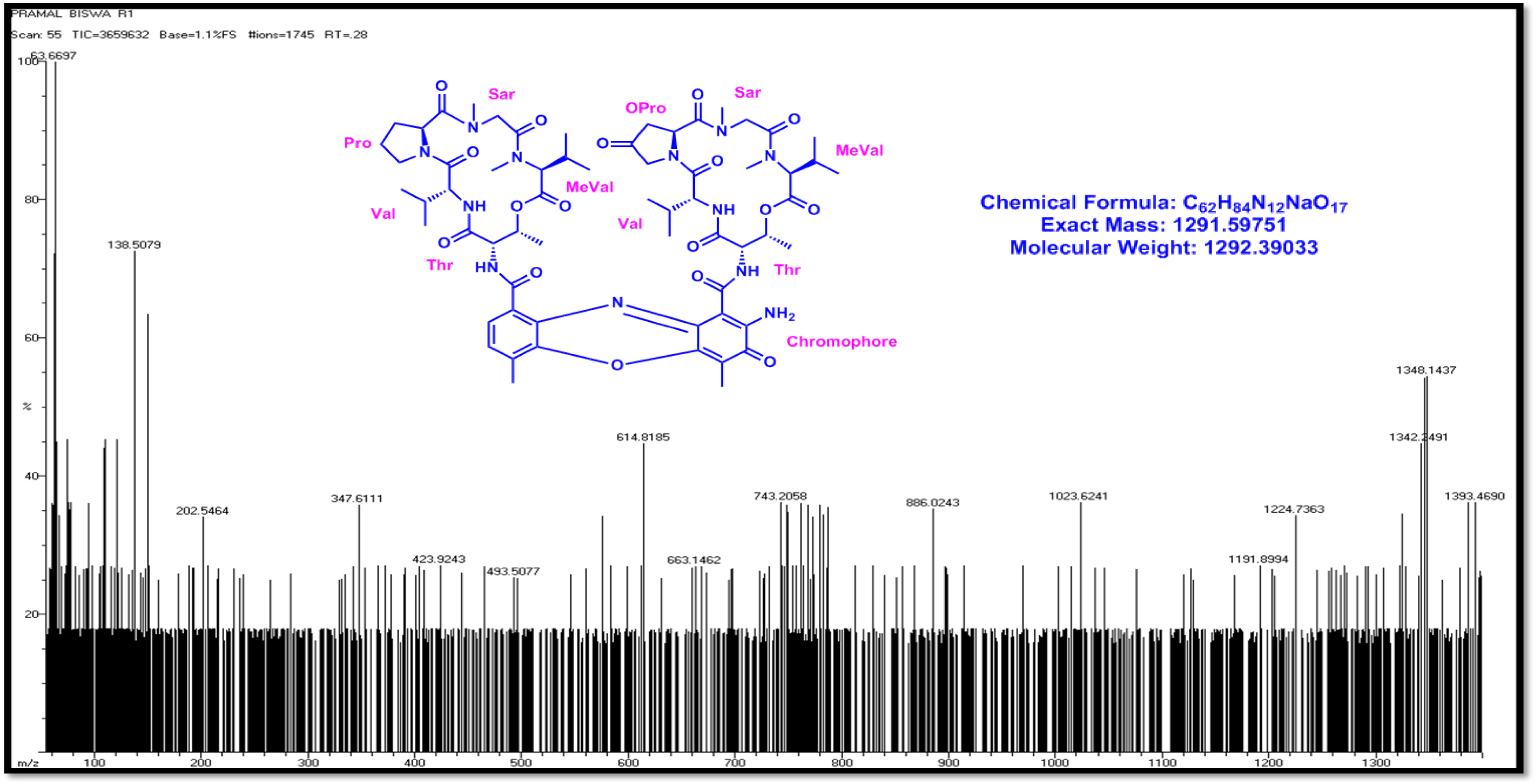
EI-MS Spectrum of Transitmycin (R1) (Positive mode)

**Figure S107.**
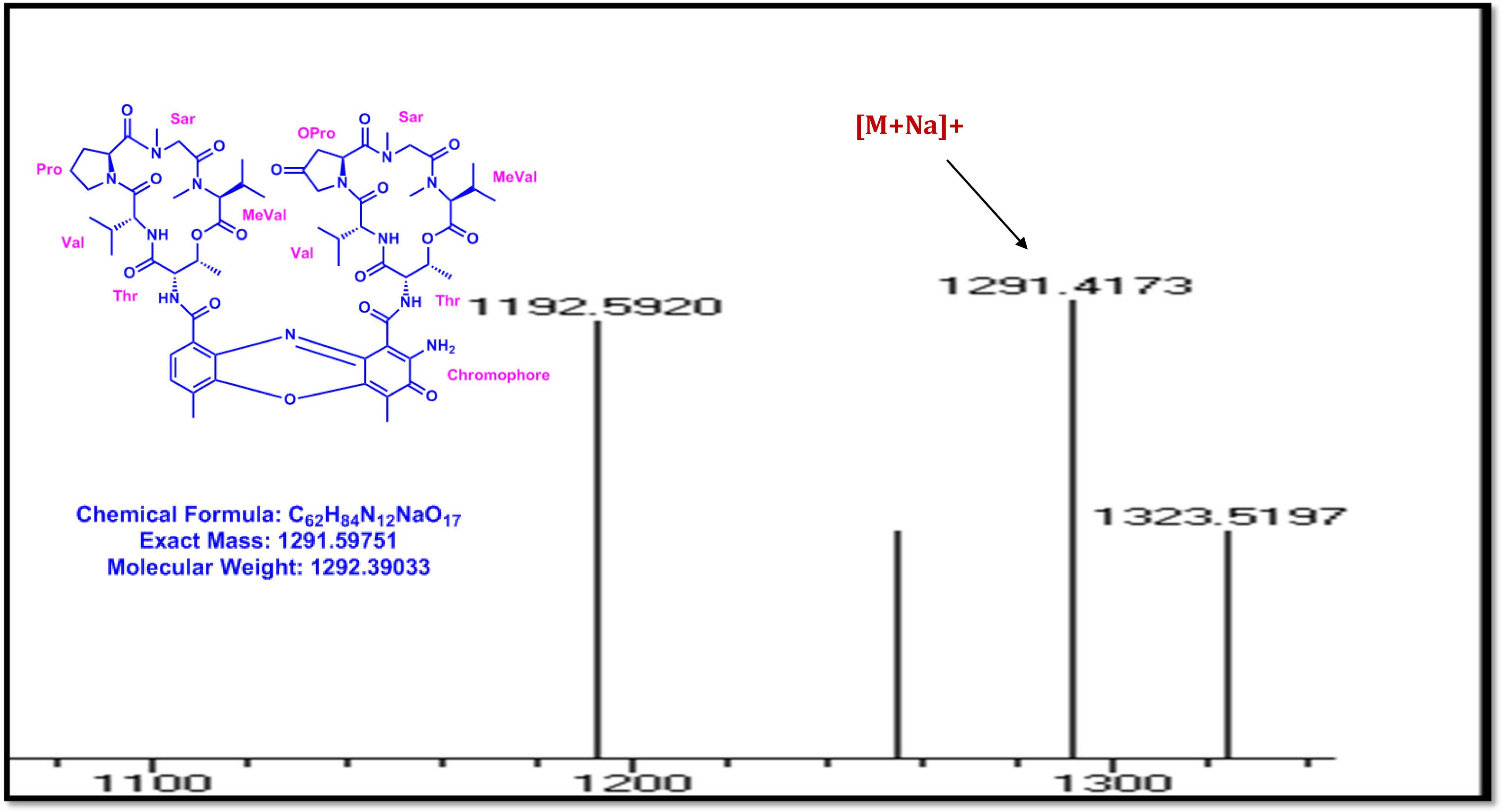
EI-MS Spectrum of (R1) (molecular ion peak) (Positive mode)

**Figure S108.**
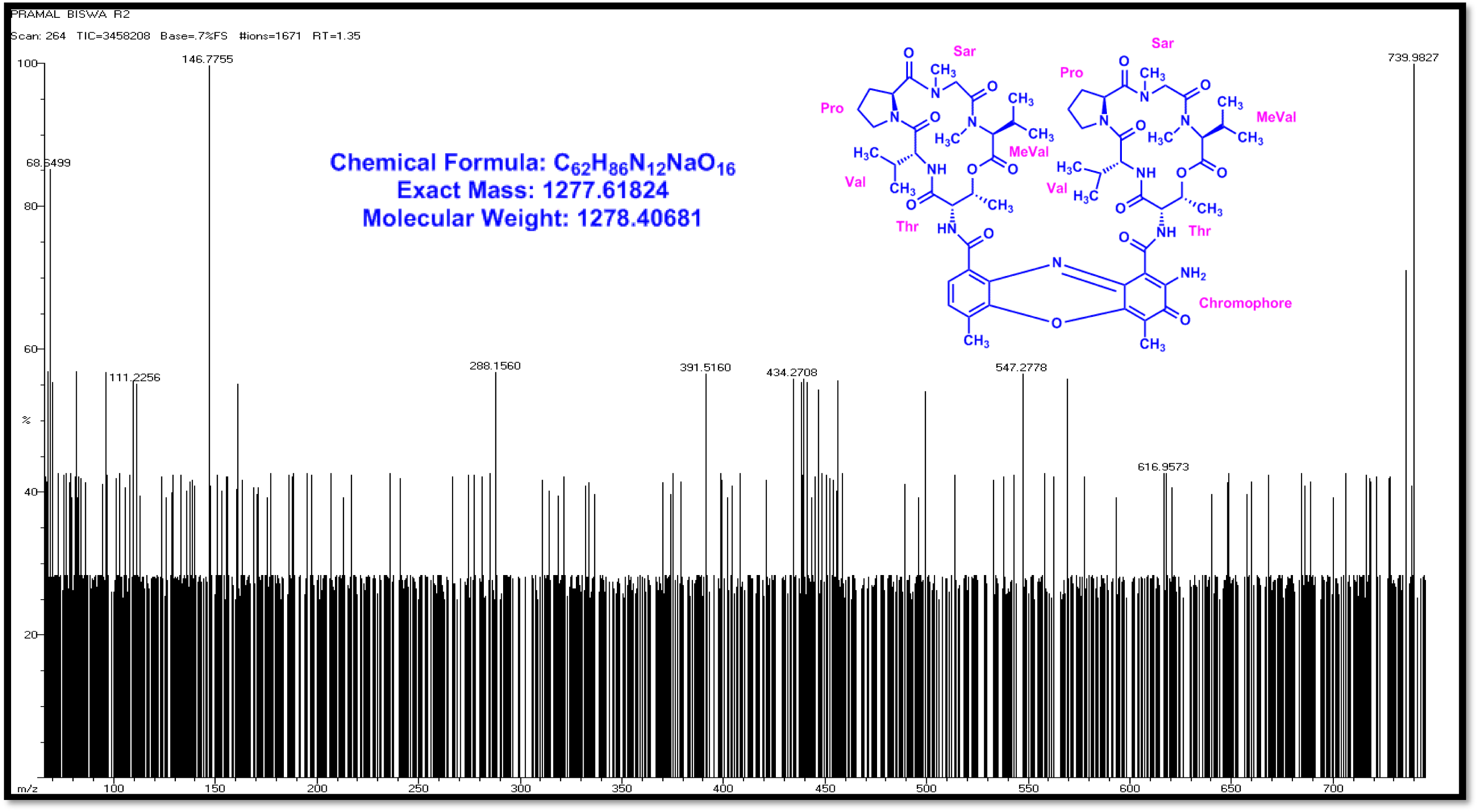
EI-MS Spectrum of (R2) (Positive mode)

**Figure S109.**
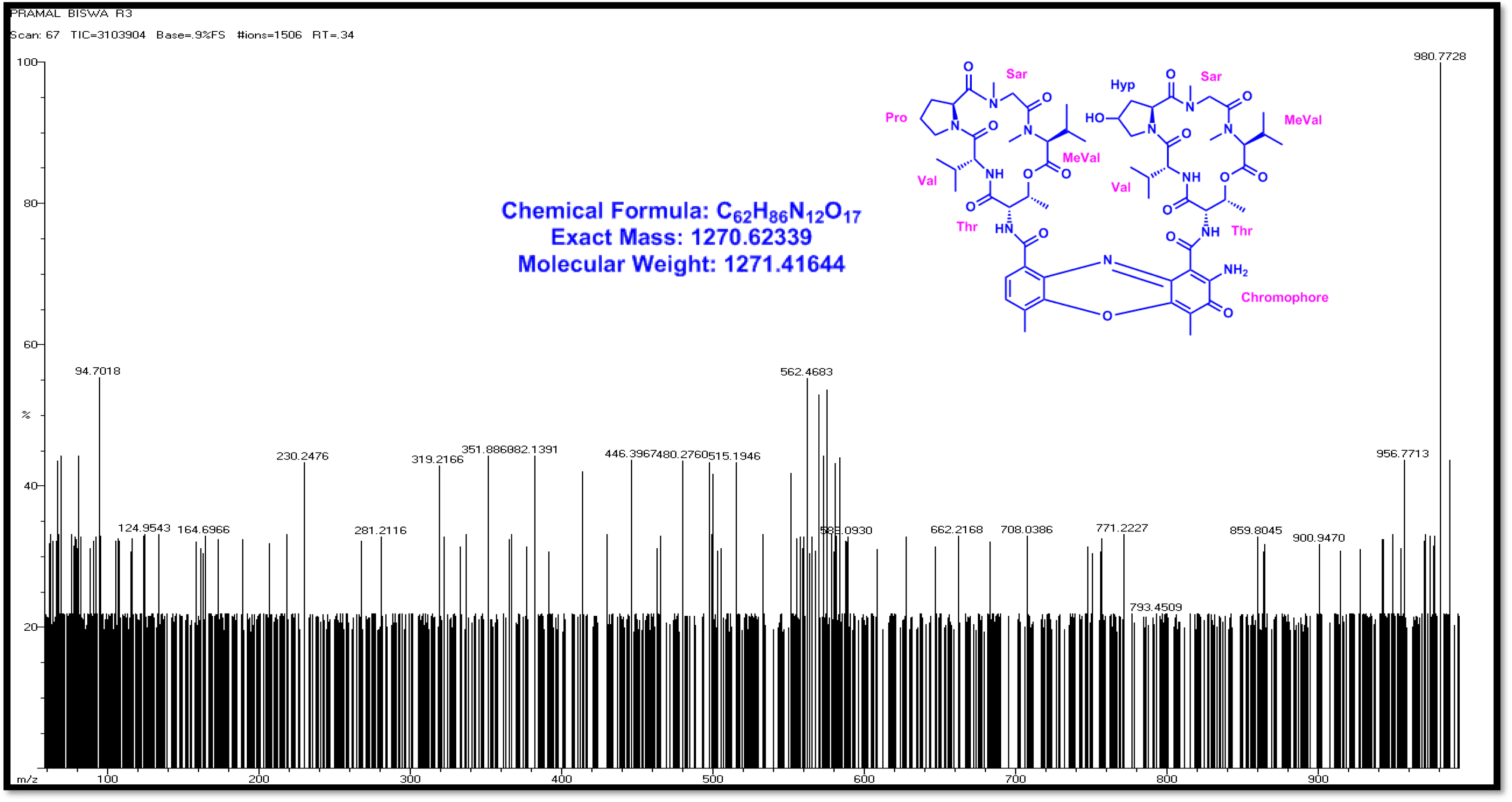
EI-MS Spectrum of (R3) (Positive mode)

**Figure S110.**
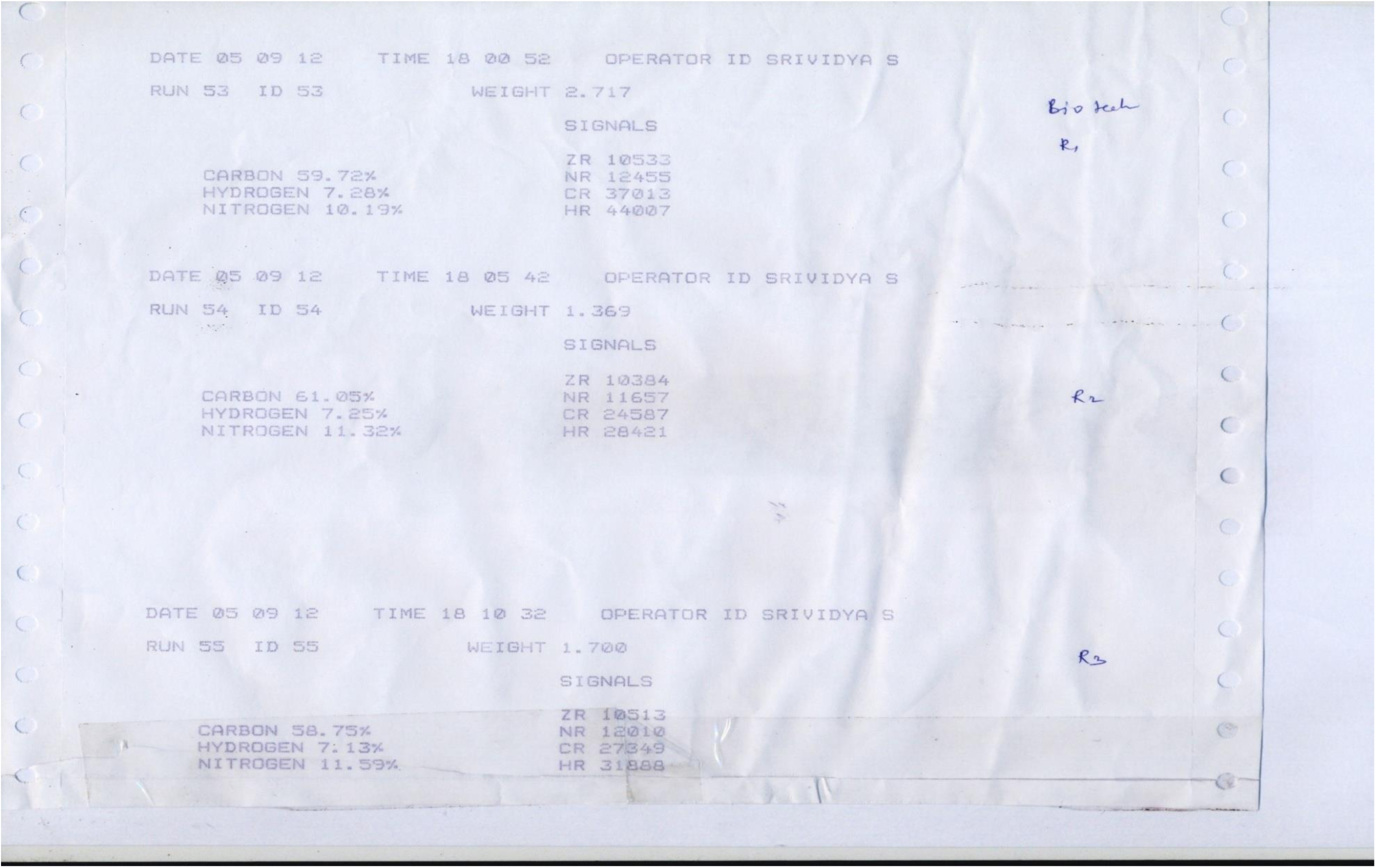
CHN analysis data of Transitmycin (R1), R2, R3.

**Figure S111.**
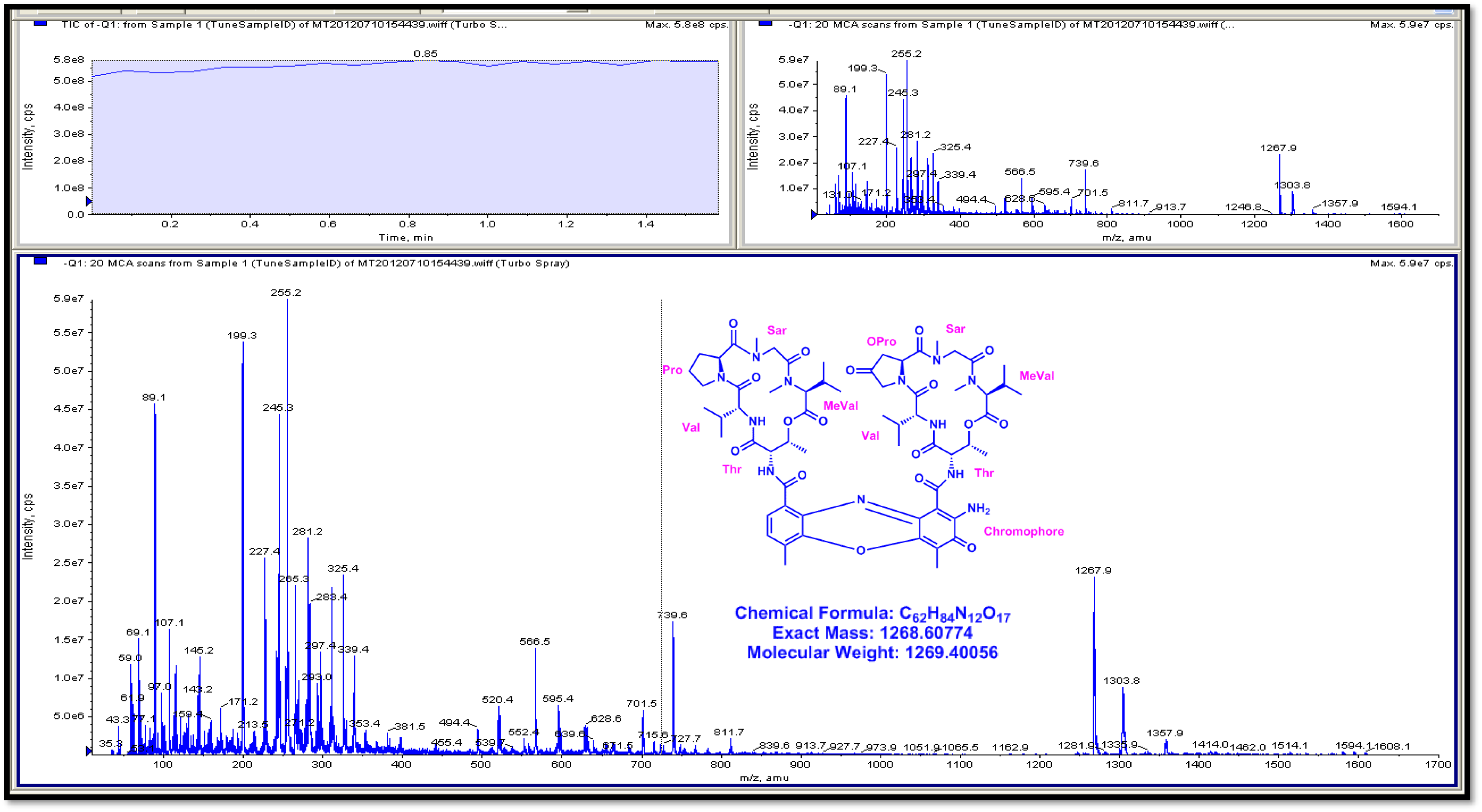
3200 QTRAP LC/MS/MS Spectrum of Transitmycin (R1) (Negative mode)

**Figure S112.**
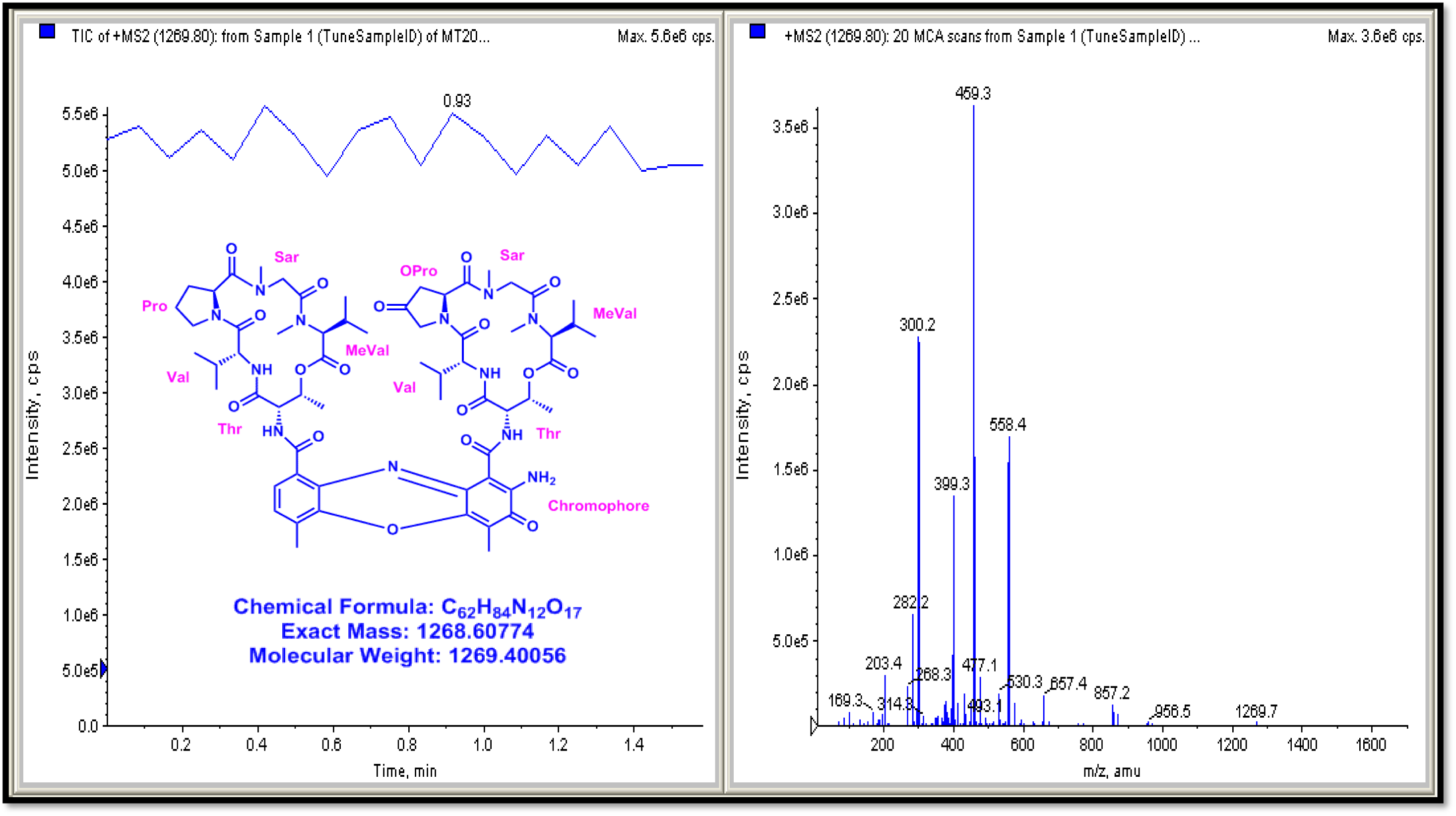
3200 QTRAP LC/MS/MS Spectrum of Transitmycin (R1) (positive mode)

**Figure S113.**
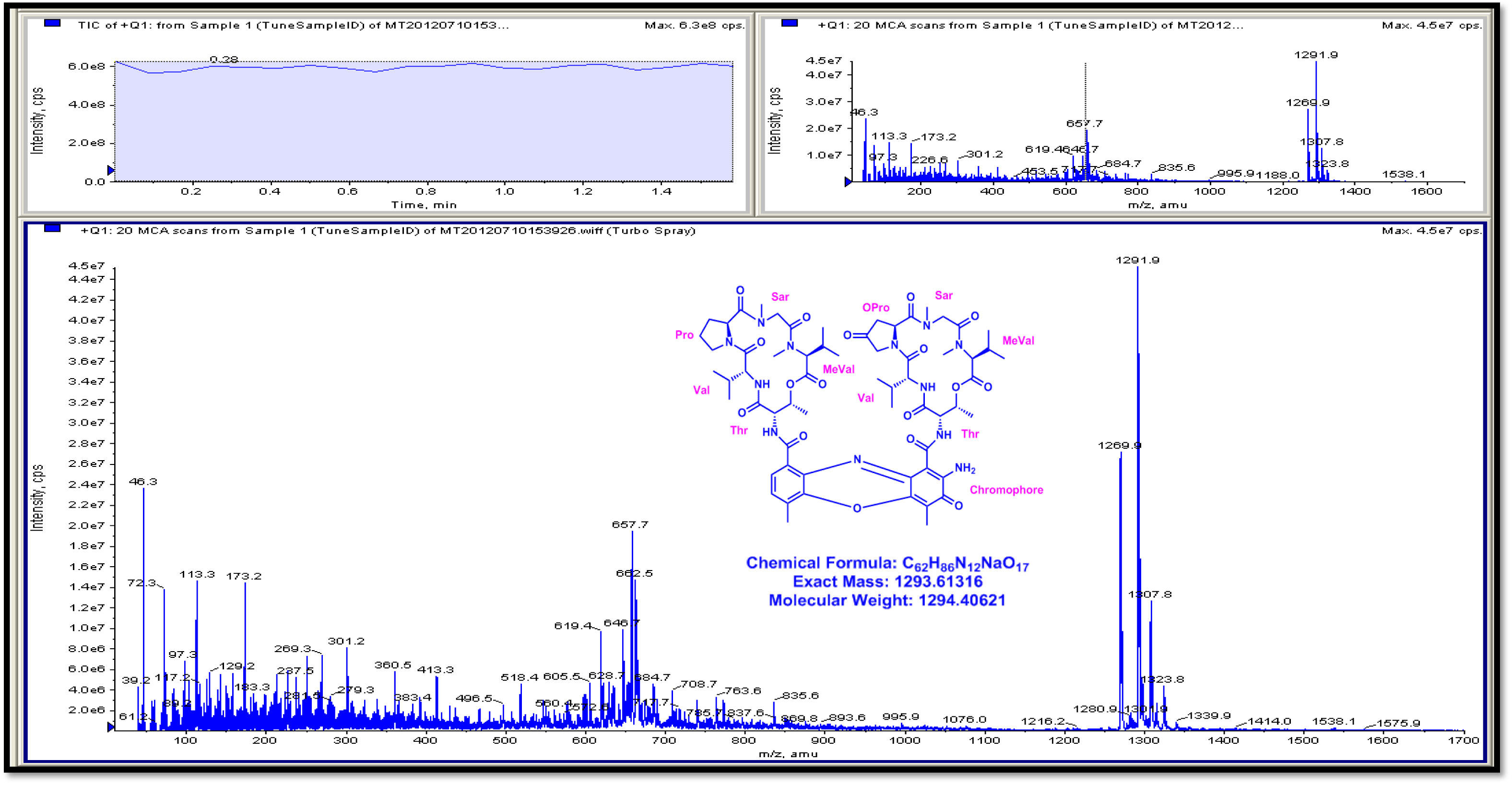
3200 QTRAP LC/MS/MS Spectrum of Transitmycin (R1) (positive mode mode)

**Figure S114.**
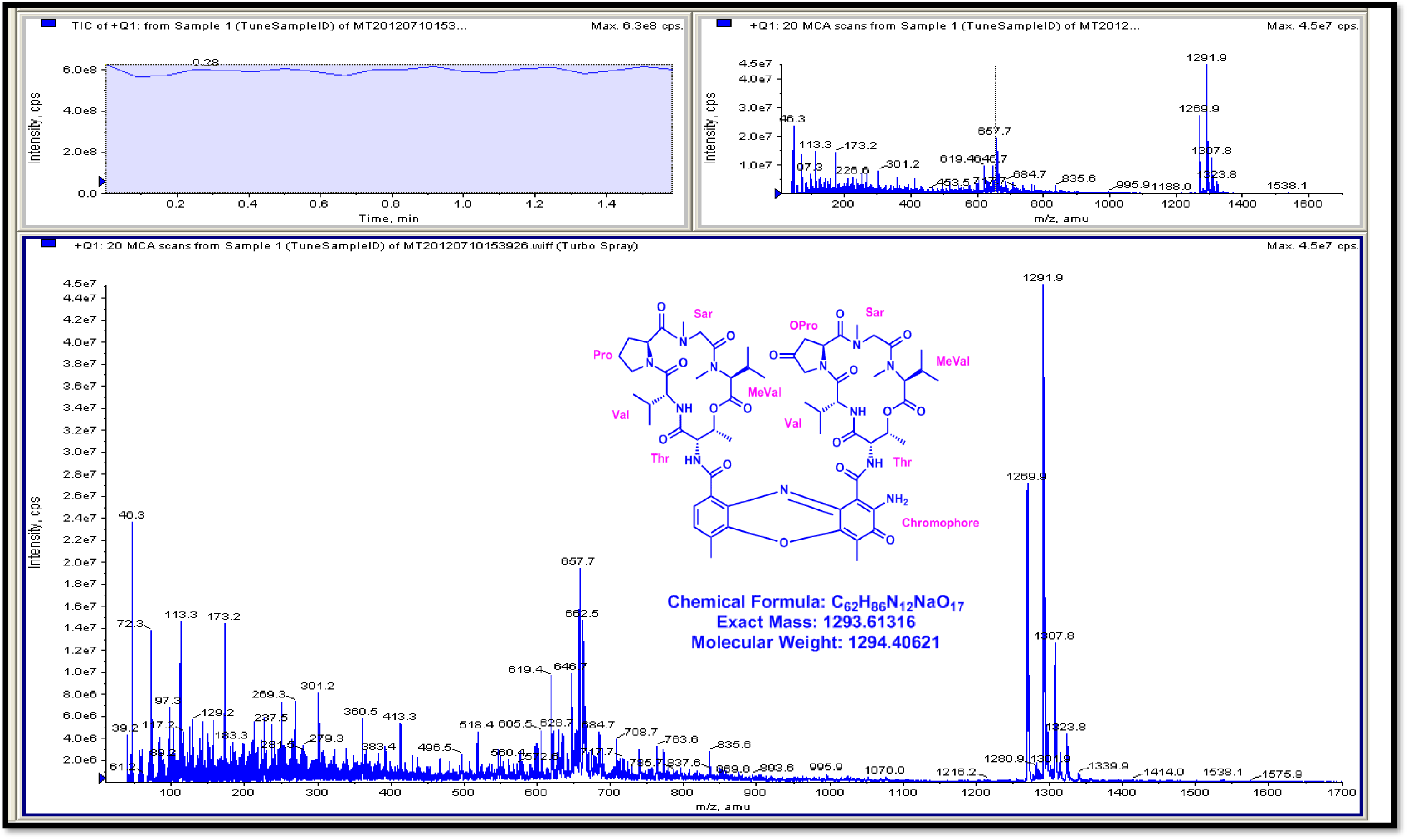
3200 QTRAP LC/MS/MS Spectrum of Transitmycin (R1) (positive mode)

**Figure S115.**
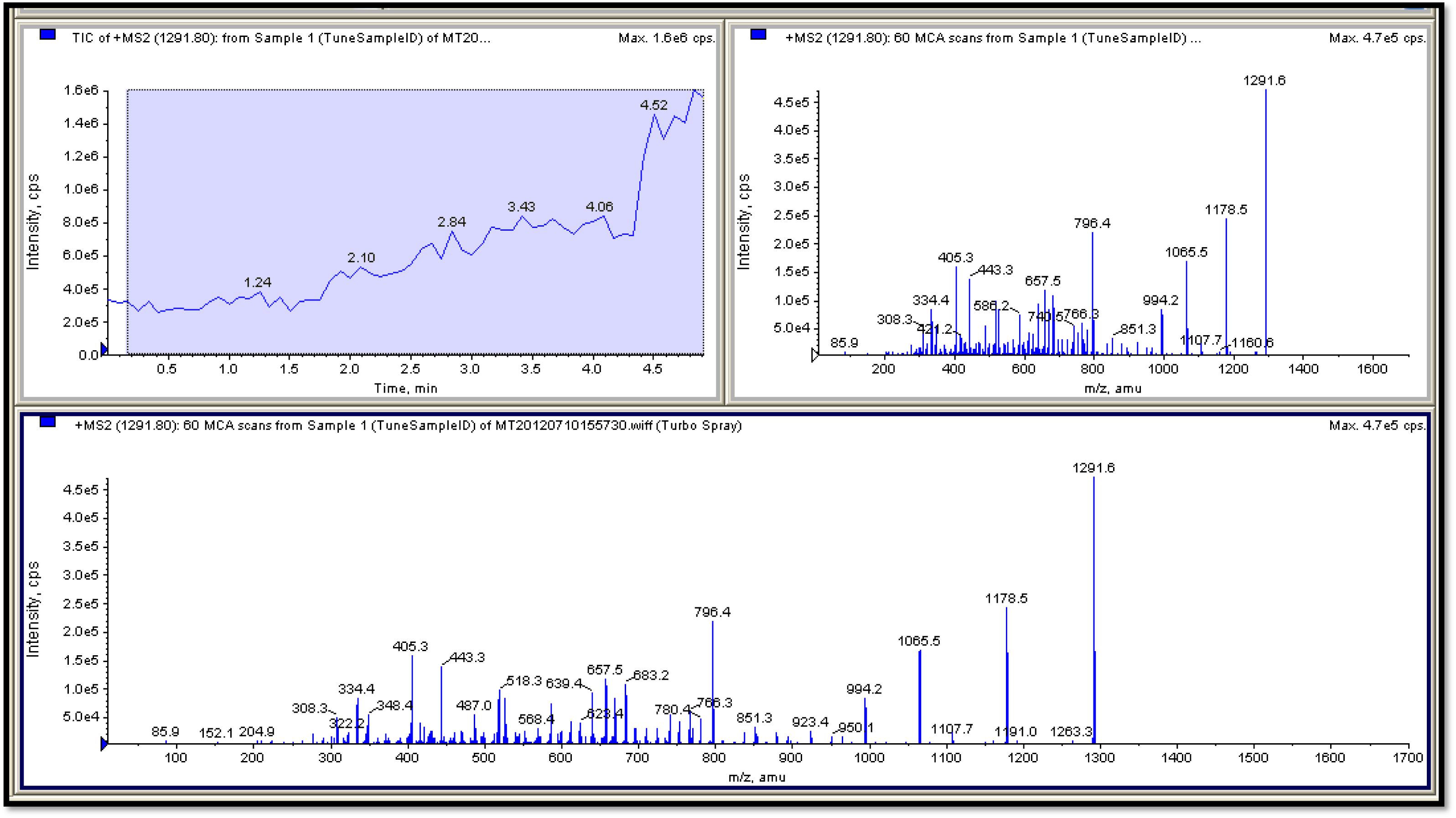
3200 QTRAP LC/MS/MS Spectrum of Transitmycin (R1) (positive mode)

**Figure S115.**
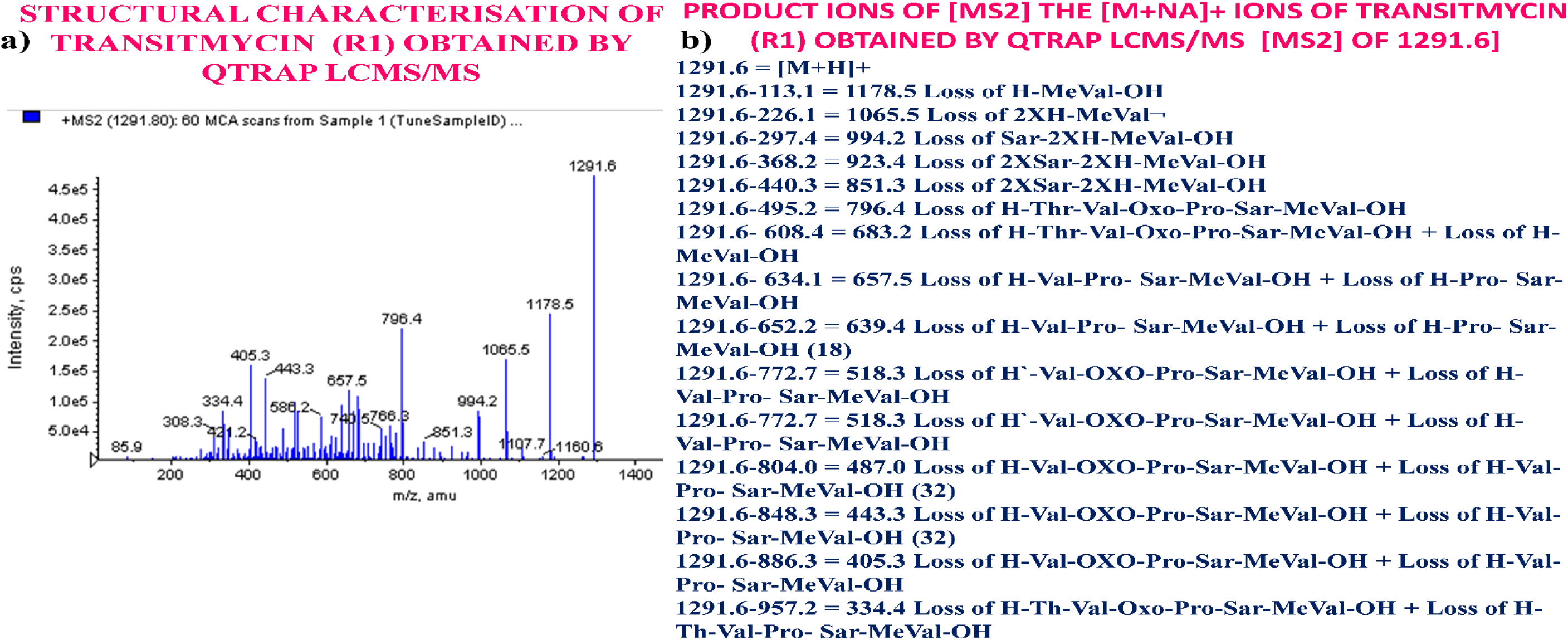
**a-b** Structural characterisation of Transitmycin (R1) obtained by QTRAP LC-MS/MS. **b** Product ions [MS2] the [M+Na]+ ions Transitmycin (R1) obtained by QTRAP LCMS/MS [MS2] of 1291.6, 1291.6 [M+H]^+^, 1178.5 Loss of H-MeVal-OH, 1065.5 Loss of 2XH- MeVal, 994.2 Loss of Sar-2XH-MeVal-OH, 923.4 Loss of 2XSar-2XH-MeVal-OH, 851.3 Loss of 2XSar-2XH-MeVal-OH, 796.4 Loss of H- Thr-Val-Oxo-Pro-Sar-MeVal-OH, 683.2 Loss of H-Thr-Val-Oxo-Pro-Sar-MeVal-OH + Loss of H-MeVal-OH, 657.5 Loss of H-Val-Pro- Sar- MeVal-OH + Loss of H-Pro- Sar-MeVal-OH, 639.4 Loss of H-Val-Pro- Sar-MeVal-OH + Loss of H-Pro- Sar-MeVal-OH (18), 518.3 Loss of H’-Val-OXO-Pro-Sar-MeVal-OH + Loss of H-Val-Pro- Sar-MeVal-OH, 518.3 Loss of H’-Val-OXO-Pro-Sar-MeVal-OH + Loss of H-Val-Pro- Sar-MeVal-OH, 487.0 Loss of H-Val-OXO-Pro-Sar-MeVal-OH + Loss of H-Val-Pro- Sar-MeVal-OH (32), 443.3 Loss of H-Val-OXO-Pro-Sar- MeVal-OH + Loss of H-Val-Pro- Sar-MeVal-OH (32), 405.3 Loss of H-Val-OXO-Pro-Sar-MeVal-OH + Loss of H-Val-Pro- Sar-MeVal-OH, 334.4 Loss of H-Th-Val-Oxo-Pro-Sar-MeVal-OH + Loss of H-Th-Val-Pro- Sar-MeVal-OH.

**Table S7a.**
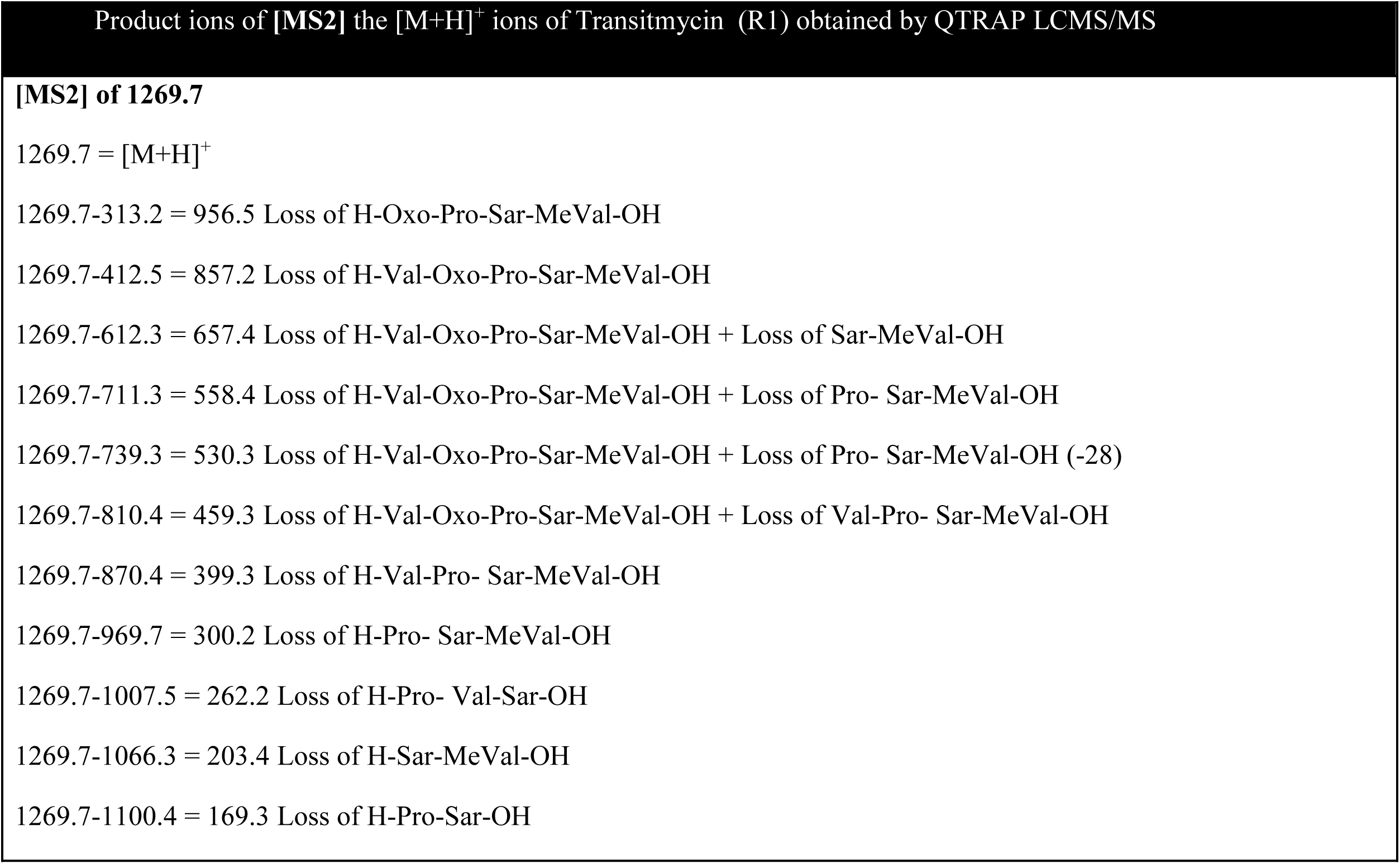
Structural characterisation of Transitmycin (R1) obtained by QTRAP LCMS/MS.

| Product ions of [MS2] the [M+H]<sup>+</sup> ions of Transitmycin (R1) obtained by QTRAP LCMS/MS |
| --- |
| **[MS2] of 1269.7** |
| 1269.7 = [M+H]<sup>+</sup> |
| 1269.7-313.2 = 956.5 Loss of H-Oxo-Pro-Sar-MeVal-OH |
| 1269.7-412.5 = 857.2 Loss of H-Val-Oxo-Pro-Sar-MeVal-OH |
| 1269.7-612.3 = 657.4 Loss of H-Val-Oxo-Pro-Sar-MeVal-OH + Loss of Sar-MeVal-OH |
| 1269.7-711.3 = 558.4 Loss of H-Val-Oxo-Pro-Sar-MeVal-OH + Loss of Pro- Sar-MeVal-OH |
| 1269.7-739.3 = 530.3 Loss of H-Val-Oxo-Pro-Sar-MeVal-OH + Loss of Pro- Sar-MeVal-OH (-28) |
| 1269.7-810.4 = 459.3 Loss of H-Val-Oxo-Pro-Sar-MeVal-OH + Loss of Val-Pro- Sar-MeVal-OH |
| 1269.7-870.4 = 399.3 Loss of H-Val-Pro- Sar-MeVal-OH |
| 1269.7-969.7 = 300.2 Loss of H-Pro- Sar-MeVal-OH |
| 1269.7-1007.5 = 262.2 Loss of H-Pro- Val-Sar-OH |
| 1269.7-1066.3 = 203.4 Loss of H-Sar-MeVal-OH |
| 1269.7-1100.4 = 169.3 Loss of H-Pro-Sar-OH |

**Table S7b.** Structural characterisation of Transitmycin (R1) obtained by QTRAP LCMS/MS.

| Product ions of [MS2] the [M+Na]<sup>+</sup> ions of Transitmycin (R1) obtained by QTRAP LCMS/MS |  |
| --- | --- |
| **[MS2] of 1291.6** |  |
| 1291.6 = [M+H]<sup>+</sup> |  |
| 1291.6-113.1 = 1178.5 | Loss of H-MeVal-OH |
| 1291.6-226.1 = 1065.5 | Loss of 2XH-MeVal |
| 1291.6-297.4 = 994.2 | Loss of Sar-2XH-MeVal-OH |
| 1291.6-368.2 = 923.4 | Loss of 2XSar-2XH-MeVal-OH |
| 1291.6-440.3 = 851.3 | Loss of 2XSar-2XH-MeVal-OH |
| 1291.6-495.2 = 796.4 | Loss of H-Thr-Val-Oxo-Pro-Sar-MeVal-OH |
| 1291.6- 608.4 = 683.2 | Loss of H-Thr-Val-Oxo-Pro-Sar-MeVal-OH + Loss of H-MeVal-OH |
| 1291.6- 634.1 = 657.5 | Loss of H-Val-Pro- Sar-MeVal-OH + Loss of H-Pro- Sar-MeVal-OH |
| 1291.6-652.2 = 639.4 | Loss of H-Val-Pro- Sar-MeVal-OH + Loss of H-Pro- Sar-MeVal-OH (18) |
| 1291.6-772.7 = 518.3 | Loss of H<sup>-</sup>-Val-OXO-Pro-Sar-MeVal-OH + Loss of H-Val-Pro- Sar-MeVal-OH |
| 1291.6-772.7 = 518.3 | Loss of H<sup>-</sup>-Val-OXO-Pro-Sar-MeVal-OH + Loss of H-Val-Pro- Sar-MeVal-OH |
| 1291.6-804.0 = 487.0 | Loss of H-Val-OXO-Pro-Sar-MeVal-OH + Loss of H-Val-Pro- Sar-MeVal-OH (32) |
| 1291.6-848.3 = 443.3 | Loss of H-Val-OXO-Pro-Sar-MeVal-OH + Loss of H-Val-Pro- Sar-MeVal-OH (32) |
| 1291.6-886.3 = 405.3 | Loss of H-Val-OXO-Pro-Sar-MeVal-OH + Loss of H-Val-Pro- Sar-MeVal-OH |
| 1291.6-957.2 = 334.4 | Loss of H-Th-Val-Oxo-Pro-Sar-MeVal-OH + Loss of H-Th-Val-Pro- Sar-MeVal-OH |

**Figure S116.**
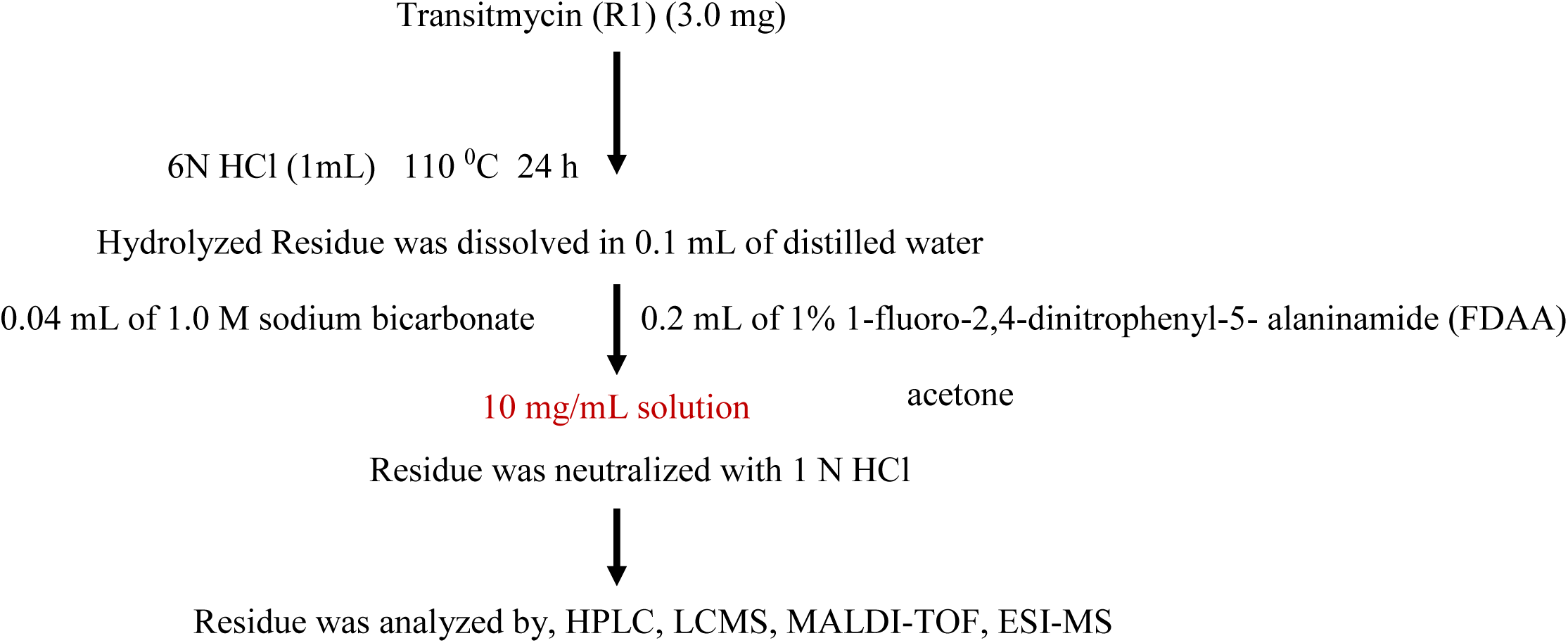
Hydrolysis of Amino Acid Residue of Transitmycin (R1), R2, R3. The Transitmycin A (3.0 mg) dissolved in 6N HCl (1 mL) and heated at 110 ^0^C for 24 h. The solvent was removed under reduced pressure, and the resulting material was subjected to further derivatization. The hydrolysate mixture (3.5 mg) or the amino acid standards (0.5 mg) were dissolved in 0.1 mL of water and treated with 0.2 mL of 1% 1-fluoro-2,4-dinitrophenyl-5-Lalaninamide (FDAA) in acetone and 0.04 mL of 1.0 M sodium bicarbonate. The vials were heated at 50 ^0^C for 90 min, and the contents after cooling at room temperature were neutralized with 1 N HCl. After degassing, an aliquot of the FDAA derivative was diluted in MeOH and purified by chromatography using a RP C-18 column (250 X 4.6 mm) and a linear gradient of acetonitrile and water containing 0.05% trifluoroacetic acid from 20:80 to 50:50 in 20 min and then isocratic. The flow rate was 1 mL/min, and the absorbance detection was at 340 nm. The chromatogram was compared with those of amino acid standards treated in the same conditions. The above same procedure for other two compounds B and C were done.

**Figure S117a.**
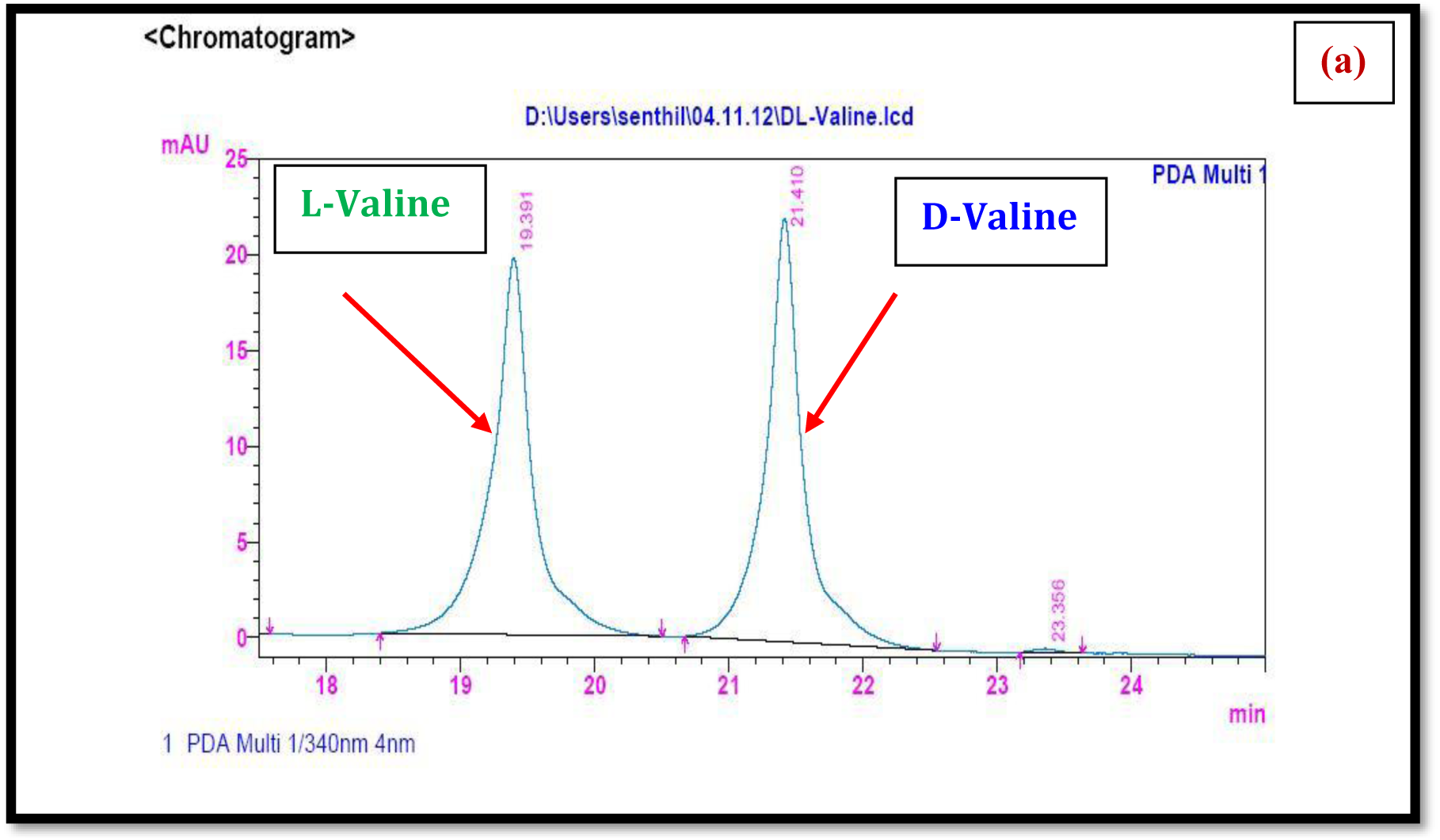
HPLC analysis of standard L-FDAA-D/L-Valine.

**Figure S117 b.**
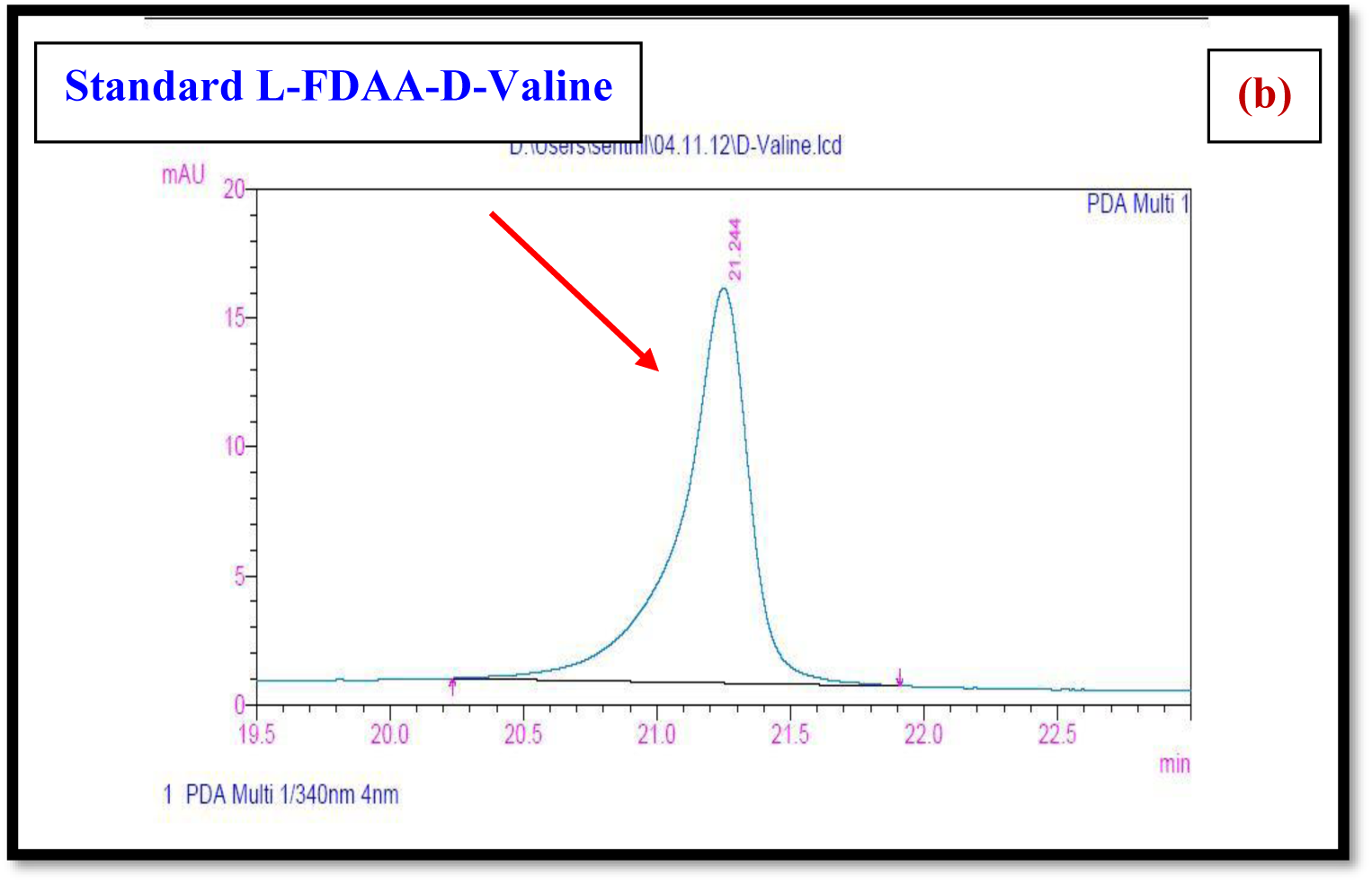
HPLC analysis of standard L-FDAA-D-Valine.

**Figure S117c.**
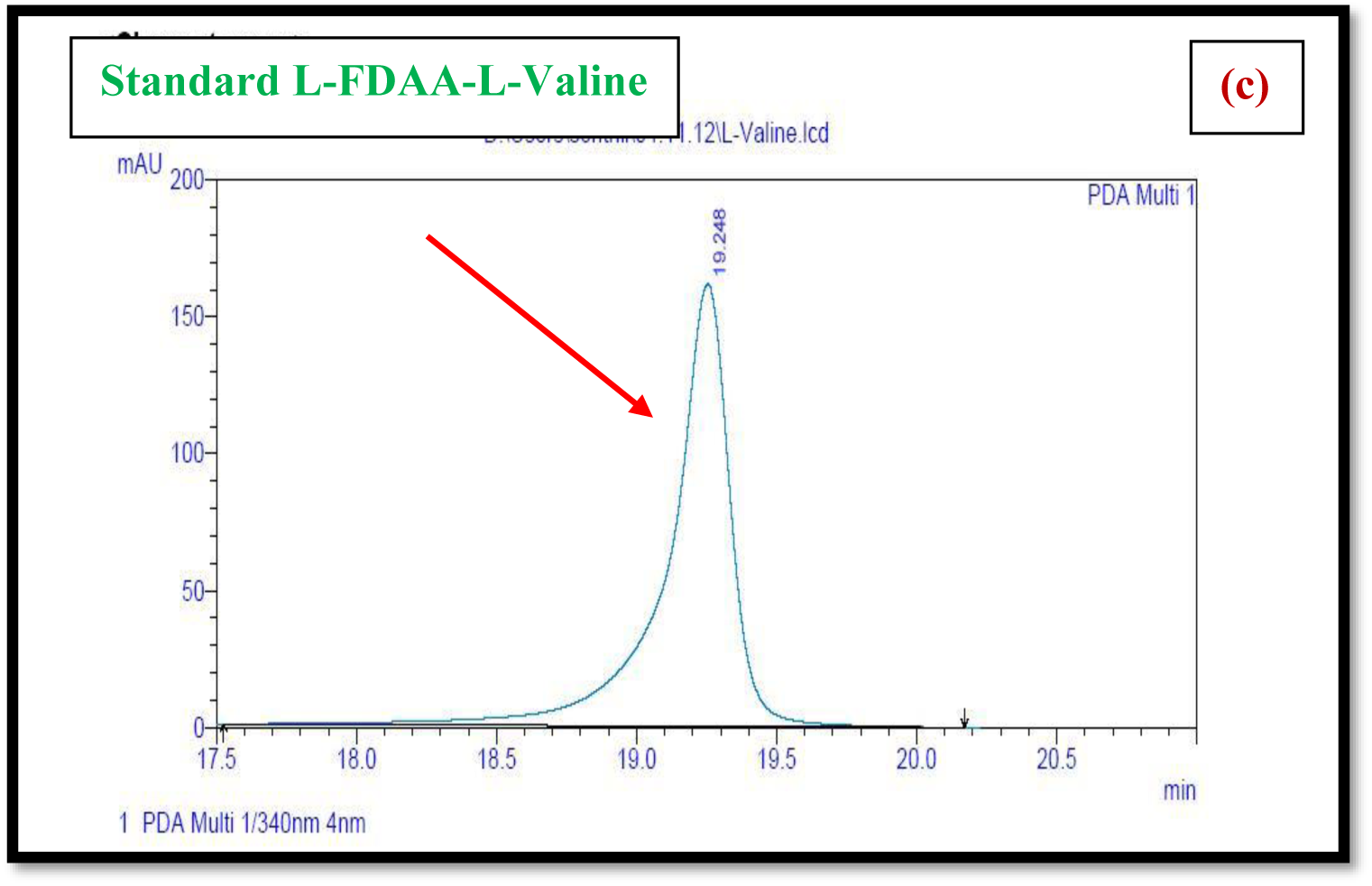
HPLC analysis of standard L-FDAA-L-Valine.

**Figure S118(a).**
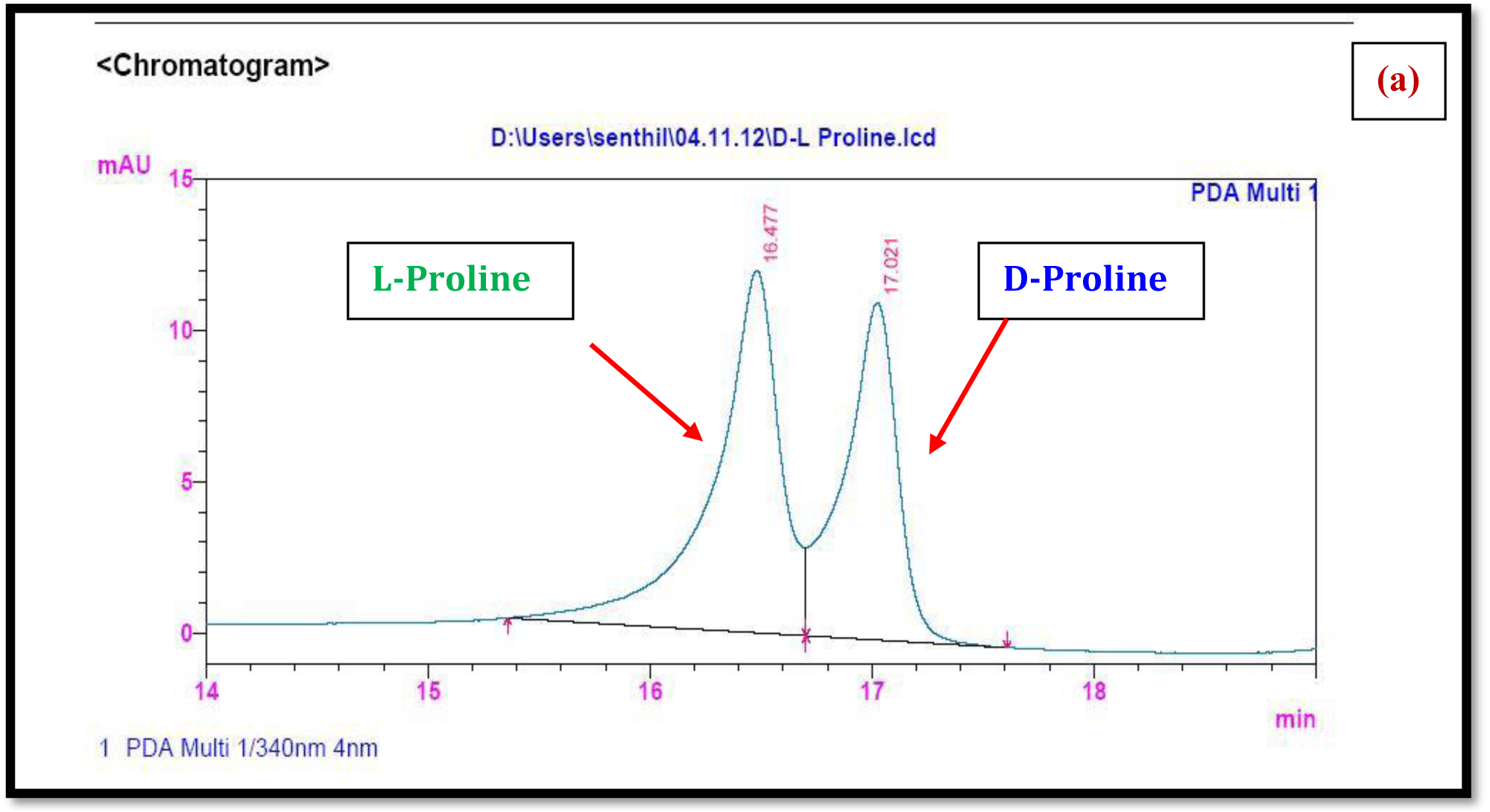
HPLC analysis of standard L-FDAA-D/L-Proline.

**Figure S118 (b).**
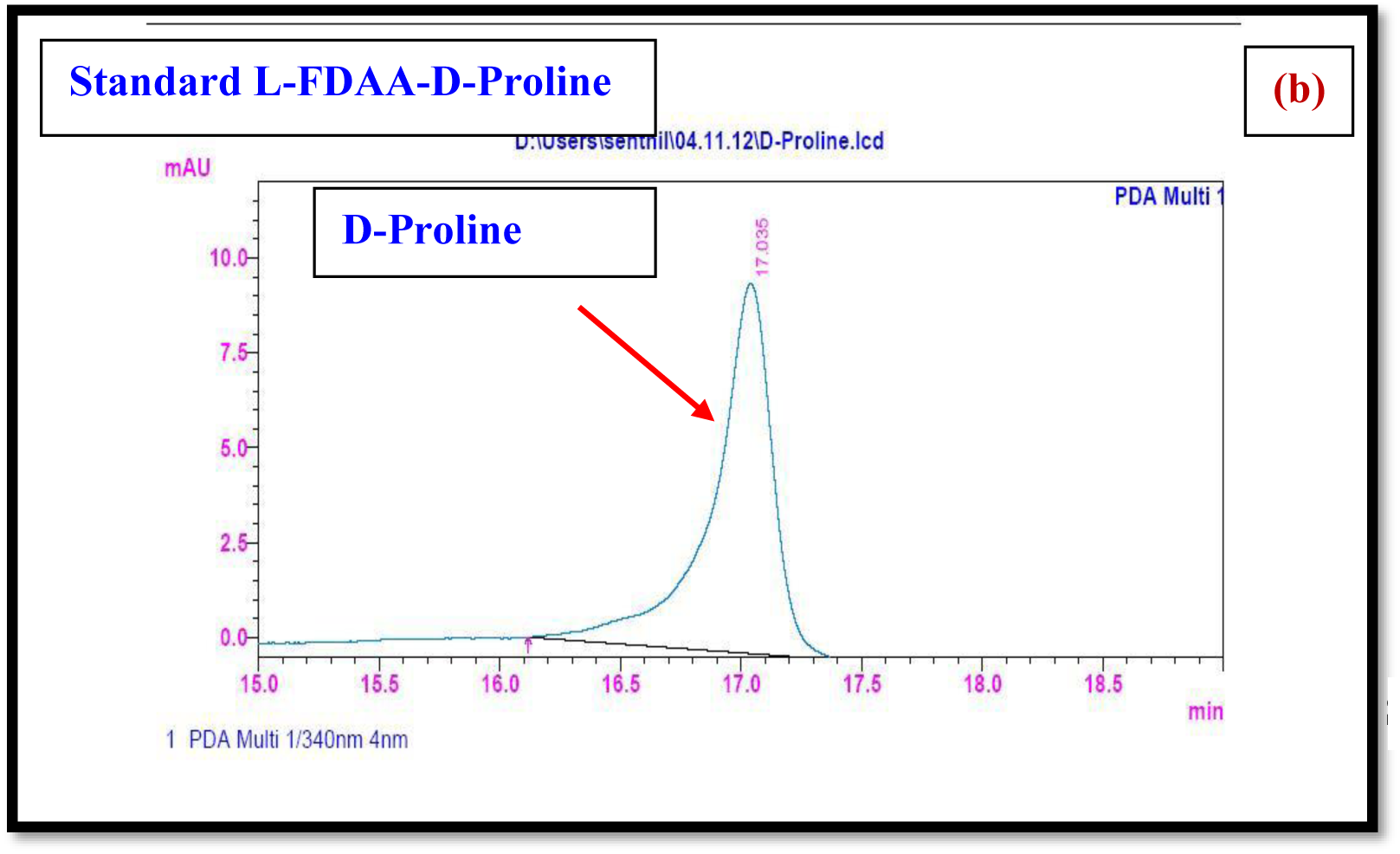
HPLC analysis of standard L-FDAA-D-Proline.

**Figure S118c.**
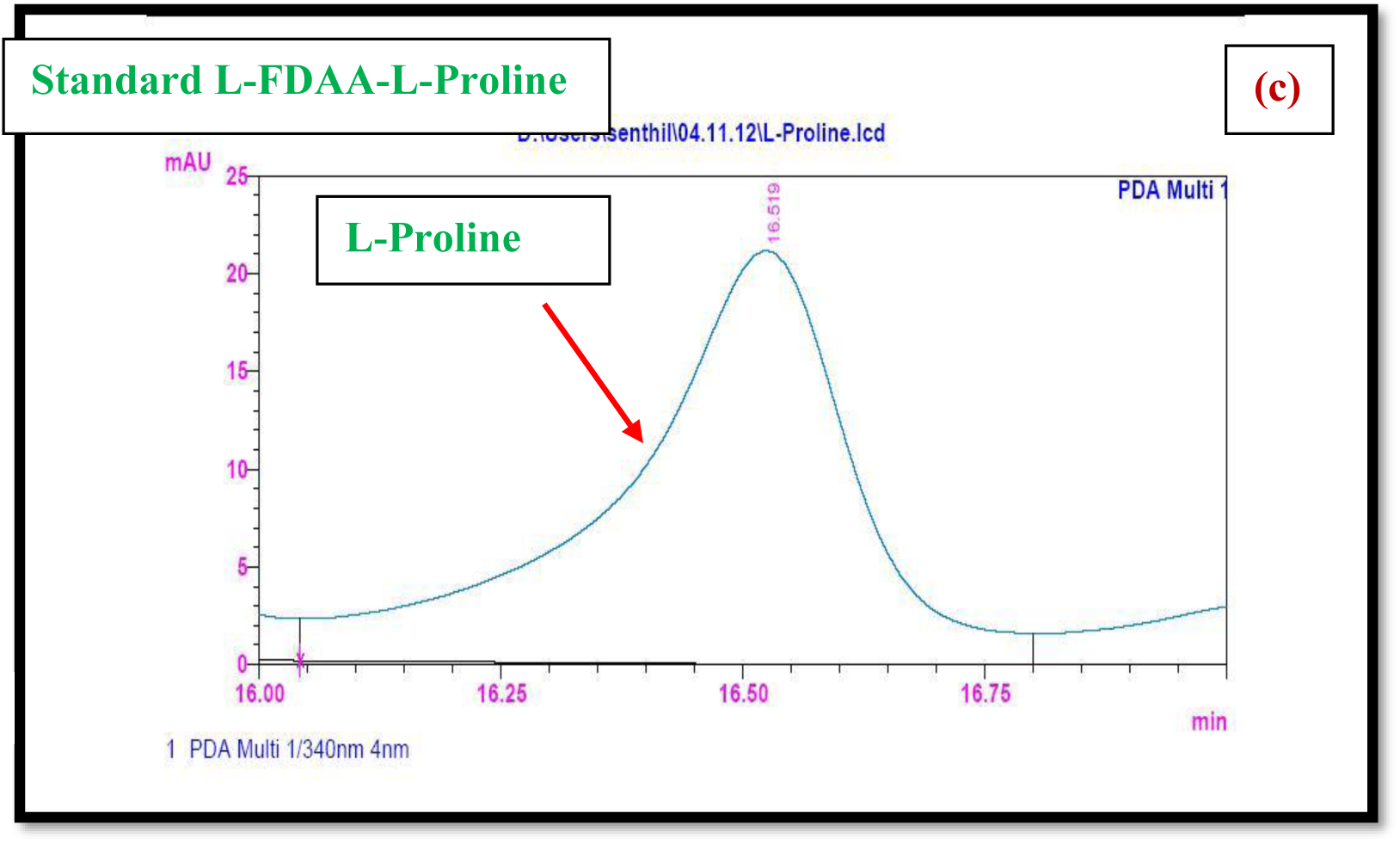
HPLC analysis of standard L-FDAA-L-Proline.

**Figure S119a.**
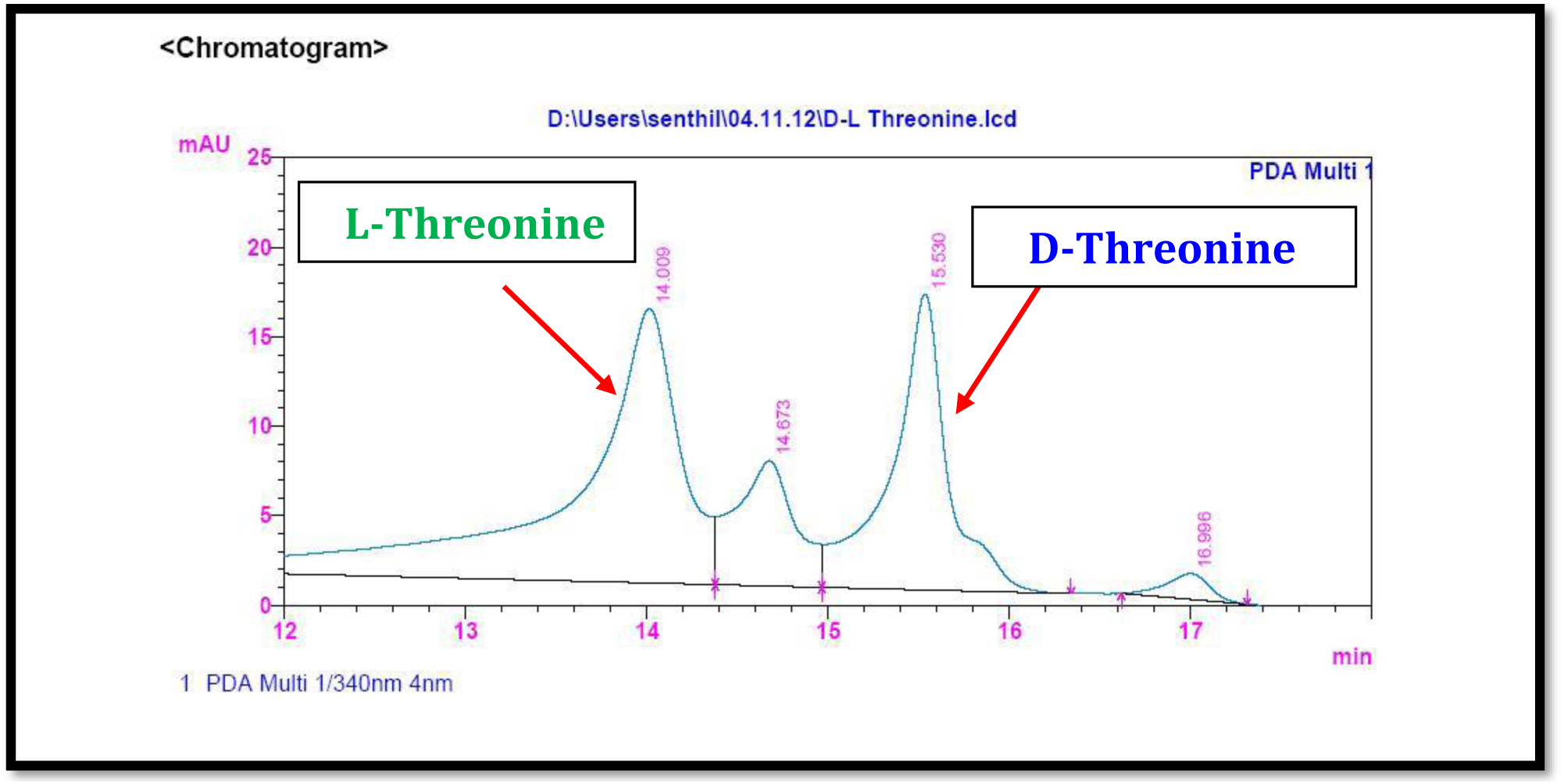
HPLC analysis of standard L-FDAA-D/L-Threonine.

**Figure S119b.**
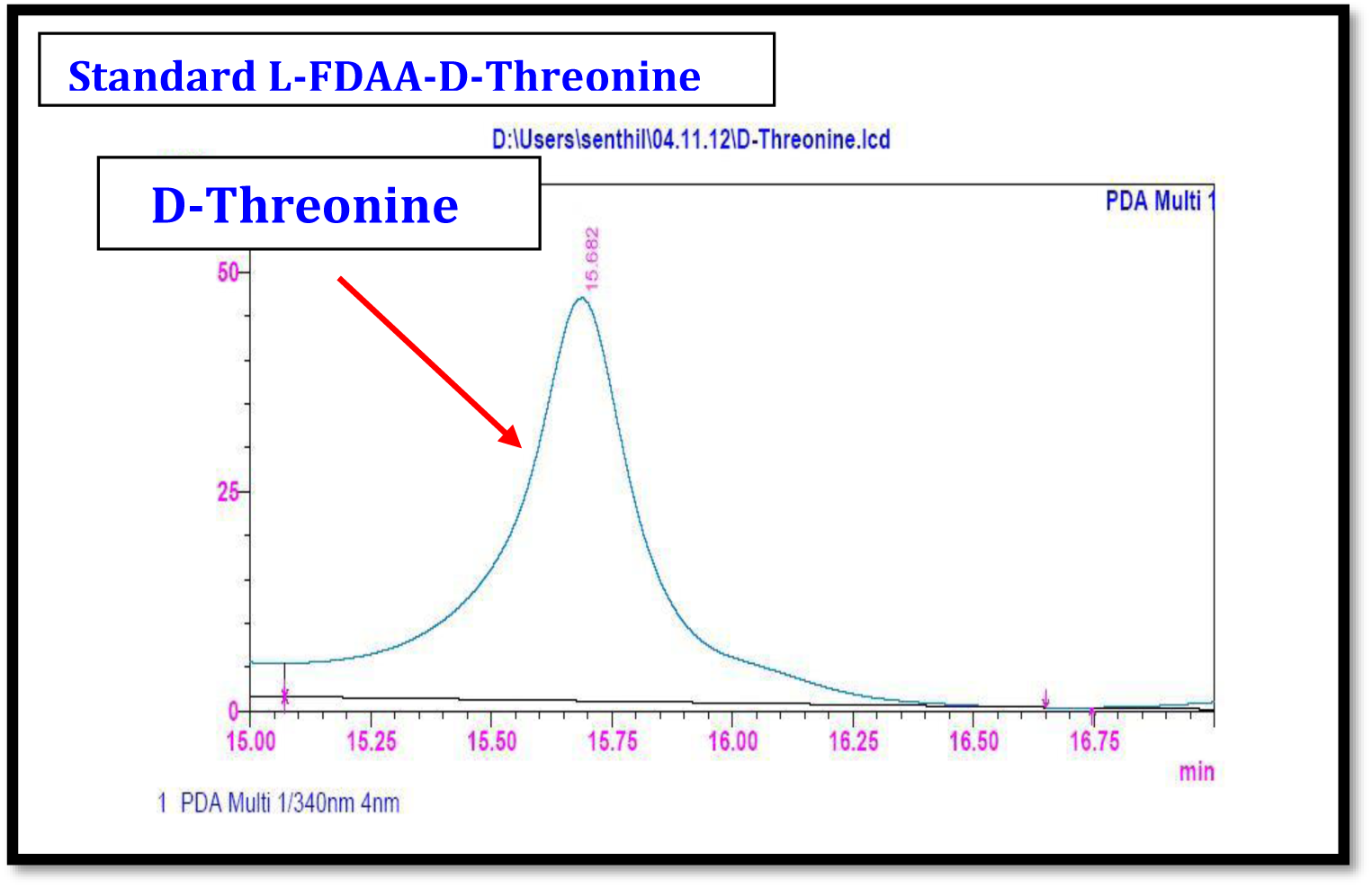
HPLC analysis of standard L-FDAA-D-Threonine.

**Figure S119c.**
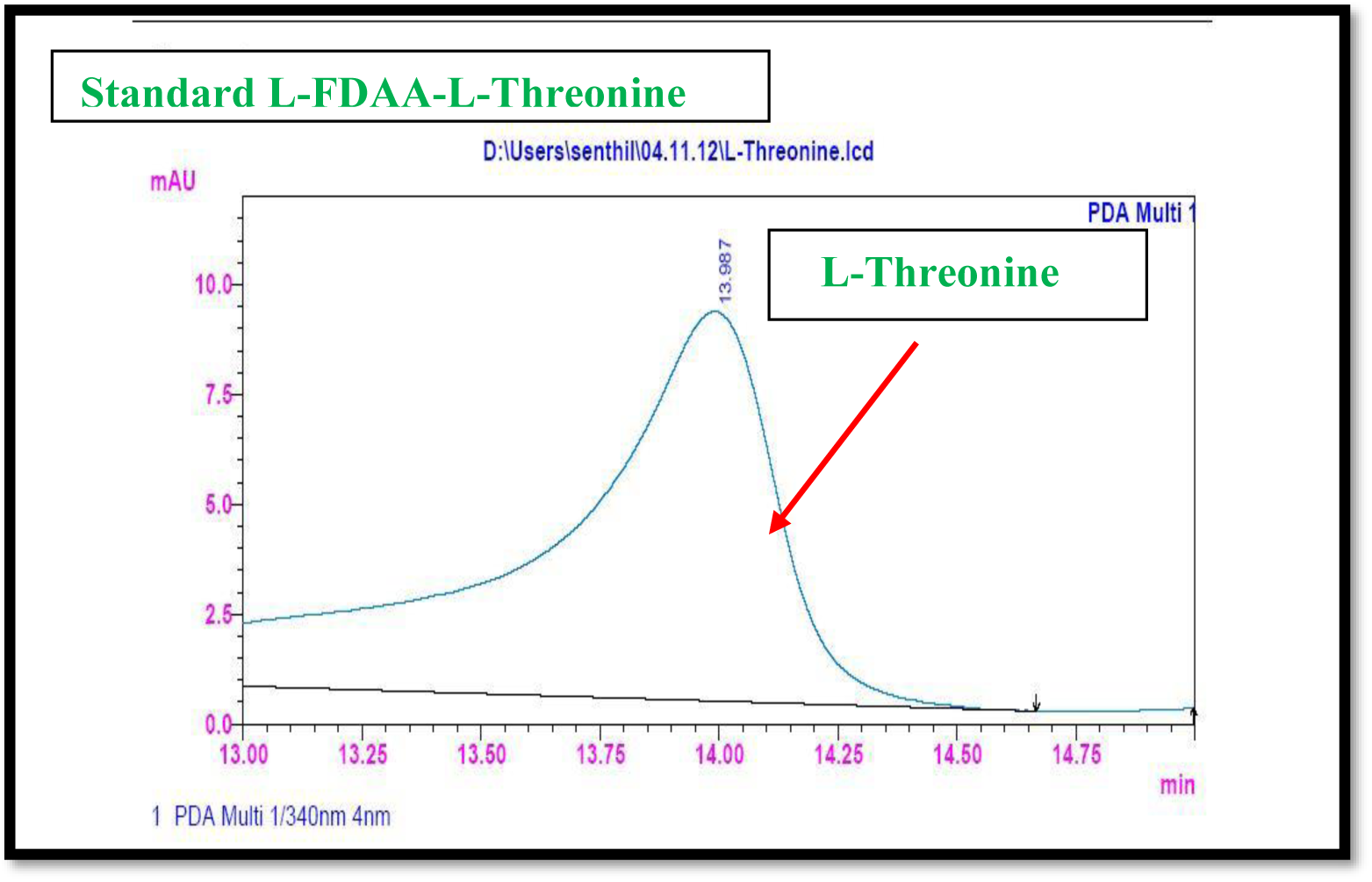
HPLC analysis of standard L-FDAA-D-Threonine.

**Figure S120a.**
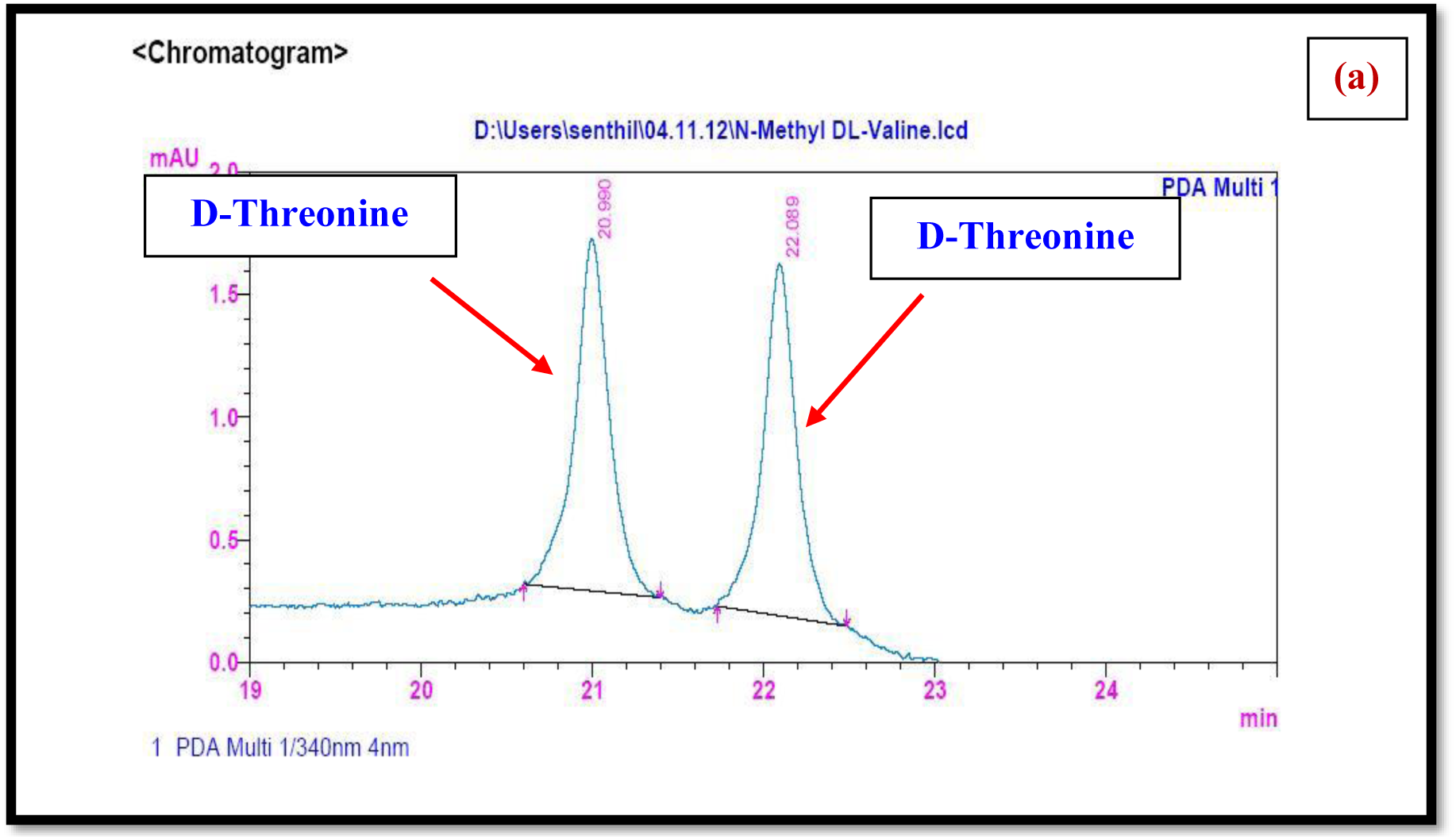
HPLC analysis of standard L-FDAA-D/L-N-Methyl Valine.

**Figure S120b.**
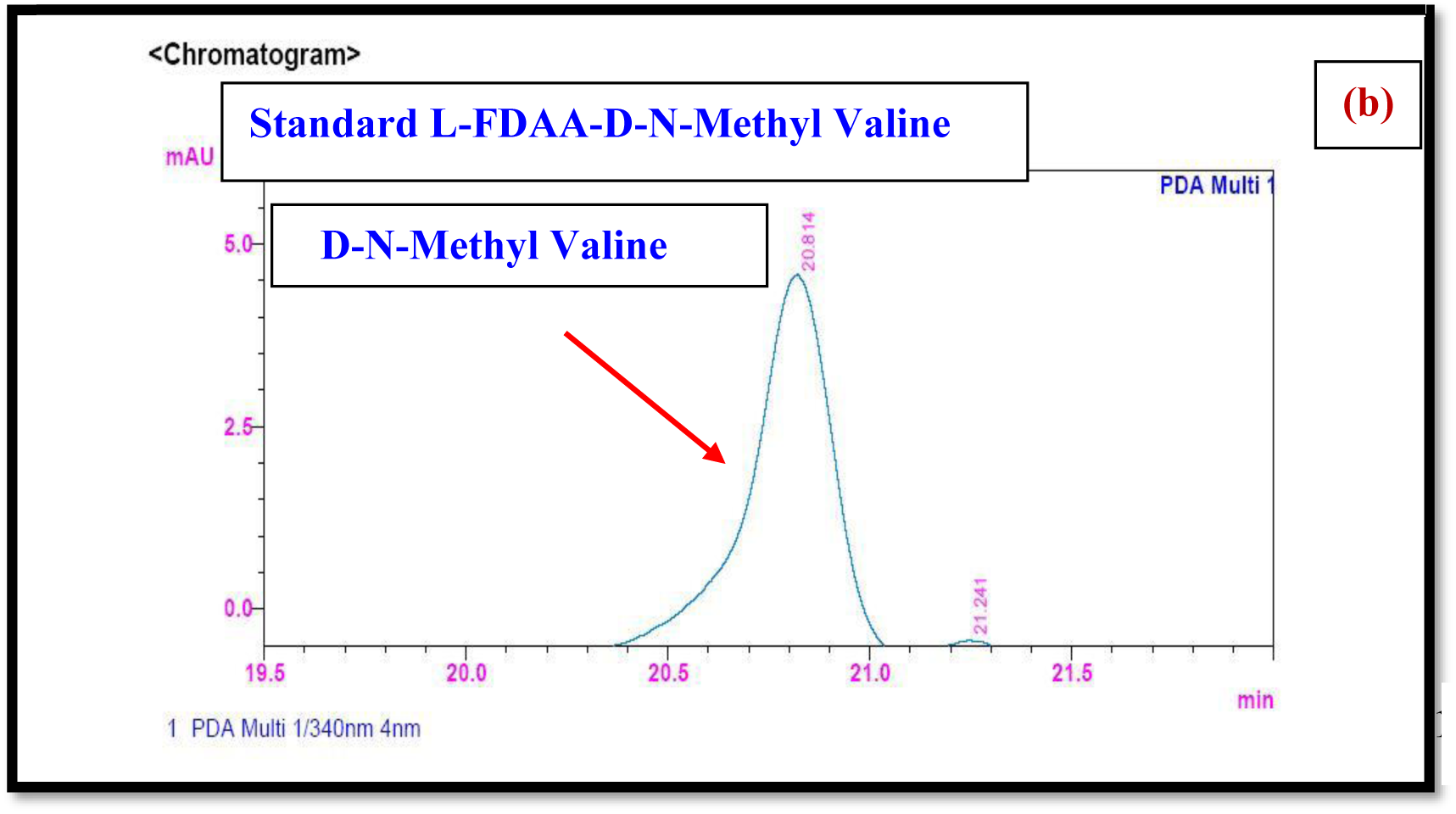
HPLC analysis of standard L-FDAA-D-N-Methyl Valine.

**Figure S120c.**
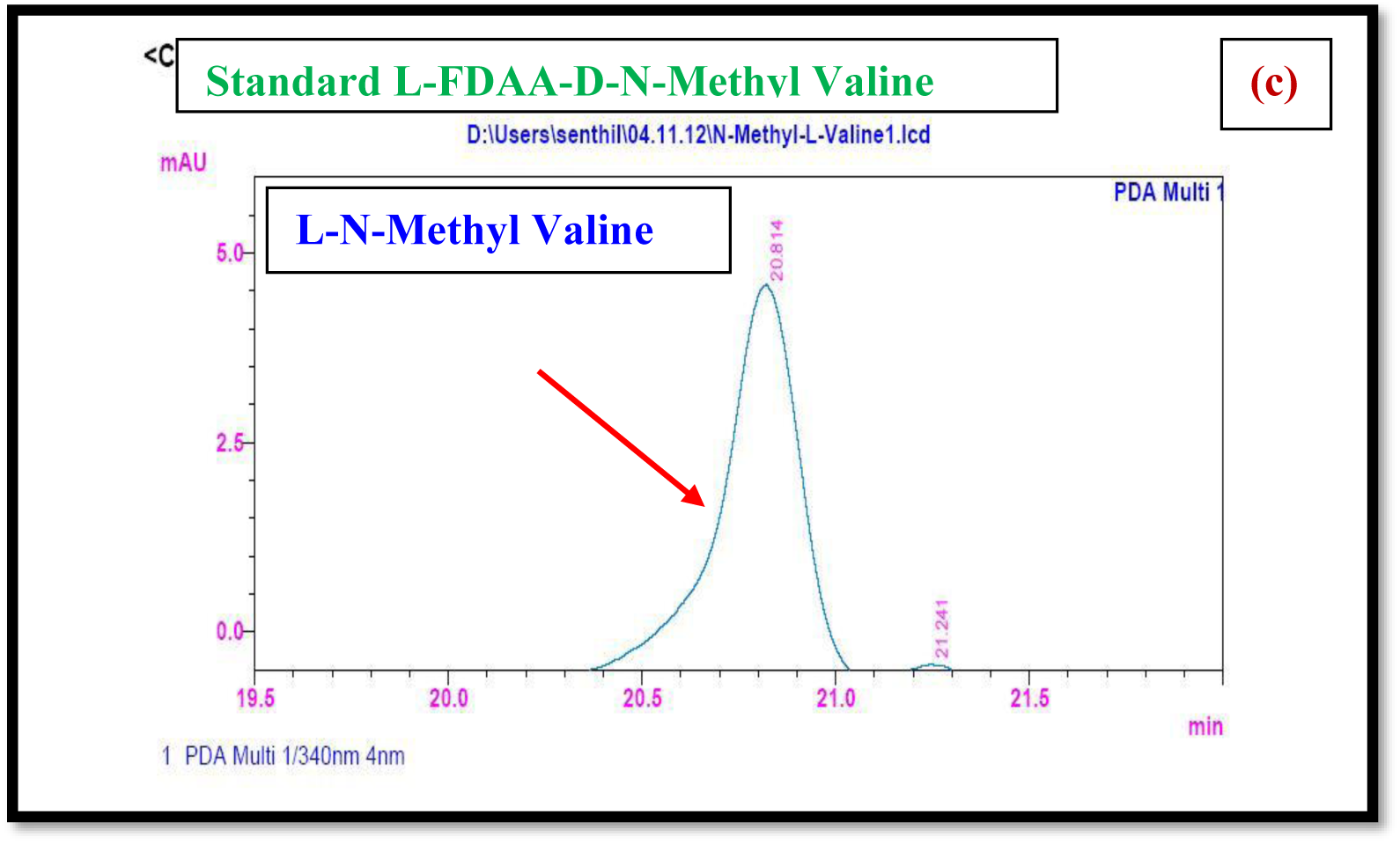
HPLC analysis of standard L-FDAA-D-N-Methyl Valine.

**Figure S121a.**
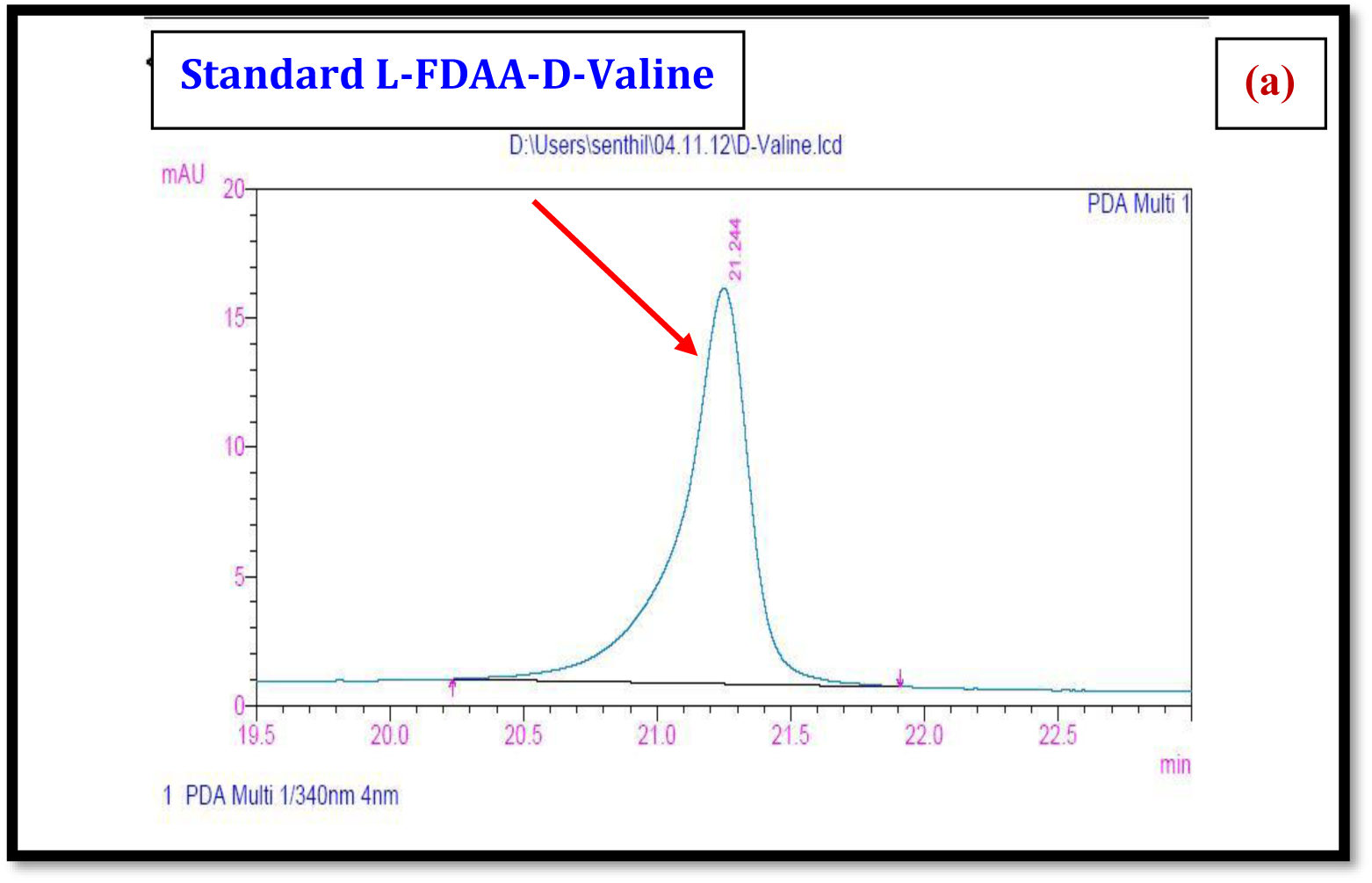
HPLC analysis of standard L-FDAA-D-Valine.

**Figure S121b.**
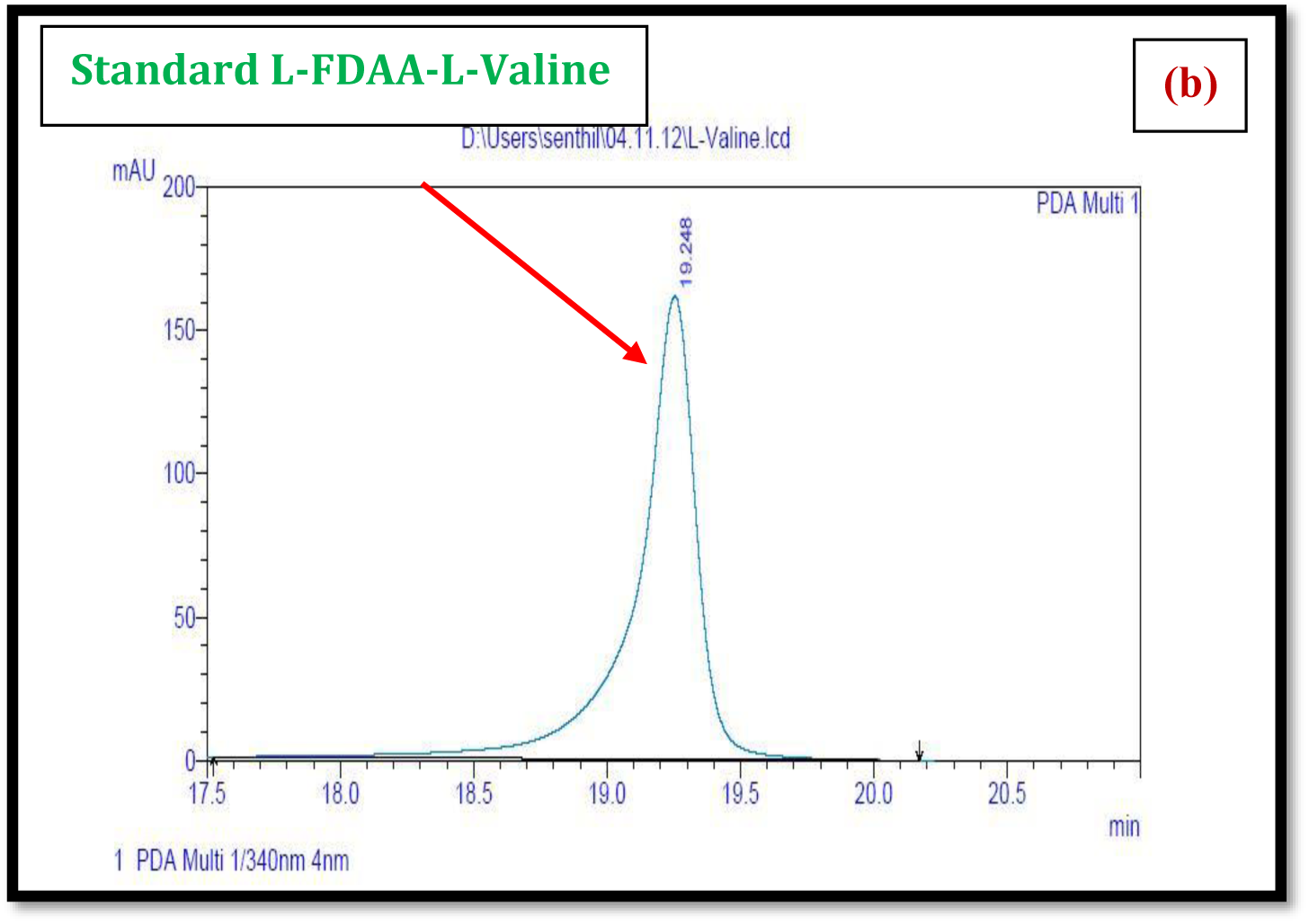
HPLC analysis of standard L-FDAA-L-Valine.

**Figure S121C.**
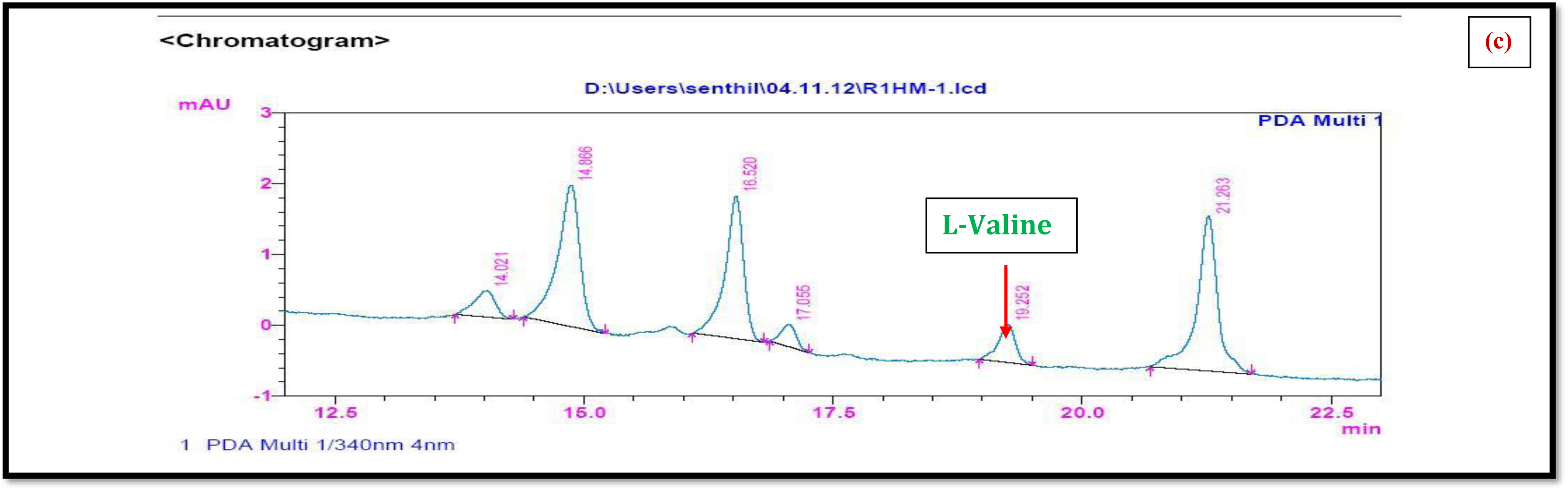
HPLC Analysis of L-FDAA Derivatives of acid hydrolysates of Transitmycin (R1)

**Figure S122a.**
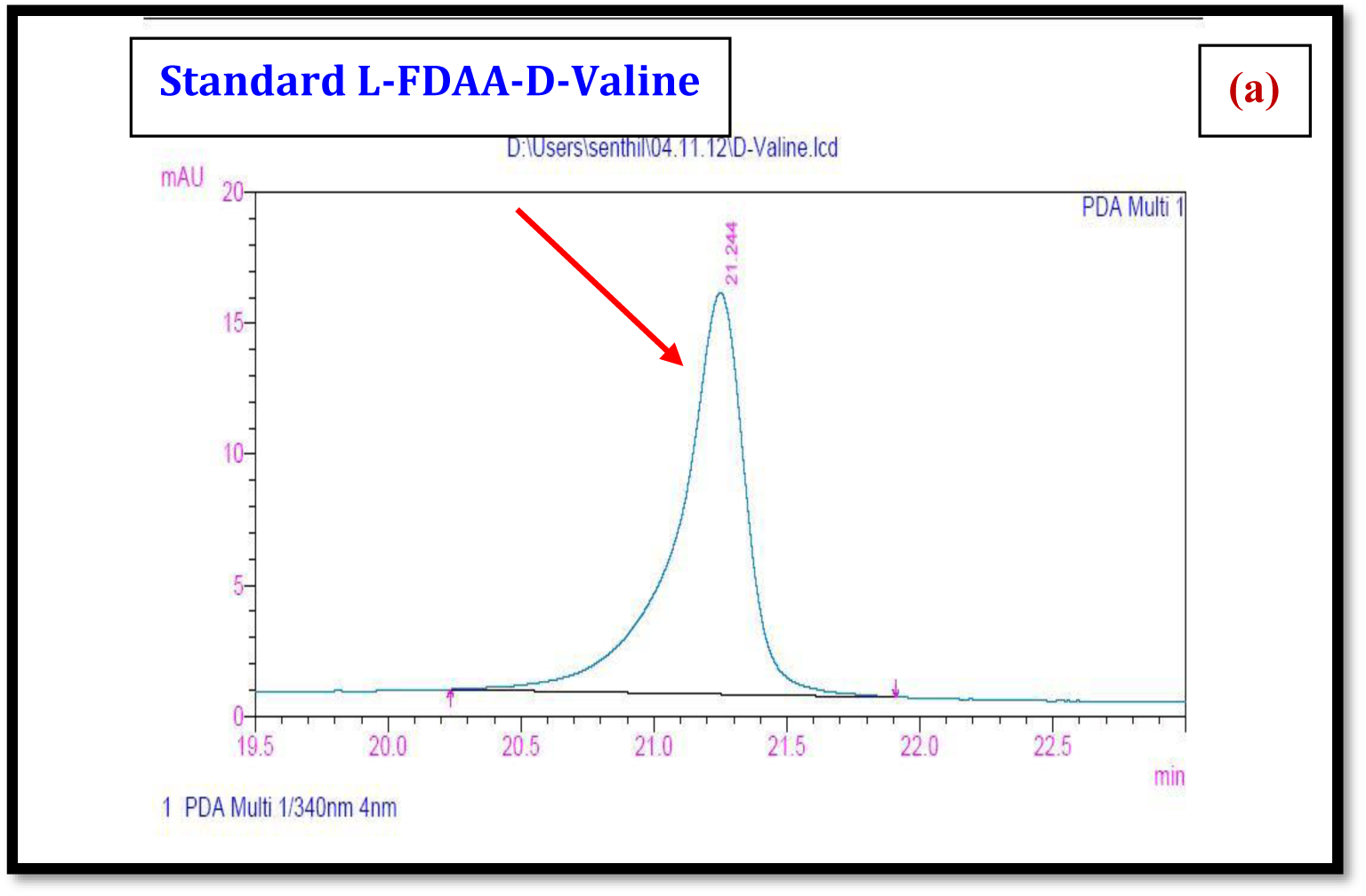
HPLC analysis of standard L-FDAA-D-Valine.

**Figure S122b.**
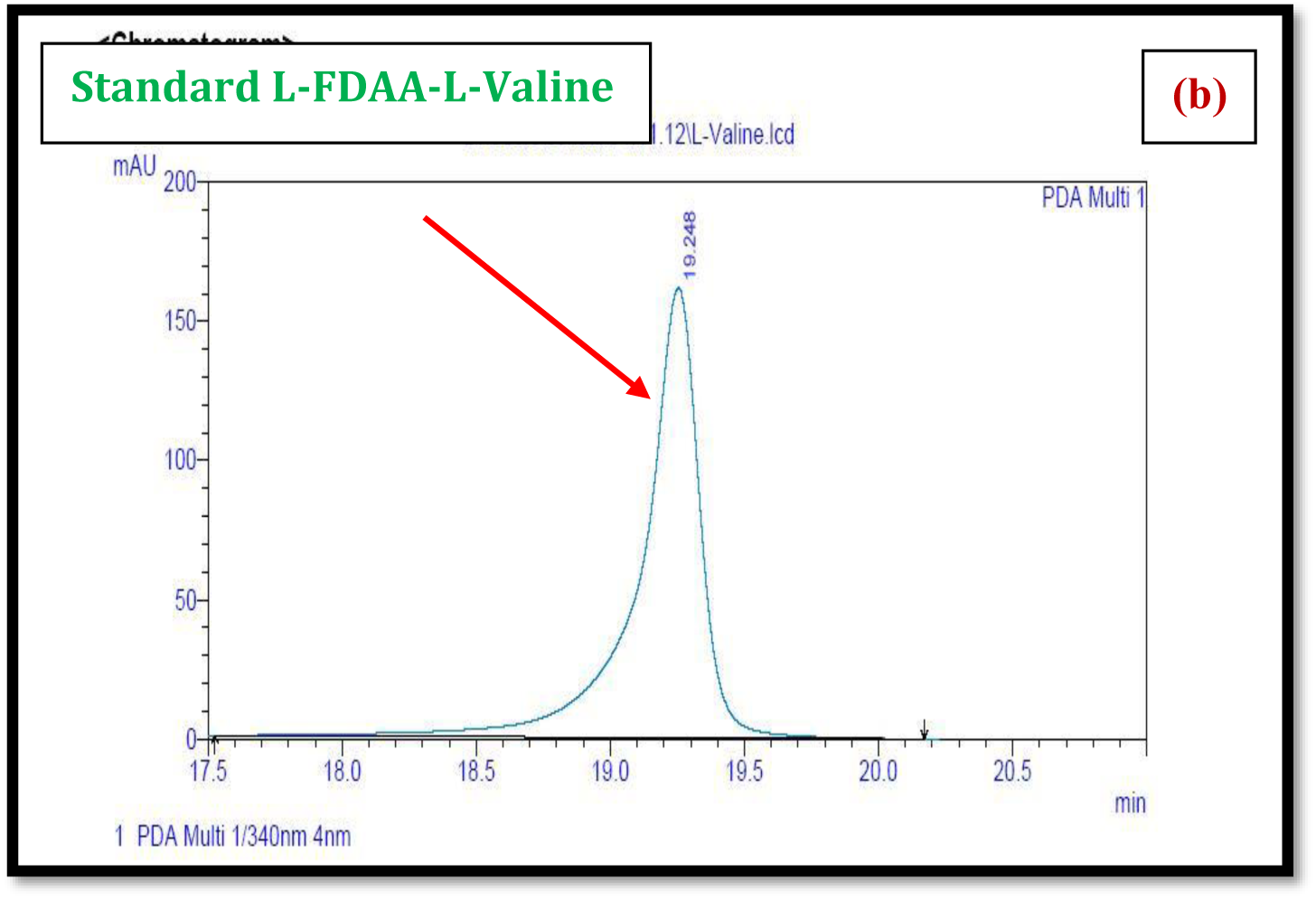
HPLC analysis of standard L-FDAA-L-Valine.

**Figure S122c.**
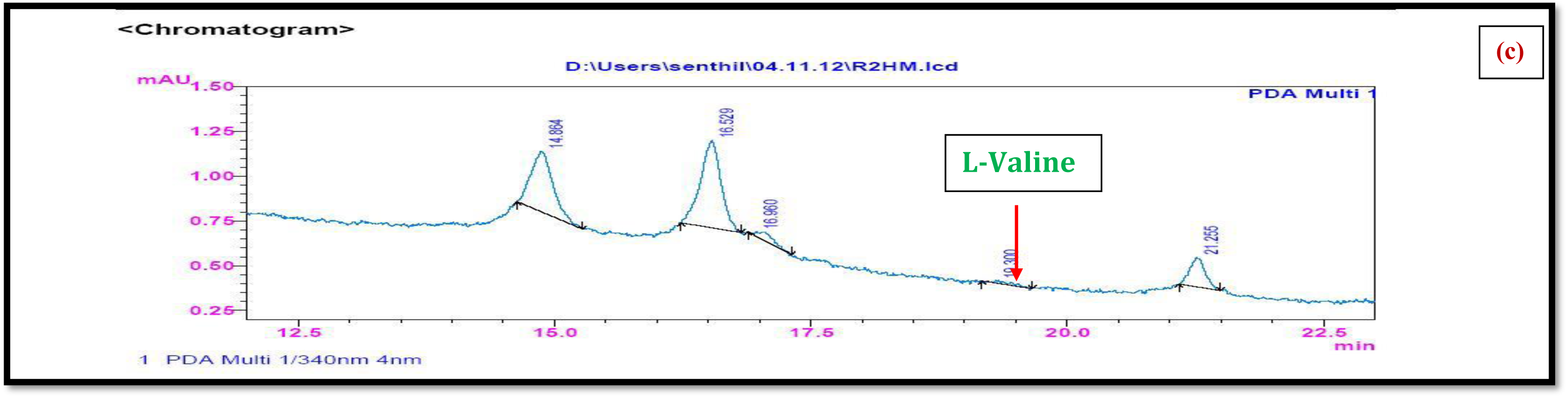
HPLC Analysis of L-FDAA Derivatives of acid hydrolysates of R2.

**Figure S123a.**
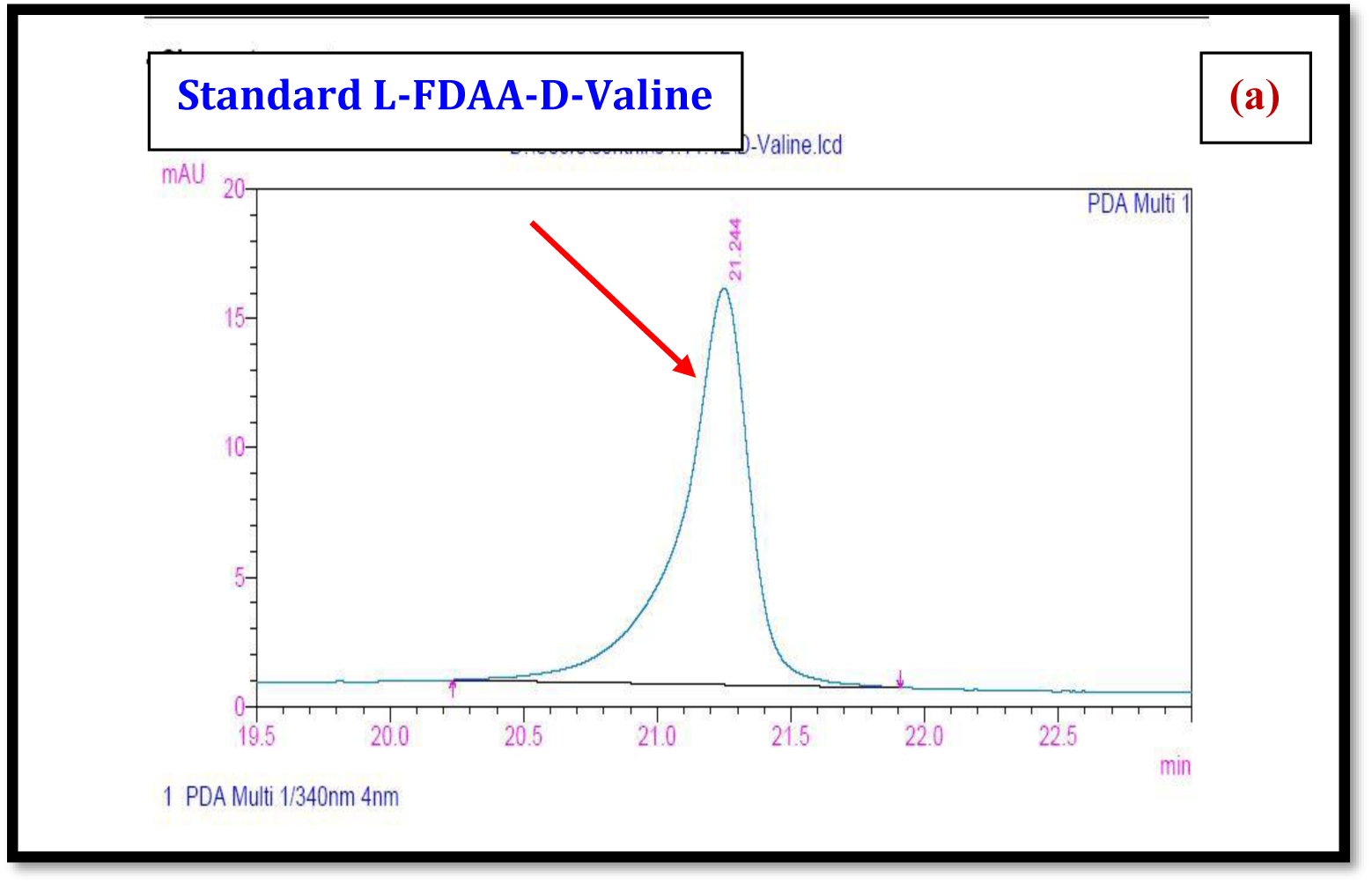
HPLC analysis of standard L-FDAA-D-Valine.

**Figure S123b.**
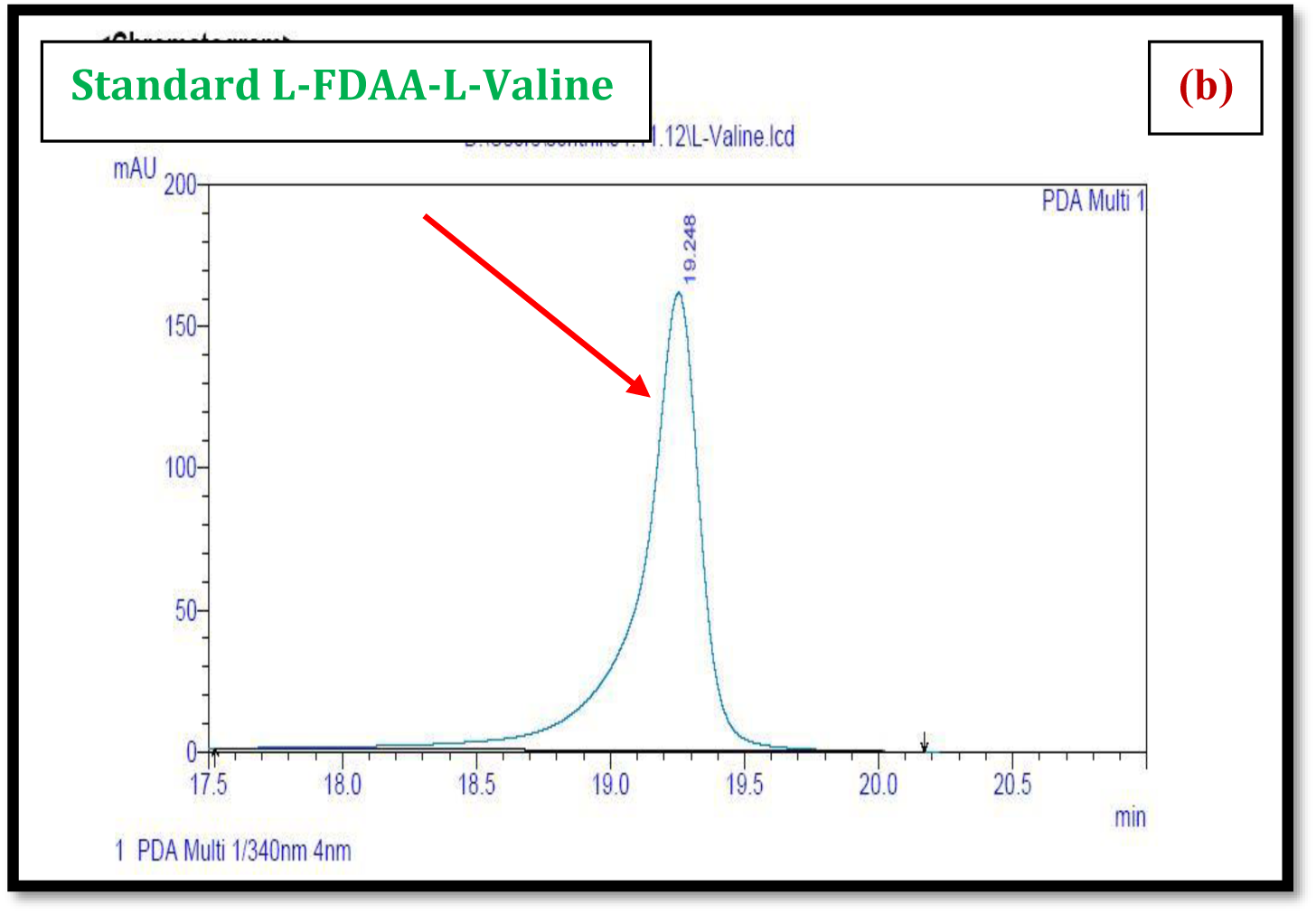
HPLC analysis of standard L-FDAA-L-Valine.

**Figure S123c.**
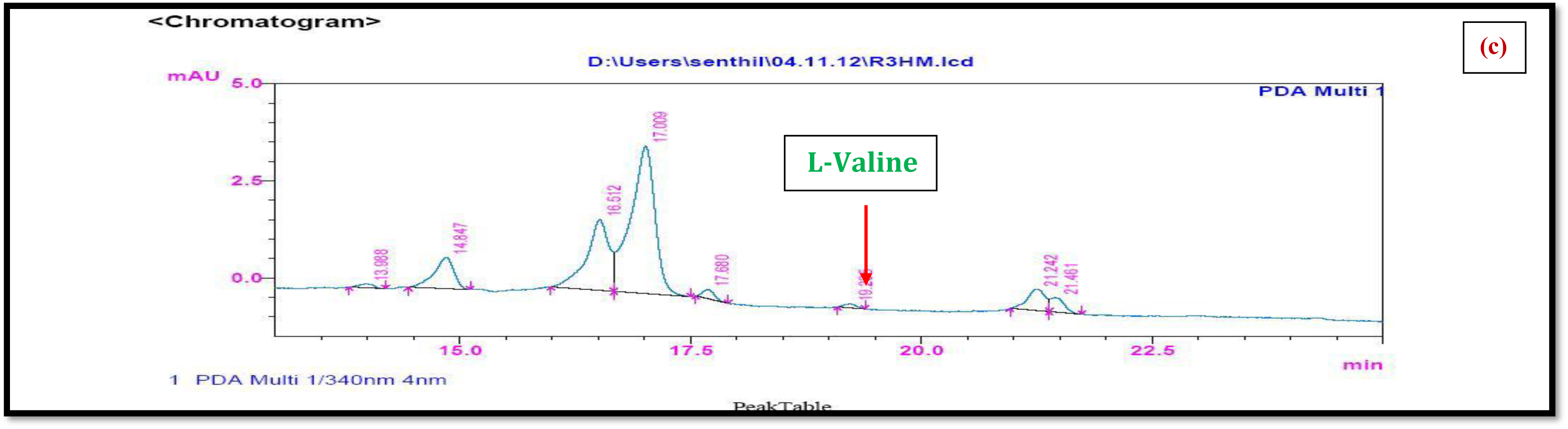
HPLC Analysis of L-FDAA Derivatives of acid hydrolysates of R3.

**Figure S124.**
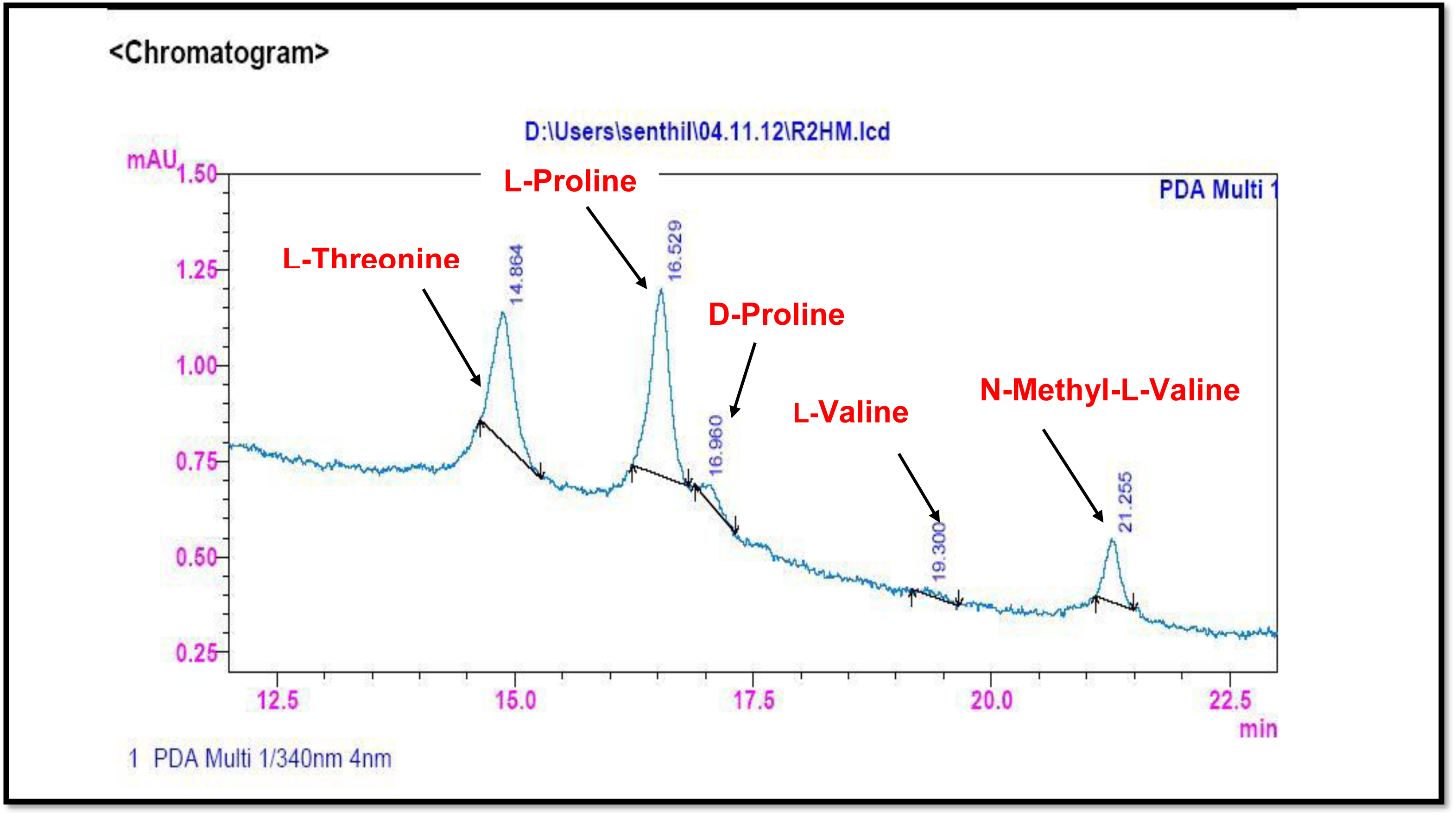
HPLC Analysis of L-FDAA Derivatives of acid hydrolysates of (R2)

**Figure S125.**
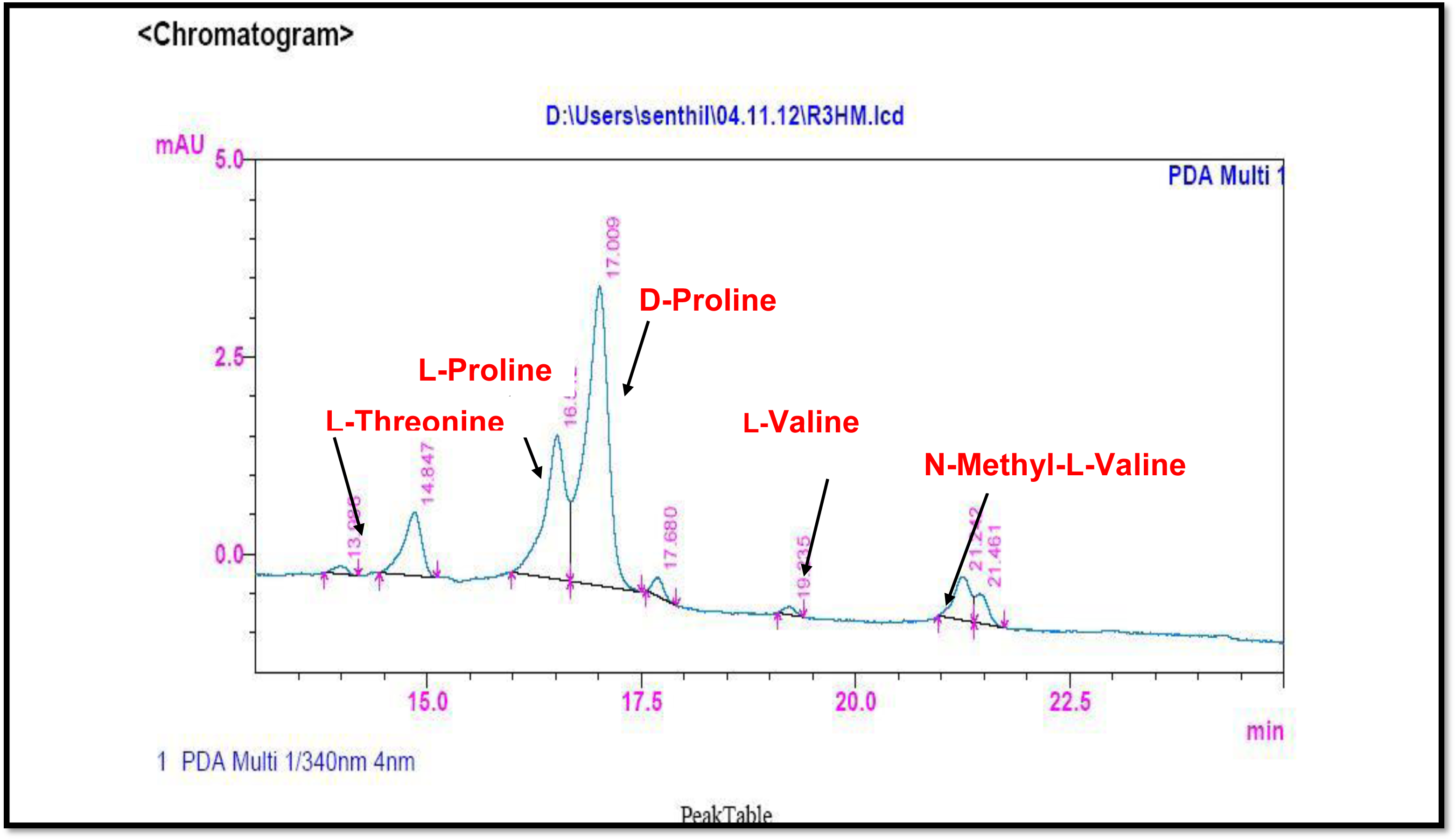
HPLC Analysis of L-FDAA Derivatives of acid hydrolysates of R3.

**Table 8a.** Analysis of L-FDAA derivates of acid hydrolysate of R2 by HPLC.

| Amino acids | HPLC retention times<br>Marfey's derivatives of standard amino acids |  | HPLC retention time<br>Marfey derivatives of<br>acid hydrolysate of<br>Transitmycin R2 | Assignment |
| --- | --- | --- | --- | --- |
|  | D | L |  |  |
| Threonine | 15.682 | 13.987 | 14.000 | L |
| Proline | 17.035 | 16.519 | 16.529 & 16.960 | D & L |
| Valine | 21.244 | 19.248 | 19.300 | L |
| N-methyl valine | 22.089 | 20.814 | 21.255 | L |

**Table 8b.** Analysis of L-FDAA derivatives of acid hydrolysate of R3 by HPLC.

| Amino acids | HPLC retention times<br>Marfey's derivatives of standard amino acids |  | HPLC retention time<br>Marfeyderivatives of<br>acid hydrolysate of<br>Transitmycin R3 | Assignment |
| --- | --- | --- | --- | --- |
|  | D | L |  |  |
| Threonine | 15.682 | 13.987 | 13.988 | L |
| Proline | 17.035 | 16.519 | 16.512 & 17.000 | D & L |
| Valine | 21.244 | 19.248 | 19.235 | L |
| N-methyl valine | 22.089 | 20.814 | 21.242 | L |

**Figure S126.**
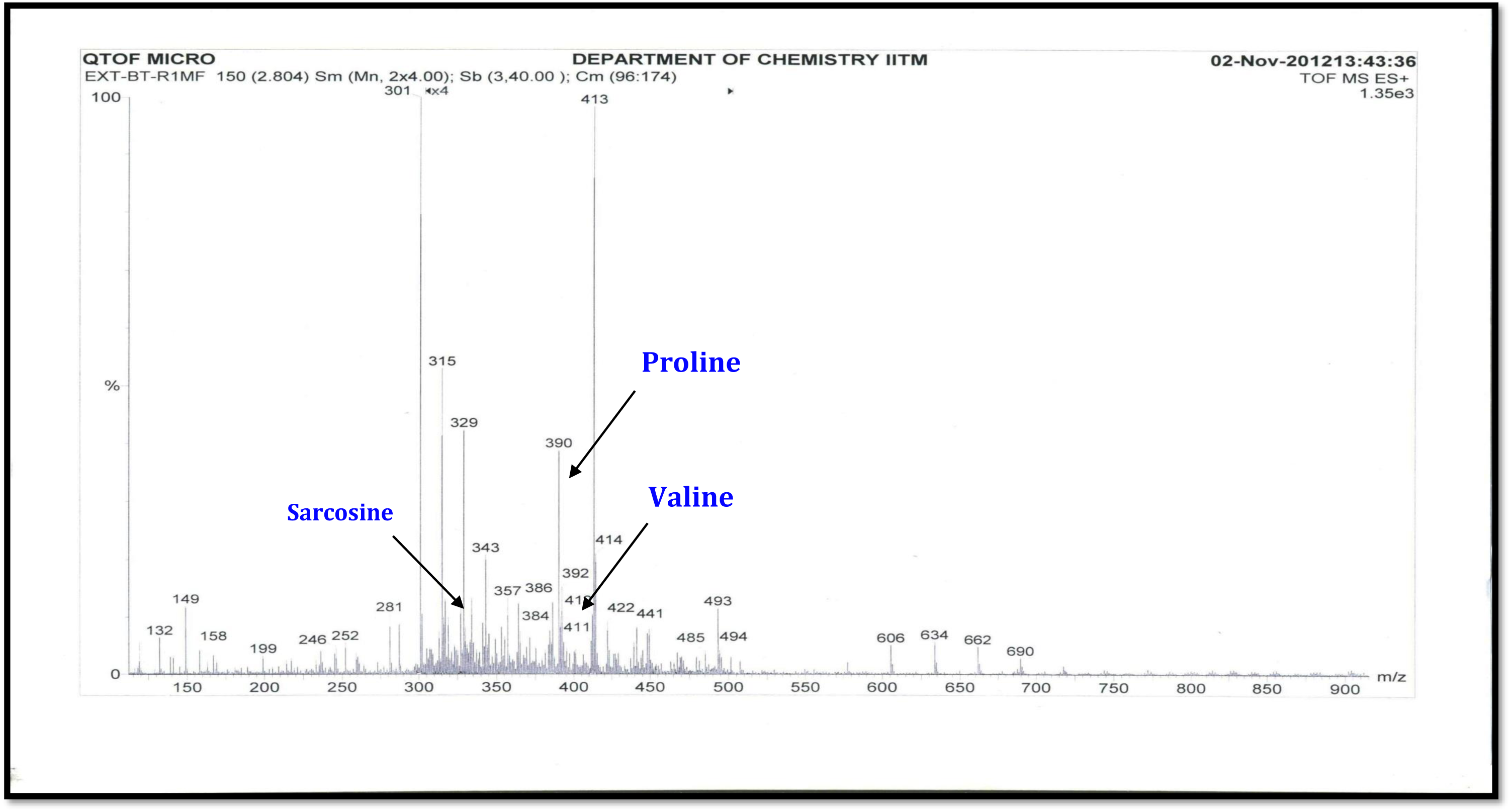
ESI- MS Spectrum of Marfey’s Derivatives of Transitmycin (R1) (Positive mode)

**Figure S127.**
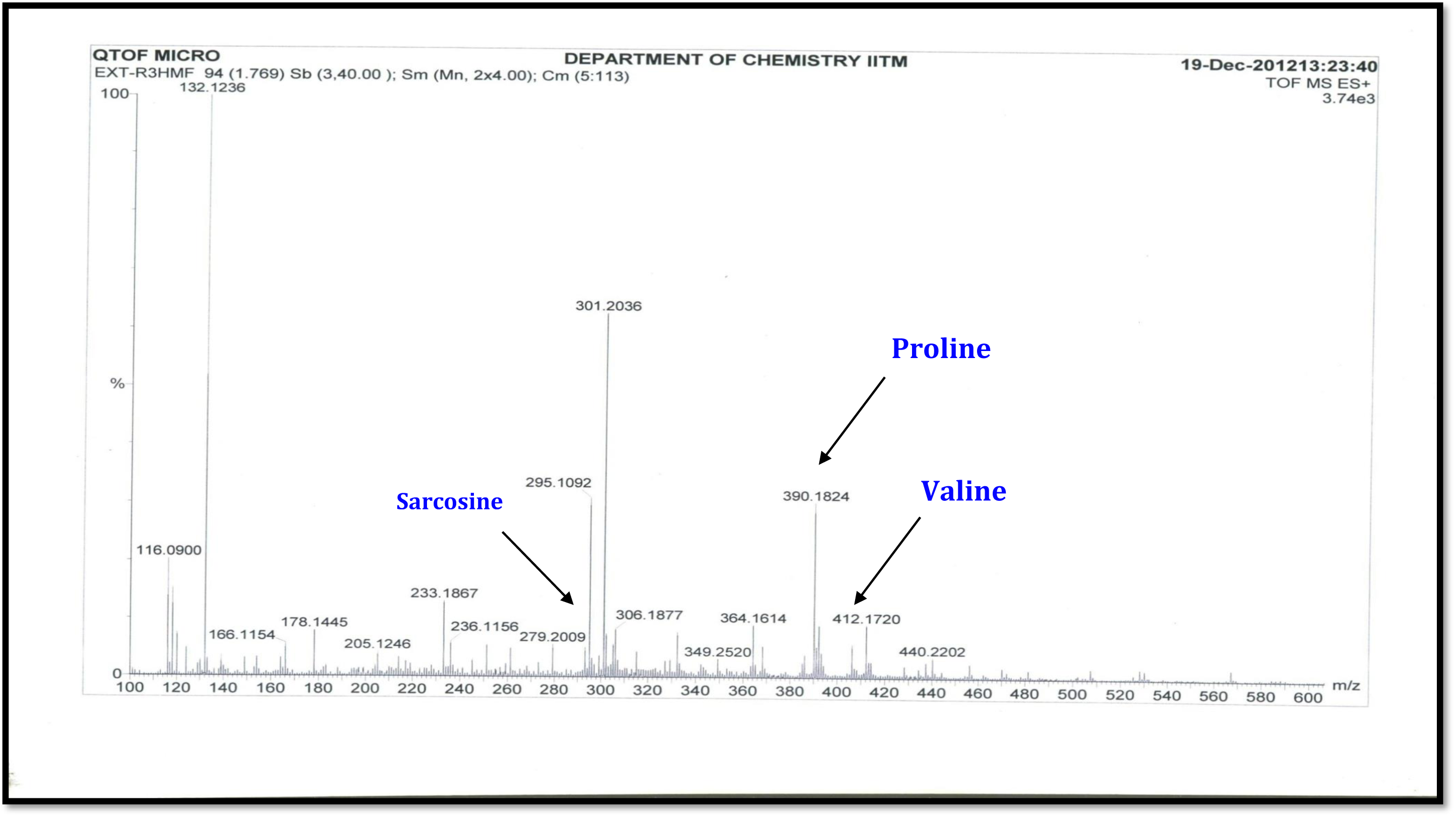
ESI- MS Spectrum of Marfey’s Derivatives of Transitmycin (R1) (Positive mode)

**Figure S128.**
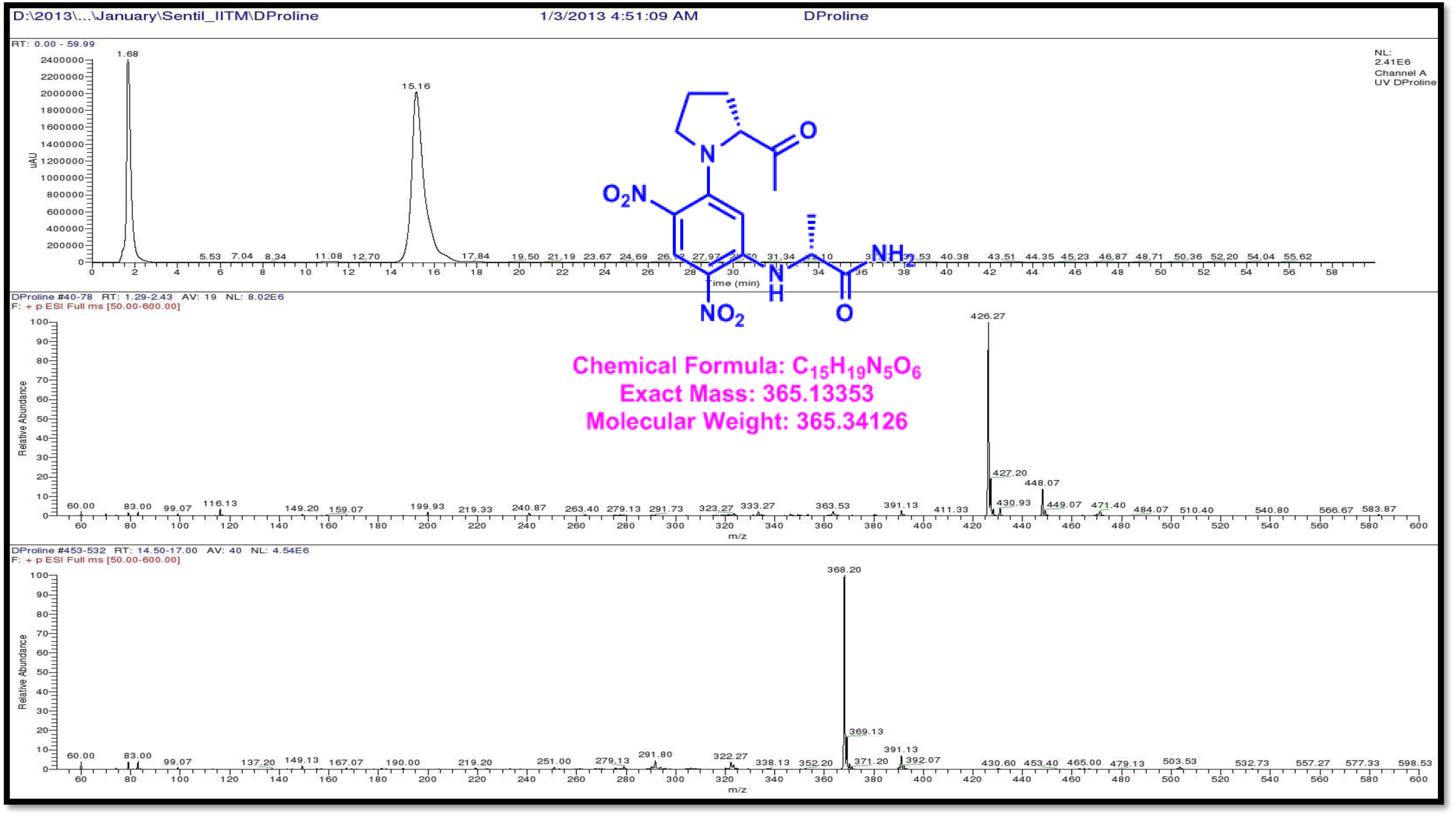
LCMS analysis of Standard L-FDAA-D-Proline (Positive mode)

**Figure S129.**
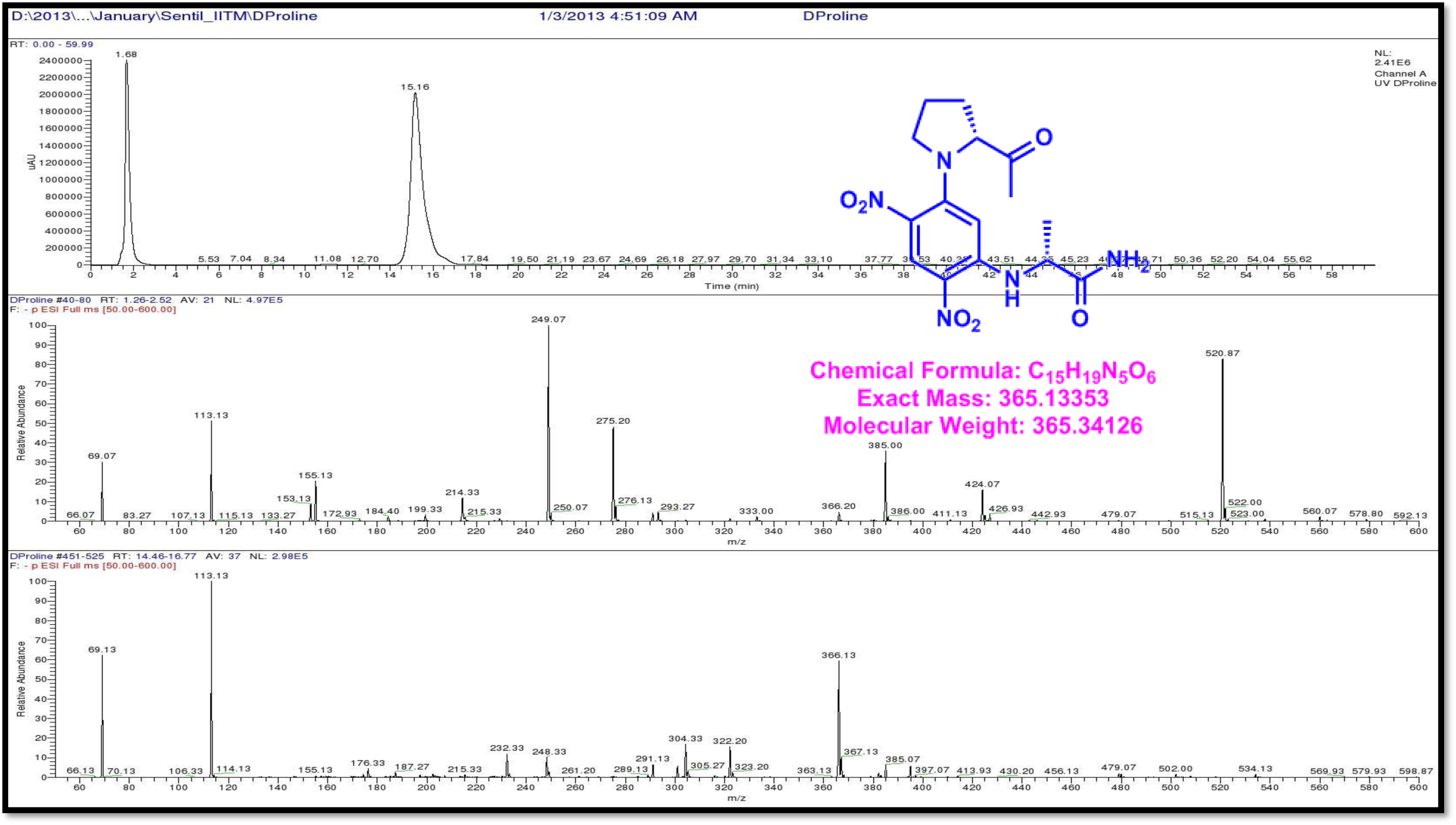
LCMS analysis of Standard L-FDAA-D-Proline (Negative mode)

**Figure S130.**
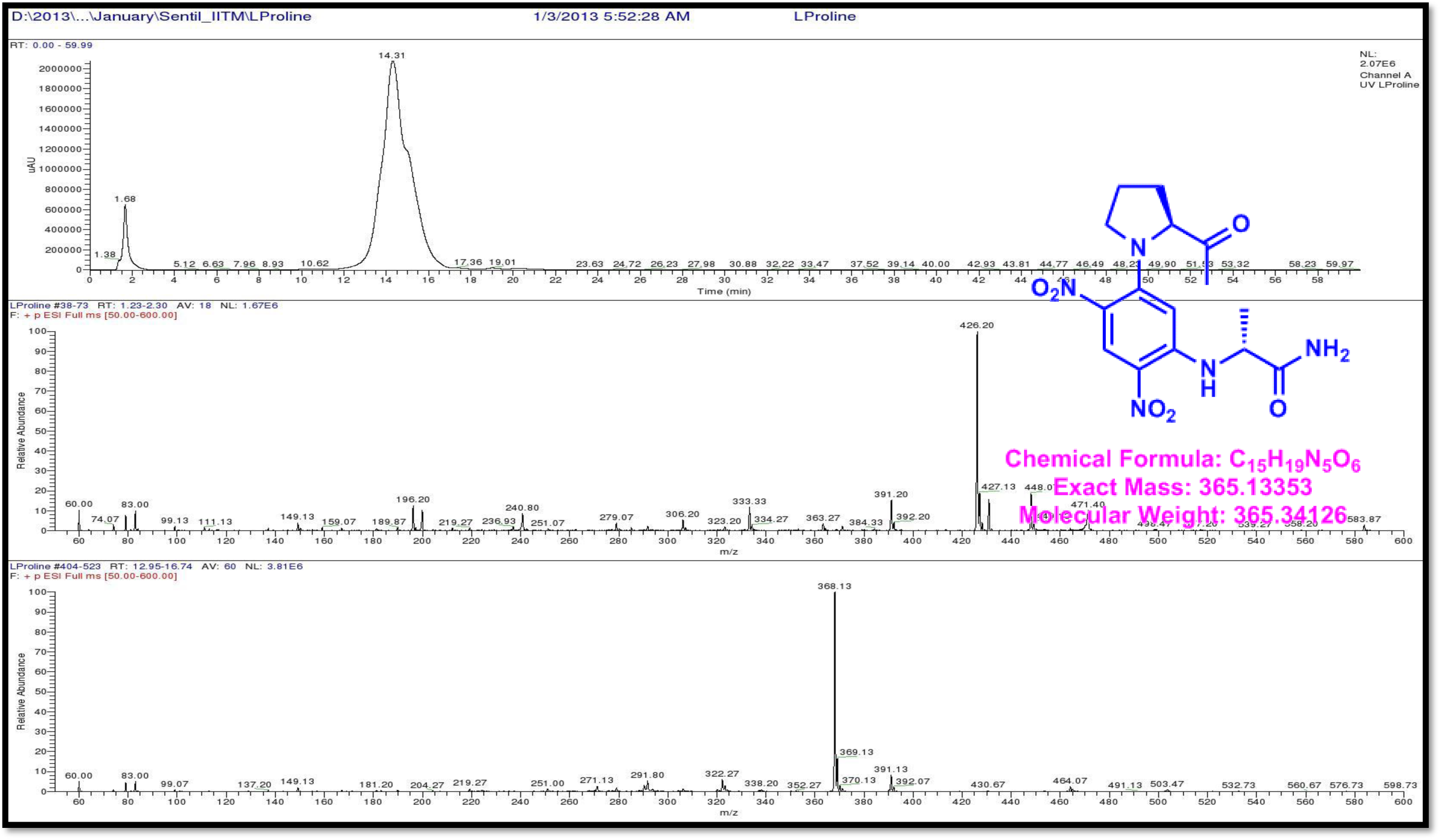
LCMS analysis of Standard L-FDAA-L-Proline (Positive mode)

**Figure S131.**
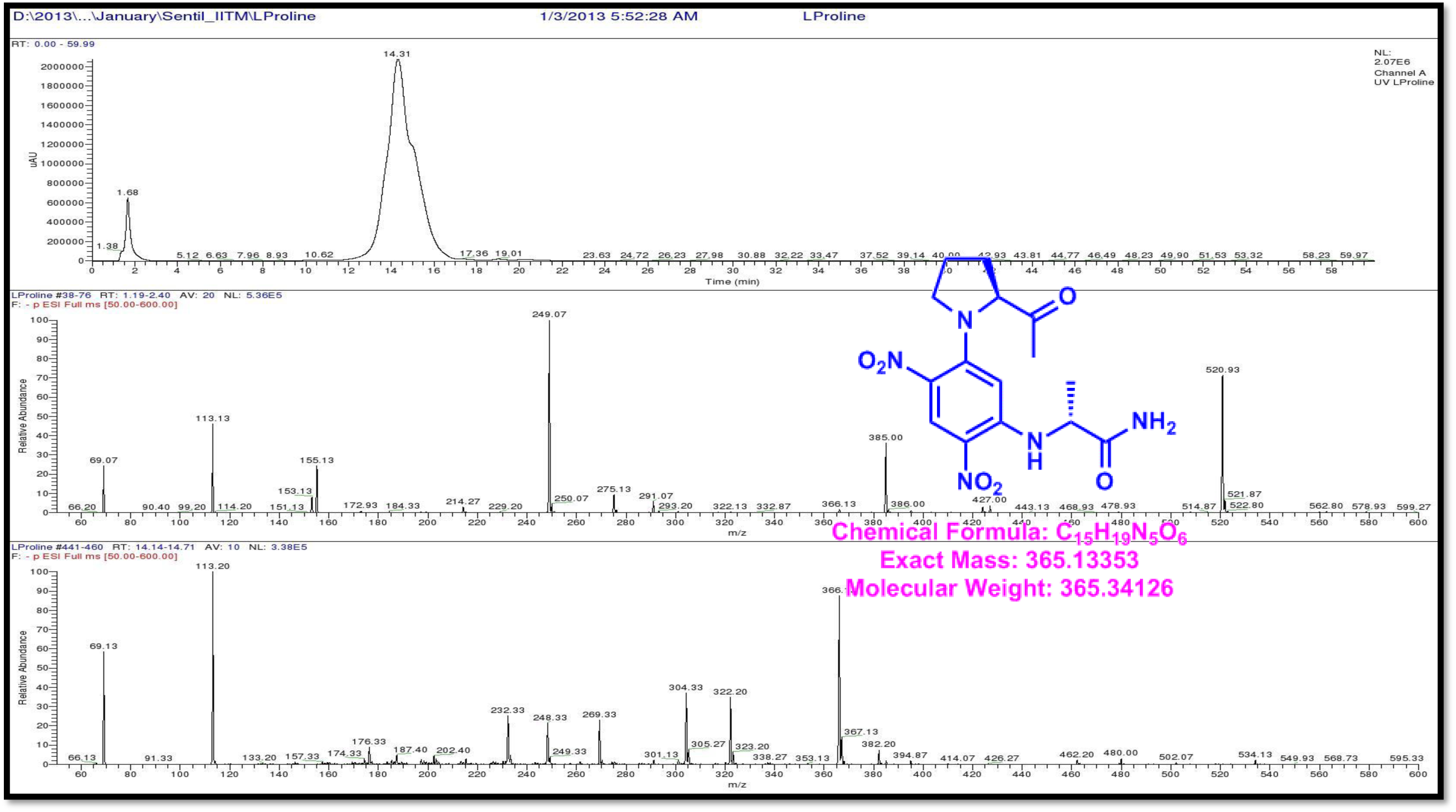
LCMS analysis of Standard L-FDAA-L-Proline (Negative mode)

**Figure S132.**
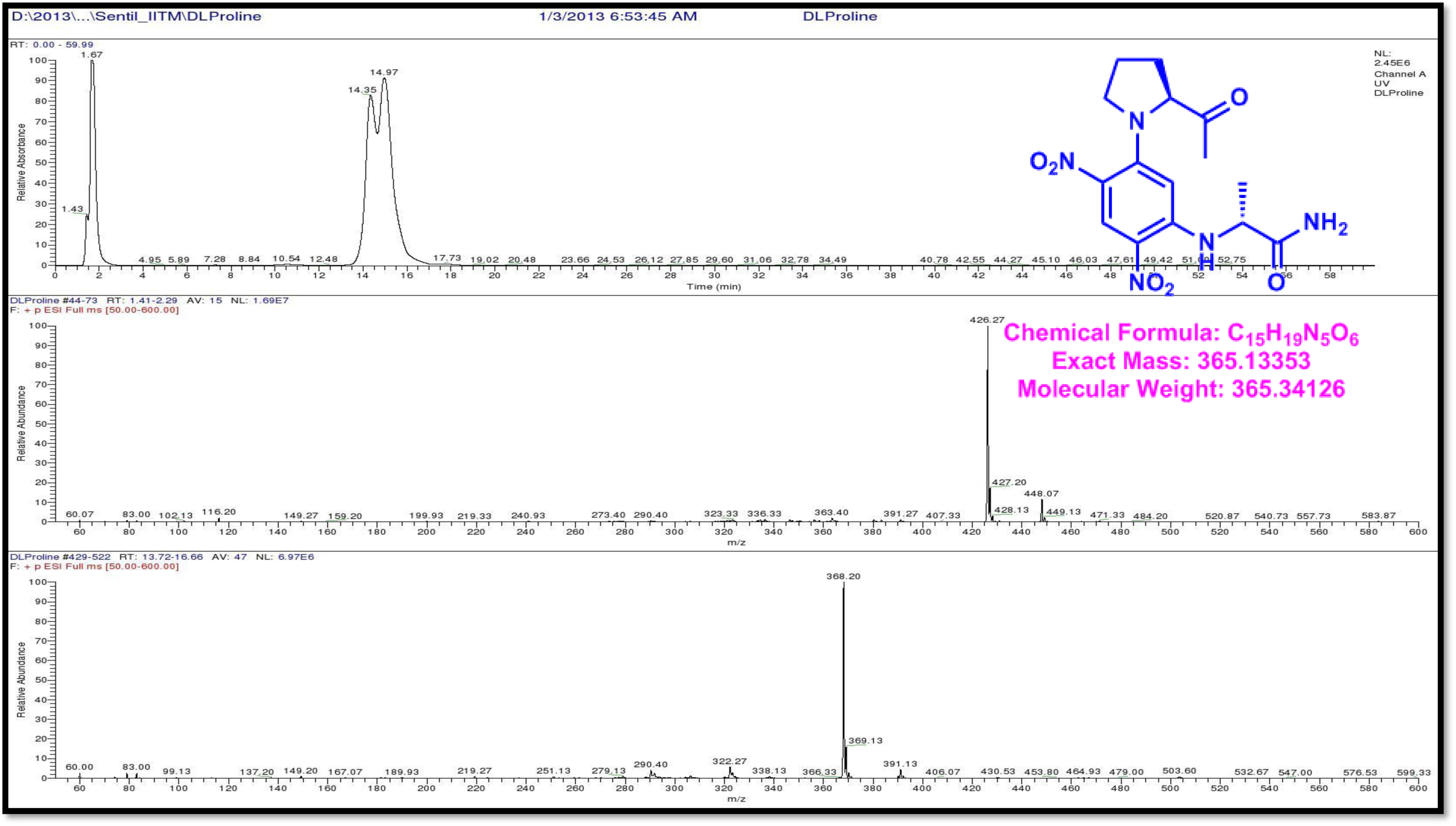
LCMS analysis of Standard L-FDAA-D/L-Proline (Positive mode)

**Figure S133.**
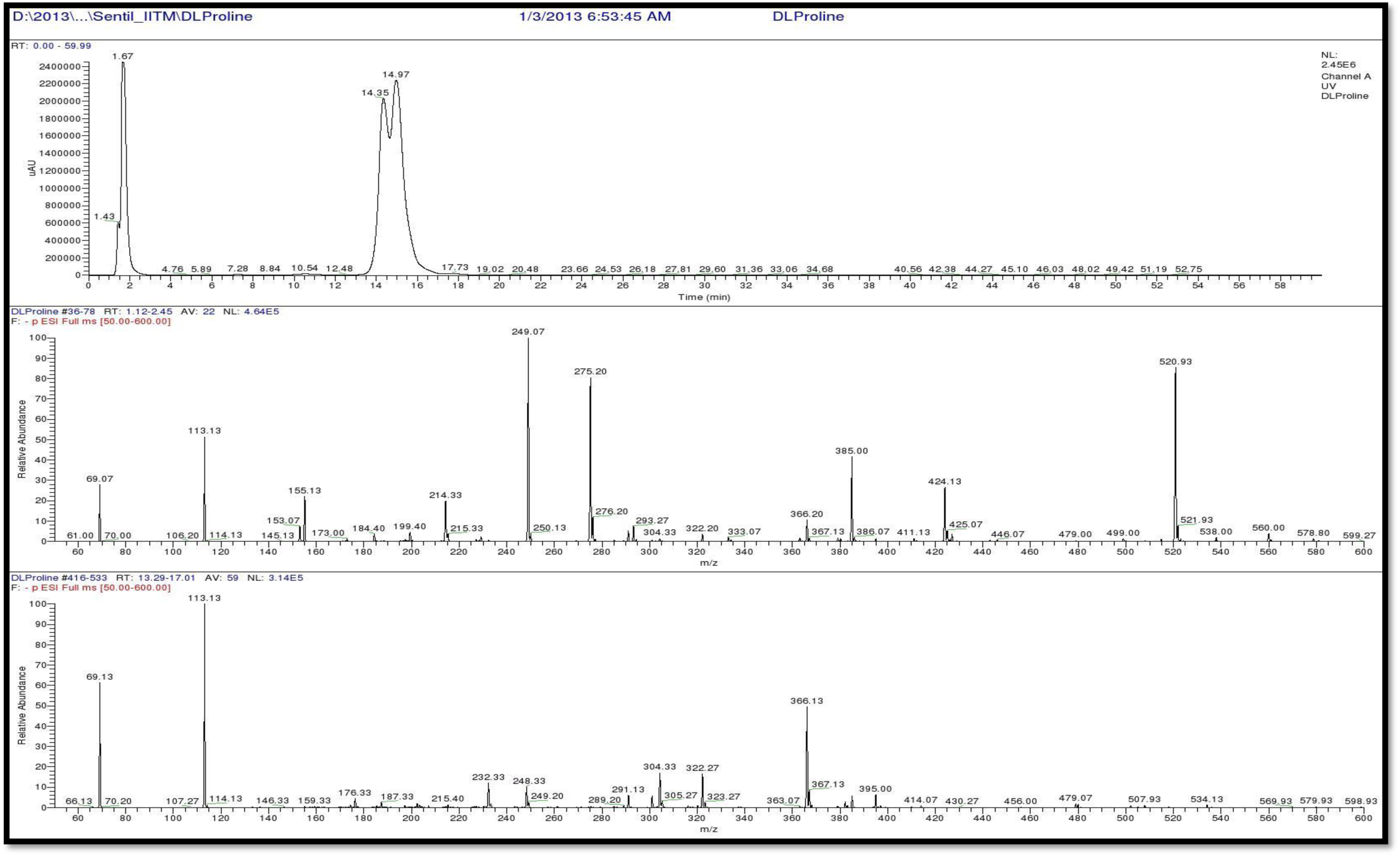
LCMS analysis of Standard L-FDAA-D/L-Proline (Negative mode)

**Figure S134.**
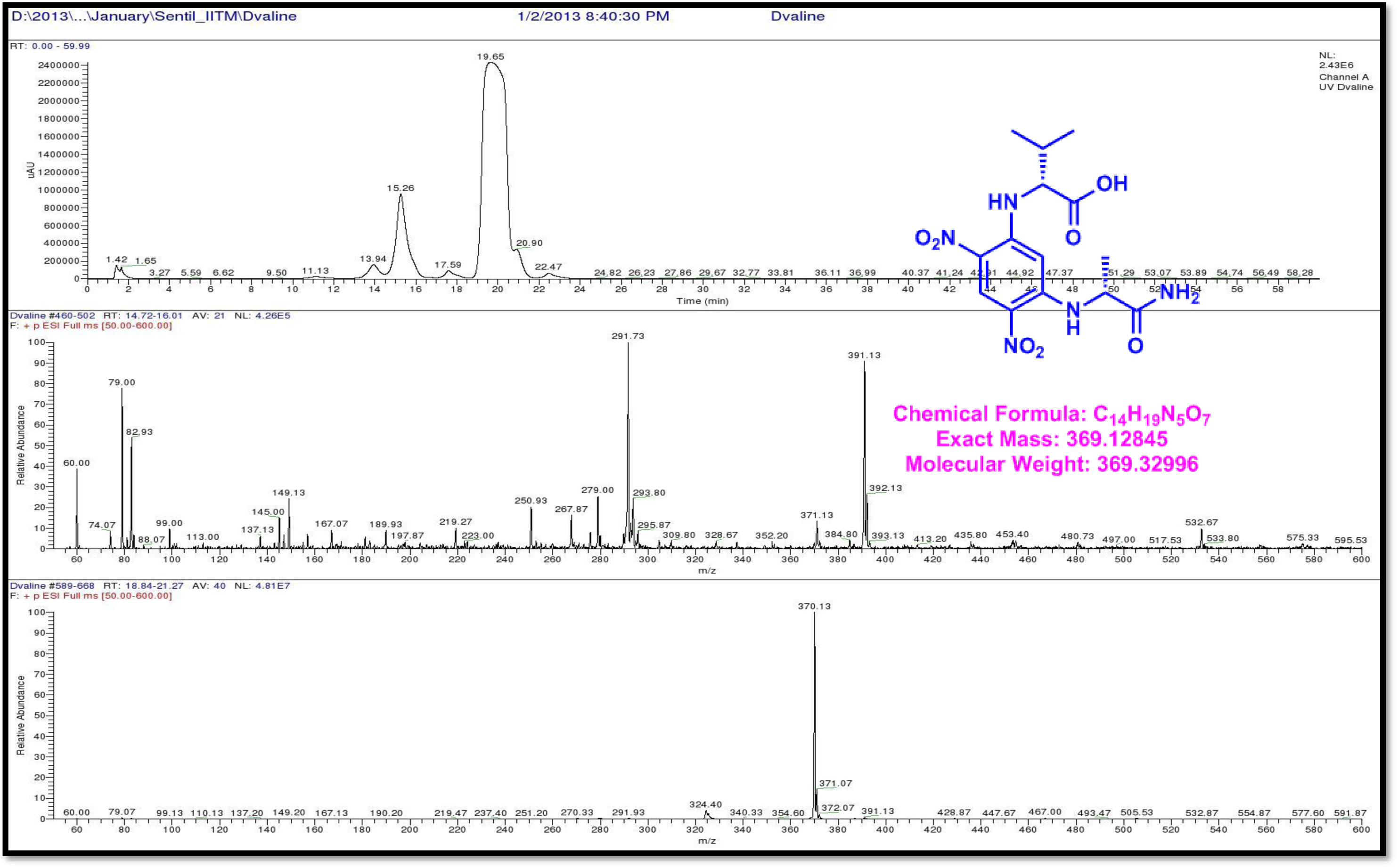
LCMS analysis of Standard L-FDAA-D-Valine (Positive mode)

**Figure S135.**
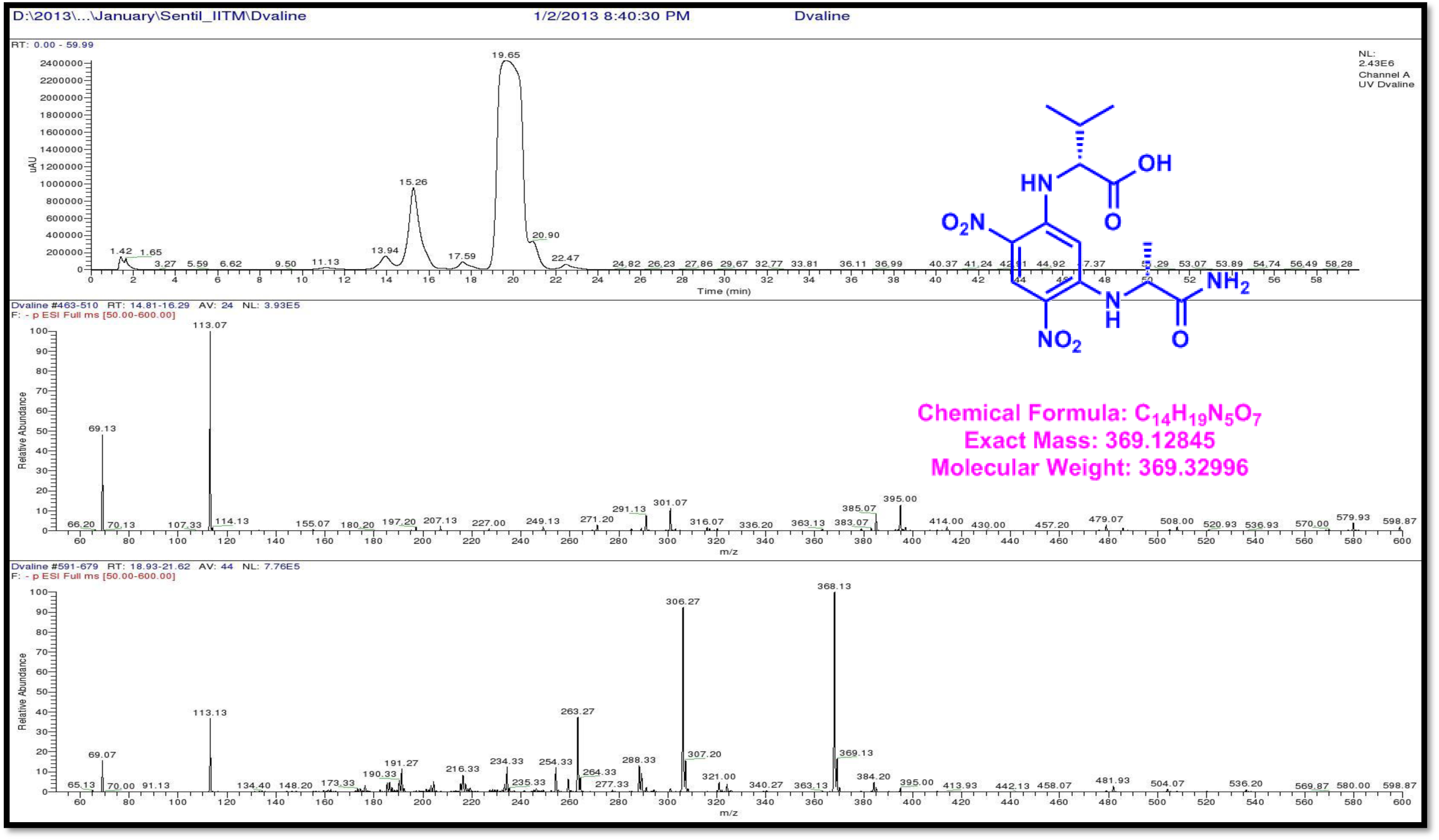
LCMS analysis of Standard L-FDAA-D-Valine (Negative mode)

**Figure S136.**
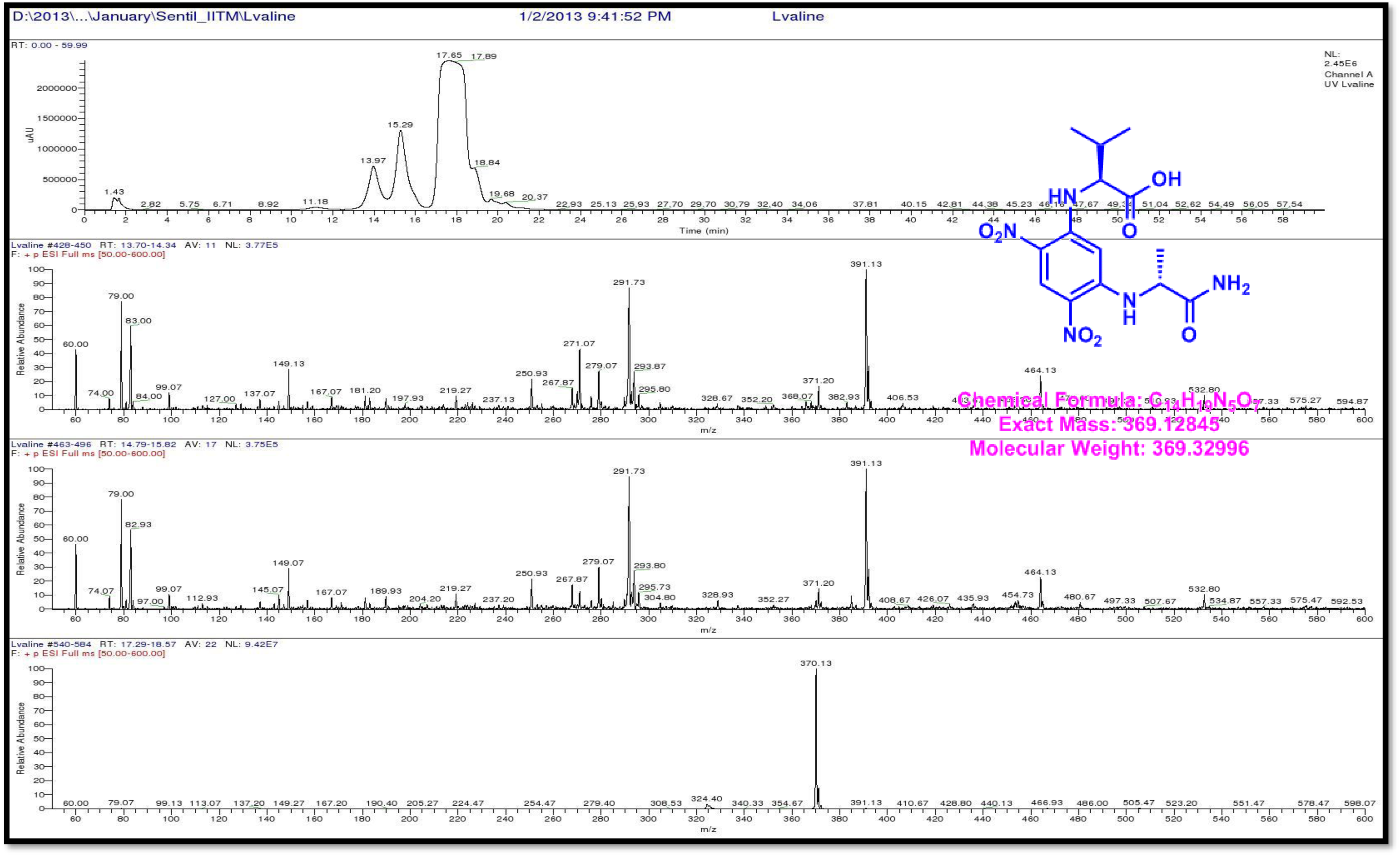
LCMS analysis of Standard L-FDAA-L-Valine (Positive mode)

**Figure S137.**
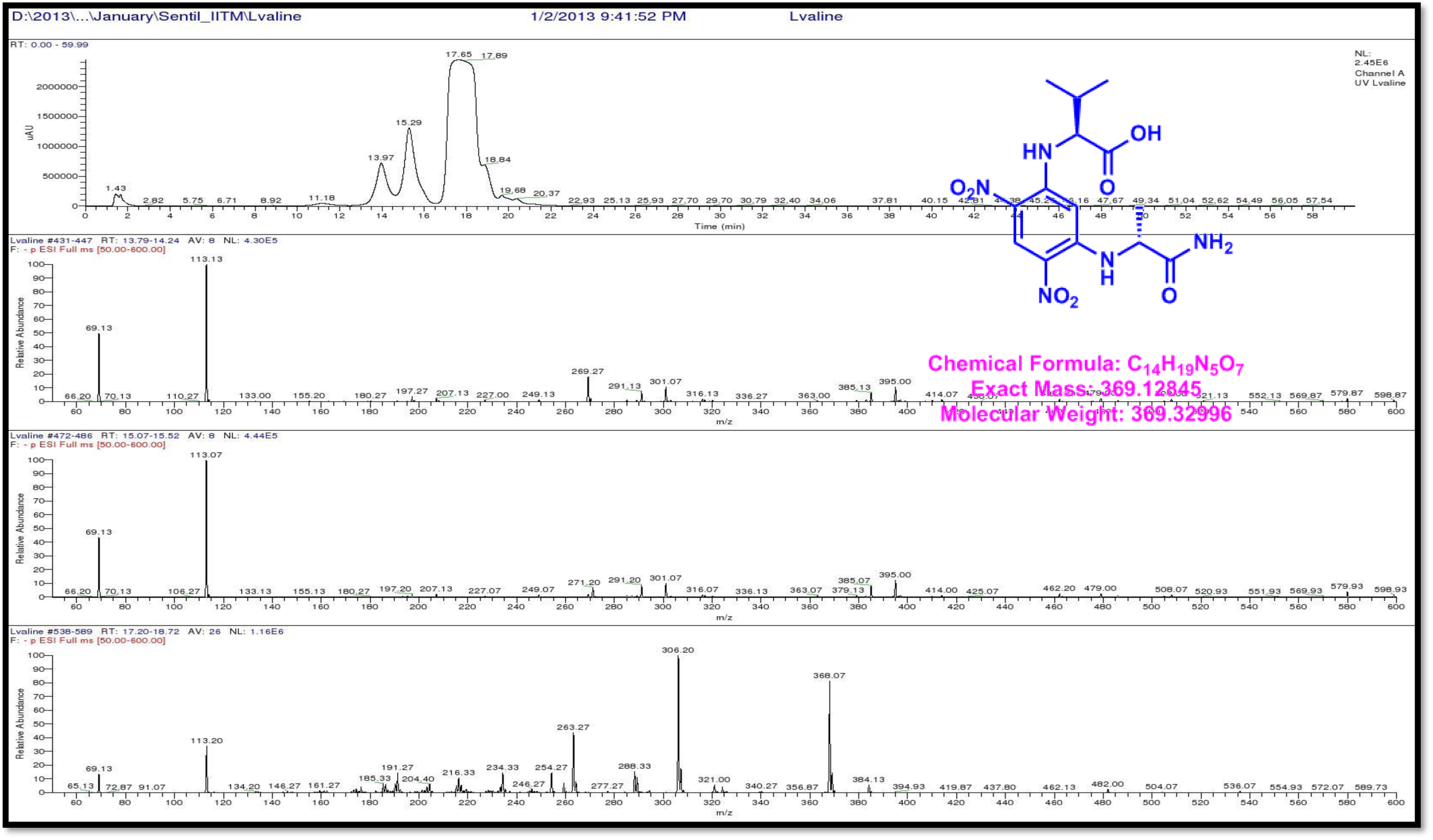
LCMS analysis of Standard L-FDAA-L-Valine (Negative mode)

**Figure S138.**
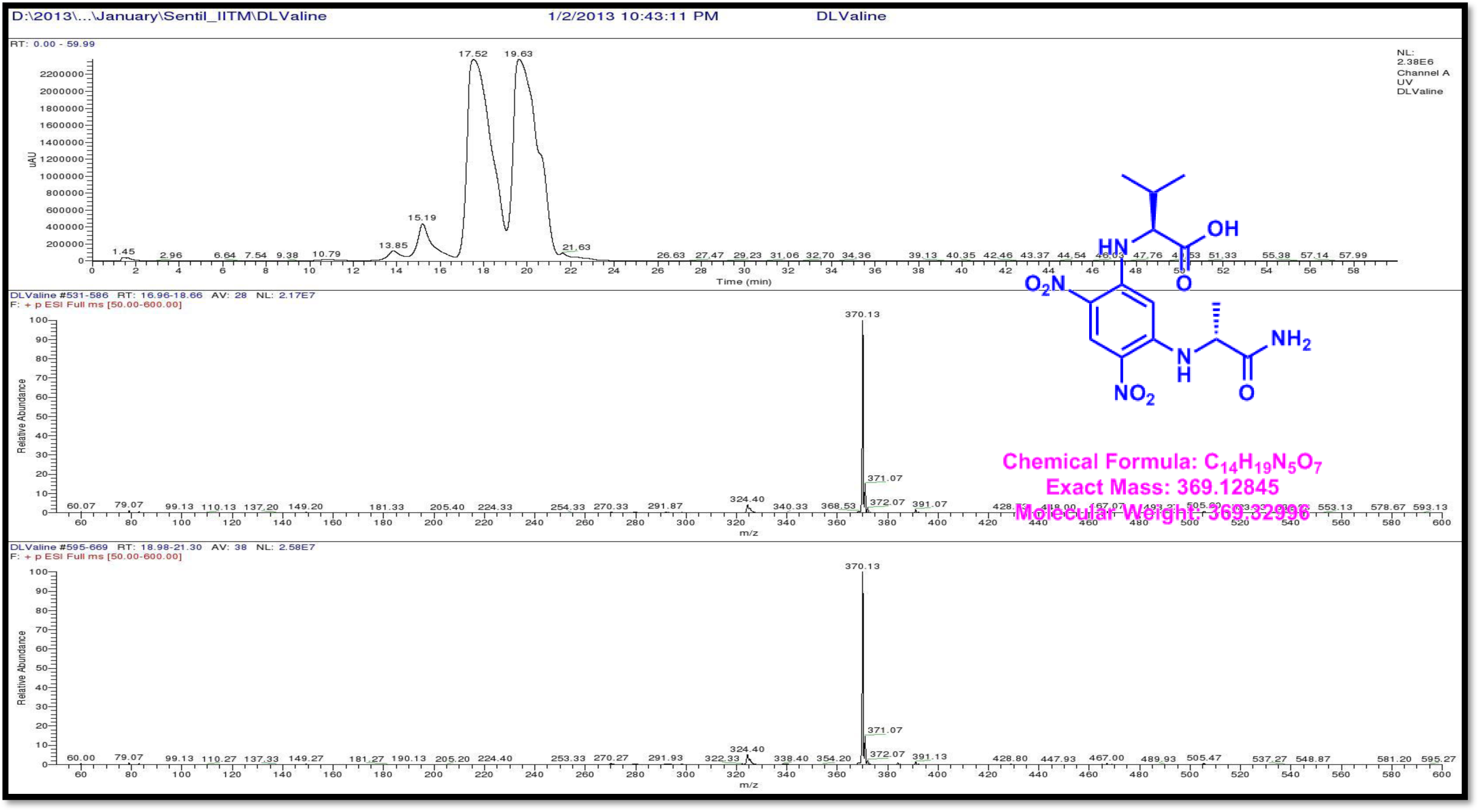
LCMS analysis of Standard L-FDAA-D/L-Valine (Positive mode)

**Figure S139.**
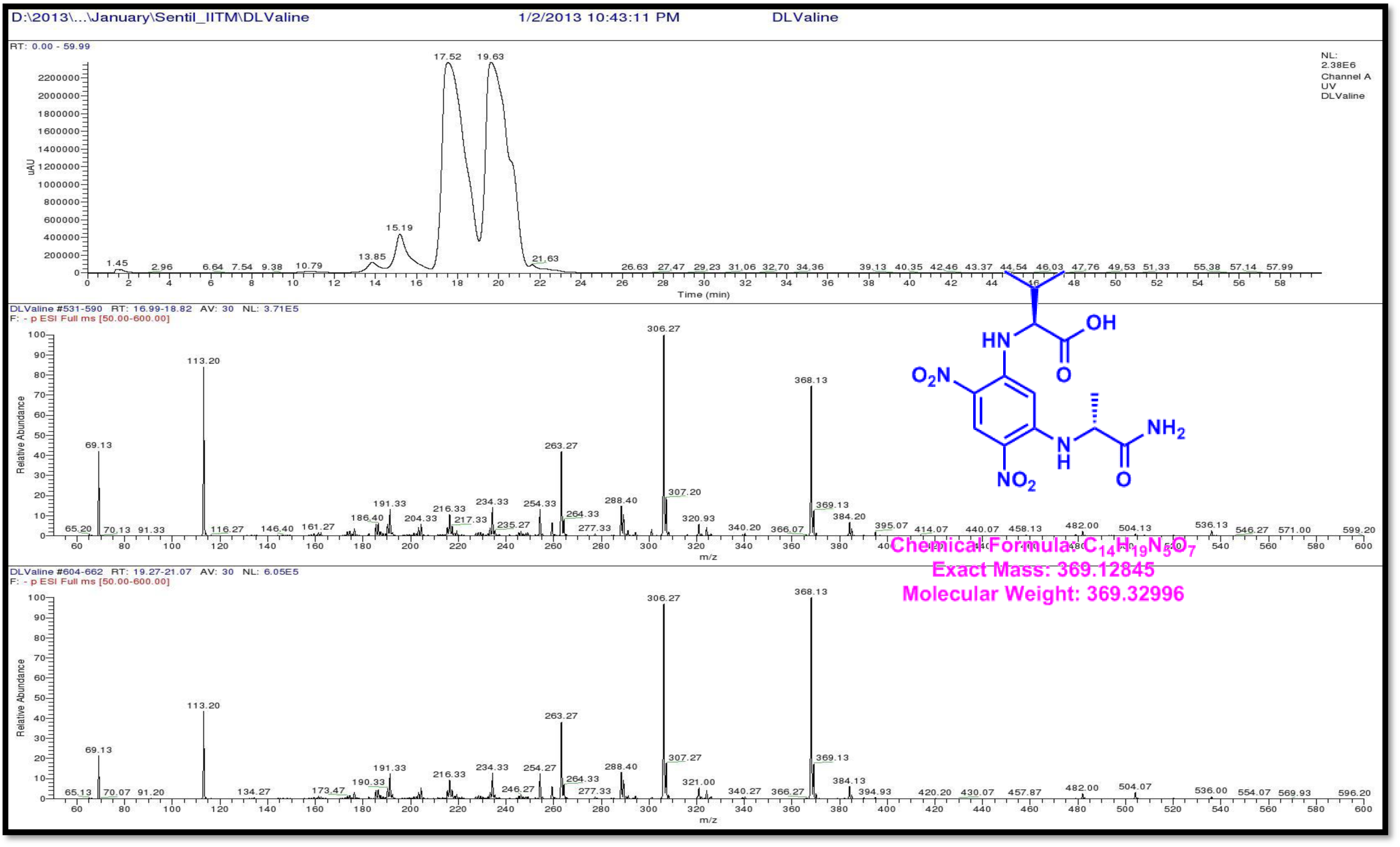
LCMS analysis of Standard L-FDAA-D/L-Valine (Negative mode)

**Figure S140.**
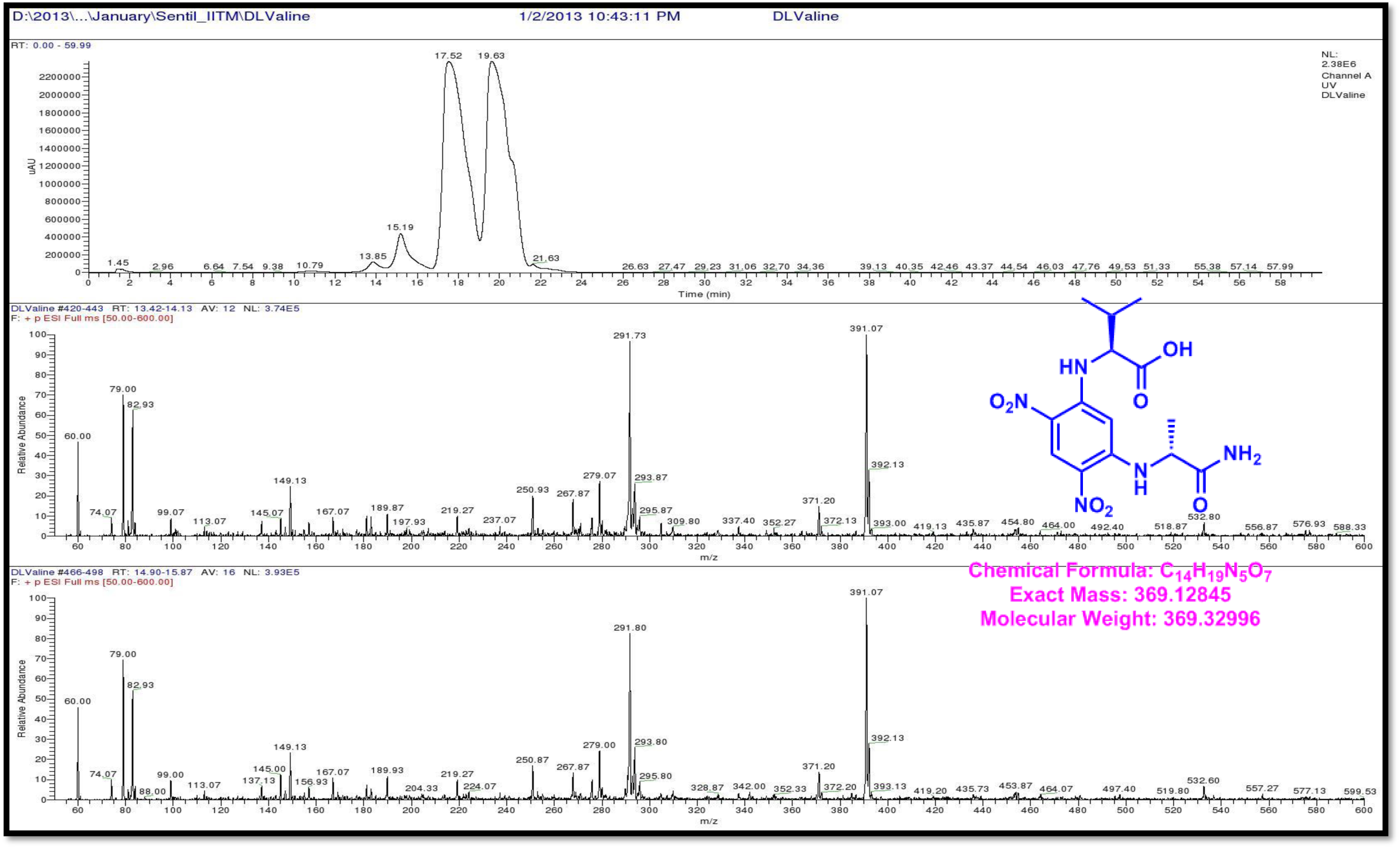
LCMS analysis of Standard L-FDAA-D/L-Valine (Positive mode)

**Figure S141.**
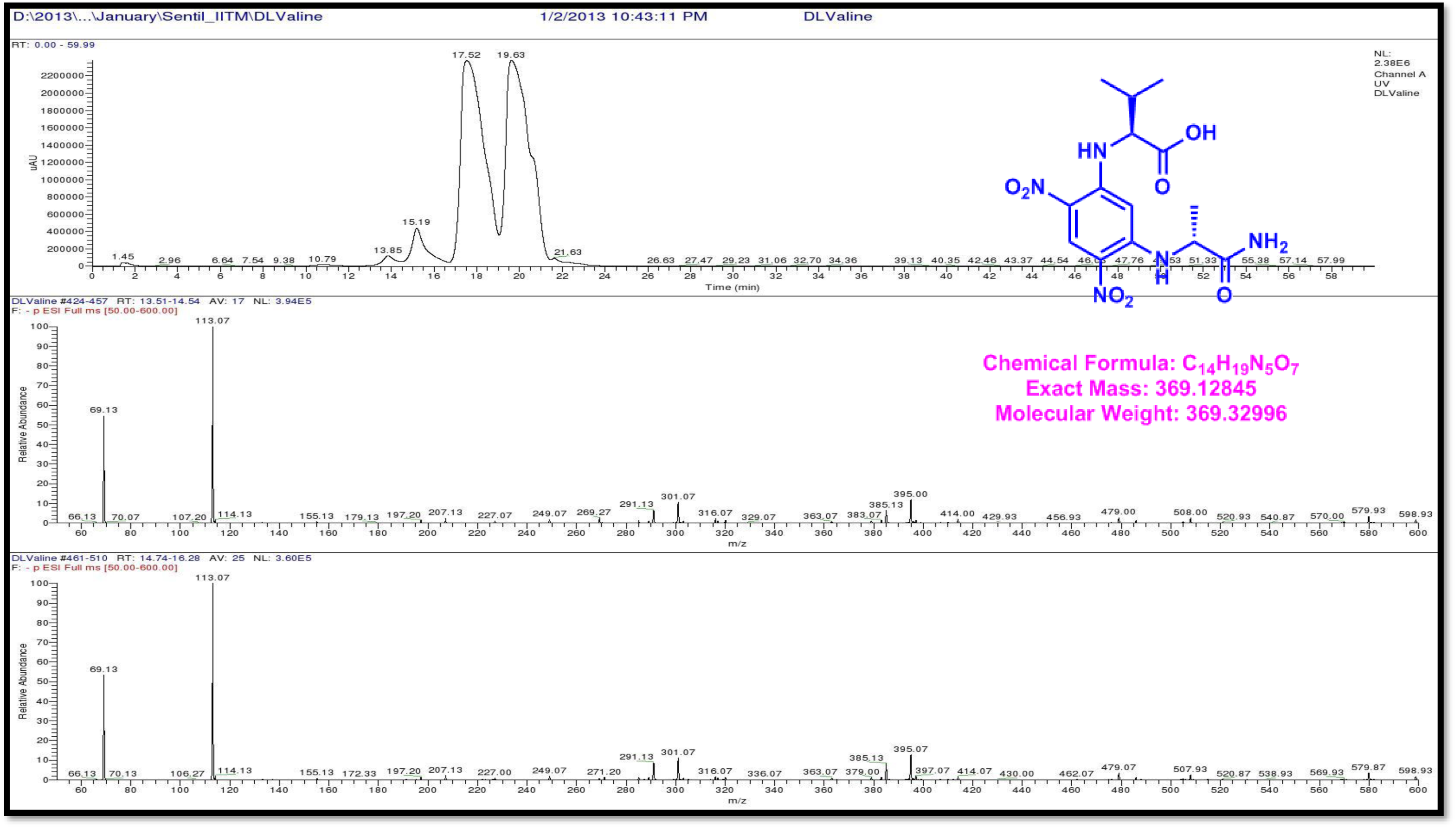
LCMS analysis of Standard L-FDAA-D/L-Valine (Negative mode)

**Figure S142.**
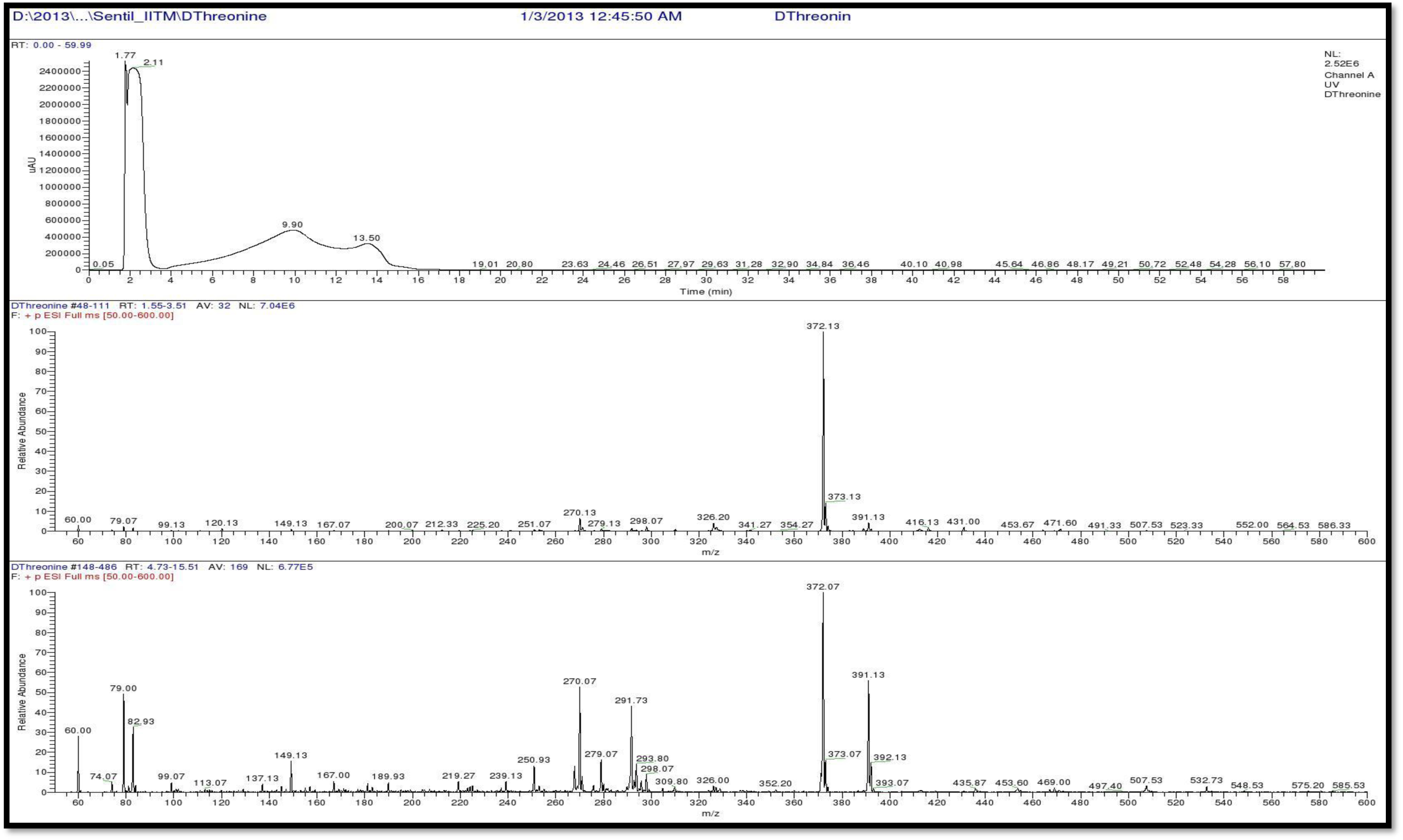
LCMS analysis of Standard L-FDAA-D-Threonine (Positive mode)

**Figure S143.**
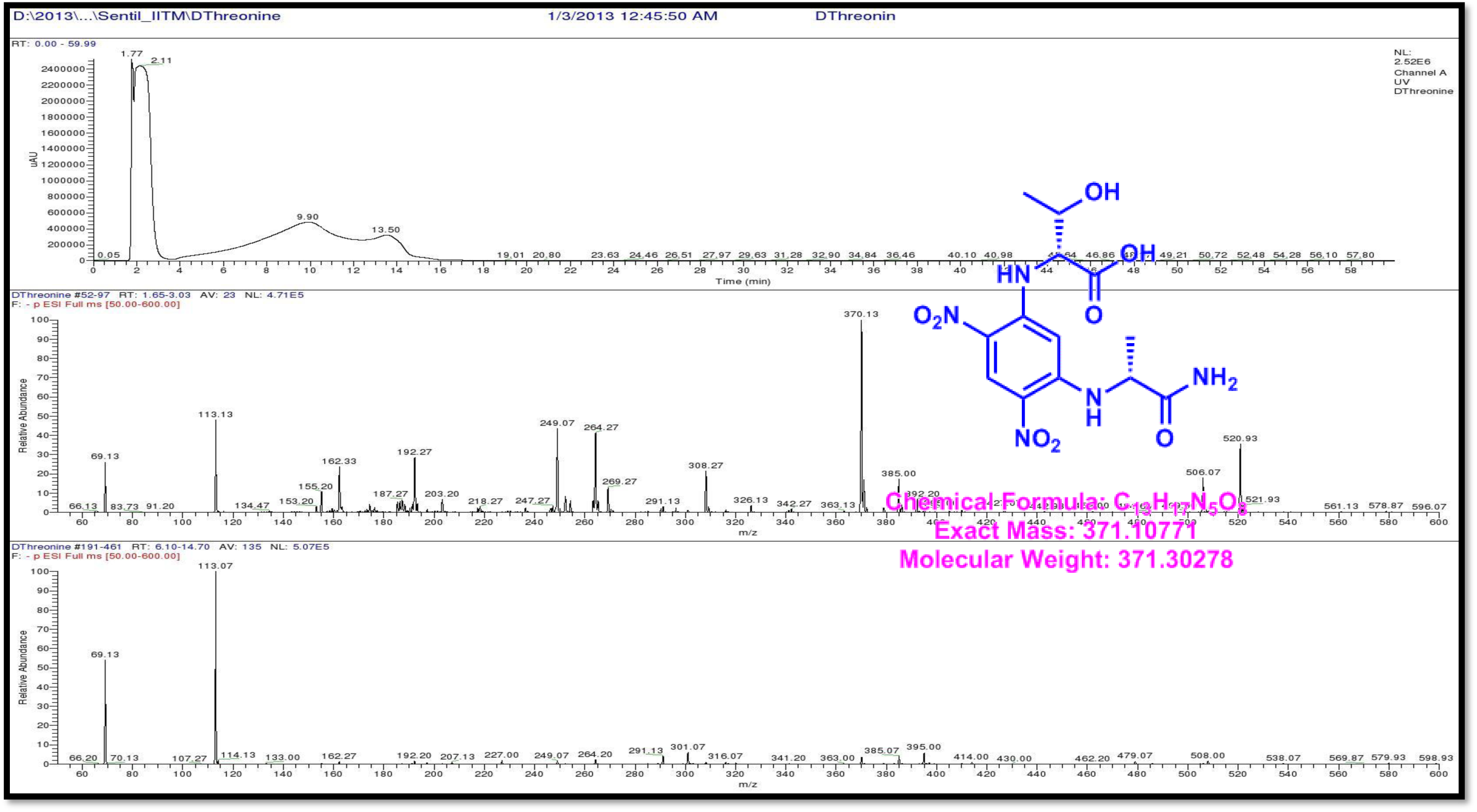
LCMS analysis of Standard L-FDAA-D-Threonine (Negative mode)

**Figure S144.**
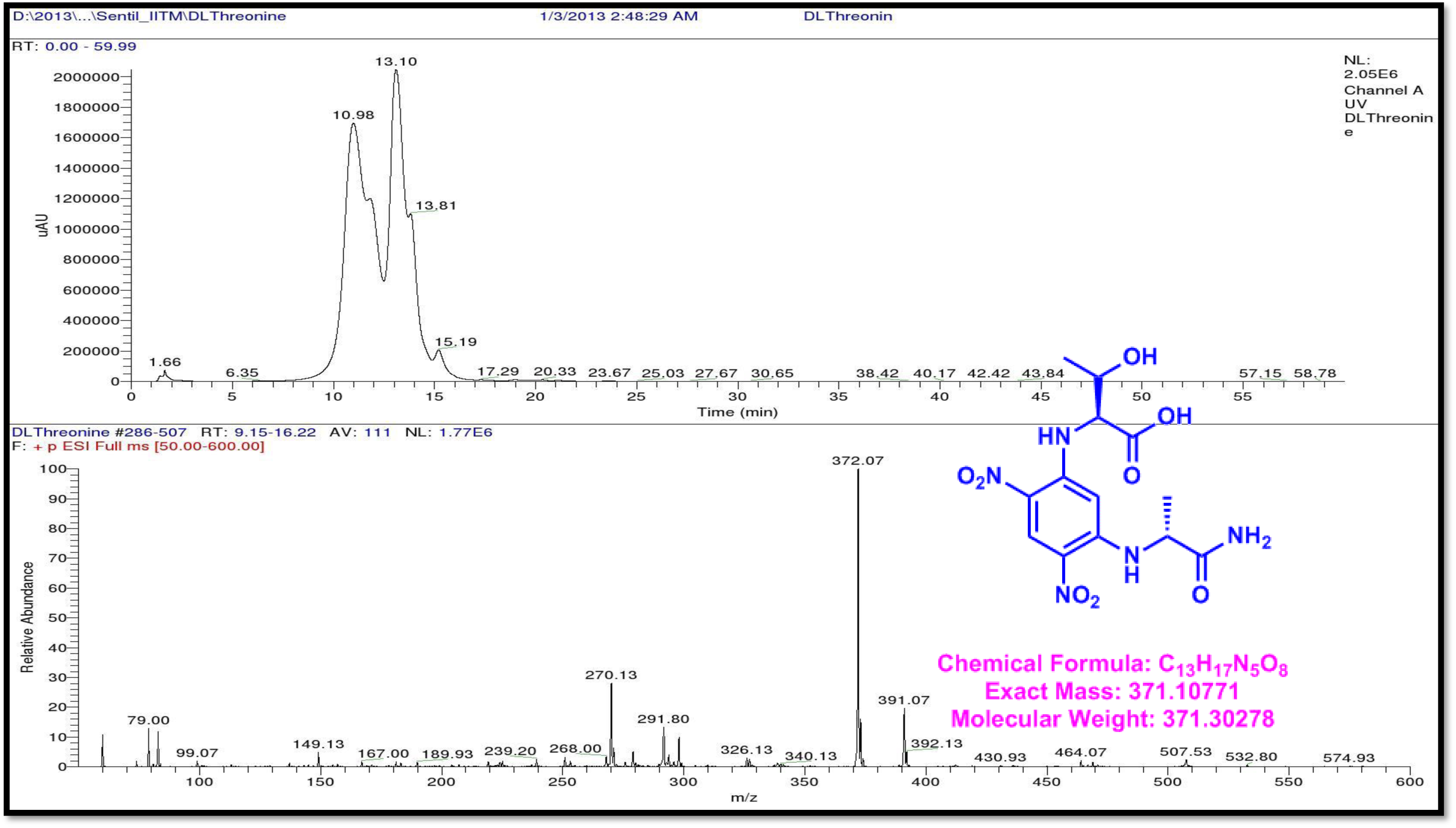
LCMS analysis of Standard L-FDAA-D/L Threonine (Positive mode)

**Figure S145.**
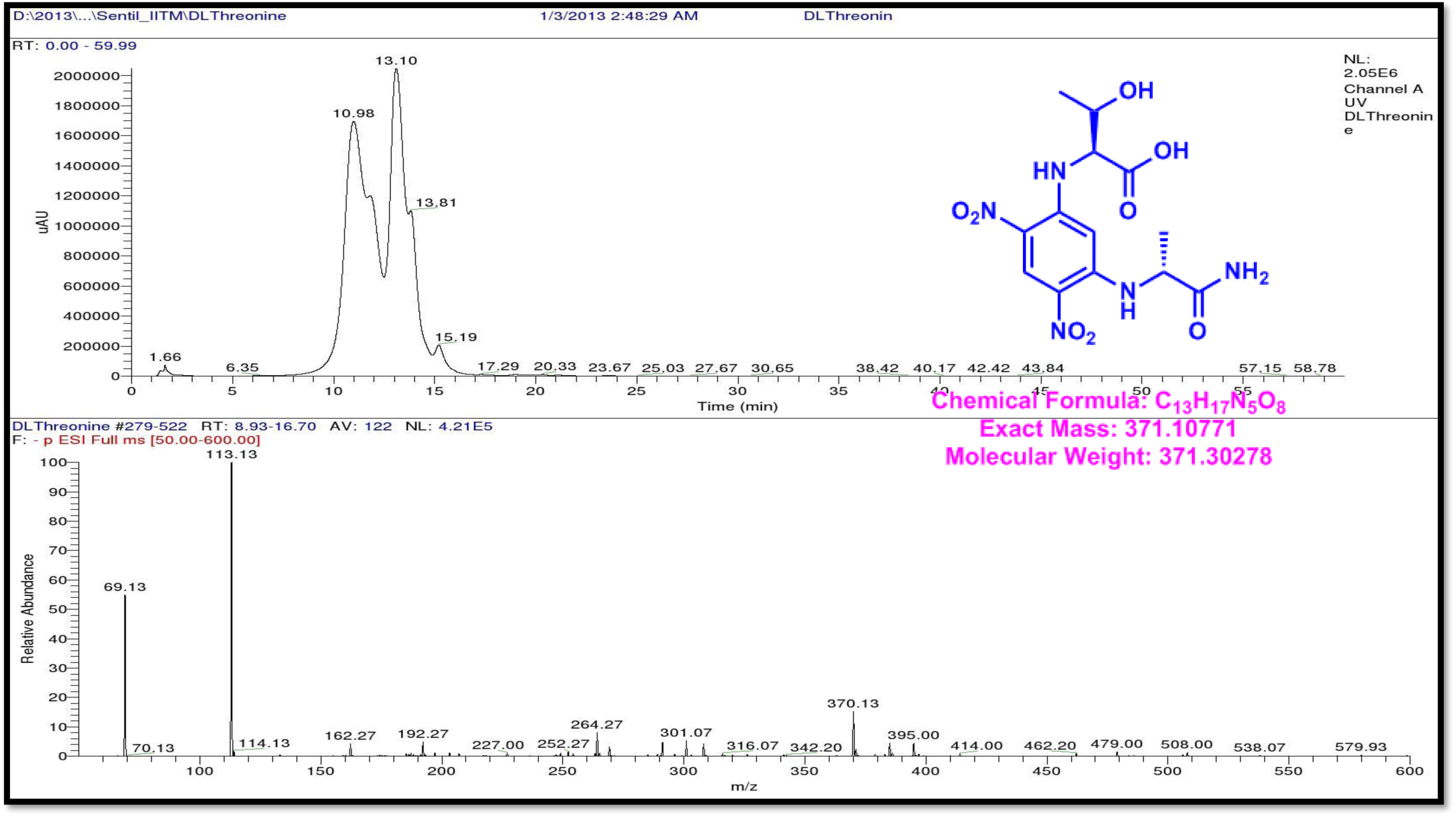
LCMS analysis of Standard L-FDAA-D/L Threonine (Negative mode)

**Figure S146.**
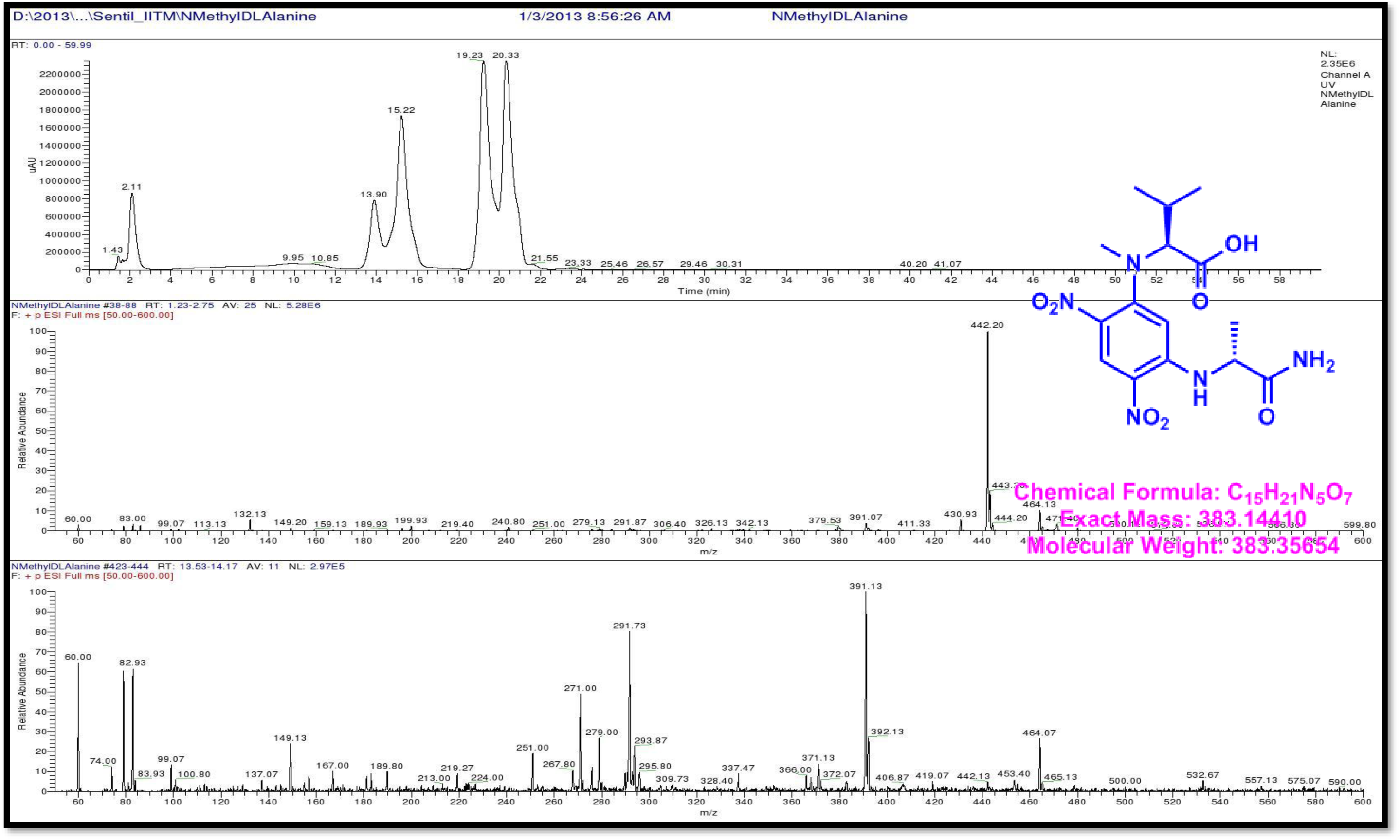
LCMS analysis of Standard L-FDAA-D/L N-Methyl valine (Positive mode)

**Figure S147.**
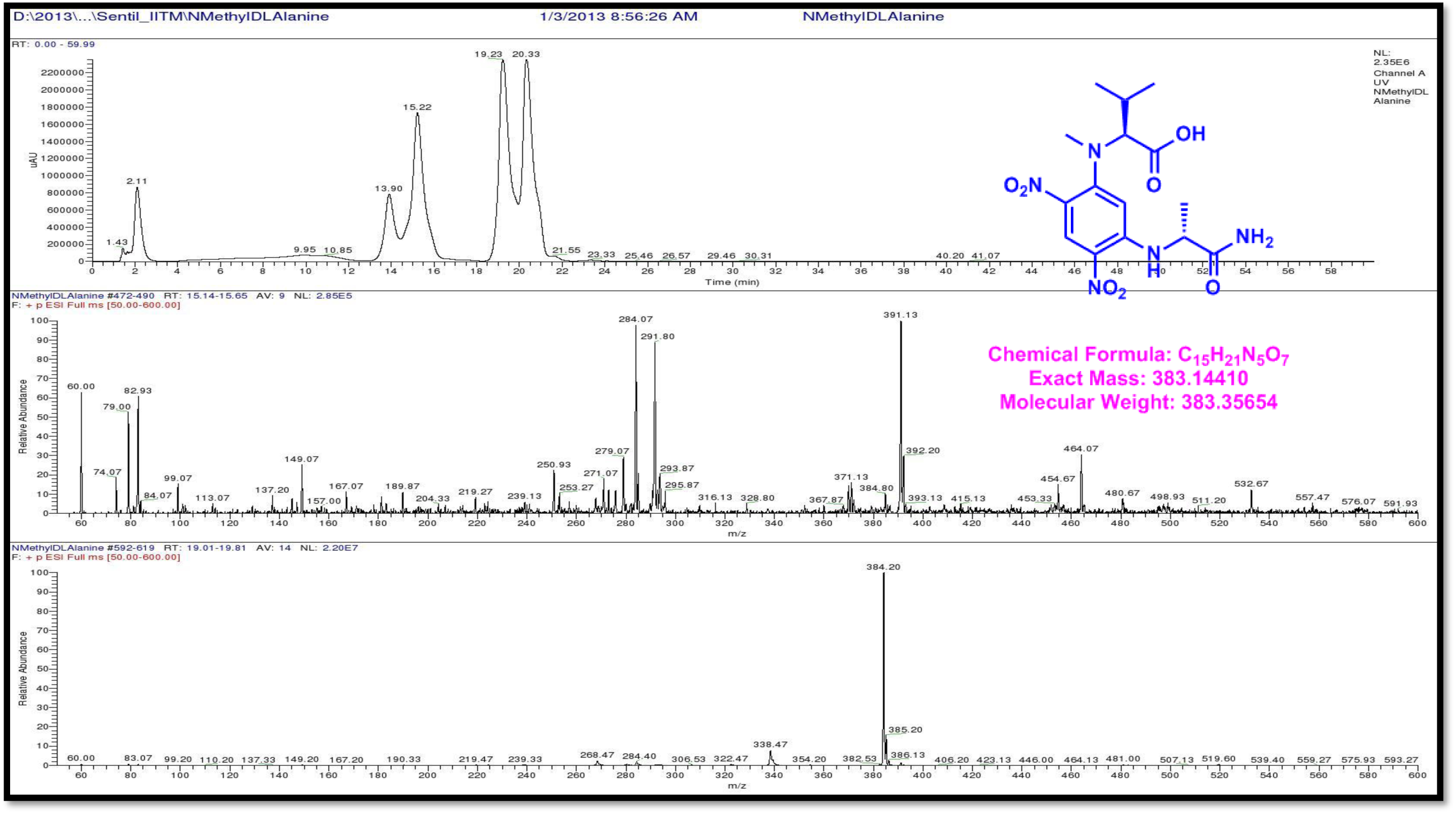
LCMS analysis of Standard L-FDAA-D/L N-Methyl Valine (Positive mode)

**Figure S148.**
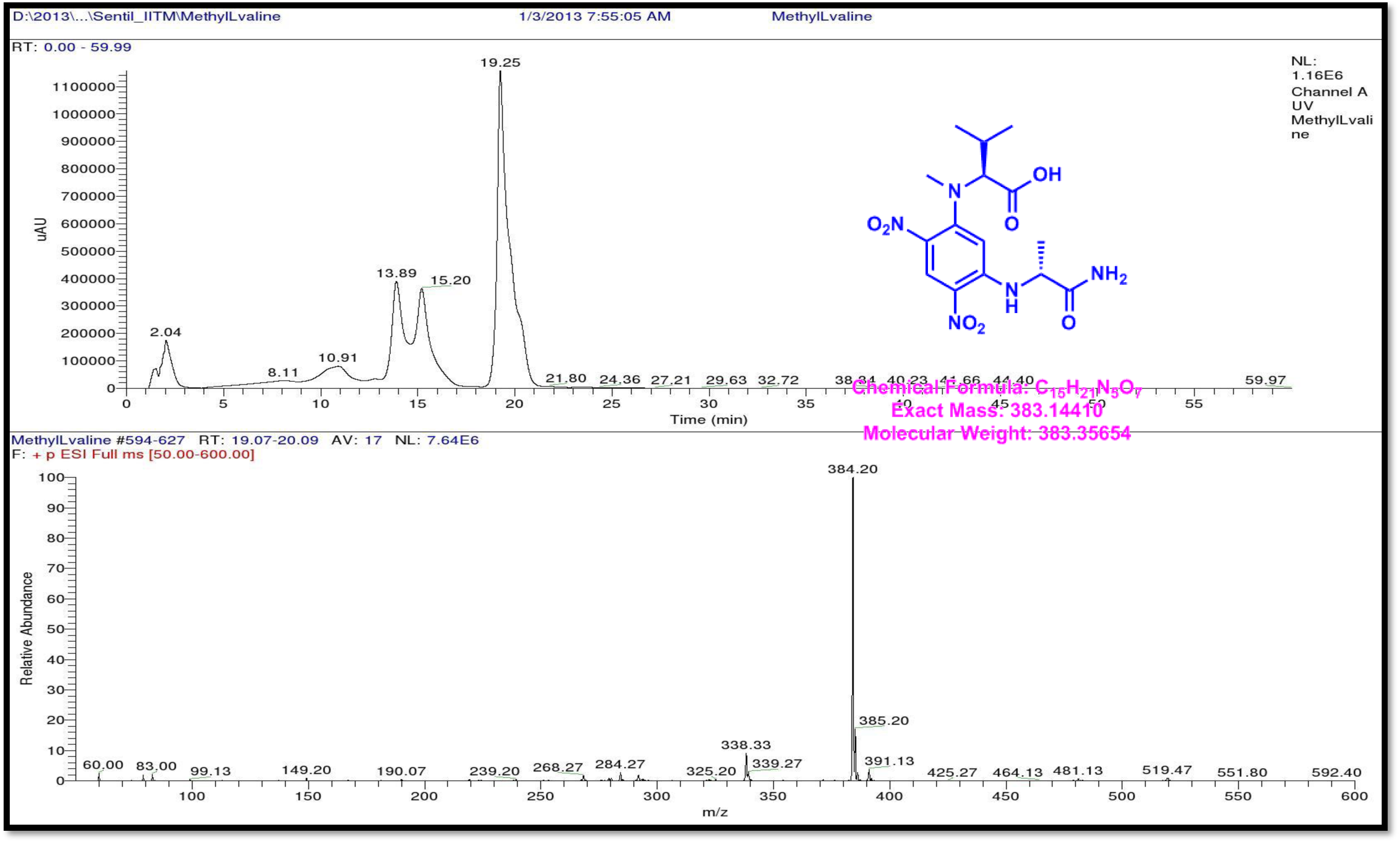
LCMS analysis of Standard L-FDAA-L N-Methyl valine (Positive mode)

**Figure S149.**
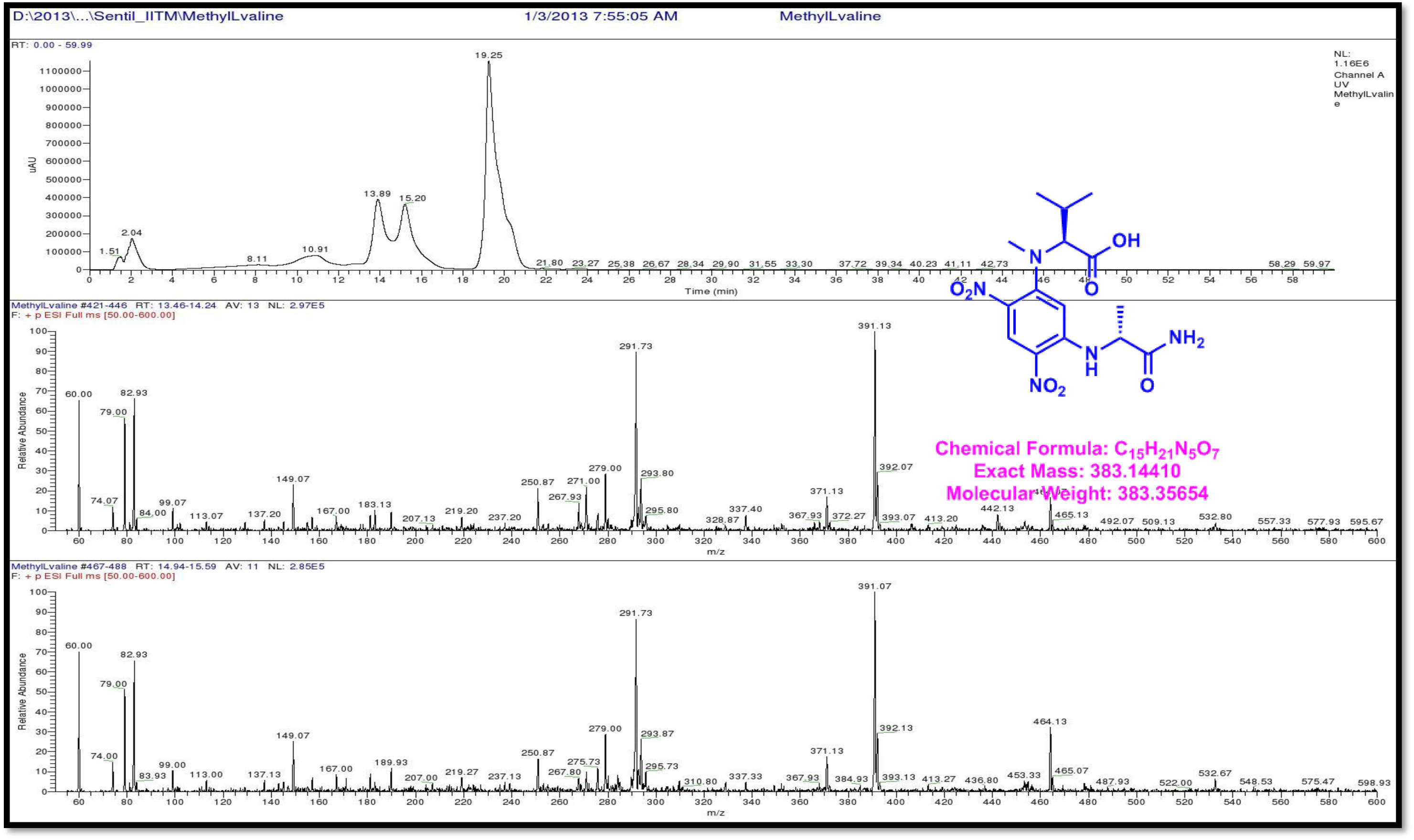
LCMS analysis of Standard L-FDAA-L- N-Methyl valine (Positive mode)

**Figure S150.**
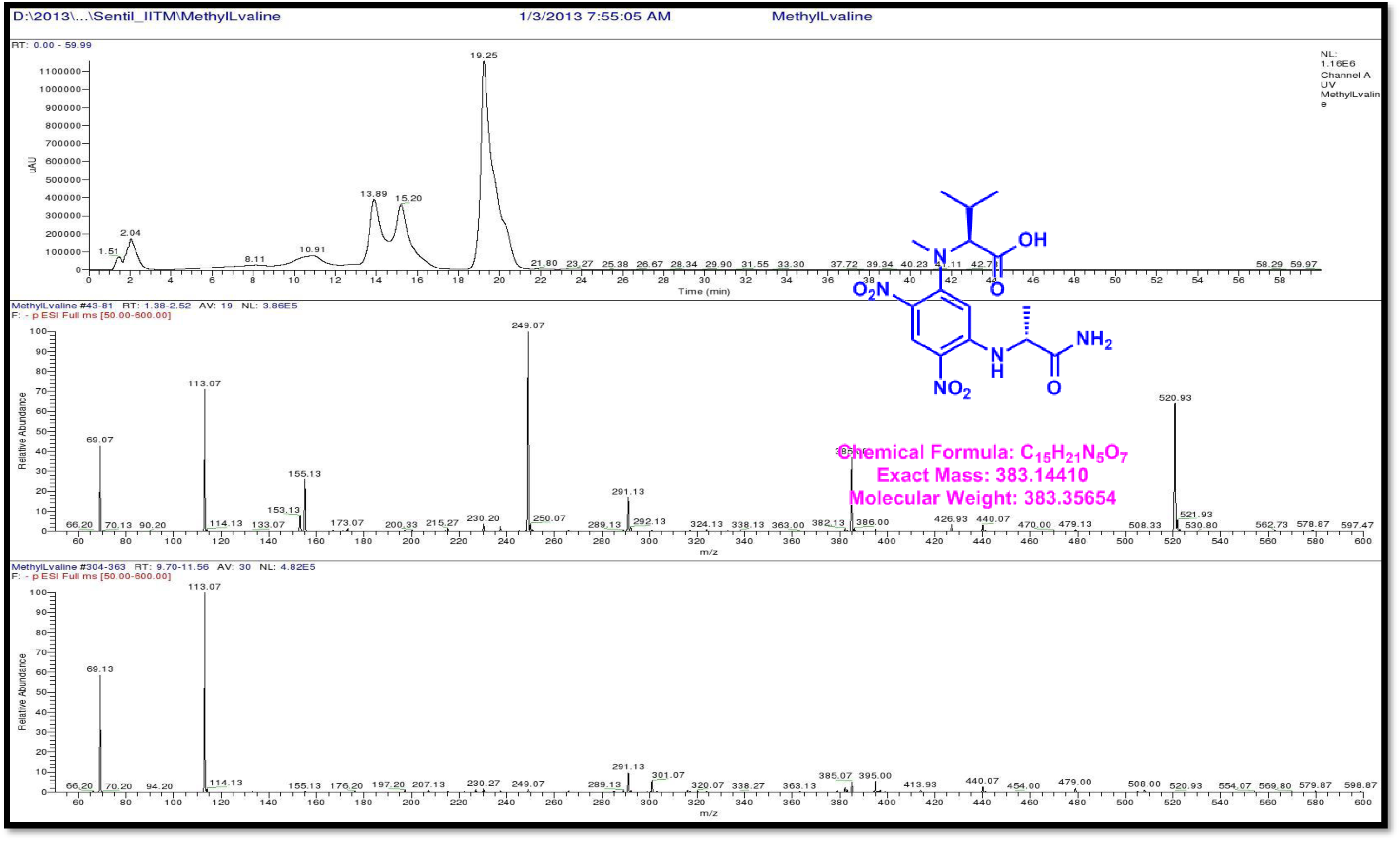
LCMS analysis of Standard L-FDAA-L N-Methyl Valine (Positive mode)

**Figure S151.**
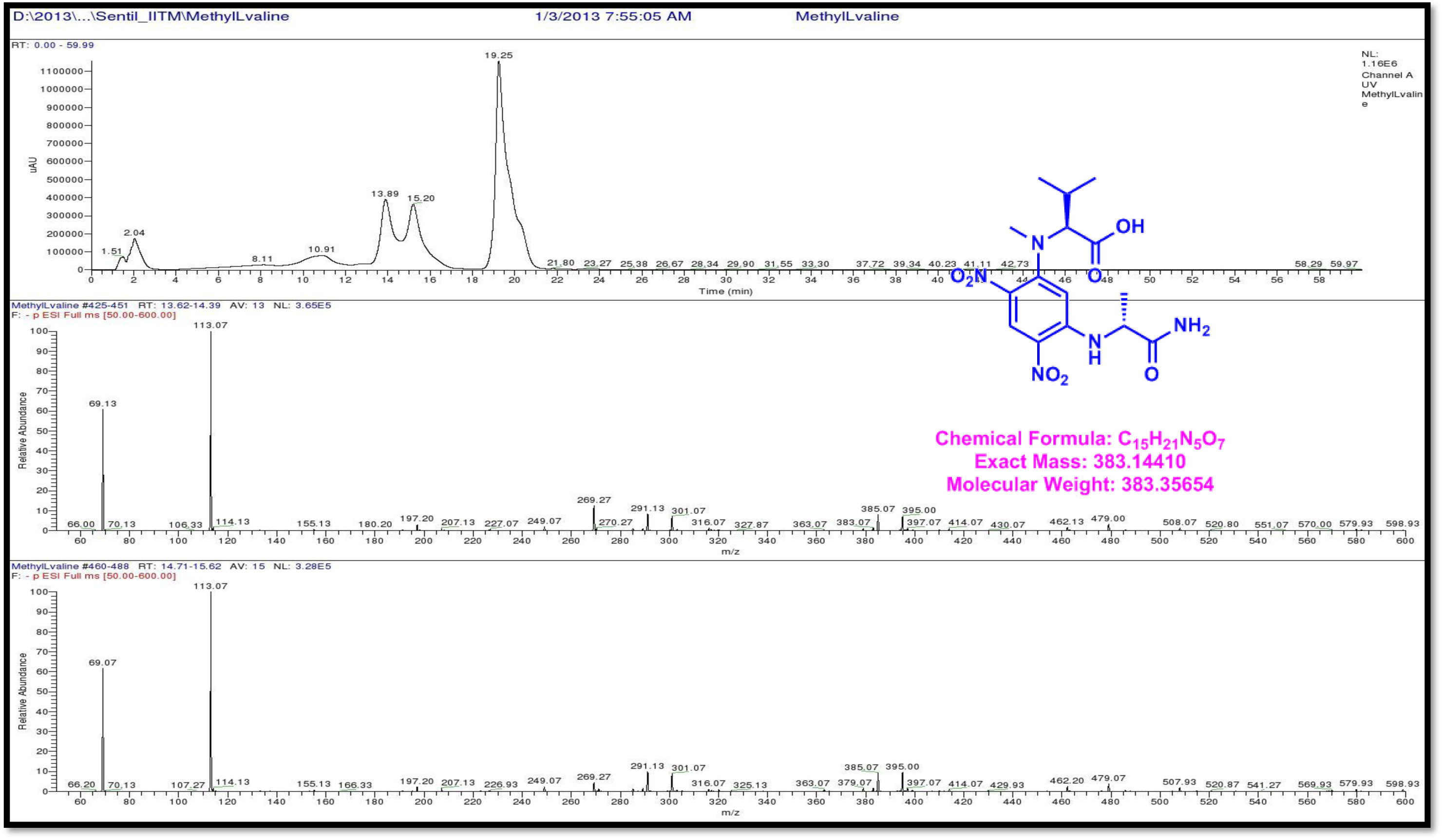
LCMS analysis of Standard L-FDAA-L N-Methyl Valine (Positive mode)

**Figure S152.**
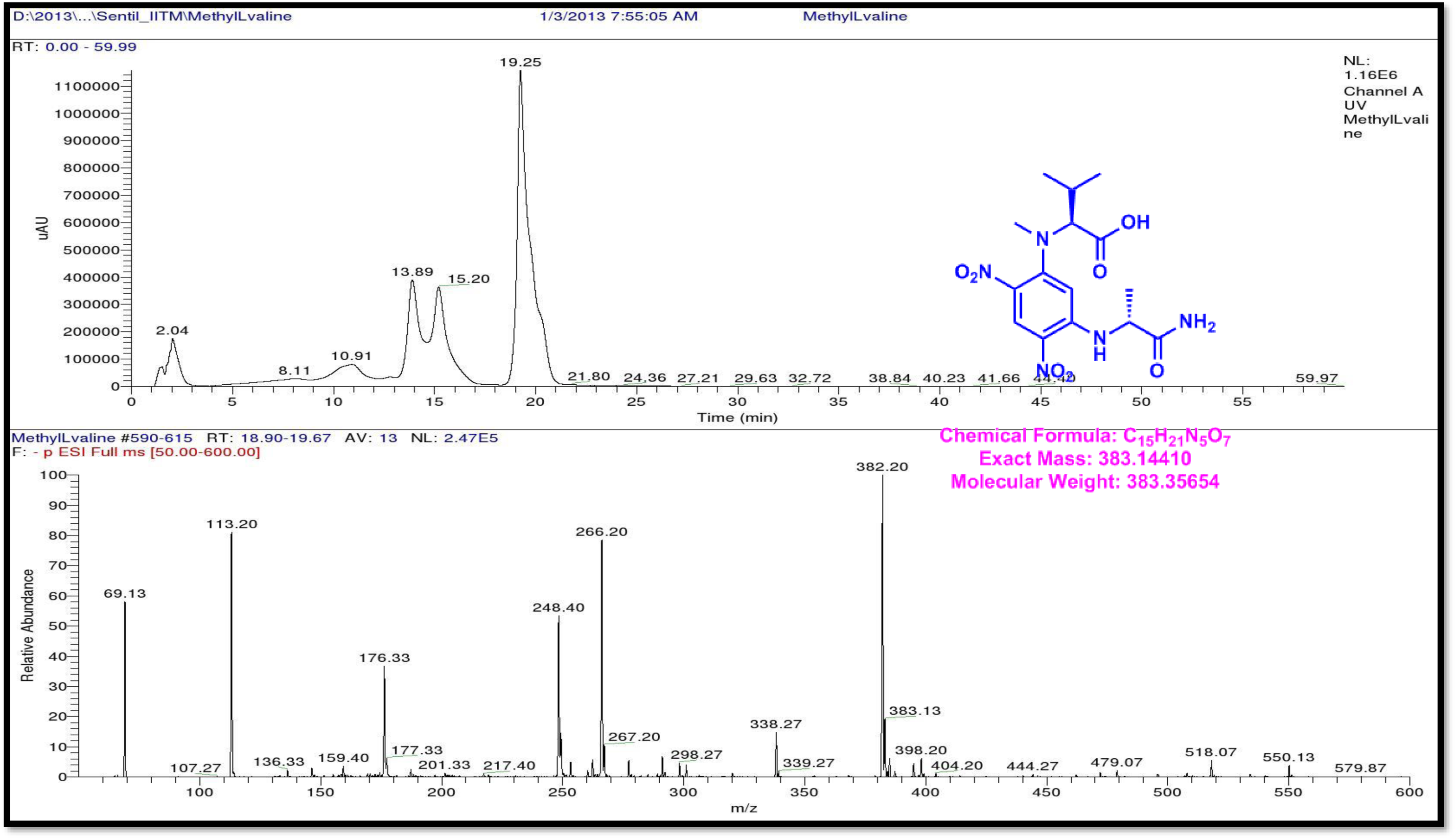
LCMS analysis of Standard L-FDAA-L N-Methyl Valine (Negative mode)

**Figure S153.**
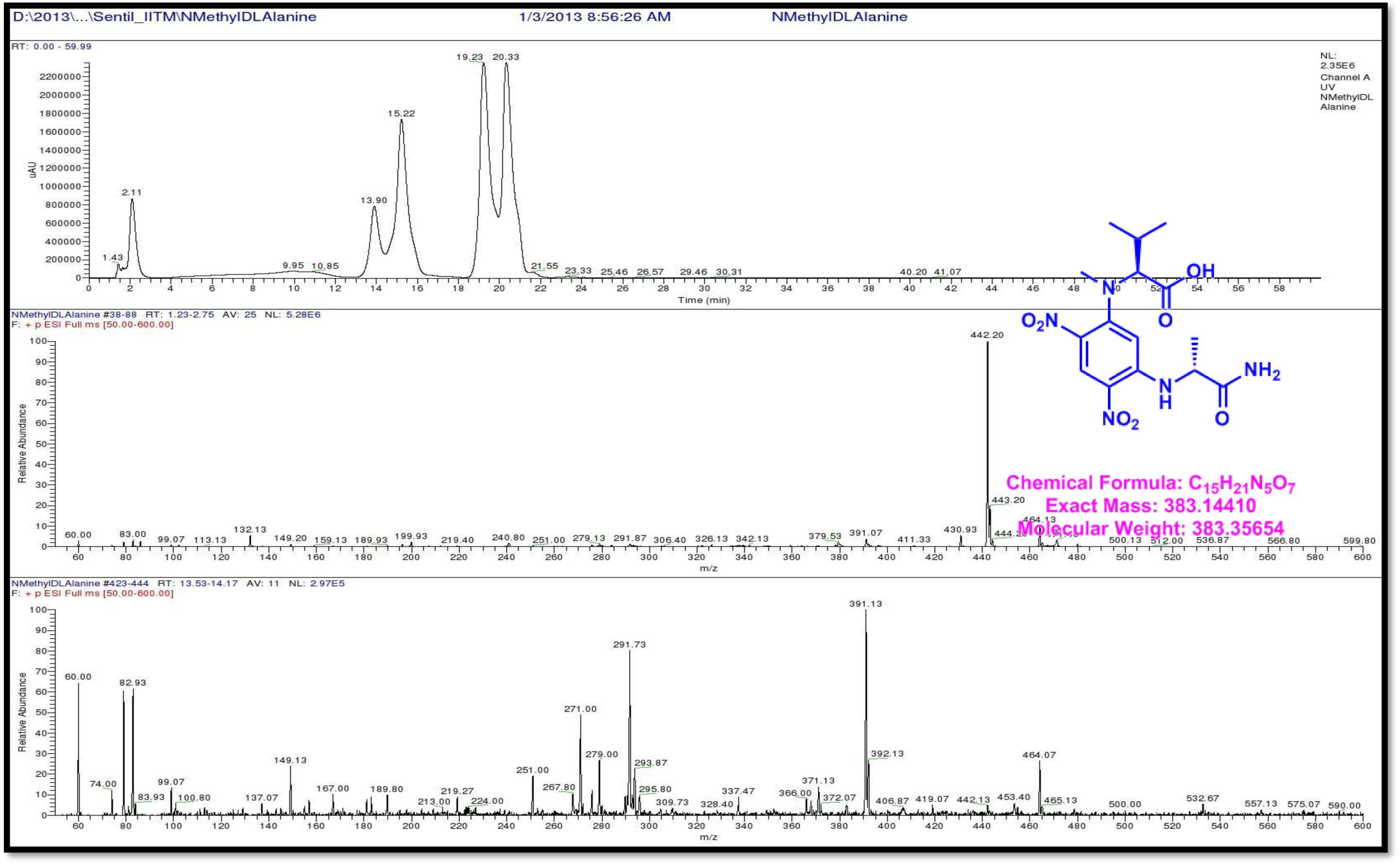
LCMS analysis of Standard L-FDAA-D/L N-Methyl Valine (Positive mode)

**Figure S154.**
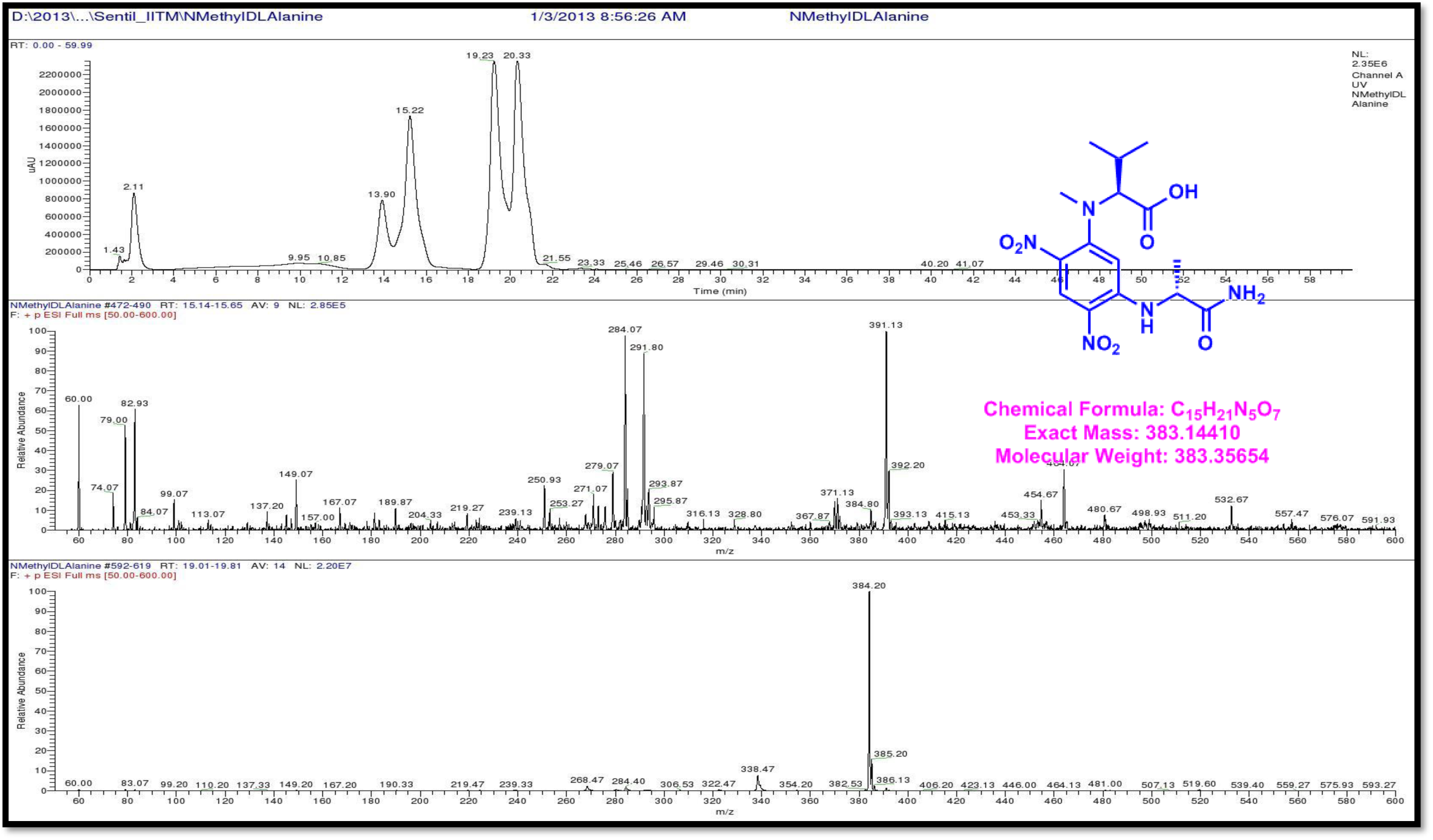
LCMS analysis of Standard L-FDAA-L N-Methyl Valine (Positive mode)

**Figure S155.**
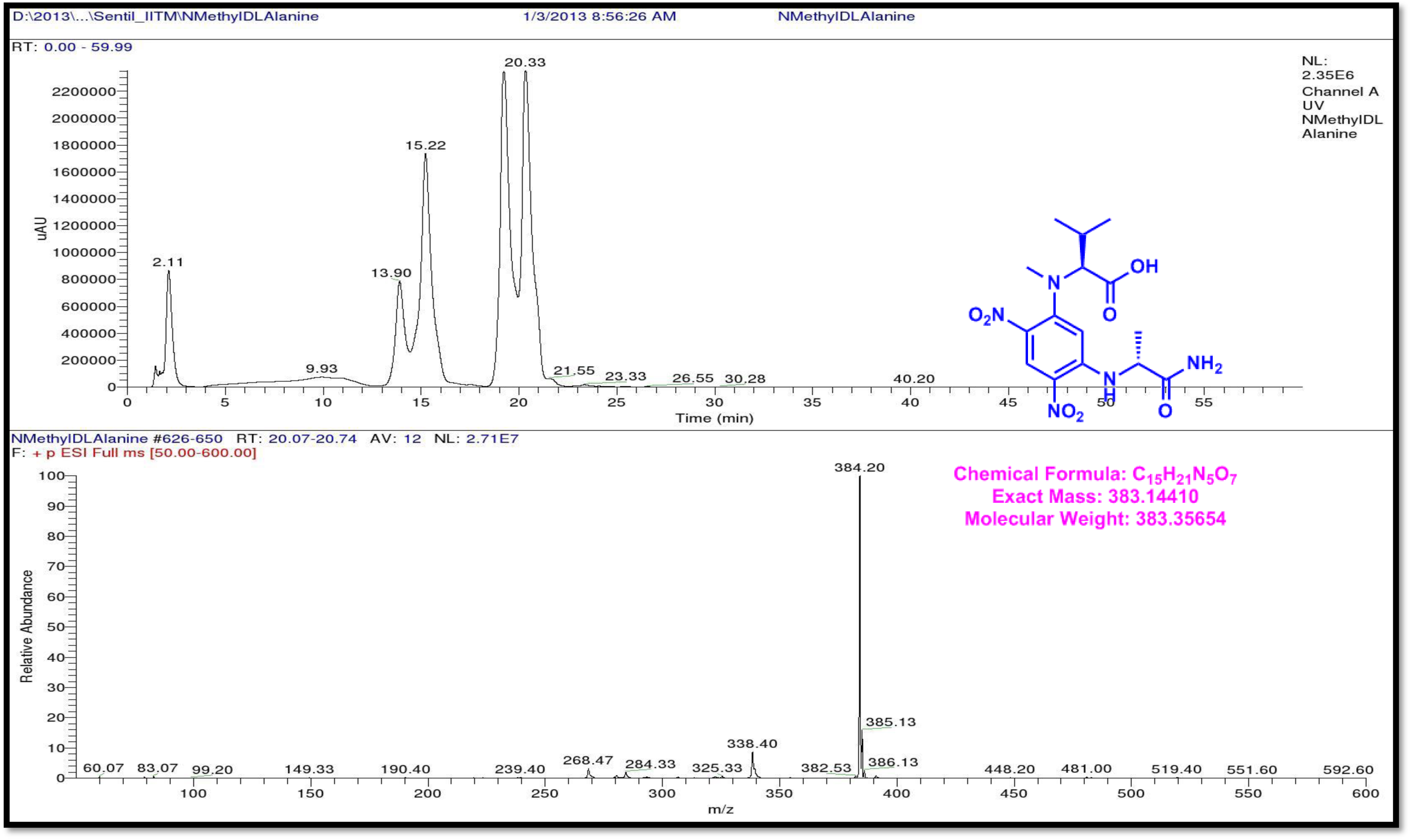
LCMS analysis of Standard L-FDAA-D/L N-Methyl Valine (Positive mode)

**Figure S156.**
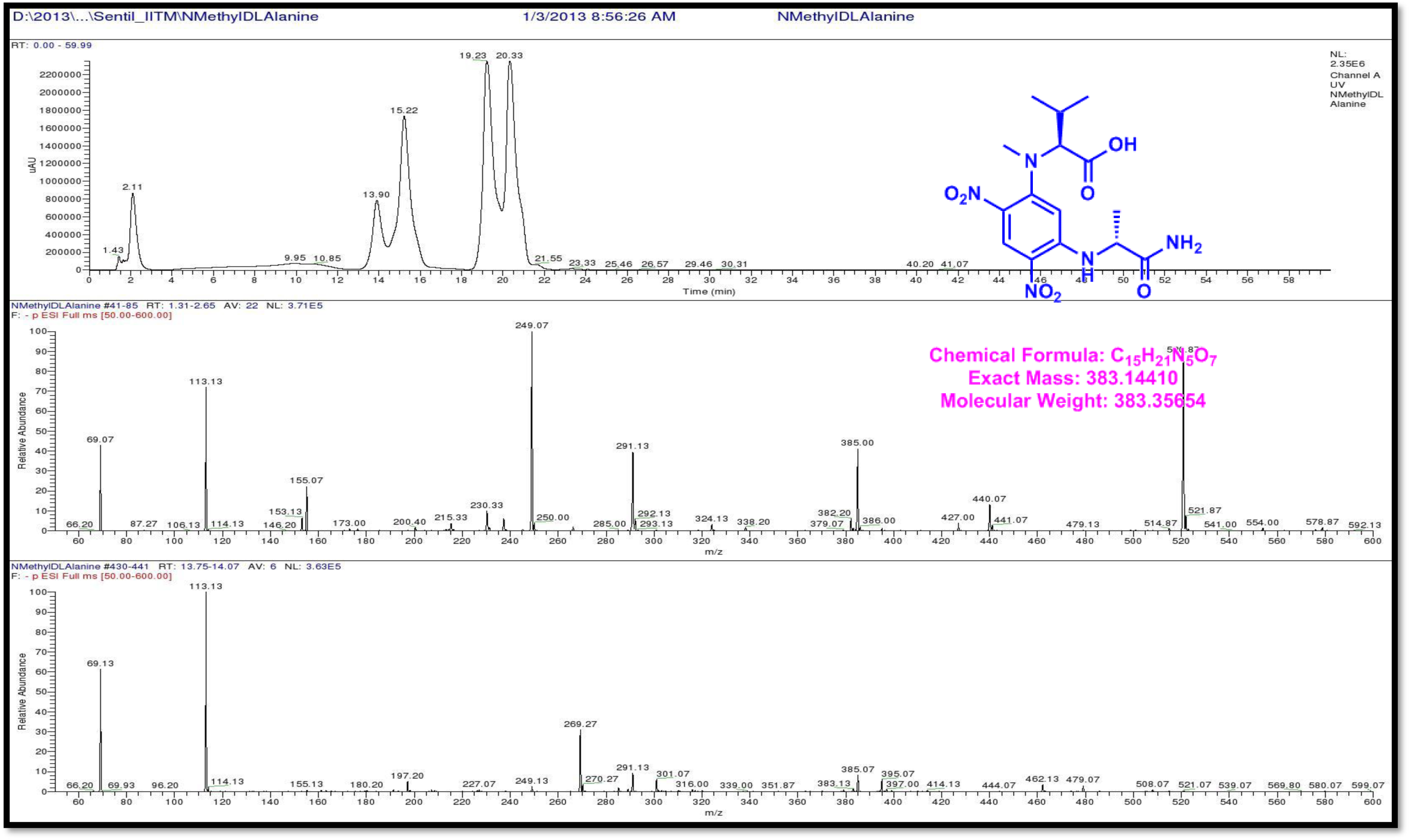
LCMS analysis of Standard L-FDAA-D/L N-Methyl Valine (Negative mode)

**Figure S157.**
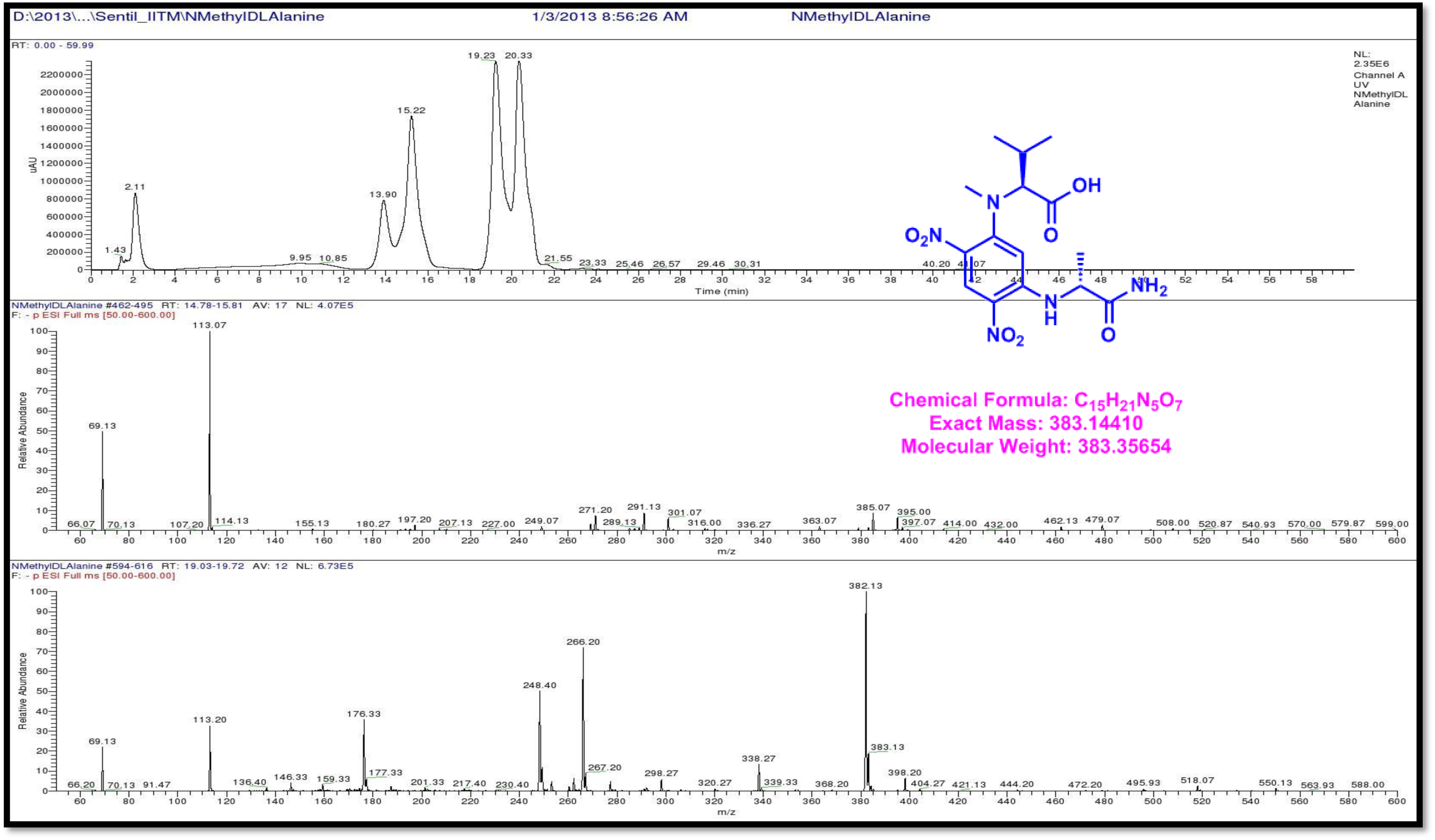
LCMS analysis of Standard L-FDAA-D/L N-Methyl Valine (Negative mode)

**Figure S158.**
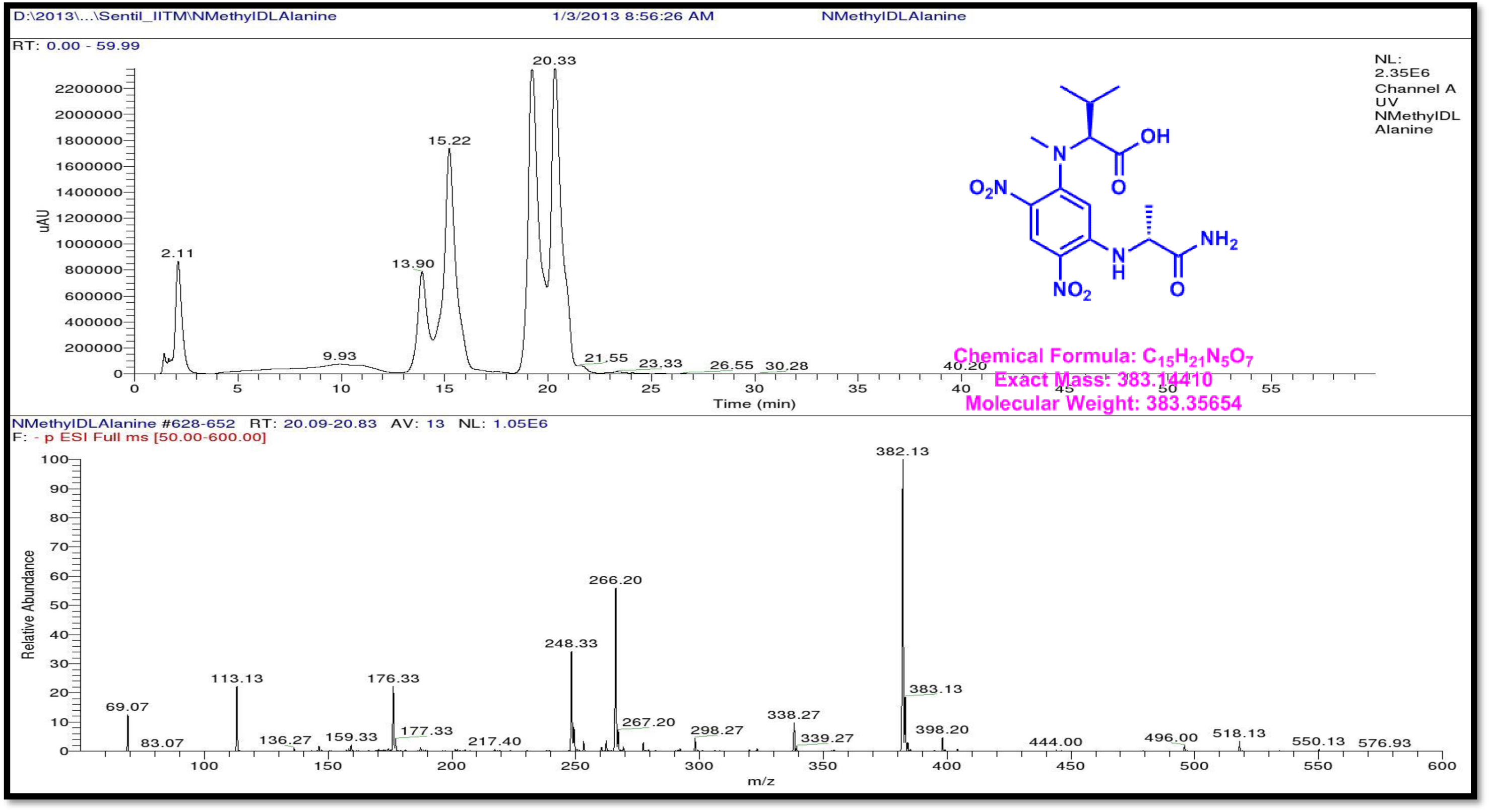
LCMS analysis of Standard L-FDAA-D/L N-Methyl Valine (Negative mode)

**Figure S159.**
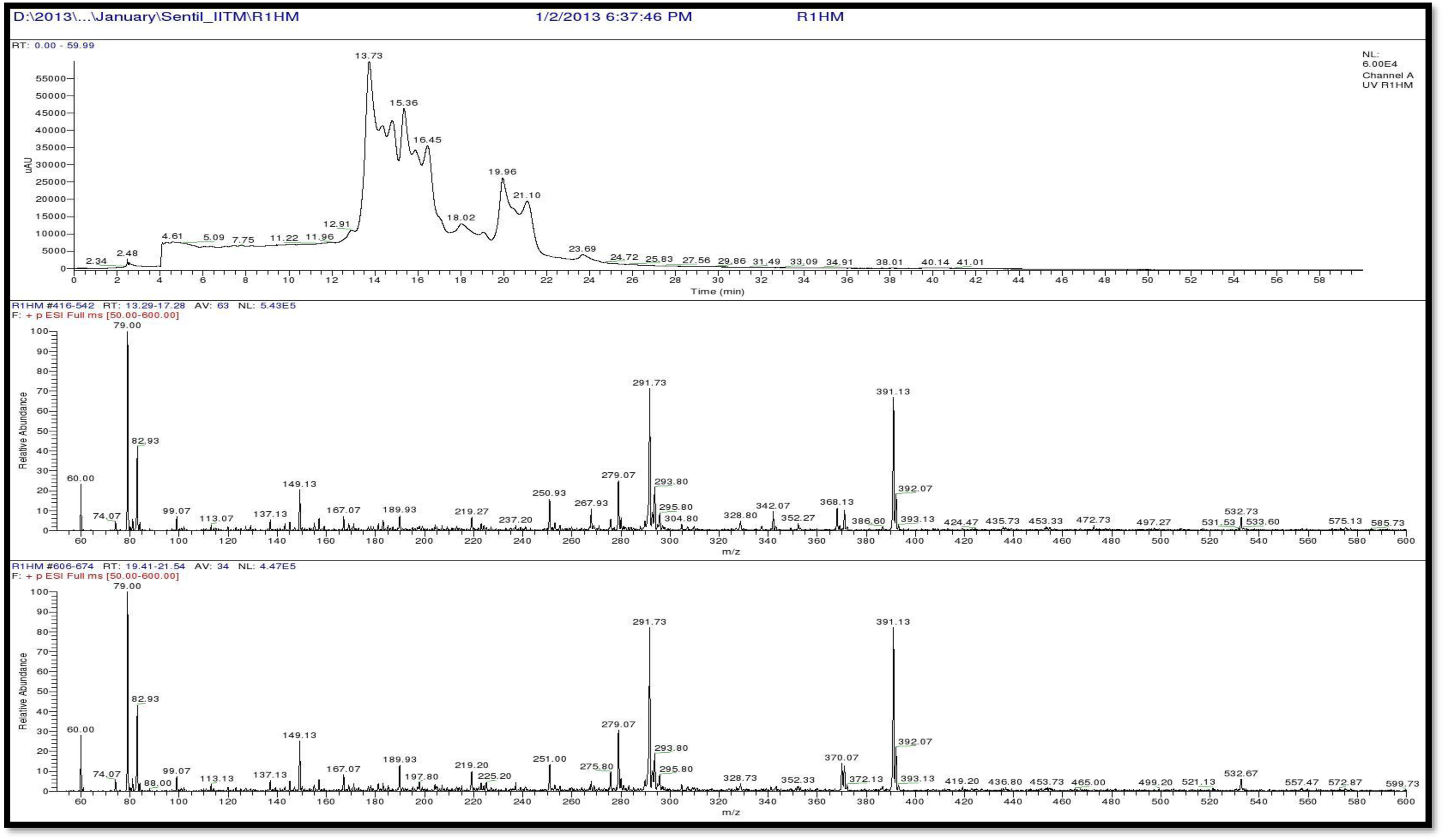
LCMS analysis of L-FDAA derivatives of Transitmycin (R1) (Positive mode)

**Figure S160.**
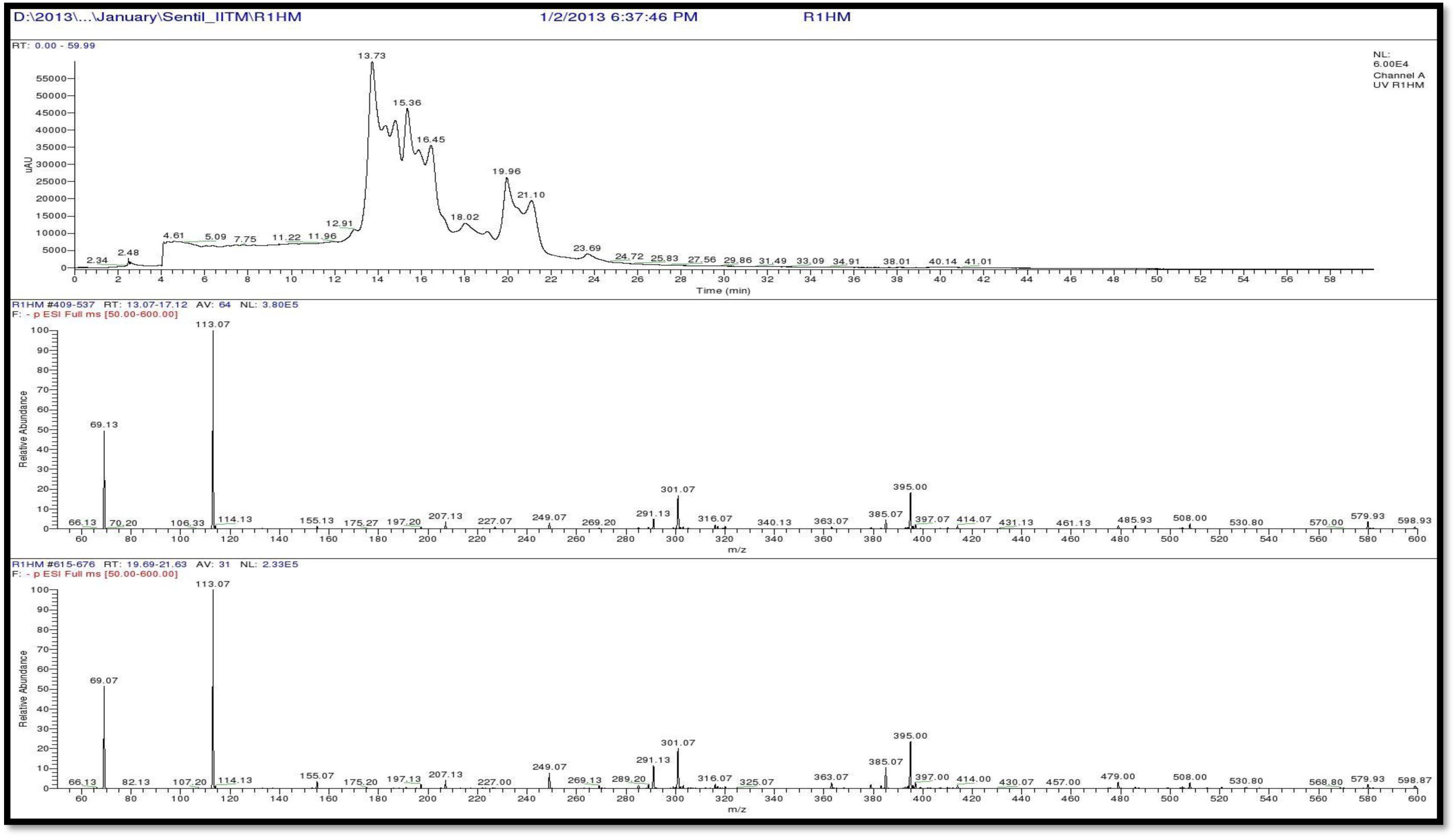
LCMS analysis of L-FDAA derivatives of R1 (Negative mode)

**Figure S161.**
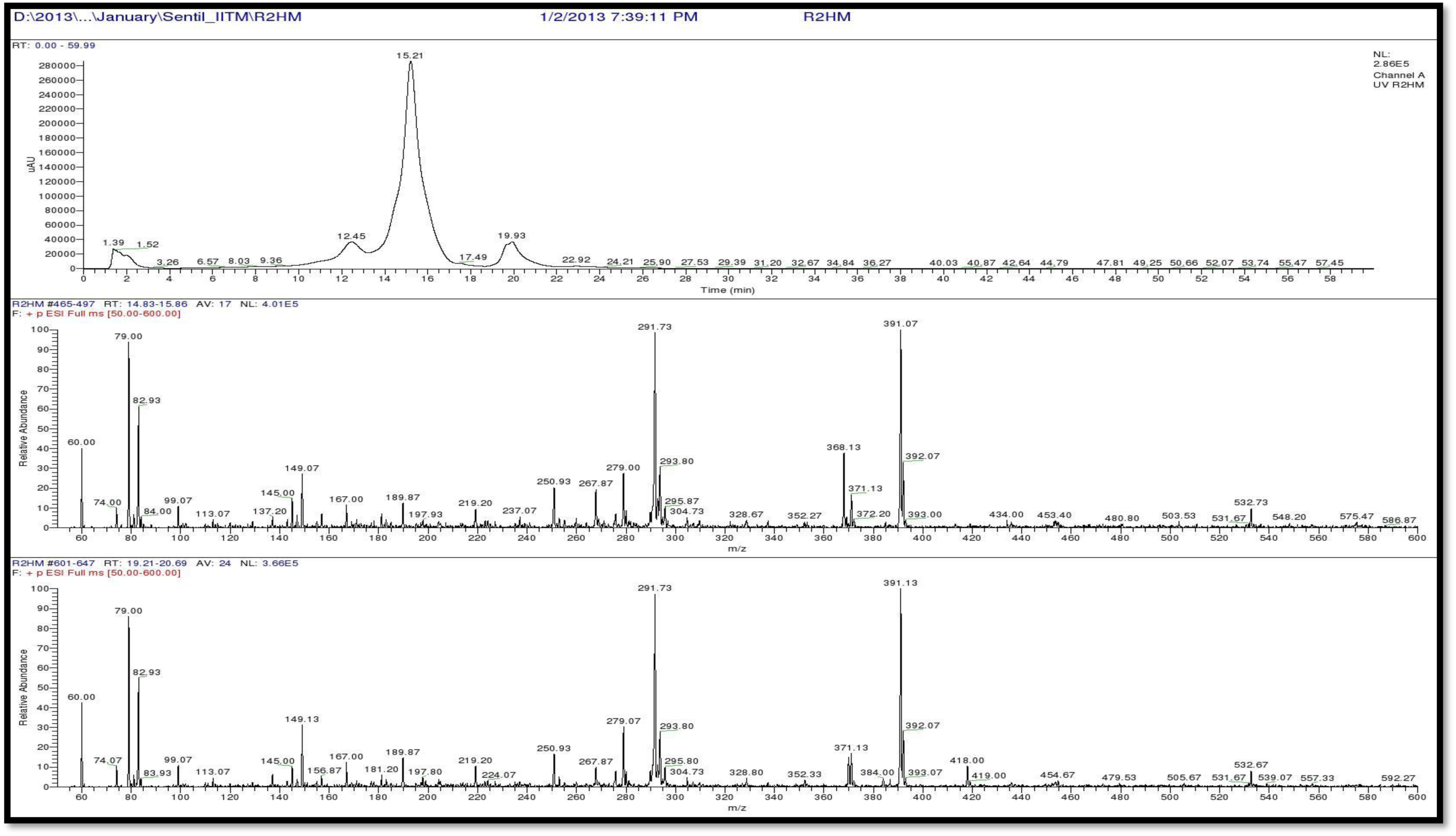
LCMS analysis of L-FDAA derivatives of R3 (Positive mode)

**Table S9a.** Analysis of L-FDAA derivates of acid hydrolysate of Transitmycin R2 by HPLCMS.

| Amino acids | LCMS retention times<br>Marfey's derivatives of standard amino acids |  | HPLC retention time<br>Marfey derivatives of<br>acid hydrolysate of<br>Transitmycin R1 | M+H]<sup>+</sup><br>m/z | [M-H]<sup>-</sup><br>m/z | Assignment |
| --- | --- | --- | --- | --- | --- | --- |
|  | D | L |  | Positive<br>mode | Negative<br>mode |  |
| Threonine | 9.90 | 13.50 | 13.73 | 372.13 | 370.13 | L |
| Proline | 15.16 | 14.31 | 14.23 & 15.36 | 366.13 | 363.07 | D&L |
| **Valine** | **19.65** | **17.65** | **18.02** | **370.13** | **368.07** | **L** |
| N-methyl valine | 20.33 | 19.25 | 19.96 | 384.20 | 382.20 | L |

**Table S9b.** Analysis of L-FDAA derivates of acid hydrolysate of R3 by HPLCMS.

| Amino acids | LCMS retention times<br>Marfey's derivatives of standard amino acids |  | HPLC retention time<br>Marfey derivatives of acid<br>hydrolysate of R3<br>minutes | [M+H]<sup>+</sup><br>m/z | [M-H]<sup>-</sup><br>m/z | Assignment |
| --- | --- | --- | --- | --- | --- | --- |
|  | D minutes | L minutes |  | Positive mode | Negative mode |  |
| Threonine | 9.90 | 13.50 | 12.45 | 372.13 | 370.13 | L |
| Proline | 15.16 | 14.31 | 14.25 | 366.13 | 363.07 | D & L |
| **Valine** | **19.65** | **17.65** | **17.49** | **370.13** | **368.07** | **L** |
| N-methyl valine | 20.33 | 19.25 | 19.93 | 384.20 | 382.20 | L |

**Table S10.**
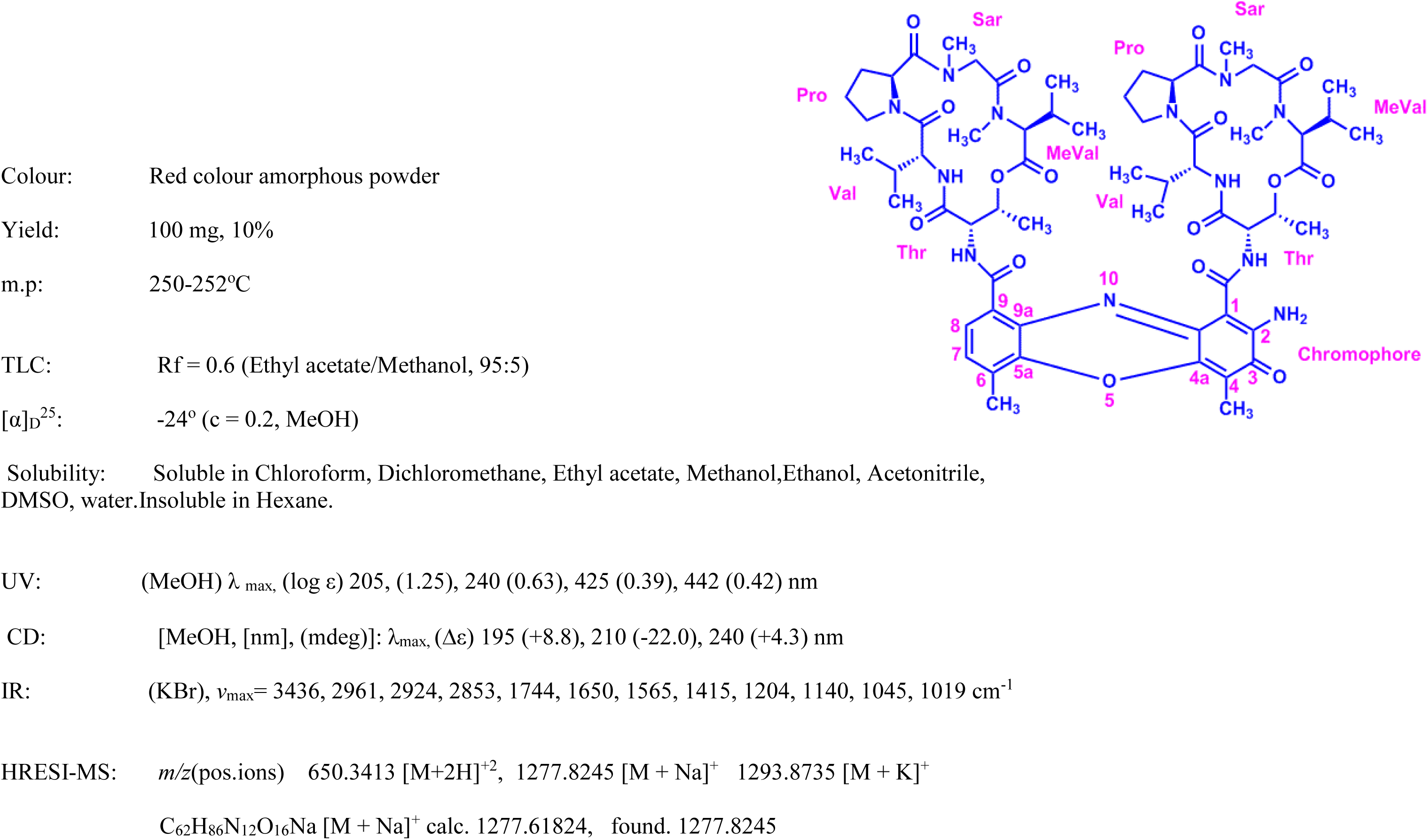
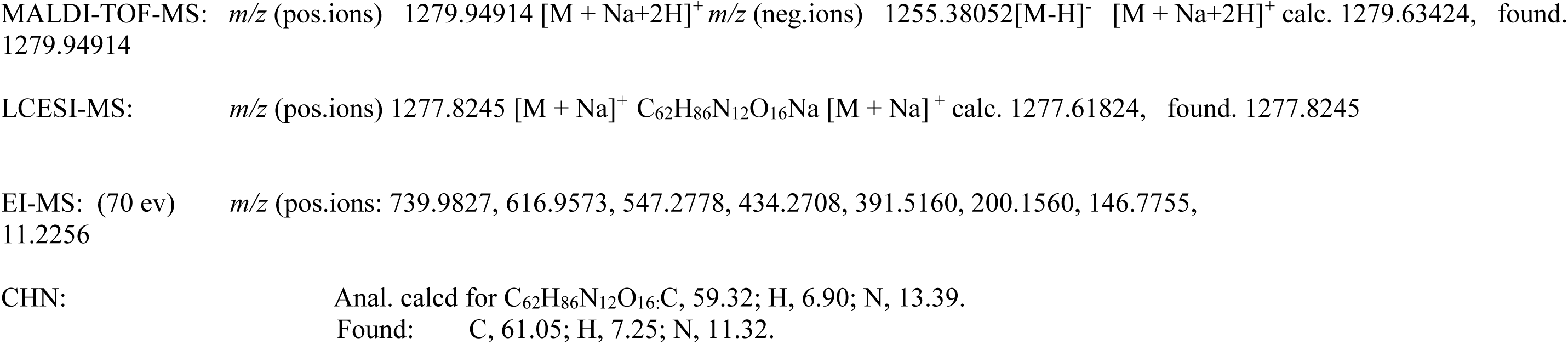
Physico-chemical properties of R2.

**Table S11.**
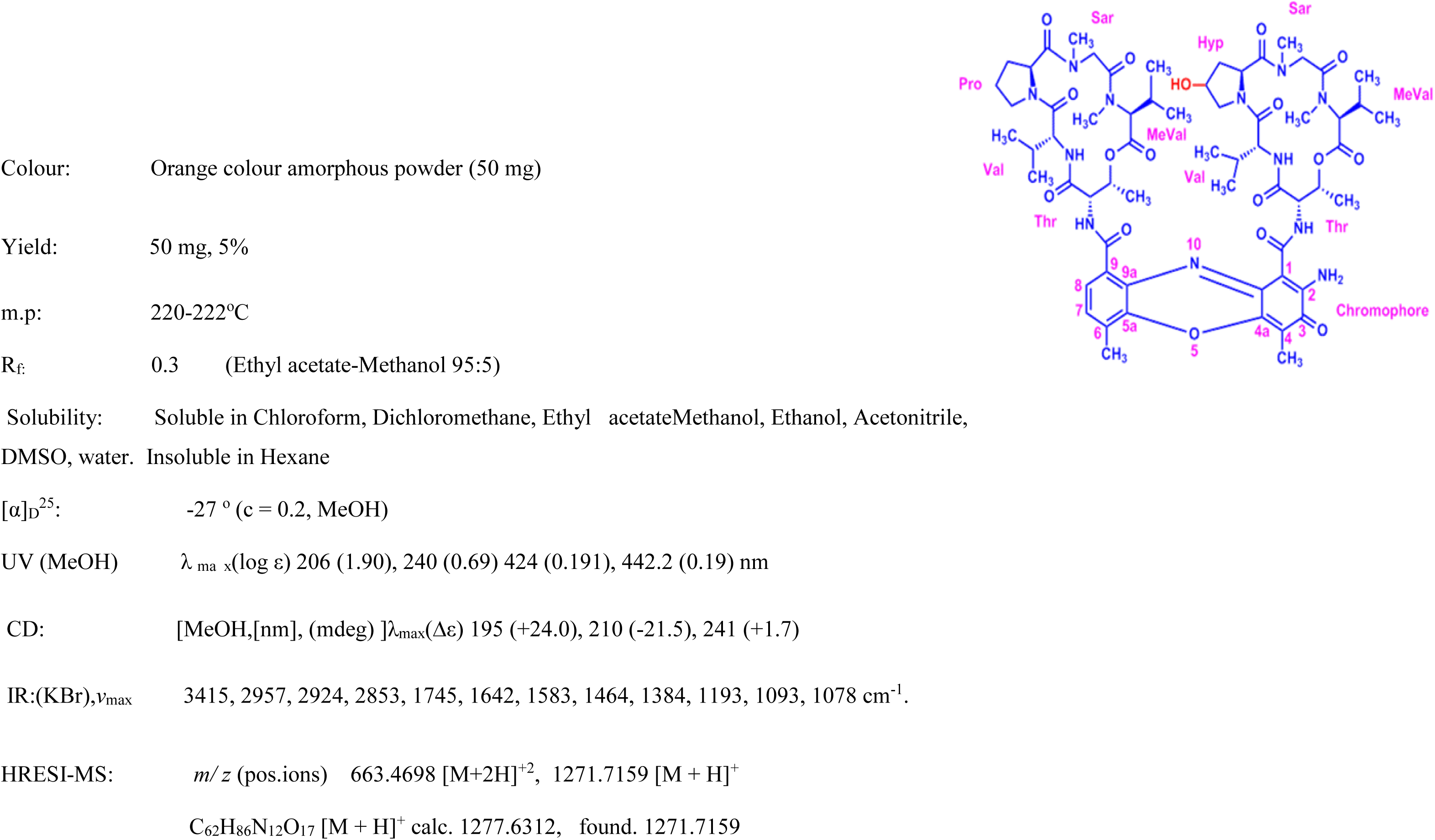
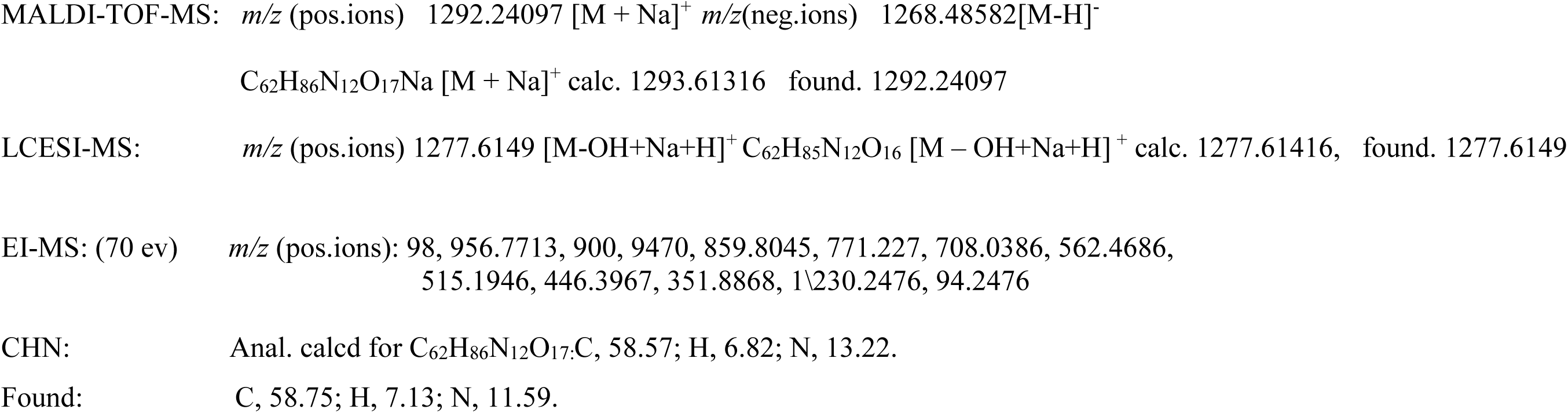
Physico-chemical properties of R3.

**Supplementary Table 12:** Drug resistant profile of the clinical isolates of *Mycobacterium tuberculosis* used to determine the anti TB activity of Transitmycin. R: Resistant; S: Susceptible. The drug resistant profile was determined by LJ based Proportion sensitivity method.

| **Culture No.** | **Isoniazid** | **Rifampicin** | **Kanamycin** | **Ofloxacin** |
| --- | --- | --- | --- | --- |
| 1-49 | S | S | S | S |
| 50 | R | S | S | S |
| 51 | R | S | S | S |
| 52 | S | R | S | S |
| 53 | R | S | S | S |
| 54 | S | S | S | R |
| 55 | R | S | S | S |
| 56 | R | S | S | R |
| 57 | R | S | S | S |
| 58 | R | S | S | S |
| 59 | R | S | R | R |
| 60 | R | R | R | S |
| 61 | S | S | R | S |
| 62 | R | S | S | R |
| 63 | R | S | S | R |
| 64 | R | S | R | R |
| 65 | R | S | S | S |
| 66 | R | S | S | R |
| 67 | R | S | S | R |
| 68 | R | S | R | R |
| 69 | S | S | S | R |
| 70 | S | S | S | R |
| 71 | R | S | S | S |
| 72 | R | S | S | R |
| 73 | R | R | S | S |
| 74 | R | S | S | S |
| 75 | R | R | S | S |
| 76 | R | R | R | S |
| 77 | R | R | S | R |
| 78 | R | R | S | R |
| 79 | R | S | S | S |
| 80 | R | S | S | R |
| 81 | R | S | S | S |
| 82 | R | R | S | S |
| 83 | R | R | S | S |
| 84 | R | R | S | R |
| 85 | R | R | S | S |
| 86 | R | R | S | R |
| 87 | S | R | S | R |
| 88 | R | R | S | S |
| 89 | R | R | S | S |
| 90 | S | R | S | R |
| 91 | S | S | S | R |
| 92 | R | R | S | S |
| 93 | R | R | S | R |
| 94 | S | R | S | R |
| 95 | R | S | S | R |
| 96 | R | R | S | S |
| 97 | R | S | S | R |

**S - Table 13.** Anti-HIV activity of Transitmycin on various HIV-1 subtypes.

| **HIV-1 subtype** | **IC<sub>50</sub> values at various concentration of Transitmycin**<br>**(in µg/mL)** |  |  |  |  | **EC<sub>50</sub> Value (µM)** |
| --- | --- | --- | --- | --- | --- | --- |
|  | **0** | **0.01** | **0.1** | **1** | **10** |  |
| **SUB A** | 4624.7 ± 348.34 | 2600.31 ± 212.27 | 1413±84.36 | 116.00 ± 13.61 | 133.32 ± 13.33 | 0.263 |
| **SUB B** | 9123.78 ± 495.29 | 8419.48 ± 549.73 | 7470.78 ± 3208.54 | 4222.52 ± 138.79 | 1693.95±35.38 | 2.347 |
| **SUB C** | 11652.5 ± 1339.97 | 10962.821± 729.96 | 11005.35±144.9 | 3435.69 ± 75.80 | 578.46 ± 65.77 | 2.167 |
| **SUB D** | 10170.22 ± 1572 | 10761.7 ± 576.85 | 7405.783±303.46 | 1339.49±287.06 | 306.53± 27.43 | 0.447 |
| **SUB E** | 3976.34 ± 68.35 | 3726.96±590.53 | 2120.49±155.83 | 970.25 ±38.08 | 258.84 ± 3.87 | 0.78 |
| **SUB A/C** | 5871.52 ± 51.10 | 5562.45±396.06 | 4654.15 ±262.26 | 589.52 ± 133.80 | 154.45 ±30.20 | 0.685 |
| **NEV RES** | 3325.76 ± 259.01 | 3636.10 ±743.95 | 3278.46 ±329.16 | 627.37 ±69.58 | 341.00 ±7.88 | 0.771 |
| **AZT RES** | 3031.83 ± 821.87 | 3040.03±703.81 | 2213.102± 48.17 | 1066.38 ± 17.34 | 786.32 ±37.46 | 0.388 |

## Acknowledgements for Characterisation of Transitmycin

The Chairman, NMR Research Centre, IISC, Bangalore

Dr.A.Mohan, Mr.Venkatesan, Department of Chemistry, IIT Madras

Dr.M.S.Moni, Dr.C. Baby, Mr. R. Bhaskar, SAIF IIT Madras for 2D NMR analysis

Prof. T. Pradeep, AnanyaBaksi, DST Unit of Nano science IIT Madras for QTRAP LC/MS/MS

Prof. T. Pradeep, Mr.Kamalesh, Mr. E. Sundaraj, DST Unit of Nano Science IIT Madras for MALDI-TOF

Mrs.Sunita Prakash, Molecular Biophysics Department, IISC Bangalore for ESI-MS/MS, MALDI TOF-MS/MS

Mrs.Santha, Mr.Thankarasu, Department of Chemistry IIT Madras for Q-TOF-ESI-MS

Mr. Sankar, SAIF, IIT Madras for IR, EI-MS

The Head, SAIF CDRI, Luknow for LC-ESI-MS

Mr.Muralidhar, SID IISC Bangalore LC-ESI-MS

Prof.Anju Chanda, Mr. T.Saravanan, Department of Biotech IIT Madras for Optical Rotation measurement

Mrs.S.Srividya, Department of Chemistry, IIT Madras.

Dr. P.M.Sivakumar, Department of Biotechnology, Indian Institute of Technology Madras, Chennai, Tamilnadu, 600036, India

Dr. Veluchamy Prabhawathi Department of Biotechnology, Indian Institute of Technology Madras, Chennai, Tamilnadu, 600036, India

## Notes

### Competing Interest Statement

The authors have declared no competing interest.

### Funding Statement

Yes

### Author Declarations

Donor PBMC were obtained from healthy volunteers after obtaining the approval of the Institutional Ethics Committee of the National Institute for Research in Tuberculosis as well as the informed consent of the participants.

### Summary of Updates

We have added more supporting data in this pdf files

